# Multi-ancestry genome-wide meta-analysis in Parkinson’s disease

**DOI:** 10.1101/2022.08.04.22278432

**Authors:** Jonggeol Jeffrey Kim, Dan Vitale, Diego Véliz Otani, Michelle Lian, Karl Heilbron, the 23andMe Research Team, Hirotaka Iwaki, Julie Lake, Caroline Warly Solsberg, Hampton Leonard, Mary B. Makarious, Eng-King Tan, Andrew B. Singleton, Sara Bandres-Ciga, Alastair J Noyce, Cornelis Blauwendraat, Mike A. Nalls, Jia Nee Foo, Ignacio Mata

## Abstract

Although over 90 independent risk variants have been identified for Parkinson’s disease using genome-wide association studies, all studies have been performed in just one population at the time. Here we performed the first large-scale multi-ancestry meta-analysis of Parkinson’s disease with 49,049 cases, 18,785 proxy cases, and 2,458,063 controls including individuals of European, East Asian, Latin American, and African ancestry. In a single joint meta-analysis, we identified 78 independent genome-wide significant loci including 12 potentially novel loci (*MTF2, RP11-360P21*.*2, ADD1, SYBU, IRS2, USP8:RP11-562A8*.*5, PIGL, FASN, MYLK2, AJ006998*.*2, Y_RNA, PPP6R2*) and finemapped 6 putative causal variants at 6 known PD loci. By combining our results with publicly available eQTL data, we identified 23 genes near these novel loci whose expression is associated with PD risk. This work lays the groundwork for future efforts aimed at identifying PD loci in non-European populations.

## Introduction

Parkinson’s disease (PD) is a neurodegenerative disease pathologically defined by Lewy body inclusions in the brain and the death of dopaminergic neurons in the midbrain. The identification of genetic risk factors is imperative to mitigating the global burden of PD, one of the fastest growing age-related neurodegenerative diseases. The largest PD Genome-Wide Association Study (GWAS) meta-analysis to date uncovered 90 independent genetic risk variants in individuals of European ancestry ^1^. Similarly, large-scale PD GWAS meta-analyses of East Asian^2^ and a single GWAS of Latin American^3^ individuals have each identified 2 risk loci that were not previously identified in Europeans. For PD, there are now large-scale efforts to sequence and analyze genomic data in under-represented populations with the goal of both identifying novel associated loci, fine mapping known loci, and addressing the inequality that exists in current precision medicine efforts^4,5^. Here we performed a large-scale multi-ancestry GWAS meta-analysis of PD by including individuals from four ancestral populations. This effort can serve as a guide for future genetic analyses to increase ancestral representation.

## Methods

In brief, we used a single joint meta-analysis study design (Figure 1) due its efficiency over a two-stage GWAS design^6^. All datasets underwent quality control filters and were harmonized to hg19. We used datasets representing four different ancestry groups: European, East Asian, Latin American, and African. In addition to previously described results from European^1^, East Asian^2^, and Latin American^3^ cohorts, we also used FinnGen and additional datasets for East Asian, Latin American and African cohorts from 23andMe, Inc. All multi-ancestry meta-analyses were conducted using a union of a random-effects model and a multi-ancestry meta-regression model using PLINK 1.9^7^ and MR-MEGA^8^. MR-MEGA has the greatest power to detect heterogeneous effects across the different cohorts, while the random-effects model has greater power to detect homogenous allelic effects^8^. In total, we included 49,049 PD cases, 18,618 proxy cases (first-degree relative with PD), and 2,458,063 neurologically-healthy controls.

**Figure 1.**
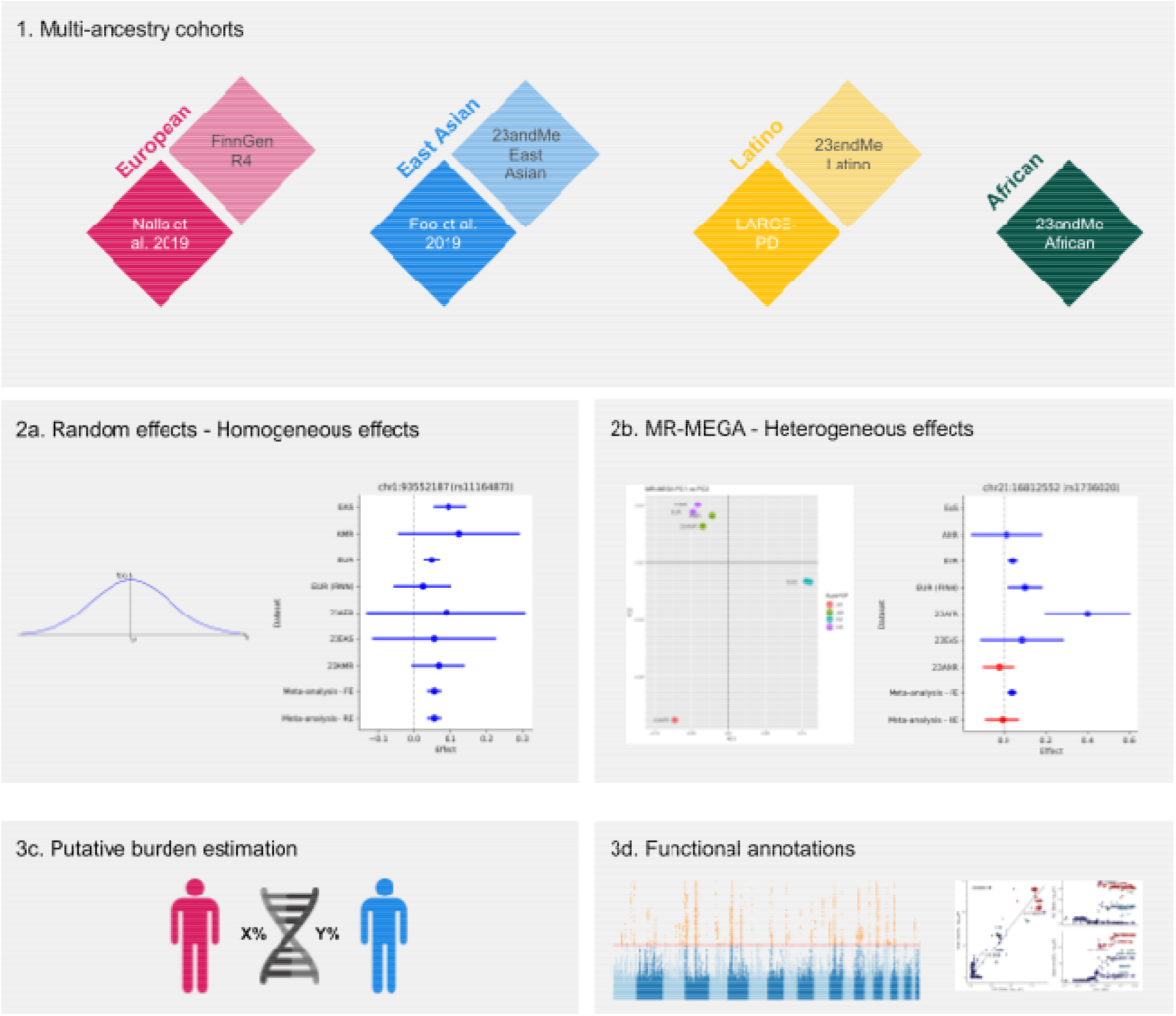
Study design of Multi-Ancestry Meta-Analysis

**Figure 2.**
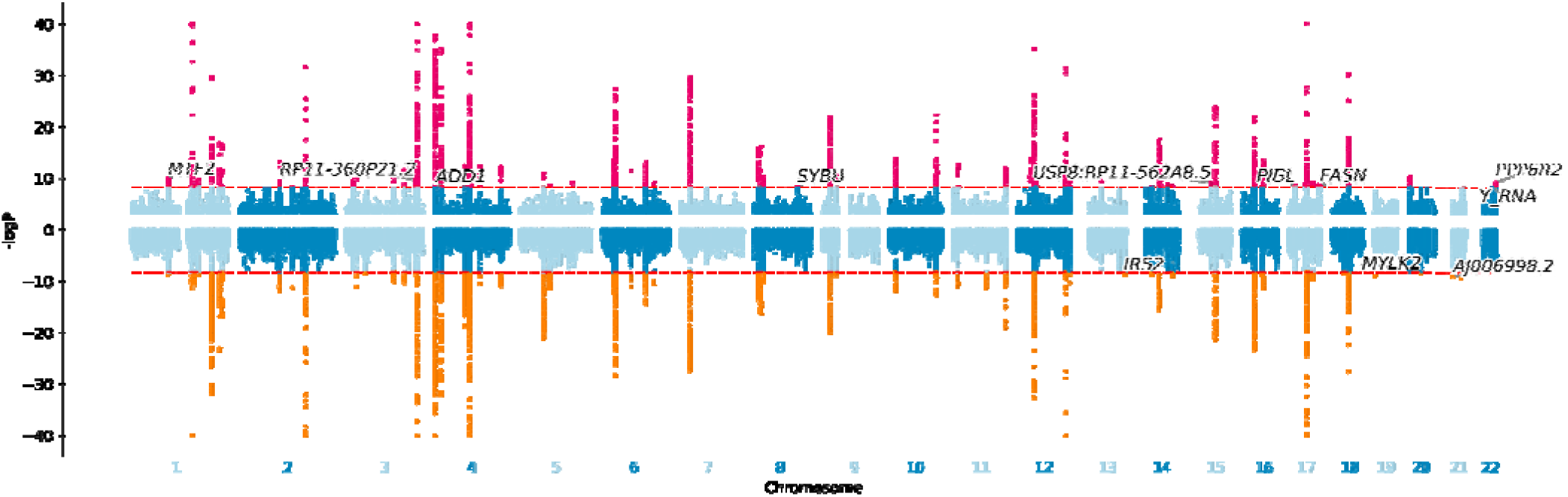
: Miami plot of random-effect and MR-MEGA meta-analyses. Top: random effect; Bottom: MR-MEGA. Dotted lines indicate the bonferroni adjusted significant threshold of P < 5 × 10^−9^. All -log10P values greater than 40 were truncated to 40 for visual clarity. SNPs significant in random-effect and MR-MEGA models are colored red and orange respectively. Novel loci are indicated by the nearest gene.

Bonferroni-adjusted alpha for genome-wide significant variants was set to a more stringent 5 × 10^−9^ for all multi-ancestry meta-analyses to account for the larger number of haplotypes resulting from the ancestrally diverse datasets^9^. Fine-mapping was also performed using MR-MEGA, which uses ancestry heterogeneity to increase fine-mapping resolution.

We ran our meta-analysis results through the Proportion of Population-specific and Shared Causal Variants (PESCA)^10^ software to estimate the population specific and shared burden on the variants found in the meta-analysis. Only East Asian and European populations were compared due to the smaller sample size of the other populations. We used the Functional Mapping and Annotation (FUMA) software^11,12^ to functionally annotate the random effect results and run Multi-marker Analysis of GenoMic Annotation (MAGMA)^13^ for gene-ontology, tissue-level, and single-cell expression data. We also searched our novel loci through GTEx v8 brain tissue eQTLs and multi-ancestry eQTL meta-analysis of the brain^14^ to correlate novel loci with gene expression data, then searched the significant-eQTL genes and genes near the loci with previously completed summary-based mendelian randomization (SMR)^15^ results to correlate said genes with PD risk. Detailed methods are available in the Online Methods and the Data and Code Availability sections.

## Results

### Multi-ancestry meta-analyses confirm 62 known loci and nominate 12 novel loci

All together we found 12 novel PD risk loci and 66 hits in known risk loci from single ancestry GWAS (Supplementary Table S2). 9 of the novel loci found in the random-effect method showed homogeneous effects across the different ancestries. The 3 loci identified exclusively in MR-MEGA showed ancestrally heterogeneous effects. All three loci showed evidence for ancestral heterogeneity (P_ANC-HET_ < 0.05) but no significant residual heterogeneity (P_RES-HET_ > 0.148), supporting the idea that the signals are due to population structural differences rather than other confounding factors. The *IRS2* locus (P_ANC-HET_ = 5.3 × 10^−3^) shows the biggest departure in allelic effects in the Finnish cohort (Supplementary Figure 4). The *MYLK2* locus has the main European cohort at odds with the Latin American and Finnish cohorts (P_ANC-HET_ = 0.035). While this is a novel single-trait GWAS locus, its lead SNP was previously discovered as a potential pleiotropic locus in a multi-trait conditional/conjunctional false discovery rate (FDR) study between schizophrenia and PD^16^. Lastly, the *AJ006998*.*2* locus had the most significant ancestral heterogeneity (P_ANC-HET_ = 4.74 × 10^−5^) and its effects were specific to European and African cohorts.

We found 17 suggestive loci that failed to meet the significance threshold at random-effect but met the more common threshold of P < 5 × 10^−8^ in fixed-effect and P < 1 × 10^−6^ in random-effect (Supplementary Table S2). 14 of these regions qualified as novel loci. Two loci near *RP11-182I10*.*3* and *HS1BP3* were exclusively found in the 23andMe Latin American and African cohorts. The lead SNPs (rs578139575, rs73919910) for these loci are very rare in European populations but more common in Africans and Latin Americans (gnomAD v3.1.2 EUR: 0.0001616, 0.002307; AFR: 0.01637, 0.08837; AMR:0.004063, 0.01905). If confirmed, these loci would confer a large amount of PD protective effect (beta: -1.3, -0.54). These loci merit further studies in the African and Latino populations.

### Fine mapping finds 6 putative SNPs in 6 loci

6 putative causal variants were found in 6 loci in *TMEM163, TMEM175, SNCA, HIP1R, MAPT*, and *LSM7* (Table 4). All putative causal variants had posterior probabilities (PP) of causality > 0.99 and overlapped with known PD loci. Our results confirmed the previous results of the *TMEM175* M393T coding variant as the likely causal variant^17^. We reported only single-variant 99% credible sets as MR-MEGA assumes a single causal variant in a GWAS signal. 99% credible sets in 57 additional loci can be found in Supplementary Table S3.

### MAGMA Gene Ontology analysis finds enrichment in brain and pituitary tissues

Out of 15,479 gene ontology sets in MsigDB c5, 111 gene sets were significantly enriched among the random effect hits, a significant increase from previous 11 gene sets in Nalls et al. 2019. Out of the 11, only 3 gene ontology terms were replicated in the multi-ancestry: vacuole (P_FDR_ = 6.9 × 10^−4^), vacuolar part (P_FDR_ = 0.009), and vacuolar membrane (P_FDR_ = 0.01). At the tissue level the genes of interest were enriched in all brain cells as well as pituitary tissue (Supplementary Figure 9), consistent with the results from Nalls et al. 2019^1^.

When analyzing single-cell RNAseq data, there was no expression enrichment across 88 brain cell types in mouse brain data DropViz ^18^ (Supplementary Figure 13). When narrowed down to substantia nigra tissue, glutamate receptor (Grin2c) in neurons were enriched (Supplementary Figure 13). In human midbrain data ^19^ oculomotor and trochlear nucleus (OMTN), dopaminergic (DA1), neuroblast GABAnergic (NbGABA), and GABA were enriched (Supplementary Figure 13).

Out of 15,479 gene ontology sets in MsigDB c5, 110 gene sets were significantly enriched among the random effect hits (Supplementary Table S6), a significant increase from previous 11 gene sets in Nalls et al. 2019. Out of the 11, only 3 gene ontology terms were replicated in the multi-ancestry: vacuole (P_FDR_ = 6.9 × 10^−4^), vacuolar part (P_FDR_ = 0.009), and vacuolar membrane (P_FDR_ = 0.01). At the tissue level the genes of interest were enriched in all brain cells as well as pituitary tissue (Supplementary Figure 9), consistent with the results from Nalls et al. 2019^1^.

### eQTL and SMR nominate 23 putative genes near novel loci

When comparing the SNPs in novel loci with multi-ancestry brain eQTL results^14^, 28 genes showed high levels of association (Supplementary Figures 8). SMR found 23 genes near four novel loci associated with PD risk. Interestingly, *PPP6R2* and *CENPV* expression changes in *substantia nigra* were associated with PD risk. *PPP6R2* or protein phosphatase 6 regulatory subunit 2 is a regulatory protein for Protein Phosphatase 6 Catalytic Subunit (*PPP6C*), which is involved in the vesicle-mediated transport pathway. *CENPV* or Centromere Protein V is involved in centromere formation and cell division. Detailed results for eQTL and the SMR can be found in Supplementary Table S8, S9, and S10.

## Discussion

This is the first large-scale GWAS meta-analysis of PD that incorporates multiple diverse ancestry populations. From the joint cohort analysis, we were able to confirm 65 independent known risk loci and identify 12 potentially novel risk loci, 9 with homogeneous effect and 3 with heterogeneous effect across the different cohorts. We further found 17 suggestive loci at the more traditional method of fixed-effect at 5 × 10^−8^ and fine-mapped 11 putative SNPs in 11 of the known loci by leveraging the diverse ancestry populations. We highlighted tissues and cell types involved with the loci, which were consistent with previous findings^1^. Finally we used SMR to nominate 23 putative genes near our novel loci.

Novel loci contain genes previously implicated with PD. The *MTF2* and *PPP6R2* loci contain genes *TMED5* and *PPP6R2*, which intersect with golgi body^20^ and vesicular transport pathways^21,22^, both implicated in PD pathogenesis^23–28^. Because *substantia nigra* deterioration is a hallmark pathogenic feature of PD, *PPP6R2*, and *CENPV* expression in *substantia nigra* and their association with PD merit additional investigation. Within a known locus, a new independent signal was found in *RILPL1* (rs28659953), which interacts with LRRK2-phosphorylated Rab10 to block primary cilia generation^29^. All potentially novel PD loci identified in this analysis will require additional replication and functional validation to elucidate their role in PD pathogenesis. Previous findings in European populations found that the known variants explained 16–36% of the heritable risk^1^, and while we did not test for genetic liability of the novel loci, they have been added to the list of variants that explain potential heritable PD risk.

We found that 26 of the 68 detected known PD loci have nominal ancestral heterogeneity (P_ANC-HET_ < 0.05) and 5 significant after bonferroni correction (P_ANC-HET_ < 0.05/4,512 significant SNPs) (Supplementary Table S2). This heterogeneity may be caused by significant differences in allele effect sizes and frequencies between the different ancestry populations and thus should be studied as potential loci with ancestrally divergent risk. 18 known loci from single-ancestry GWASes failed to replicate (Supplementary Table S4) which implies that the variants associated in these loci may only be relevant in certain populations. However, it is worth noting that there are large differences in statistical power for each included ancestry. Additional population-specific loci will likely reach significance when larger sample sizes are available for non-European datasets.

Our fine mapping isolated several putative causal variants in previously discovered loci. *TMEM175*-rs34311866 has been previously identified as functionally relevant to PD risk^17^, which is consistent with our fine-mapping results. While the fine-mapping results provided by MR-MEGA are sufficient to identify putative causal variants for loci driven by one independent signal, multiple variants in a locus can contribute to complex traits. The additive and epistatic effects of multiple causal variants in a locus can be difficult to interpret when the effects associated with each independent signal are small.

Although this is the largest multi-ancestry PD meta-analysis GWAS to date, the European population is still overrepresented in this analysis. Around 79% of effective PD cases are of European descent. Individuals of African descent were particularly underpowered at just 0.5% of the effective PD cases. The discoveries in our study warrant future efforts to expand the studies in more diverse populations. The Global Parkinson’s Genetics Program (GP2) is partnering with institutions that care for underrepresented populations to generate data for these underserved communities all over the world^5^, and we will continue the ongoing analysis as more participants are genotyped. Just as the first PD GWASs failed to identify significant signals ^30,31^, we are confident that future diverse-ancestry GWAS will produce impactful association results as sample sizes increase. Further efforts in multi-ancestry and non-European GWAS will identify loci that are more relevant to the global population and will continue to facilitate fine mapping efforts to identify the genetic variants that drive these associations.

## Data Availability

The analysis pipeline code is available on GP2 github: https://github.com/GP2code/GP2-WorkingGroups/tree/main/CD-DAWG-Complex-Data-Analysis/PROJECTS/GP2-Multiancestry-PD
GWAS summary statistics for 23andMe datasets (post-Chang and data included in Chang et al. 2017 and Nalls et al. 2014) will be made available through 23andMe to qualified researchers under an agreement with 23andMe that protects the privacy of the 23andMe participants. Please visit research.23andme.com/collaborate/#publication for more information and to apply to access the data. An immediately accessible version of the summary statistics is available here https://drive.google.com/file/d/1TmDZNFgyQvsOZ0xu-aZmBpVCpeUUa0UX/
excluding Nalls et al. 2014, 23andMe post-Chang et al. 2017 and Web-Based Study of Parkinson's Disease (PDWBS) but including all analyzed SNPs. After applying with 23andMe, the full summary statistics including all analyzed SNPs and samples in this GWAS meta-analysis will be accessible to the approved researcher(s).

https://drive.google.com/file/d/1TmDZNFgyQvsOZ0xu-aZmBpVCpeUUa0UX/

## Data and code availability

The analysis pipeline code is available on GP2 github: https://github.com/GP2code/GP2-WorkingGroups/tree/main/CD-DAWG-Complex-Data-Analysis/PROJECTS/GP2-Multiancestry-PD

GWAS summary statistics for 23andMe datasets (post-Chang and data included in Chang et al. 2017 and Nalls et al. 2014) will be made available through 23andMe to qualified researchers under an agreement with 23andMe that protects the privacy of the 23andMe participants. Please visit research.23andme.com/collaborate/#publication for more information and to apply to access the data. An immediately accessible version of the summary statistics is available here https://drive.google.com/file/d/1TmDZNFgyQvsOZ0xu-aZmBpVCpeUUa0UX/ excluding Nalls et al. 2014, 23andMe post-Chang et al. 2017 and Web-Based Study of Parkinson’s Disease (PDWBS) but including all analyzed SNPs. After applying with 23andMe, the full summary statistics including all analyzed SNPs and samples in this GWAS meta-analysis will be accessible to the approved researcher(s).

## Acknowledgements

M.A.N.’s research was supported in part by the Intramural Research Program of the NIH, National Institute on Aging (NIA), National Institutes of Health, Department of Health and Human Services; project number ZO1 AG000535, as well as the National Institute of Neurological Disorders and Stroke (NINDS), and participation in this project was part of a competitive contract awarded to Data Tecnica International LLC by the National Institutes of Health to support open science research.

I.F.M.’s research is supported by grants from the National Institute of Neurological Disorders and Stroke under (R01NS112499), the Parkinson’s Foundation (Stanley Fahn Junior Faculty Award and an International Research Grants Program award), the Michael J Fox Foundation, Aligning Science Across Parkinson’s Global Parkinson’s Genetic Project (ASAP-GP2) and the American Parkinson’s Disease Association.

E.K.T. is supported by a grant from the National Medical Research Council Singapore, Open Fund Large Collaborative Grant (MOH-OFLCG-000207). J.N.F. is supported by the Singapore National Research Foundation Fellowship (NRF-NRFF2016-03) and the National Medical Research Council Singapore, Open Fund Individual Research Grant (MOH-000559).

A.J.N. reports grants from Parkinson’s UK, Barts Charity, Cure Parkinson’s, NIHR, Innovate UK, Virginia Keiley benefaction, Alchemab, Aligning Science Across Parkinson’s Global Parkinson’s Genetics Program (ASAP-GP2) and Michael J Fox Foundation.

This research has been conducted using the UK Biobank Resource under Application Number 33601. We want to acknowledge the participants and investigators of FinnGen study.

We thank the research participants and employees of 23andMe. The following members of the 23andMe Research Team contributed to this study:

Stella Aslibekyan, Adam Auton, Elizabeth Babalola, Robert K. Bell, Jessica Bielenberg, Katarzyna Bryc, Emily Bullis, Paul Cannon, Daniella Coker, Gabriel Cuellar Partida, Devika Dhamija, Sayantan Das, Sarah L. Elson, Nicholas Eriksson, Teresa Filshtein, Alison Fitch, Kipper Fletez-Brant, Pierre Fontanillas, Will Freyman, Julie M. Granka, Karl Heilbron, Alejandro Hernandez, Barry Hicks, David A. Hinds, Ethan M. Jewett, Yunxuan Jiang, Katelyn Kukar, Alan Kwong, Keng-Han Lin, Bianca A. Llamas, Maya Lowe, Jey C. McCreight, Matthew H. McIntyre, Steven J. Micheletti, Meghan E. Moreno, Priyanka Nandakumar, Dominique T. Nguyen, Elizabeth S. Noblin, Jared O’Connell, Aaron A. Petrakovitz, G. David Poznik, Alexandra Reynoso, Madeleine Schloetter, Morgan Schumacher, Anjali J. Shastri, Janie F. Shelton, Jingchunzi Shi, Suyash Shringarpure, Qiaojuan Jane Su, Susana A. Tat, Christophe Toukam Tchakouté, Vinh Tran, Joyce Y. Tung, Xin Wang, Wei Wang, Catherine H. Weldon, Peter Wilton, Corinna D. Wong

This research was funded by Aligning Science Across Parkinson’s through the Michael J. Fox Foundation for Parkinson’s Research (MJFF).

## Online Methods

### Study Design and Cohort Descriptions

We used a single joint meta-analysis study design to maximize statistical power ^6^. We used datasets representing four different ancestry groups: European, East Asian, Latin American, and African. The meta-analysis included 49,049 PD cases, 49,049 PD proxy cases (participant with a parent with PD) and 2,458,063 neurologically normal controls (Table 1, Supplementary Table S1). GWAS results of European^1^, East Asian^2^, and Latin American^3^ populations were previously reported. African dataset as well as the additional Latin American and East Asian PD GWAS summary statistics were provided by 23andMe. The Finnish PD GWAS summary statistics was acquired from FinnGen Release 4 (G6_PARKINSON_EXMORE). For the FinnGen data, we chose the endpoint “Parkinson’s Disease (more controls excluded)” (G6_PARKINSON_EXMORE), which excludes control participants with psychiatric diseases or neurological diseases. While some FinnGen GWAS results also include UK Biobank participants, our FinnGen data did not include any UK Biobank participants.

**Table 1.**
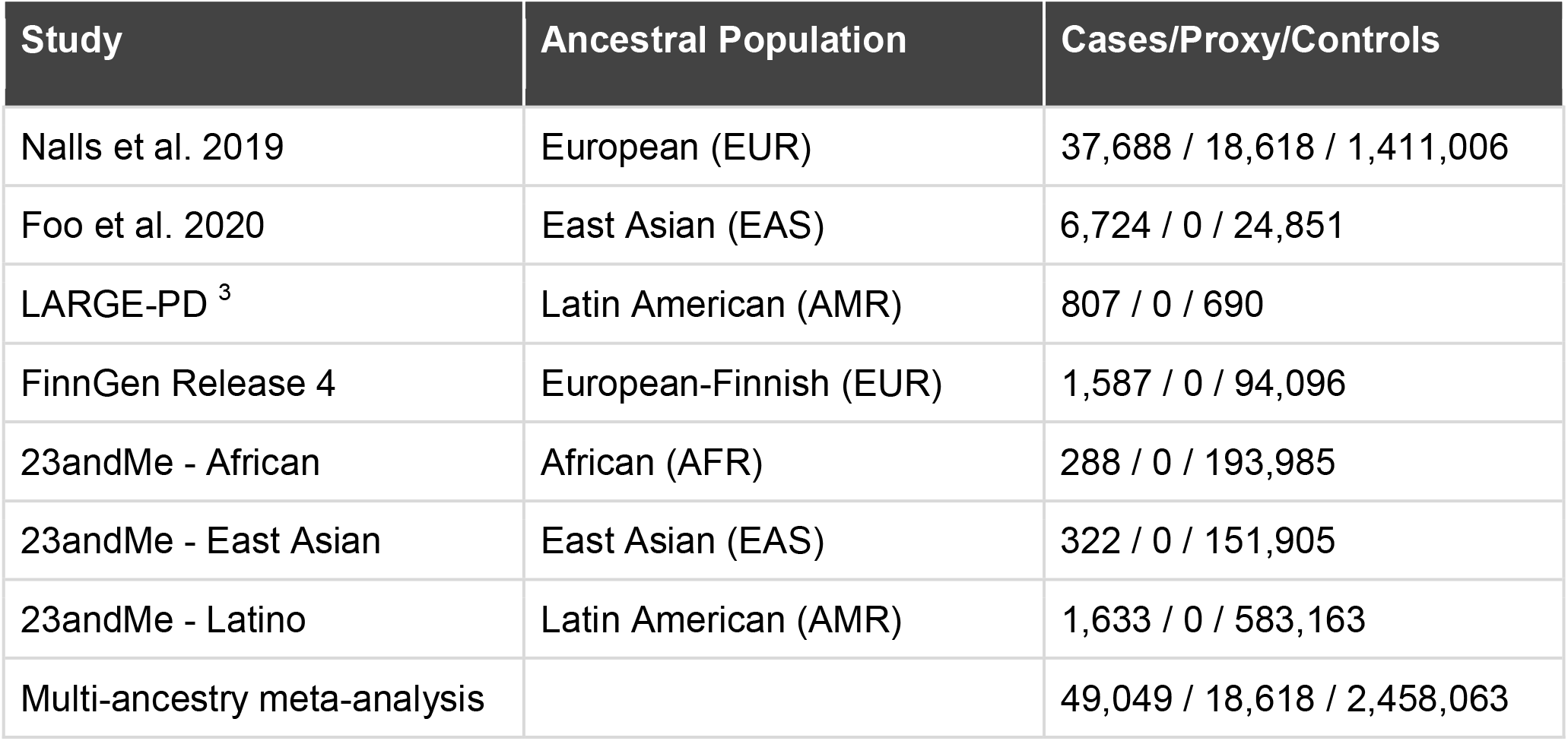
Cohort Description

| Study | Ancestral Population | Cases/Proxy/Controls |
| --- | --- | --- |
| Nalls et al. 2019 | European (EUR) | 37,688 / 18,618 / 1,411,006 |
| Foo et al. 2020 | East Asian (EAS) | 6,724 / 0 / 24,851 |
| LARGE-PD <sup>3</sup> | Latin American (AMR) | 807 / 0 / 690 |
| FinnGen Release 4 | European-Finnish (EUR) | 1,587 / 0 / 94,096 |
| 23andMe - African | African (AFR) | 288 / 0 / 193,985 |
| 23andMe - East Asian | East Asian (EAS) | 322 / 0 / 151,905 |
| 23andMe - Latino | Latin American (AMR) | 1,633 / 0 / 583,163 |
| Multi-ancestry meta-analysis |  | 49,049 / 18,618 / 2,458,063 |

**Table 2.** Meta-analysis results of lead SNPs in the novel loci. RE – random effect, A1 – effect allele, A2 – alternate allele, ANC-HET – ancestry heterogeneity, RES-HET – residual heterogeneity

| rsID | Nearest Gene | CHR:BP:A1:A2 | BETA(RE) | SE | P(RE) | P(MR-MEGA) | P(ANC-HET) | P(RES-HET) |
| --- | --- | --- | --- | --- | --- | --- | --- | --- |
| rs11164870 | *MTF2* | 1:93552187:C:G | 0.054 | 0.009 | **1.15E-10** | **2.64E-09** | 0.229 | 0.928 |
| rs6806917 | *RP11-360P21.2* | 3:178861417:T:C | -0.070 | 0.011 | **1.65E-10** | **3.43E-09** | 0.215 | 0.762 |
| rs16843452 | *ADD1* | 4:2849168:T:C | -0.068 | 0.012 | **4.11E-09** | 3.19E-07 | 0.747 | 0.687 |
| rs6469271 | *SYBU* | 8:110644774:T:C | -0.056 | 0.010 | **3.62E-09** | 2.04E-07 | 0.590 | 0.954 |
| rs1078514 | *IRS2* | 13:110463168:T:C | 0.068 | 0.026 | 0.004817 | **2.30E-09** | **5.30E-03** | 0.261 |
| rs28648524 | *USP8:RP11-562A8.5* | 15:50787409:A:T | 0.064 | 0.010 | **6.45E-10** | 2.58E-08 | 0.406 | 0.661 |
| rs11650438 | *PIGL* | 17:16234260:A:G | 0.050 | 0.009 | **2.93E-09** | 1.46E-07 | 0.528 | 0.288 |
| rs4485435 | *FASN* | 17:80045086:C:G | 0.082 | 0.014 | **2.61E-09** |  |  |  |
| rs6060983 | *MYLK2* | 20:30420924:T:C | 0.069 | 0.037 | 0.03221 | **3.86E-09** | **0.035** | 0.149 |
| rs1736020 | *AJ006998.2* | 21:16812552:A:C | 0.006 | 0.005 | 0.8853 | **1.12E-09** | **4.74E-05** | 0.638 |
| rs73174657 | *Y_RNA* | 22:41434158:A:G | -0.059 | 0.010 | **3.81E-09** | 4.90E-07 | 0.983 | 0.655 |
| rs10775809 | *PPP6R2* | 22:50808017:A:T | 0.092 | 0.015 | **4.09E-10** | 5.61E-08 | 0.943 | 0.903 |

**Table 3.** MR-MEGA fine mapping results for loci with a single SNP within the 99% credible set. Known PD genes are either known PD risk genes (*SNCA, TMEM175, MAPT*) or genes with the highest score in the nearest known PD locus by the PD GWAS Locus Browser^52^

| Locus | Number of significant SNPs | Nominated variant | CHR:BP:A1:A2 | Nearest Gene | Known PD Genes +/- 1MB | Functional Consequence | CADD | RDB |
| --- | --- | --- | --- | --- | --- | --- | --- | --- |
| 11 | 6 | rs57891859 | 2:135464616:A:G | *TMEM163* | *TMEM163* | intronic | 6.746 | 4 |
| 19 | 926 | rs34311866 | 4:951947:C:T | *TMEM175* | *TMEM175* | exonic | 11.09 | NA |
| 23 | 1483 | rs356182 | 4:90626111:A:G | *RP11-115D19.1* | *SNCA* | ncRNA intronic | 8.962 | NA |
| 45 | 1371 | rs10847864 | 12:123326598:G:T | *HIP1R* | *HIP1R* | intronic | 2.403 | 2b |
| 56 | 1060 | rs62053943 | 17:43744203:C:T | *CRHR1:RP11-105N13.4* | *MAPT* | ncRNA intronic | 0.183 | 5 |
| 60 | 1 | rs55818311 | 19:2341047:C:T | *SPPL2B* | *LSM7* | ncRNA exonic | 1.096 | 5 |

### 23andMe diverse ancestry data

All self-reported PD cases and controls from 23andMe provided informed consent and participated in the research online, under a protocol approved by the external AAHRPP-accredited IRB, Ethical & Independent Review Services (E&I Review). Participants were included in the analysis on the basis of consent status as checked at the time data analyses were initiated. The name of the IRB at the time of the approval was Ethical & Independent Review Services. Ethical & Independent Review Services was recently acquired, and its new name as of July 2022 is Salus IRB (https://www.versiticlinicaltrials.org/salusirb). Samples were genotyped on one of five genotyping platforms: V1 and V2, which are variants of Illumina HumanHap550+ BeadChip; V3, Illumina OmniExpress+ BeadChip; V4, Illumina custom array that includes SNPs overlapping V2 and V3 chips; or V5, Illumina Infinium Global Screening Array. For inclusion, samples needed a minimal call rate of 98.5%. Genotyped samples were then phased using either Finch or Eagle2^32^ and imputed using Minimac3 and a reference panel of 1000 Genomes Phase III^33^ and UK10K data^34^. For this replication study, samples were classified as African, East Asian, or Latino using a genotype-based pipeline^35^ consisting of a support vector machine and a hidden Markov model, followed by a logistic classifier to differentiate Latinos from African Americans. Unrelated individuals were included in the analysis, as determined via identity-by-descent (IBD). Variants were tested for association with PD status using logistic regression, adjusting for age, sex, the first five PCs, and genotyping platform. Reported p-values were from a likelihood ratio test.

### Multi-ancestry Meta-Analysis

We performed a multi-ancestry meta-analysis of GWAS results using Meta-Regression of Multi-Ethnic Genetic Association (MR-MEGA)^8^ and PLINK 1.9. MR-MEGA performs a meta-regression by generating axes of genetic variation for each cohort, which are then used as covariates in the meta-analysis to account for differences in population structure. While MR-MEGA was able to generate 4 principal components as axes of genetic variation, 3 principal components visibly separated the super population ancestries and explained 98% of the population variance (Supplementary Figure 7). Therefore we used 3 principal components to minimize overfitting. MR-MEGA has reduced power to detect associations for variants with homogeneous effects across populations. It is therefore recommended to run MR-MEGA alongside another meta-analysis method. PLINK 1.9 was used to perform random-effect (RE) meta-analysis to detect homogenous allelic effects.

Before the analysis, all datasets were harmonized to genome build hg19 using CrossMap^36^ and all variants were filtered by imputation score (r^2^ > 0.3) and minor allele frequency (MAF >= 0.001). Only autosomal variants were kept in the final results. In total 20,590,839 variants met the inclusion criteria. Only 5,662,641 SNPs were analyzed in the MR-MEGA analysis due to cohort-number requirements. Bonferroni-adjusted alpha was set to a more stringent 5 × 10^−9^ for all multi-ancestry meta-analyses to account for the larger number of haplotypes resulting from the ancestrally diverse datasets^9^. Genomic inflations were measured for all cohorts and the meta-analysis. Inflation for cohorts with large discrepancy between the case and control numbers were normalized to 1000 cases and 1000 controls. All inflation was nominal and below 1.02 (Supplementary Figure 3, Supplementary Table S1). No genomic control was applied prior to meta-analysis.

We identified genomic risk loci within our meta-analysis results using Functional Mapping and Annotation (FUMA) v1.3.8^11,12^. In brief, FUMA first identifies independent significant SNPs in the GWAS results by clumping all significant variants with the r^2^ threshold < 0.6, then a locus is defined by merging linkage disequilibrium (LD) blocks of all independent significant SNPs within 250kb of each other. Start and end of a locus is defined by identifying SNPs in LD with the independent significant SNPs (r^2^ ≥□0.6) and defining a region that encompasses all SNPs within the locus. The 1000 Genome reference panel with all ancestries were used to calculate the r^2^.

To determine if any associated loci in the meta-analysis were not previously identified, all significant SNPs were compared to the 92 known PD risk variants found in the previous two major meta-analyses^1,2^. Two variants identified in the Latin American admixture population^3^ could not be replicated as the variants and their proxies were removed during quality control of the discovery cohort. When a genomic risk loci was scanned for significant variants in previous European and East Asian results. If the locus contained a significant hit in either results, the locus was annotated with the closest previously nominated risk SNP within 250kb. Any loci that did not have previously significant variants or failed the annotation step was considered a novel hit.

### Fine-Mapping

Fine-mapping was performed using MR-MEGA^8^, which approximates a single-SNP Bayes’ factor in favor of association. This is reported as the natural log of Bayes’ factor (lnBF) per SNP in the MR-MEGA meta-analysis summary statistics. SNPs were selected at meta-GWAS significance level (P < 5 × 10^−9^). Posterior probabilities (PPs) of driving the association signal at each locus were calculated from the Bayes’ factor as follows:

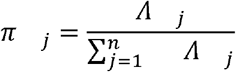

Where *⋀*_*j*_ is the Bayes’ factor of the *j*th SNP within a locus with *n* number of SNPs. Credible set of SNPs with PP (*π*_*j*_) greater than 0.99 were accepted as putative causal variants.

### Putative variant burden estimation by population

Proportion of population-specific and shared causal variants (PESCA)^10^ was used to estimate the population-specific and shared burden of the variants tested in the meta-analysis. In brief, genome-wide heritability was estimated for the European and East Asian GWAS summary statistics using LD Score regression (LDSC)^37,38^. Summary statistics of both populations were intersected with common variants with the 1000 Genome reference panels provided by PESCA, which have already been LD pruned (R^2^ > 0.95) and low frequency SNPs removed (minor allele frequency < 0.05). The intersected variants were further split according to independent LD regions from the European and East Asian populations. The genome-wide prior probabilities of population-specific and shared causal variants were calculated using default parameters or as otherwise recommended by PESCA, then the results were used to calculate the PP for each variant. When the lead SNP was unavailable in the results, proxy variants (r^2^ > 0.8) were used to approximate the PP for each variant. Other cohorts were not included due to sample size constraints for this method.

### Functional Annotation

Functional annotation of the discovery results utilizing publicly available gene expression and ontology data was done using FUMA v1.3.8^11,12^. The summary statistics were annotated by ANNOVAR^39^ through the FUMA platform. Our meta-analysis results were analyzed using Multi-marker Analysis of GenoMic Annotation (MAGMA)^13^ to compare gene expression data from gene sets and tissues in GTEx v8^40^. FUMA tested 15,479 gene sets and gene ontology terms from MsigDB v7^41^ as well as single-cell RNAseq expression data from mouse brain samples in DropViz^18^ and human ventral midbrain samples^19^. Test parameters were set to default. Results were adjusted for multiple tests using Benjamini–Hochberg False Discovery Rate (FDR) correction with the alpha of 0.05. Additional pathway analyses of genes mapped by FUMA SNP2GENE were performed through GENE2FUNC with default parameters.

SNPs in the novel loci were searched in multi-ancestry brain eQTL meta-analysis results^14^. We used a P-value cutoff of 10^−6^ as previously described^14^. eQTL and GWAS comparison plots were generated using LocusCompare^42^. Multi-SNP Summary-Based Mendelian Randomization (SMR) was used to test if DNA methylation and/or RNA expression of genes near the novel loci were associated with PD risk^15^. The nearest genes from the lead SNPs, significant genes in the multi-ancestry meta-analysis brain eQTL results, and significant genes in GTEx v8 brain tissue were chosen for SMR. In total 44 genes near the novel loci were searched in a list of previously completed PD SMR results^15,43–51^. The association signals were adjusted using FDR correction with the alpha of 0.05 and all signals with P_HEIDI_ > 0.05 were removed due to heterogeneity.

