## Supplementary Tables and Figures for "Multi-ancestry genome-wide meta-analysis in Parkinson’s disease": MAMA_PD-Supplemental Figures 1-7,9.docx

### Supplementary Figures

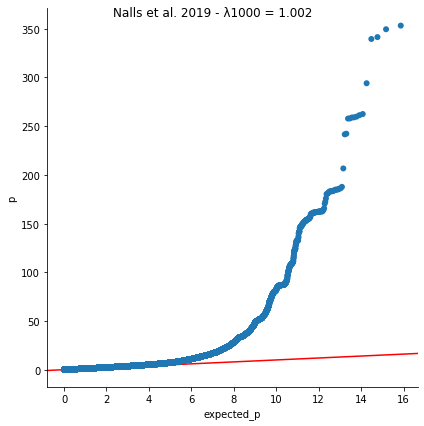

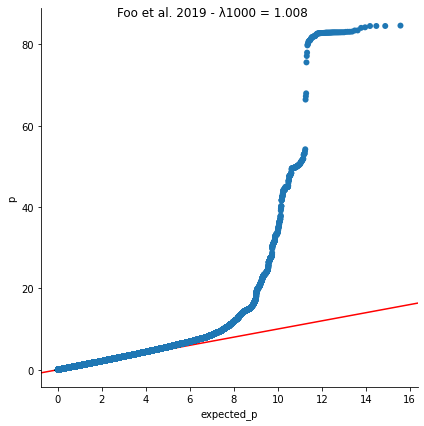

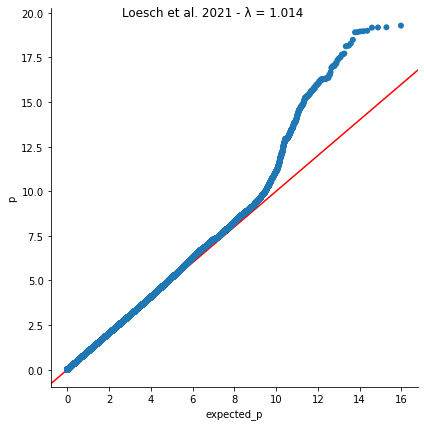

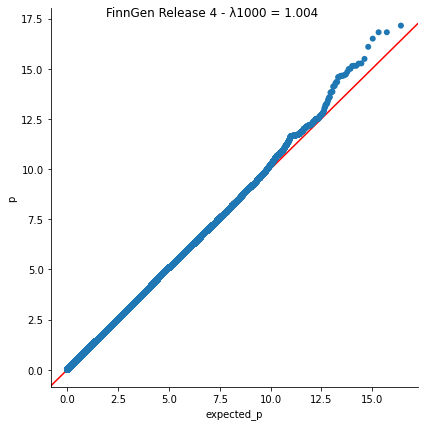

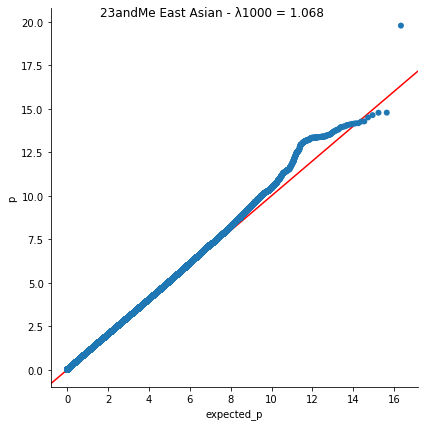

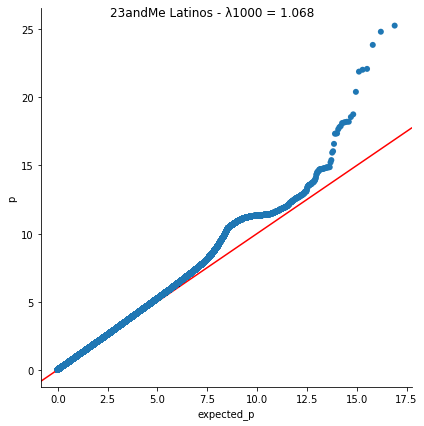

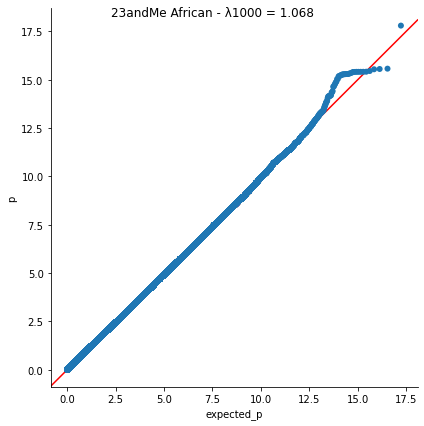

Supplementary Figure 1 – QQ Plots and Genomic Inflation (λ) of each ancestry cohorts. Genomic inflation was normalized to 1000 cases and 1000 controls for all datasets outside of Loesch et al 2021 due to large discrepancy between number of cases and controls.

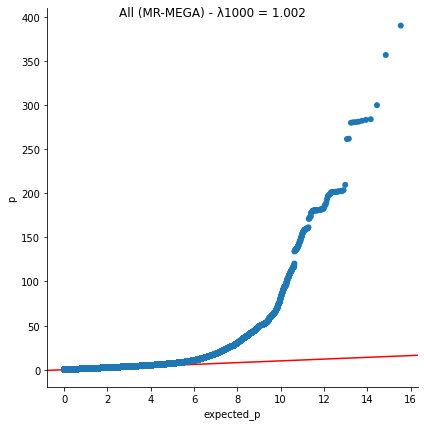

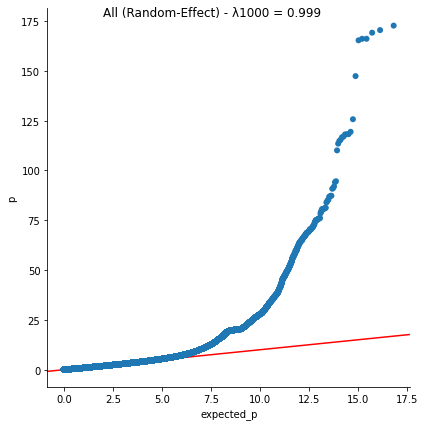

Supplementary Figure 2 – QQ Plots and normalized Genomic Inflation (λ) of the two meta-analyses. Genomic inflation was normalized to 1000 cases and 1000 controls for all datasets due to large discrepancy between number of cases and controls.

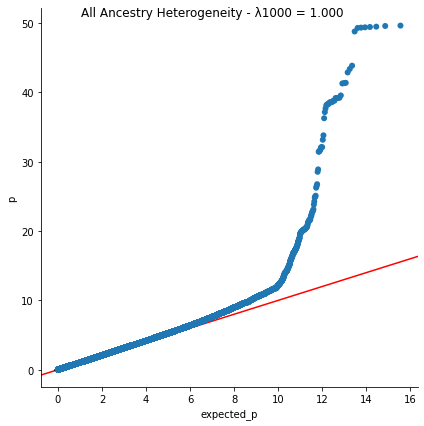

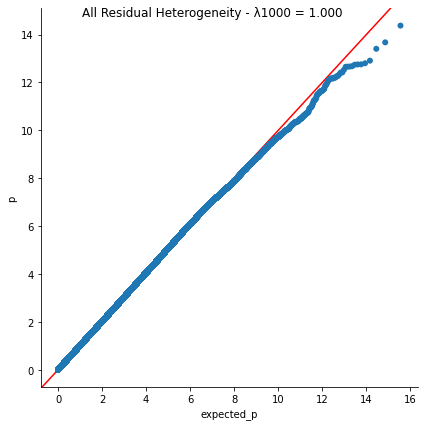

Supplementary Figure 3 - QQ Plot and Genomic Inflation of MR-MEGA ancestry heterogeneity and residual heterogeneity. Genomic inflation was normalized to 1000 cases and 1000 controls.

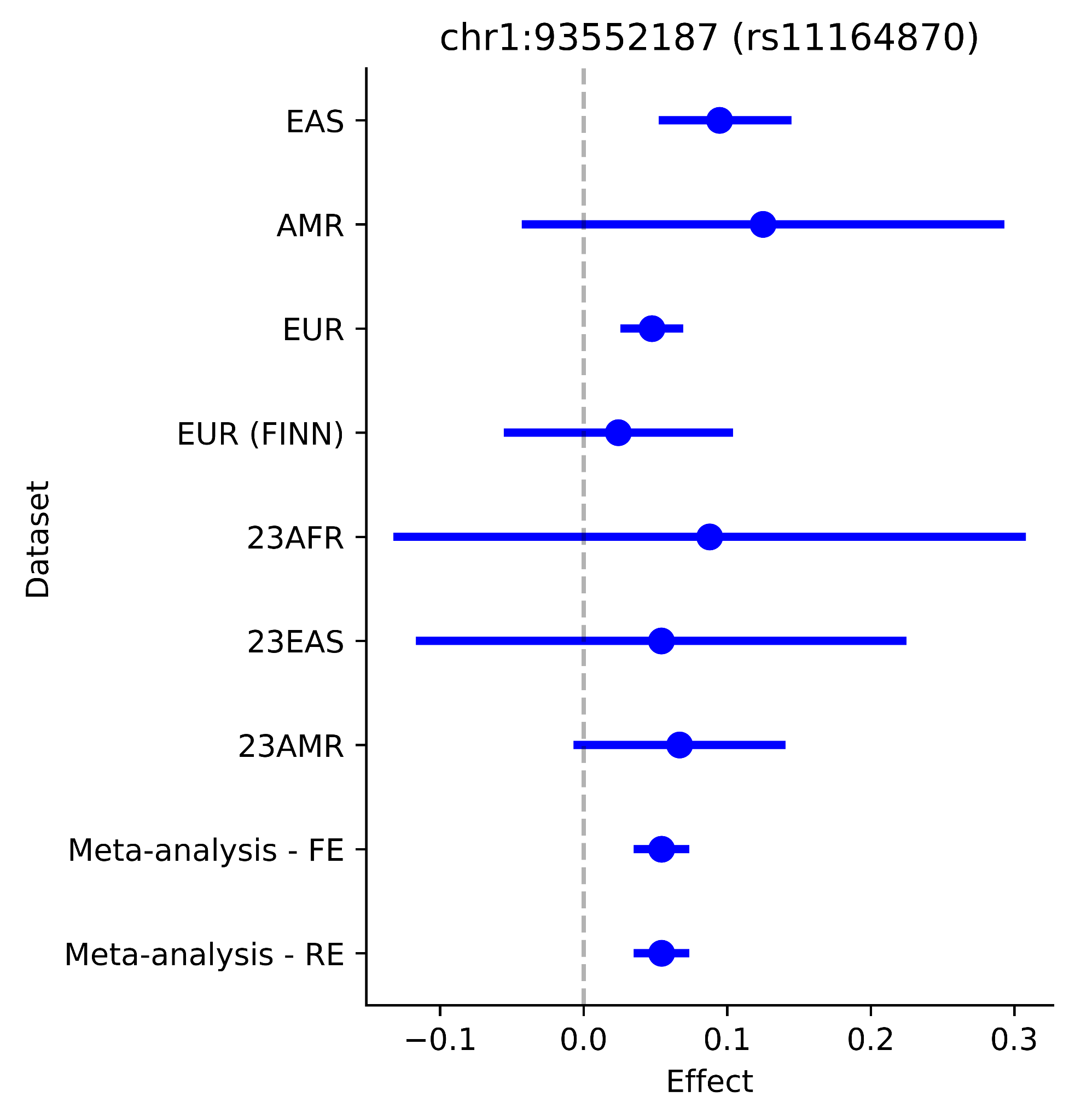

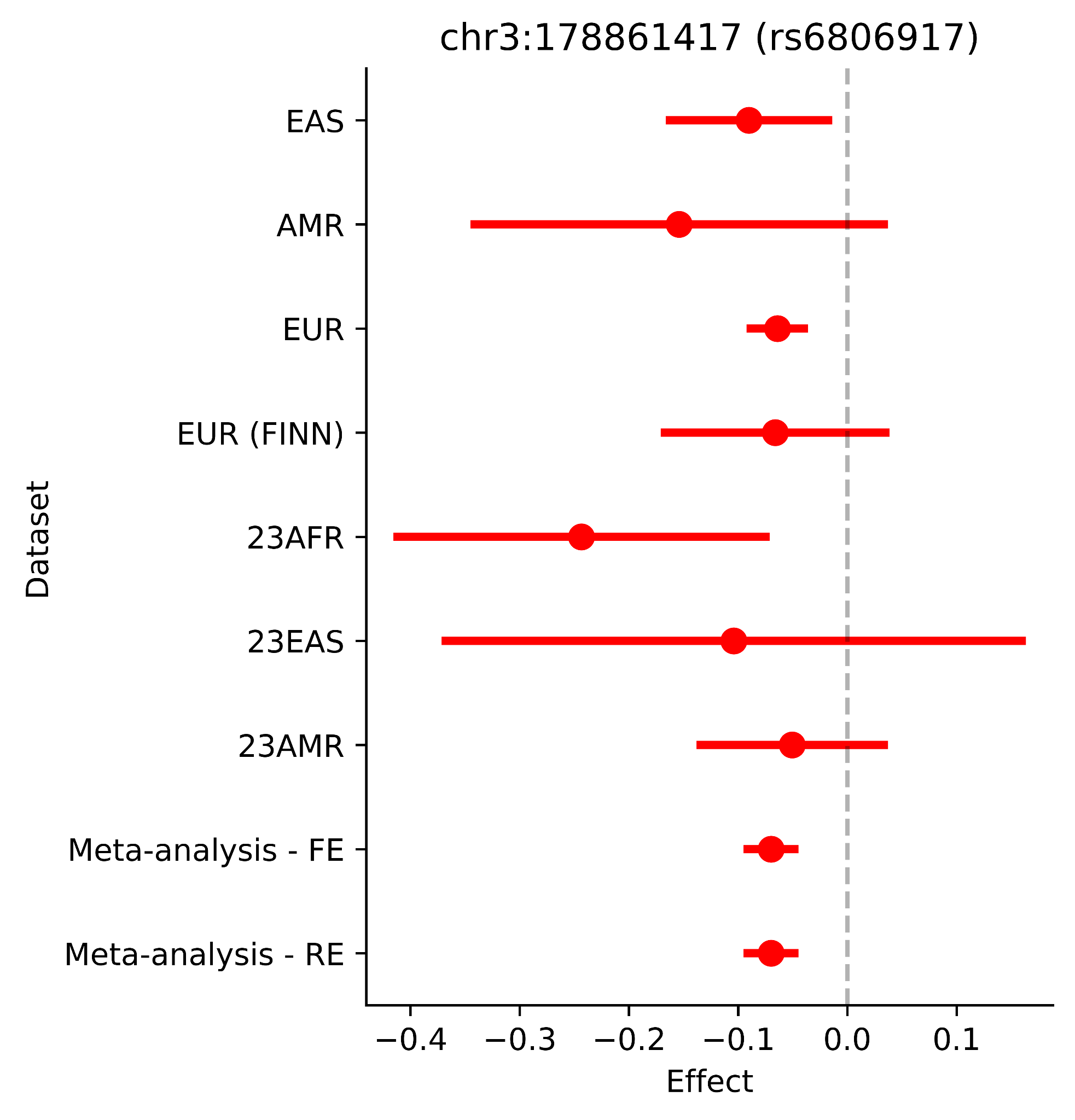

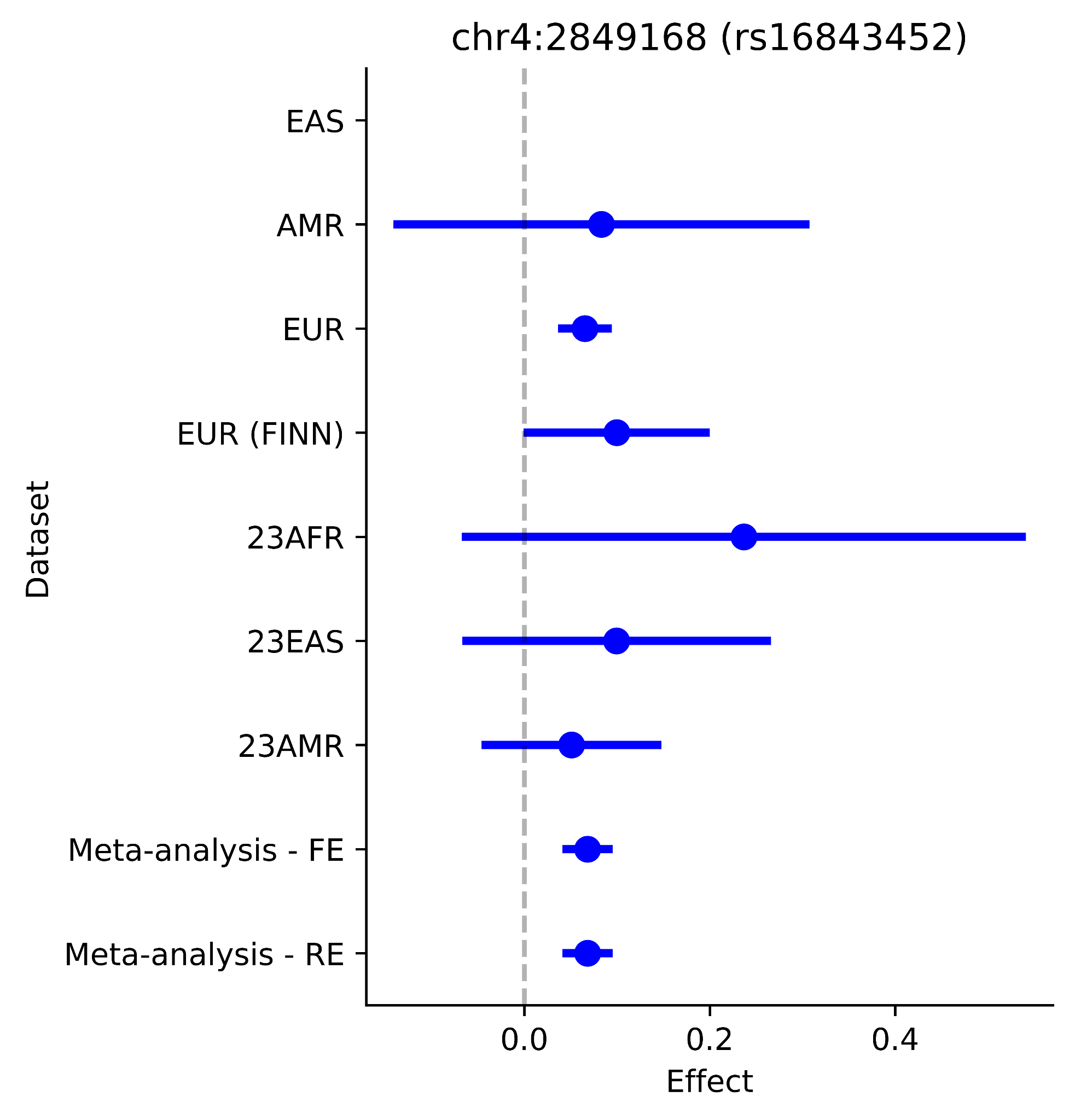

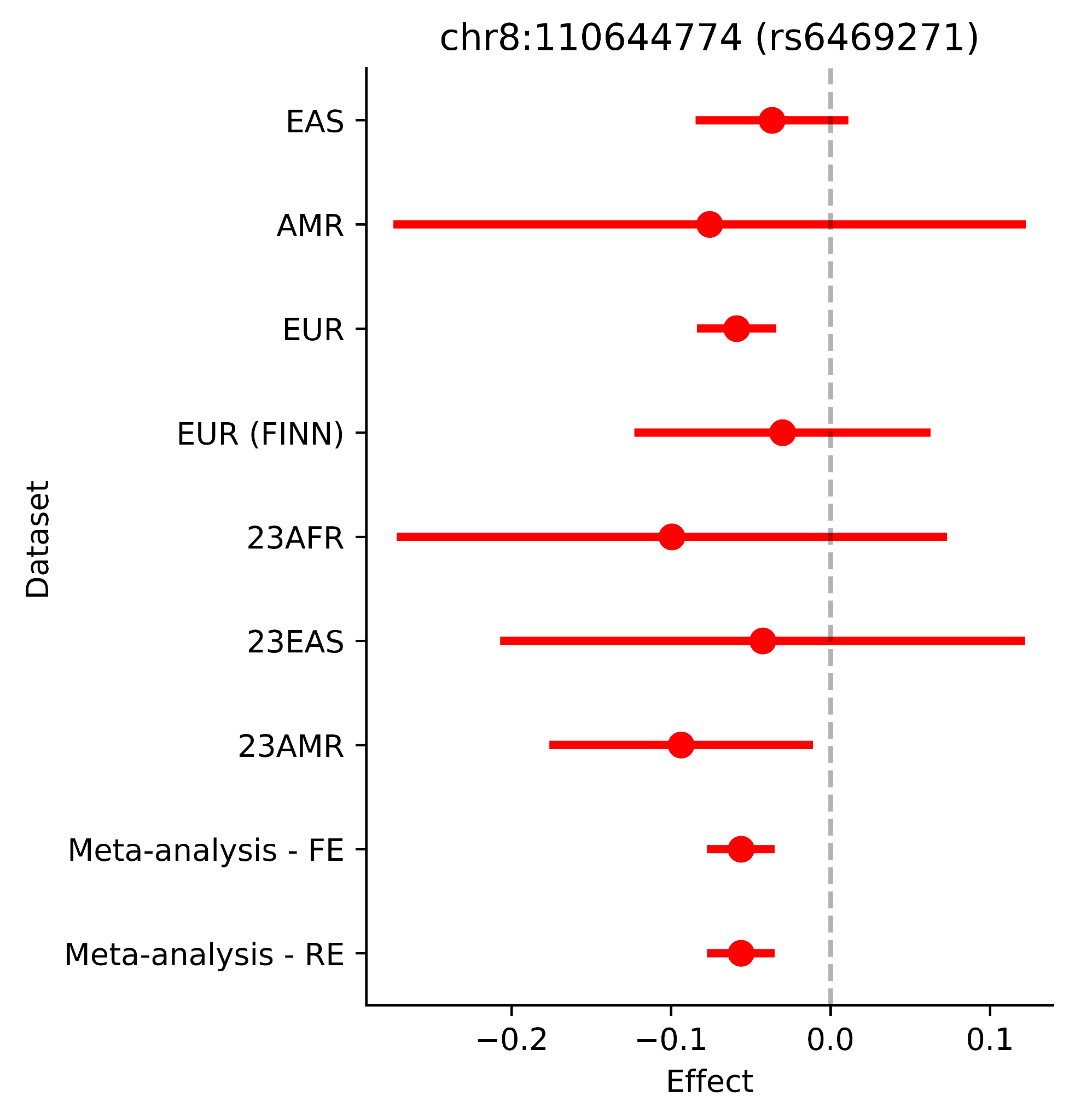

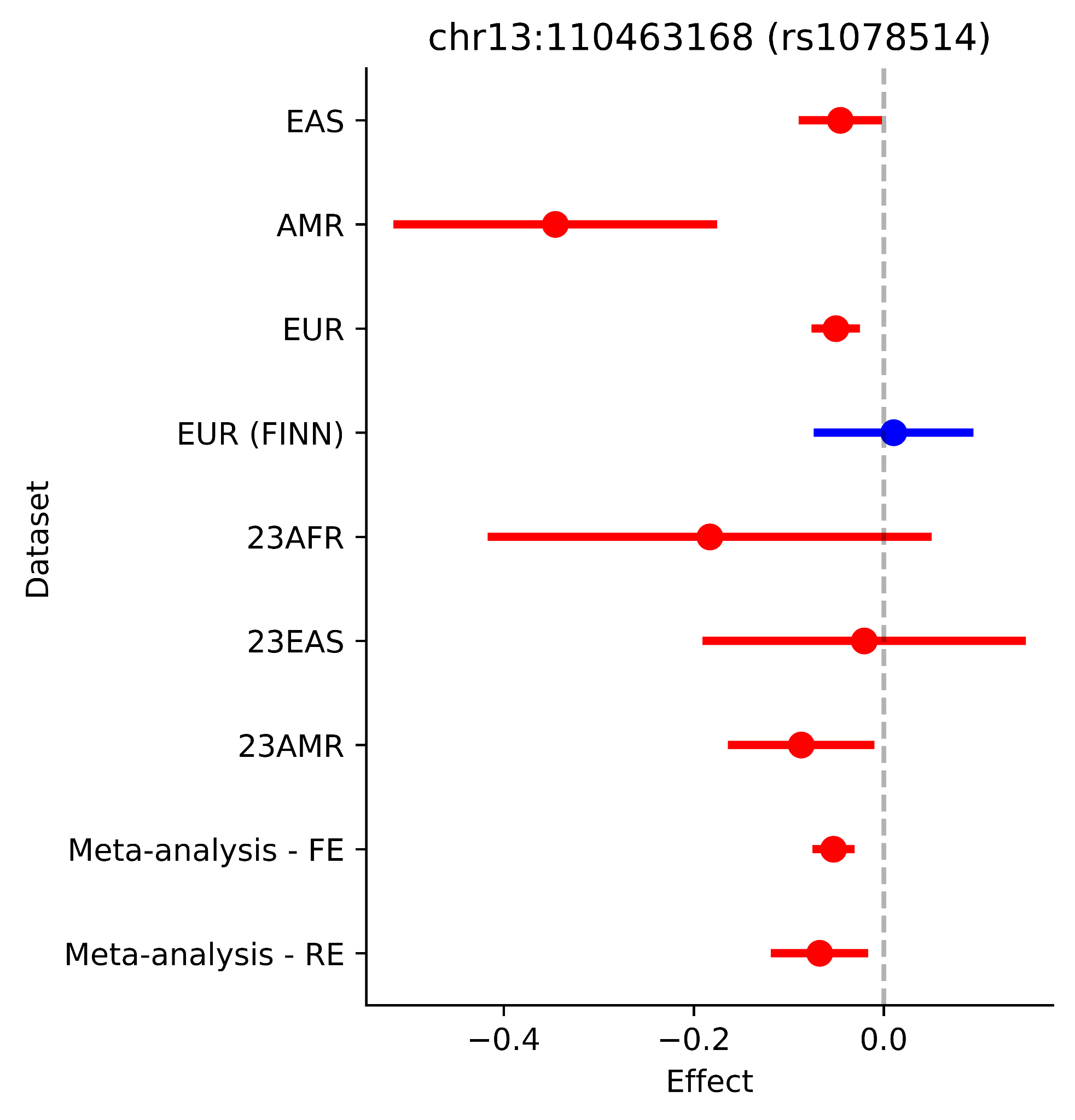

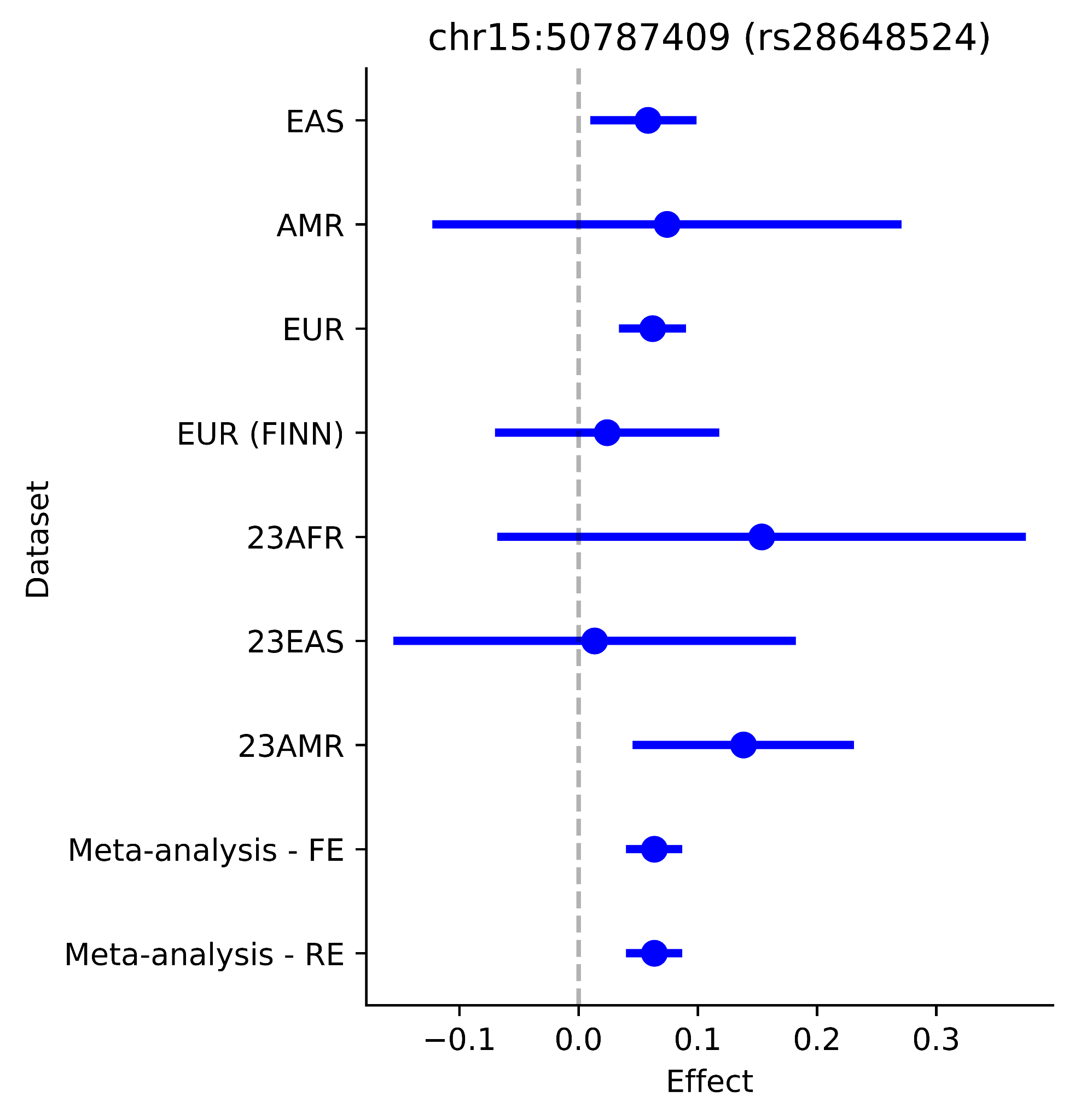

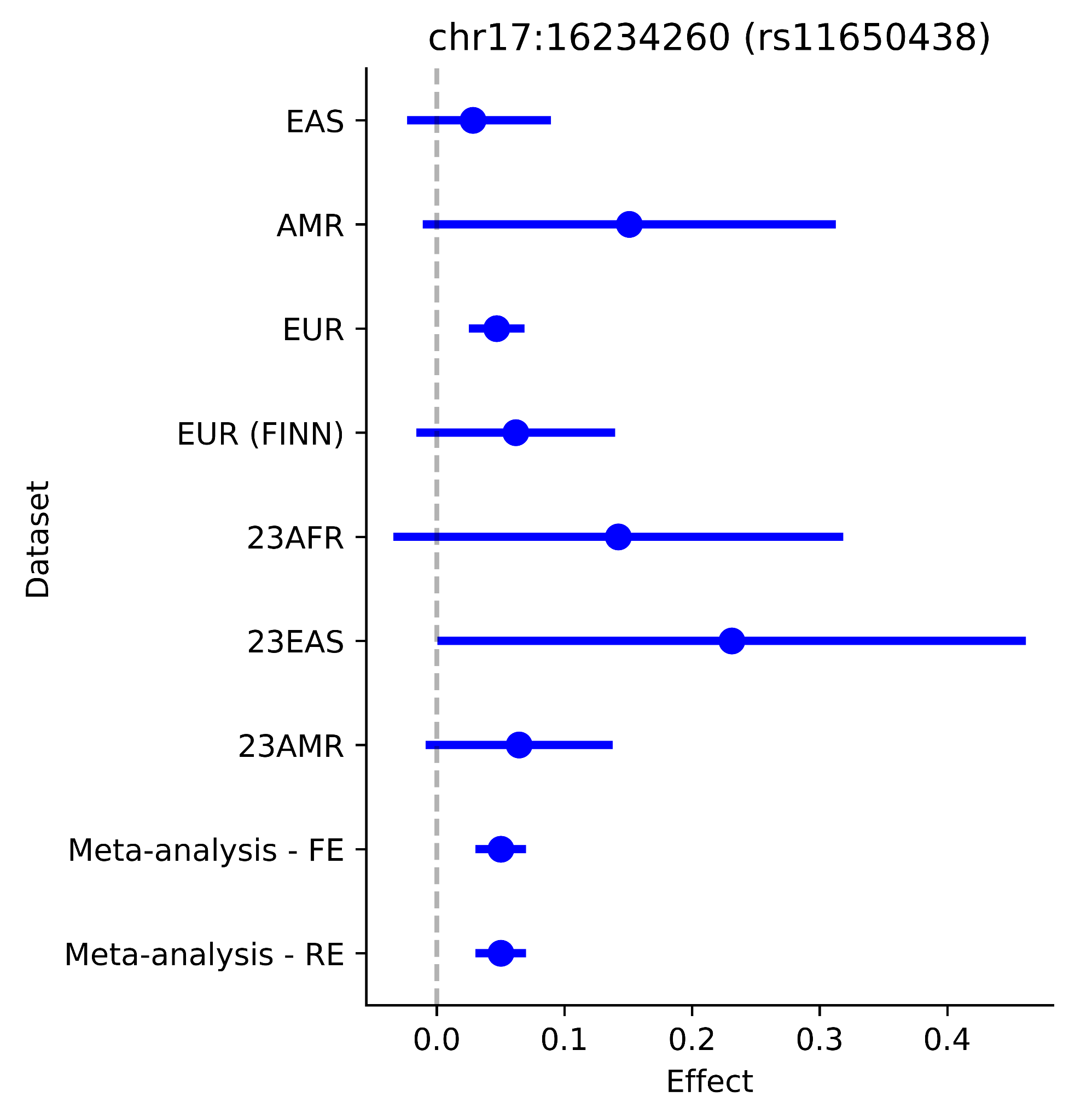

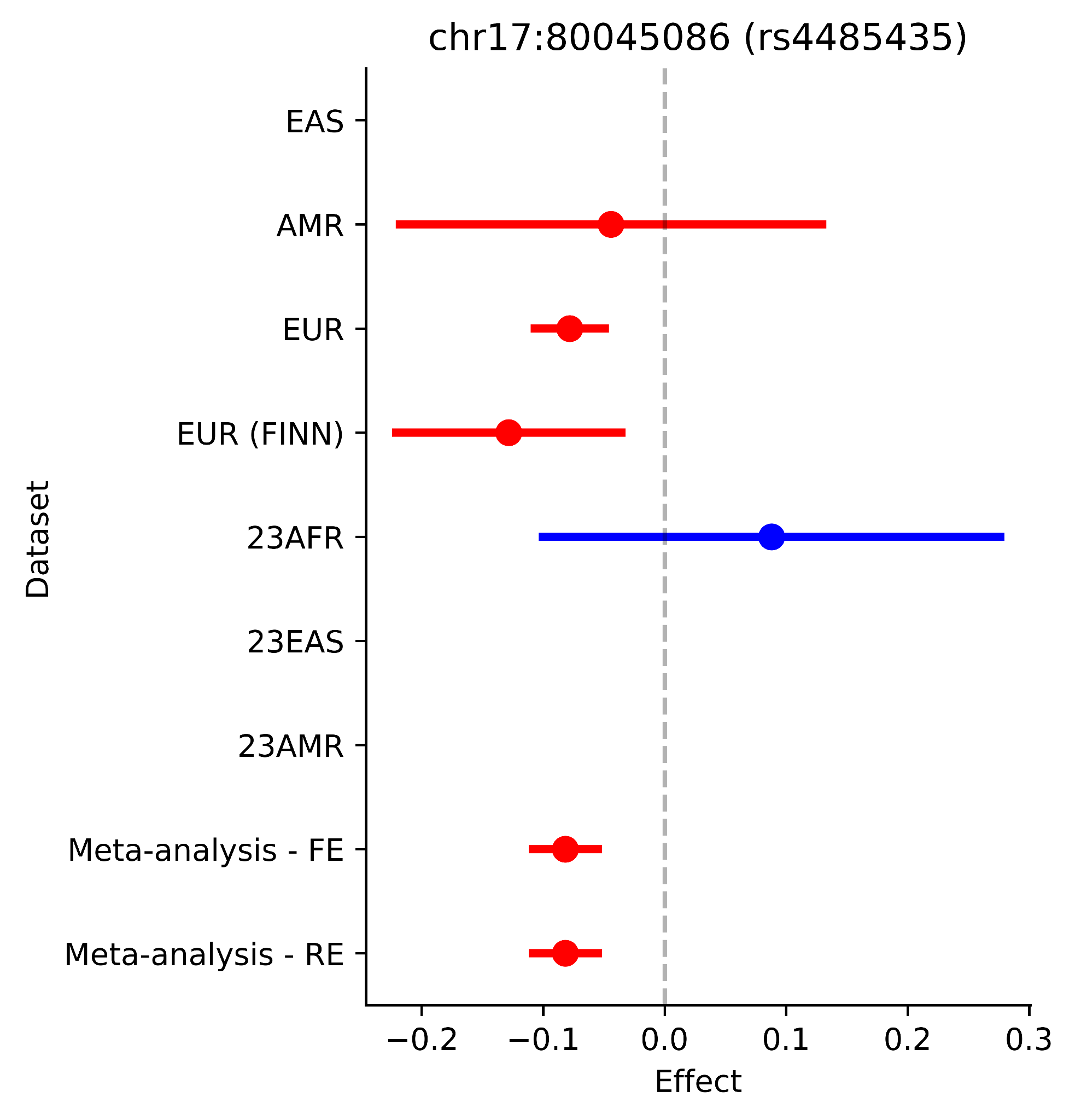

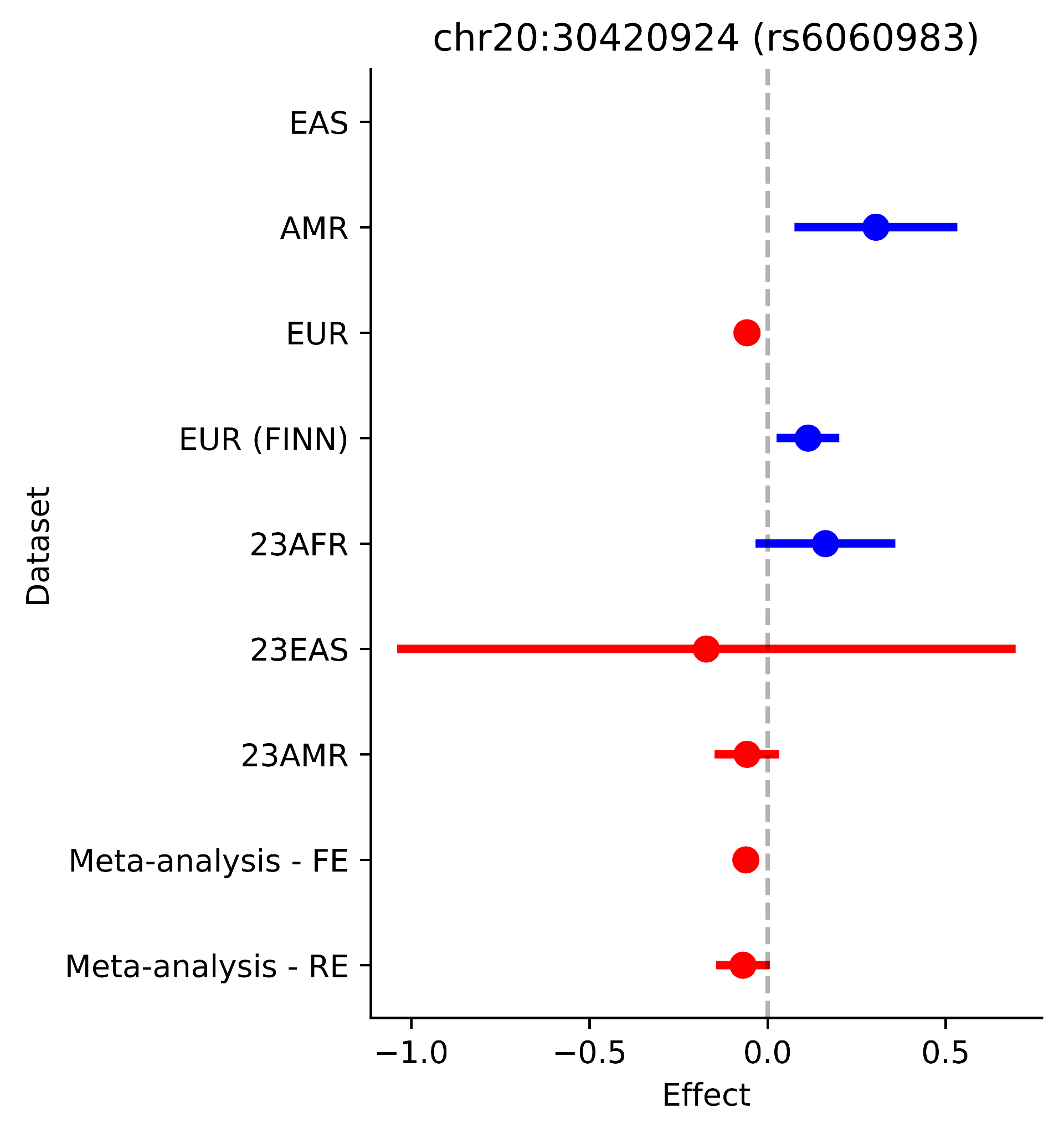

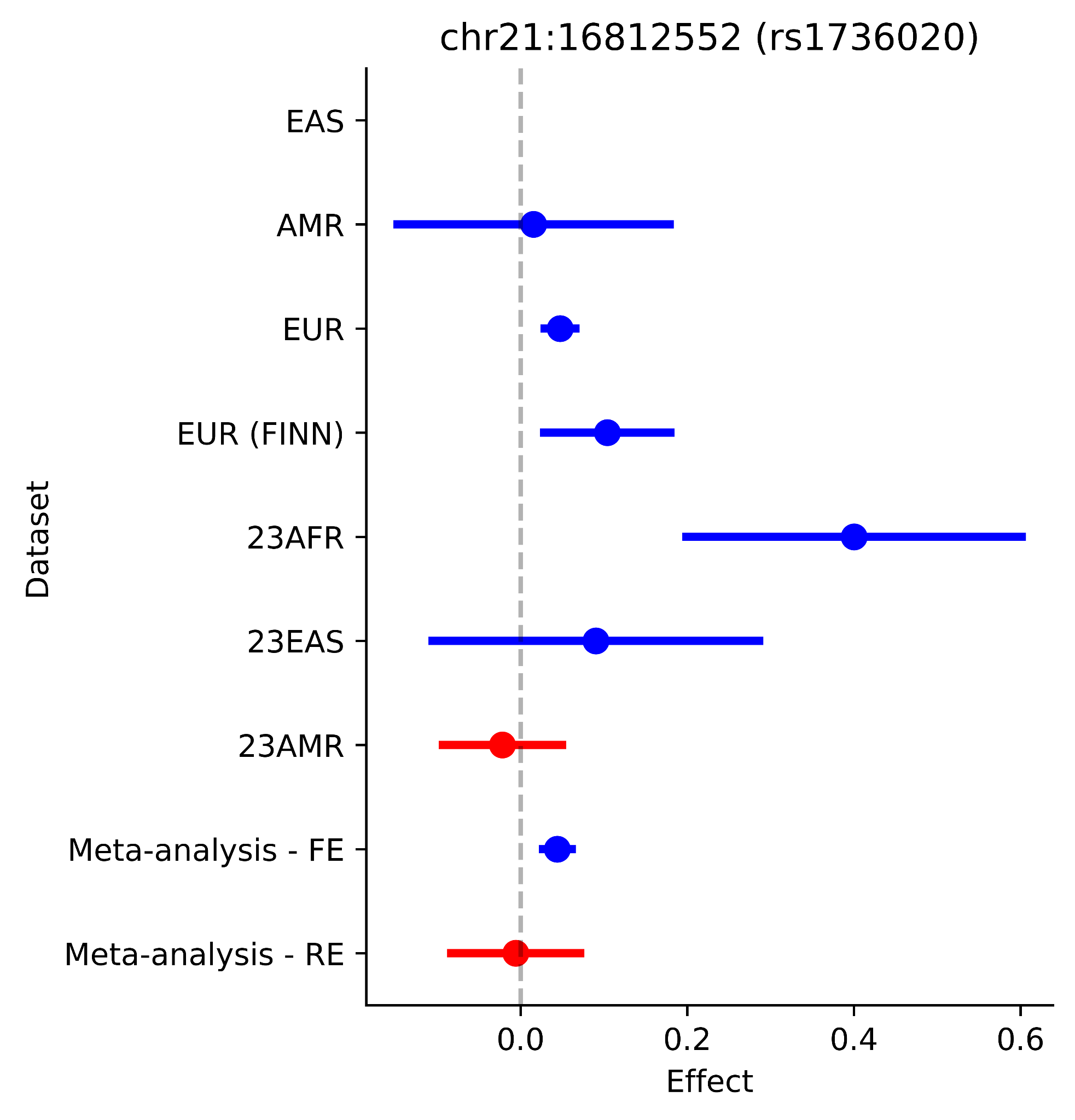

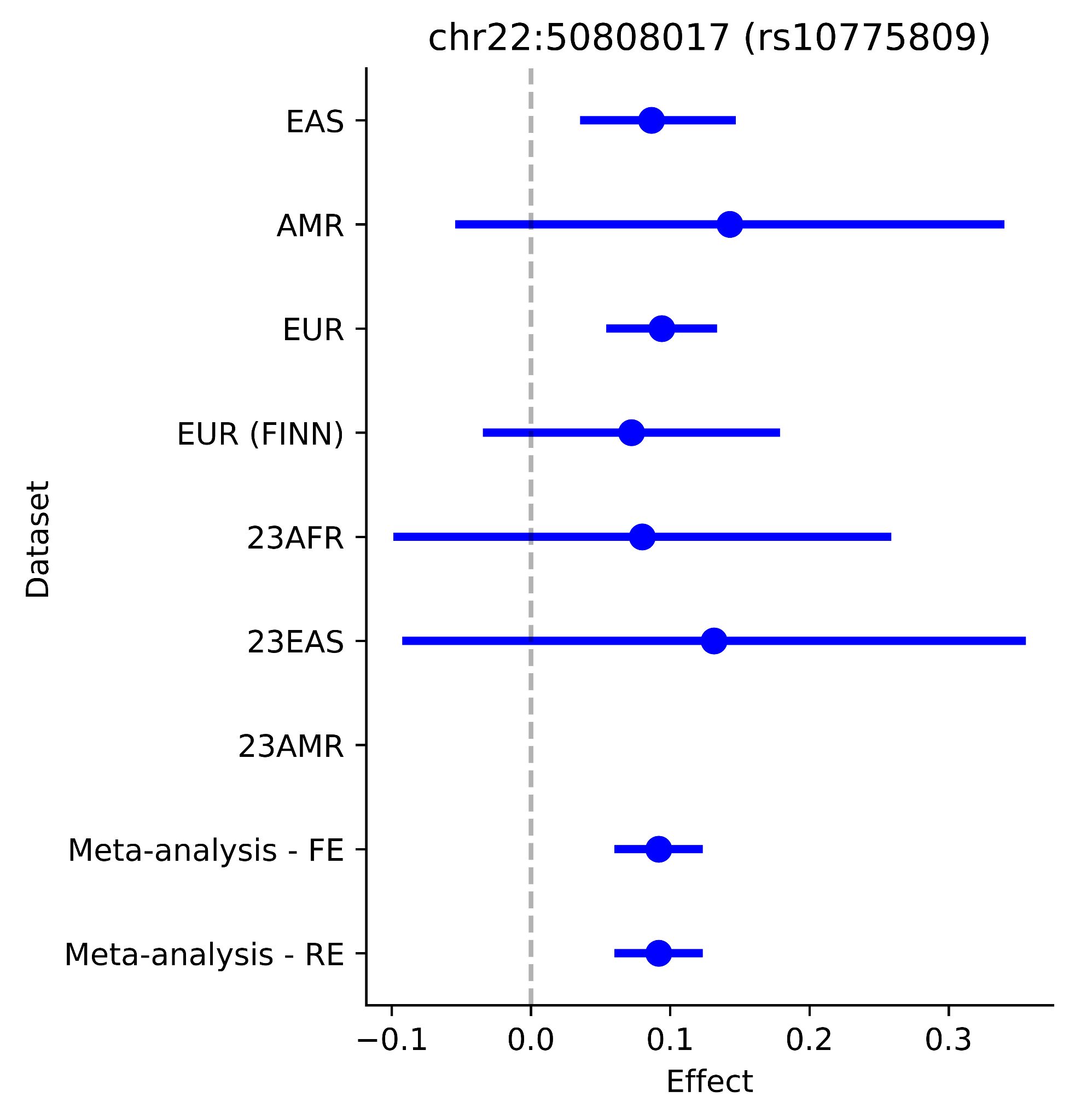

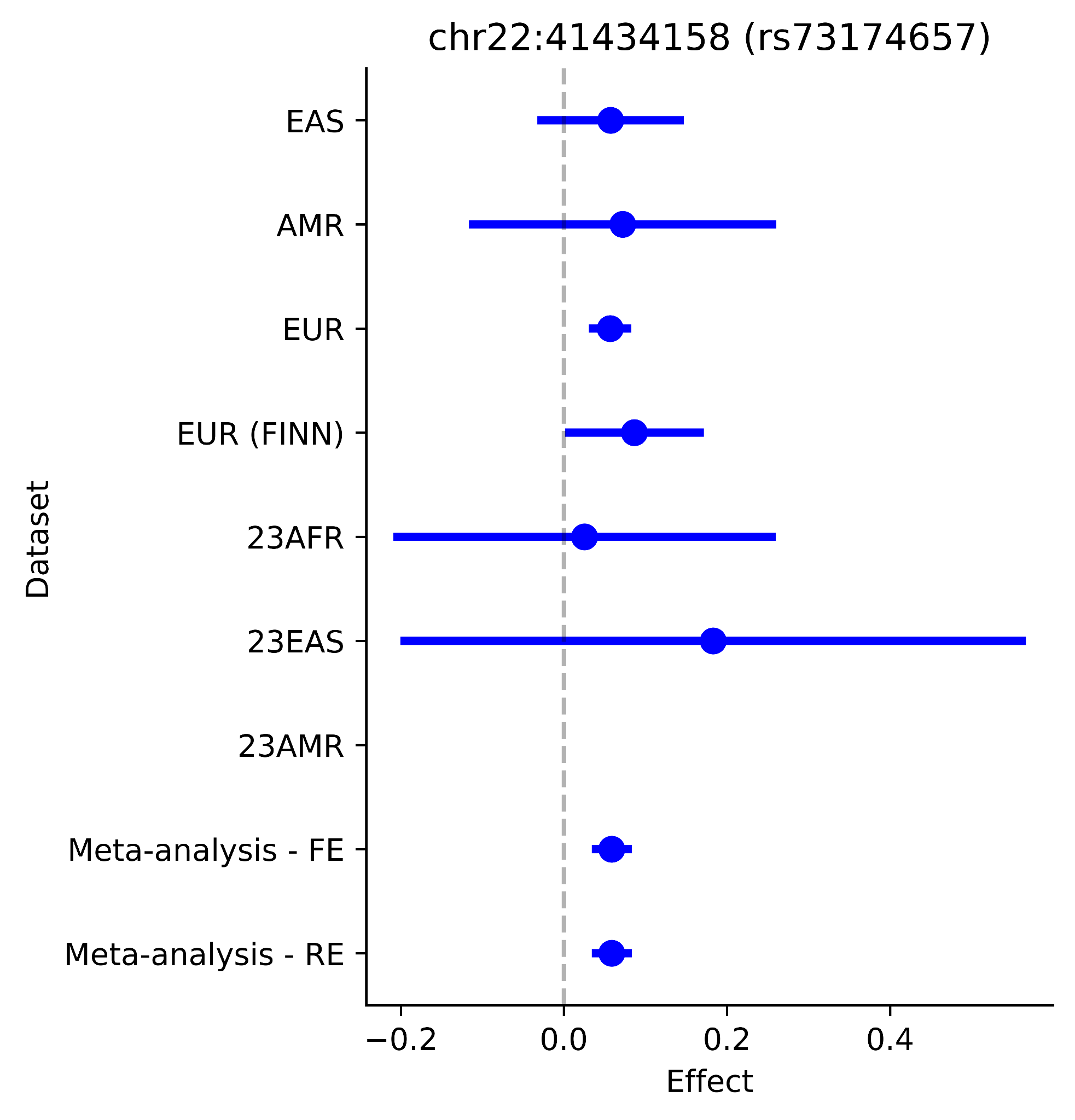

Supplementary Figure 4 - Forest plots of the lead SNPs of novel loci in the ancestrally distinct cohorts and the random effect meta-analysis results. MR-MEGA results were not represented as they do not provide effect direction or size. Red represents negative effect direction and blue represents positive effect direction

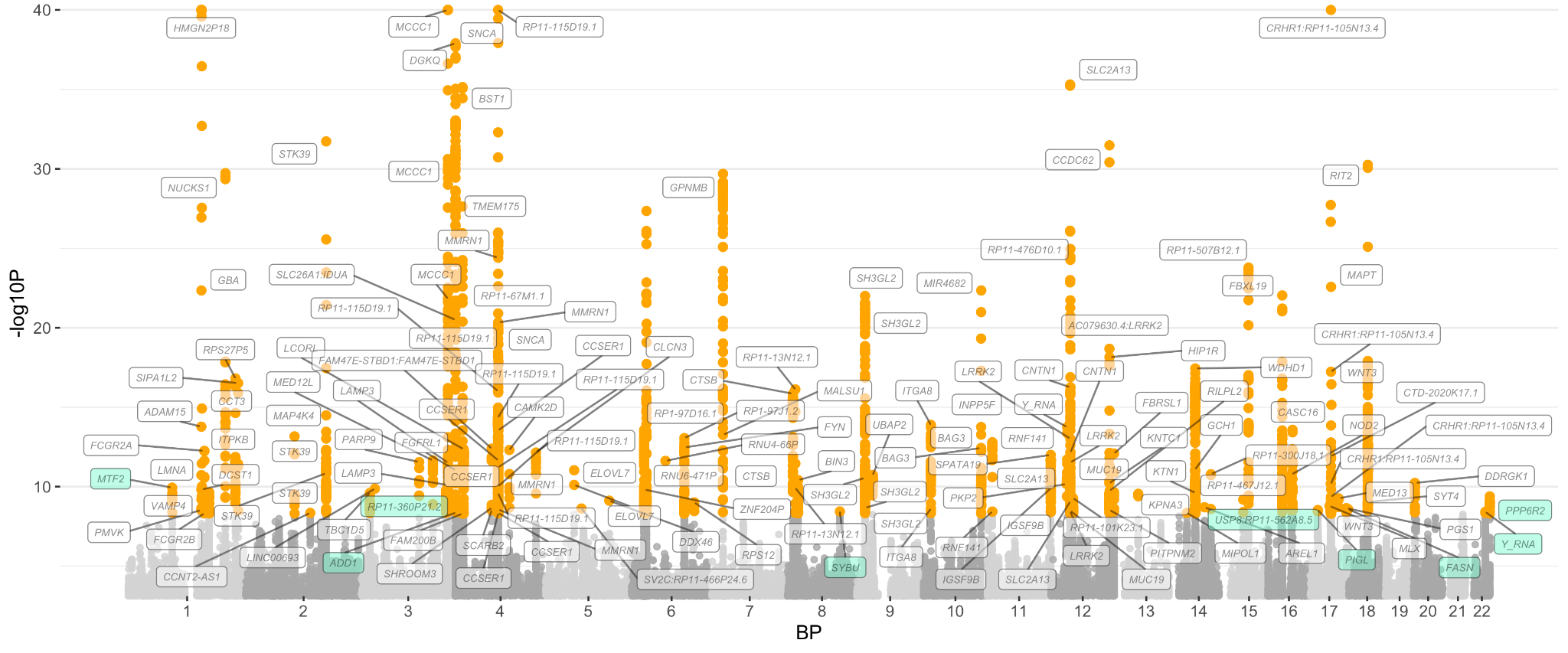

Supplementary Figure 5 - Manhattan plot of the random effect meta-analysis result. Orange points indicate a significant variant at P < 5 x 10^-9^. The lead SNPs are annotated with the nearest gene, with those in novel loci highlighted in green.

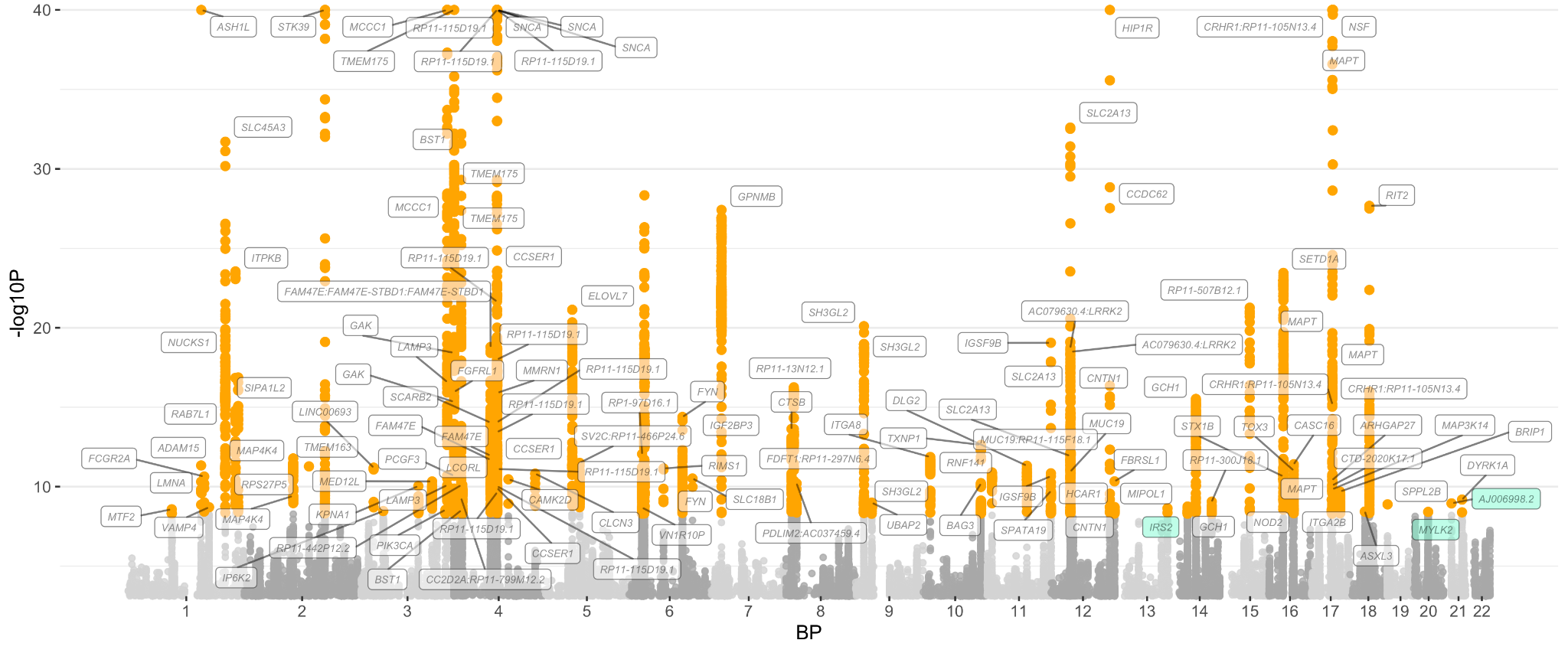

Supplementary Figure 6 - Manhattan plot of the MR-MEGA meta-analysis result. Orange points indicate a significant variant at P < 5 x 10^-9^. The lead SNPs are annotated with the nearest gene, with those in novel loci unique to MR-MEGA highlighted in green.

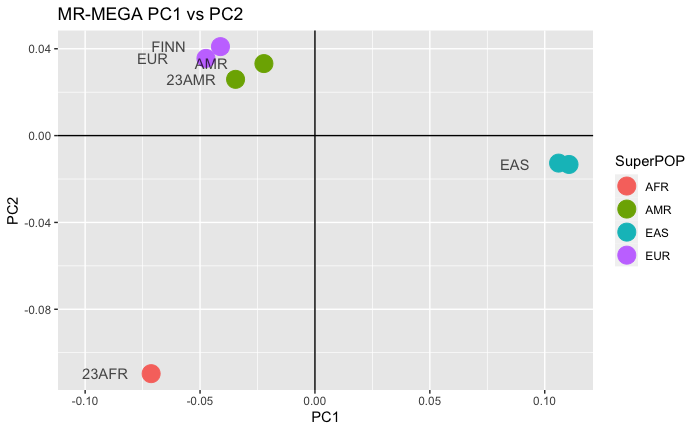

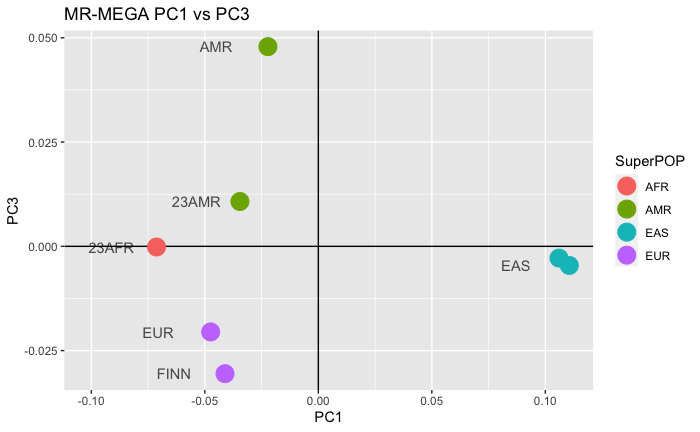

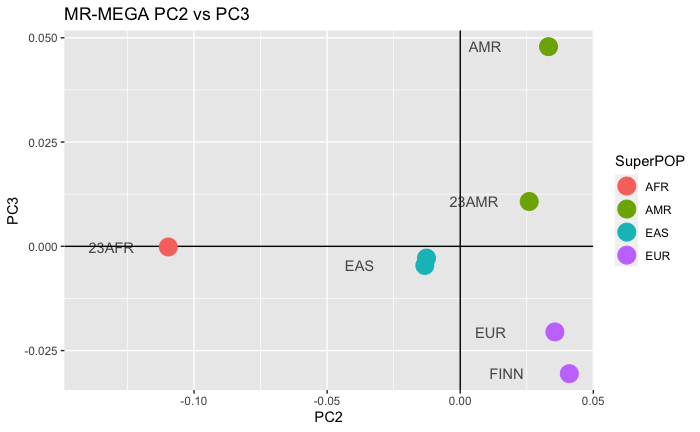

Supplementary Figure 7 - Principal Component of Ancestry Heterogeneity from MR-MEGA.

[See in folder]

Supplementary Figure 8 - LocusCompare Plots of novel loci comparing PD MAMA association and brain eQTL results. Left is the PP plot between the PD MAMA GWAS results and eQTL results. Right are locuszoom plots, with the GWAS results on top and eQTL results on the bottom. Violet diamond denotes the lead SNP, determined by summing the p-values of the two studies and selecting the lowest sum. Linkage disequilibrium reference is based on the 1000 Genome dataset, EUR populations. Brain eQTL data is from the meta-analysis study by Zeng et. al 2022. The plots only include eQTL order 1 to improve visibility.

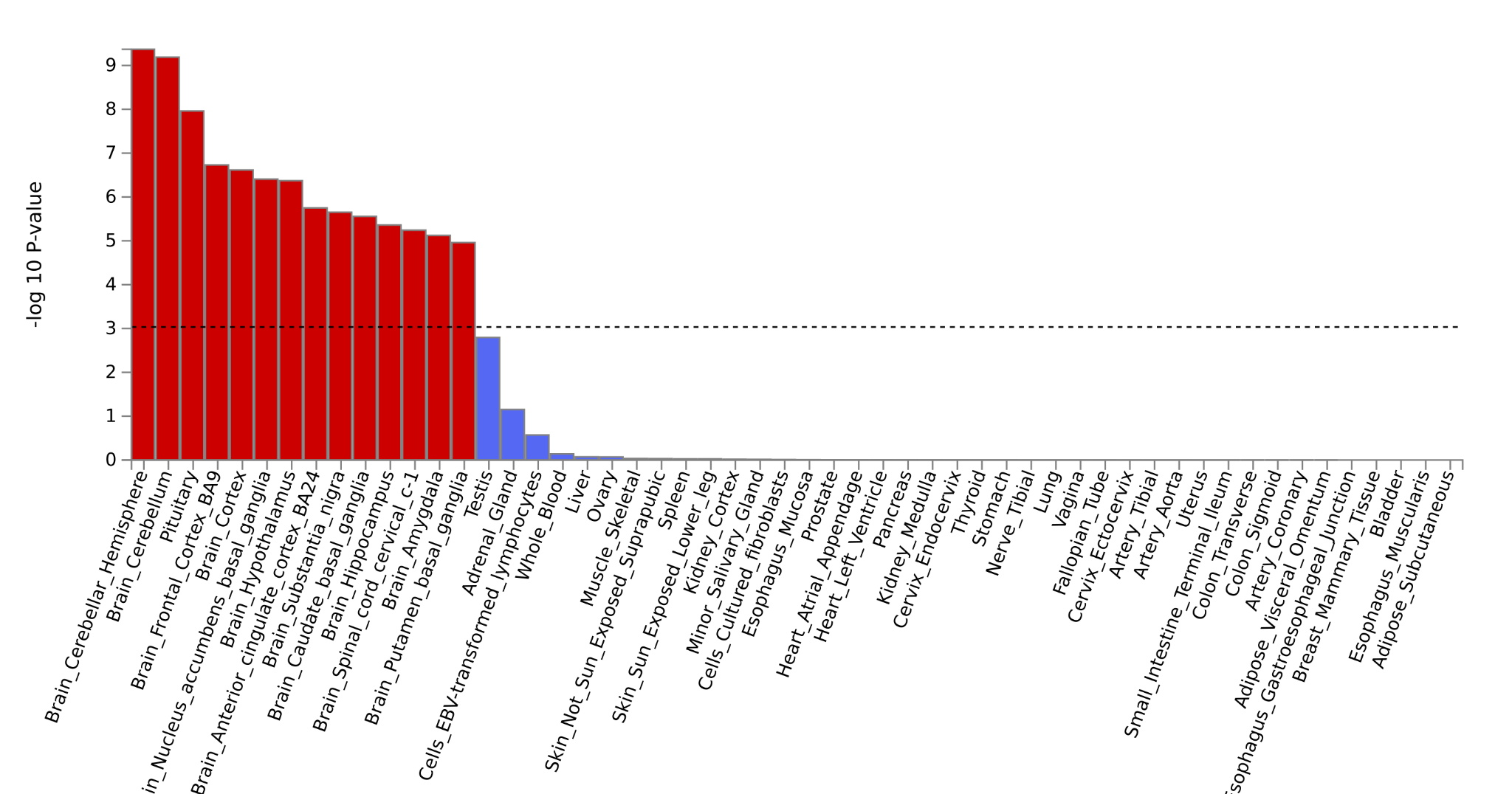

Supplementary Figure 9 - Expression enrichment by tissue from FUMA. FUMA analysis using GTEx v8 is consistent with the published summary statistics from Nalls et al. 2019.

[See in folder]

Supplementary Figure 10 - FUMA GENE2FUNC analysis of FUMA SNP2GENE mapped genes. The plots show enrichment analysis in A) biological processes B) cellular components C) molecular function D) GWAS Catalog
