## Supplementary figures and images for "Multi-ancestry genome-wide meta-analysis in Parkinson’s disease"

### ADORA2B.png

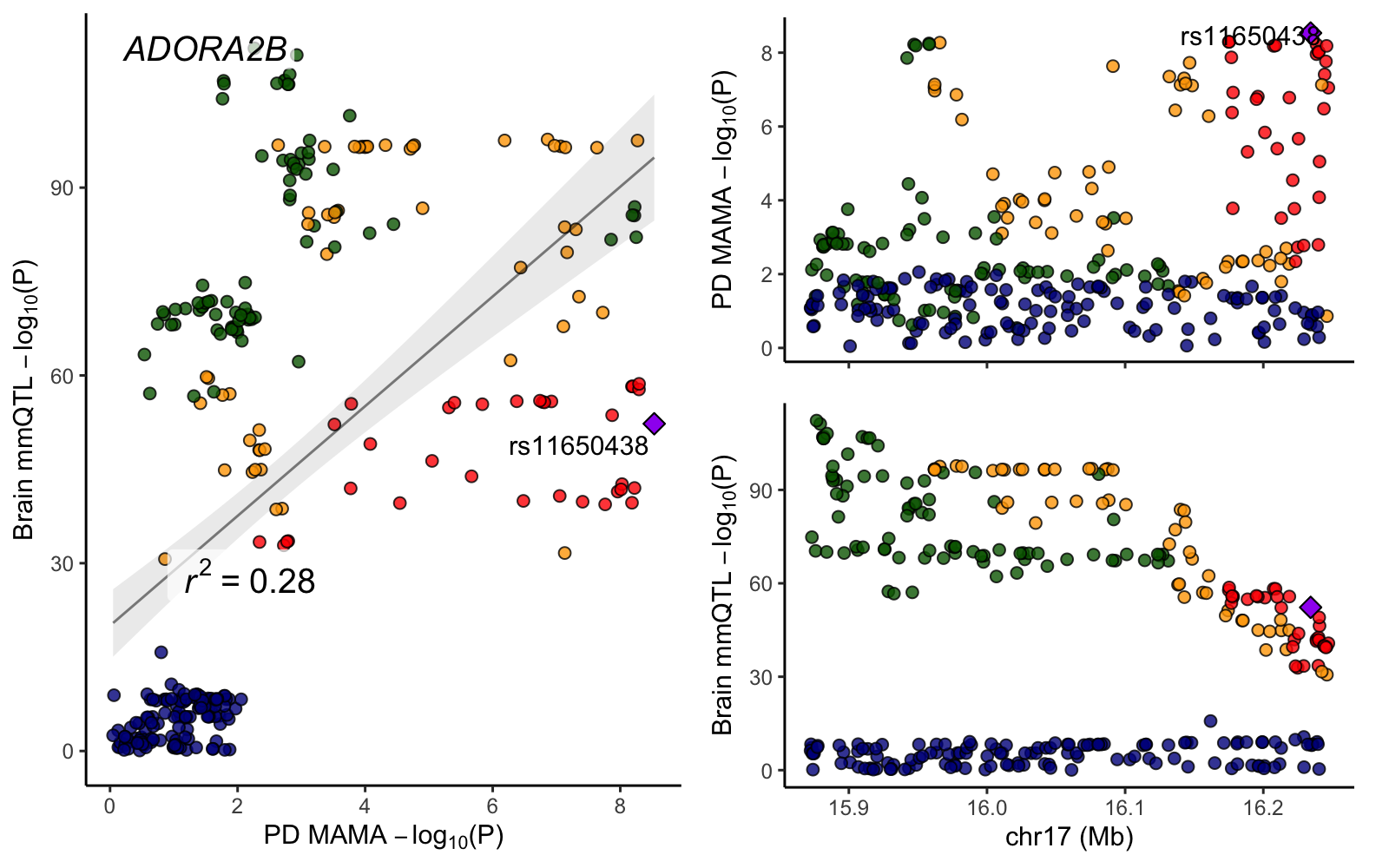

### CCDC18-AS1.png

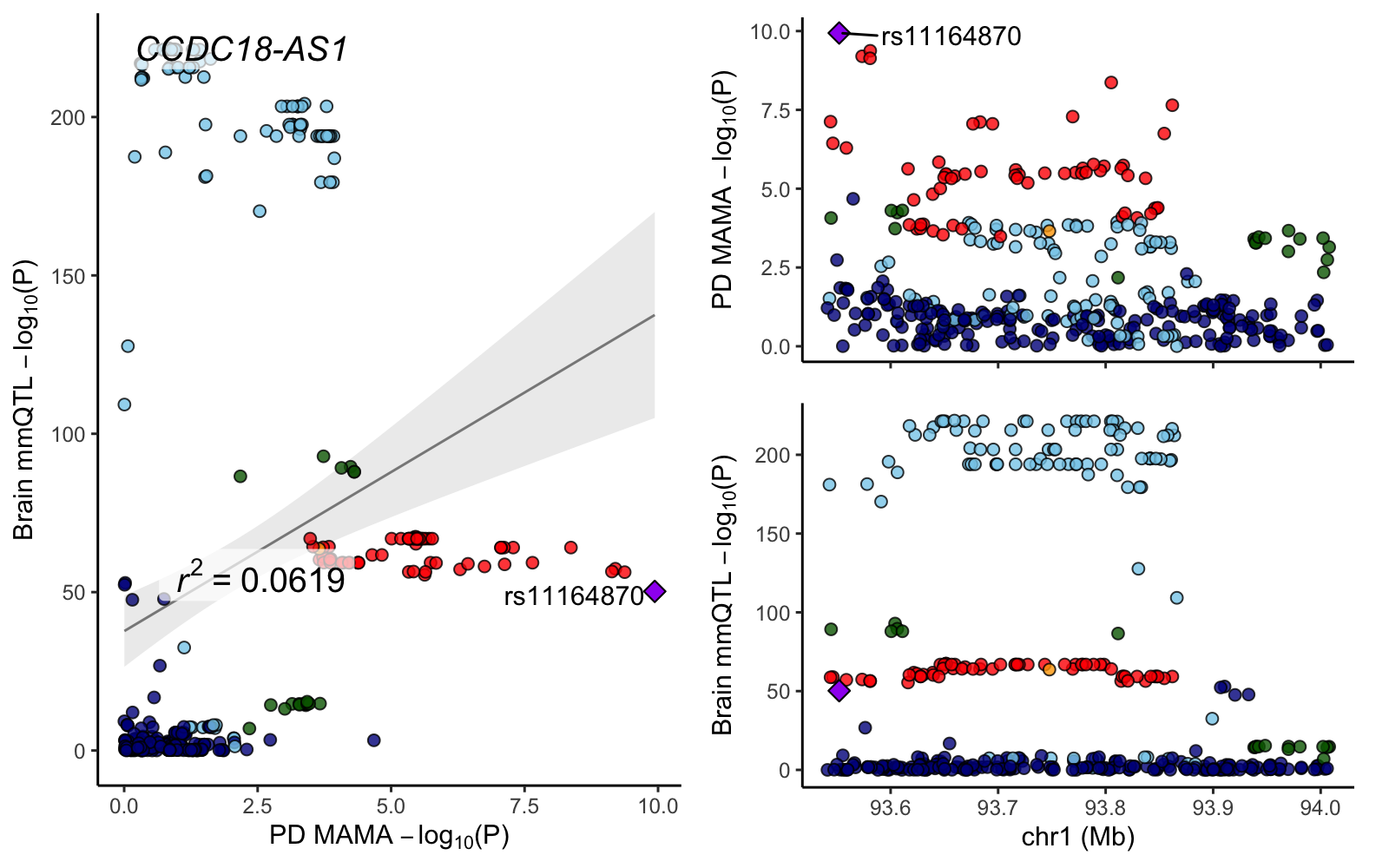

### CCDC18.png

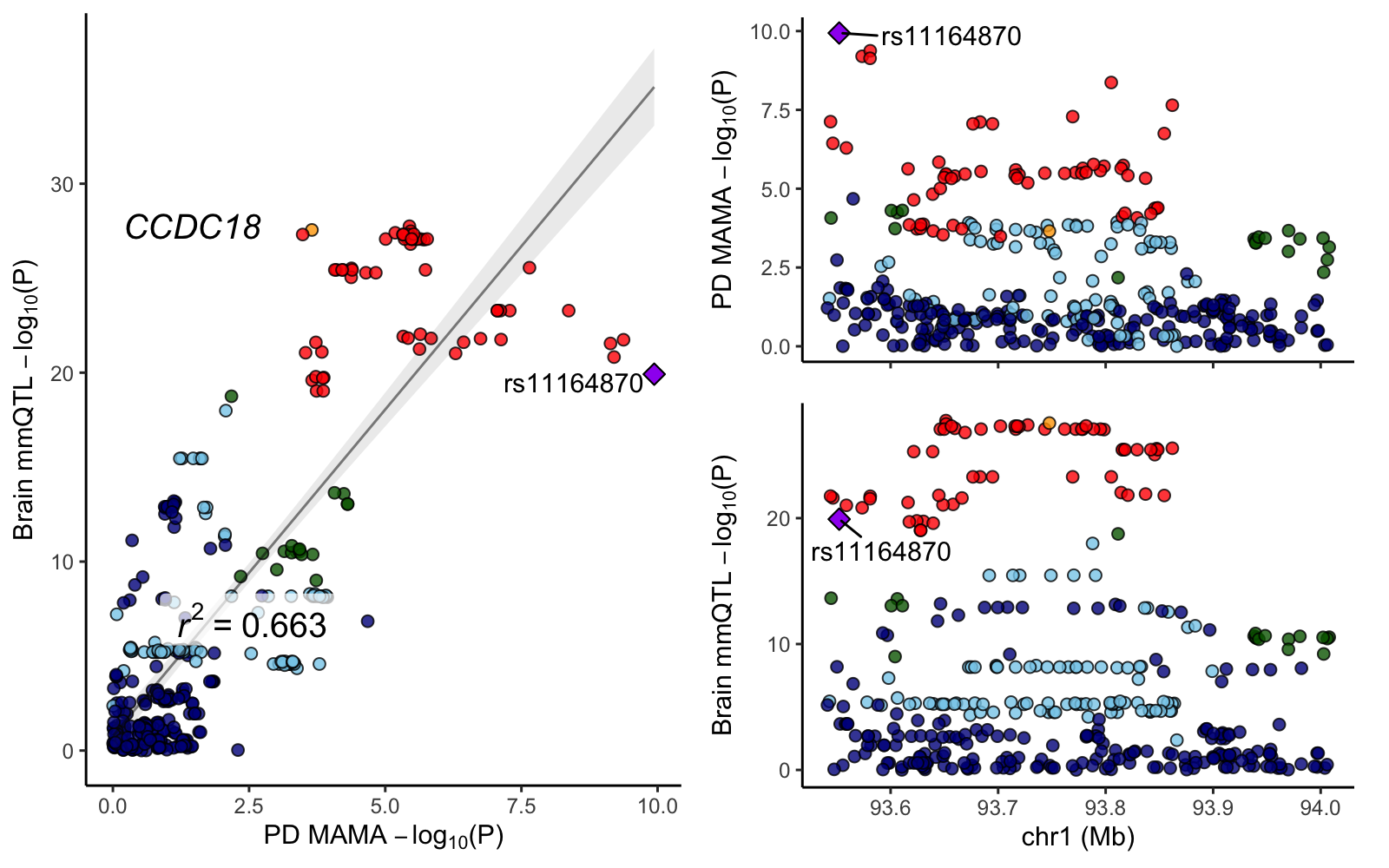

### CENPV.png

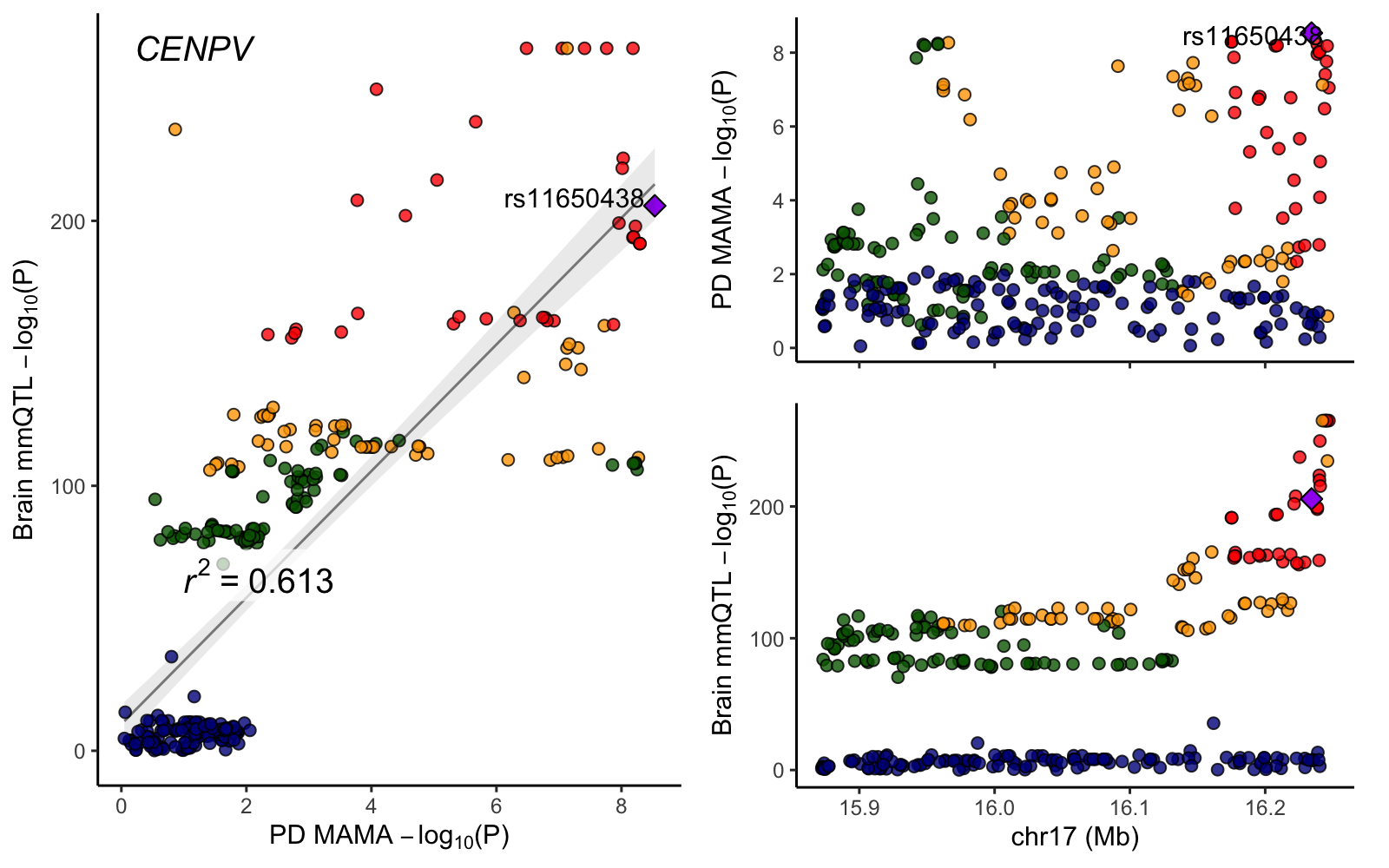

### chr2_168860394-169360394.png

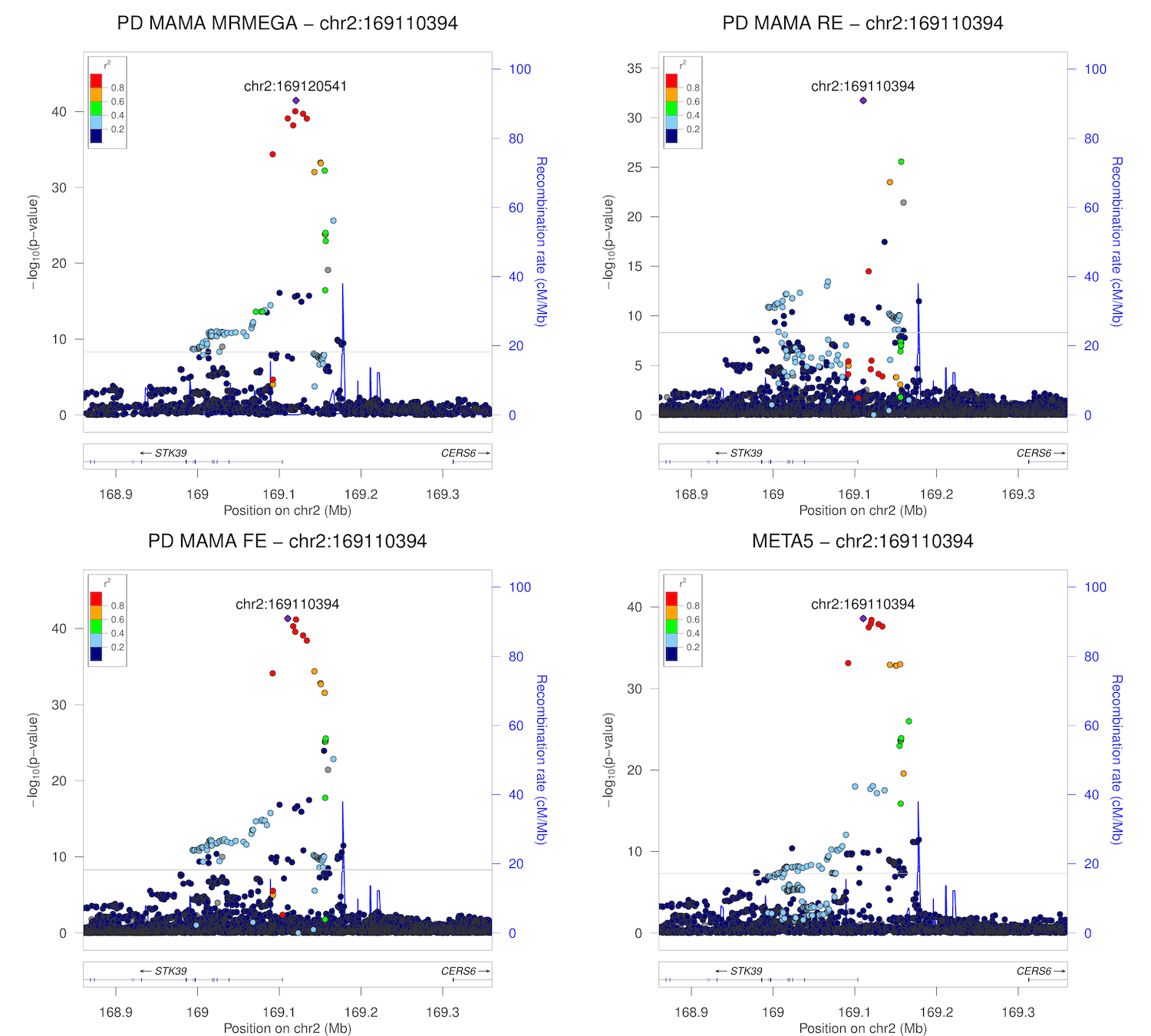

### chr3_18111759-18611759.png

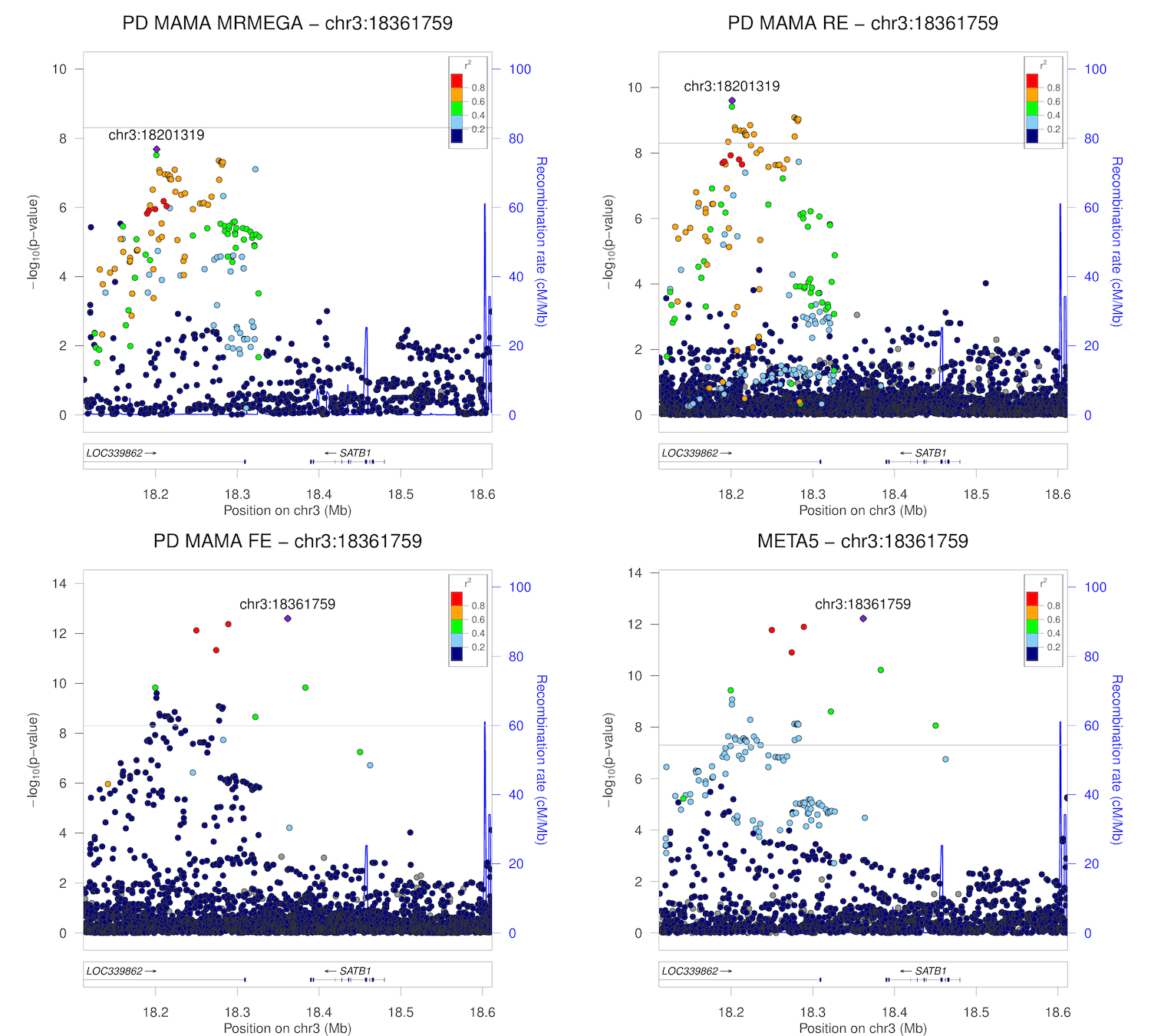

### chr4_76860365-77360365.png

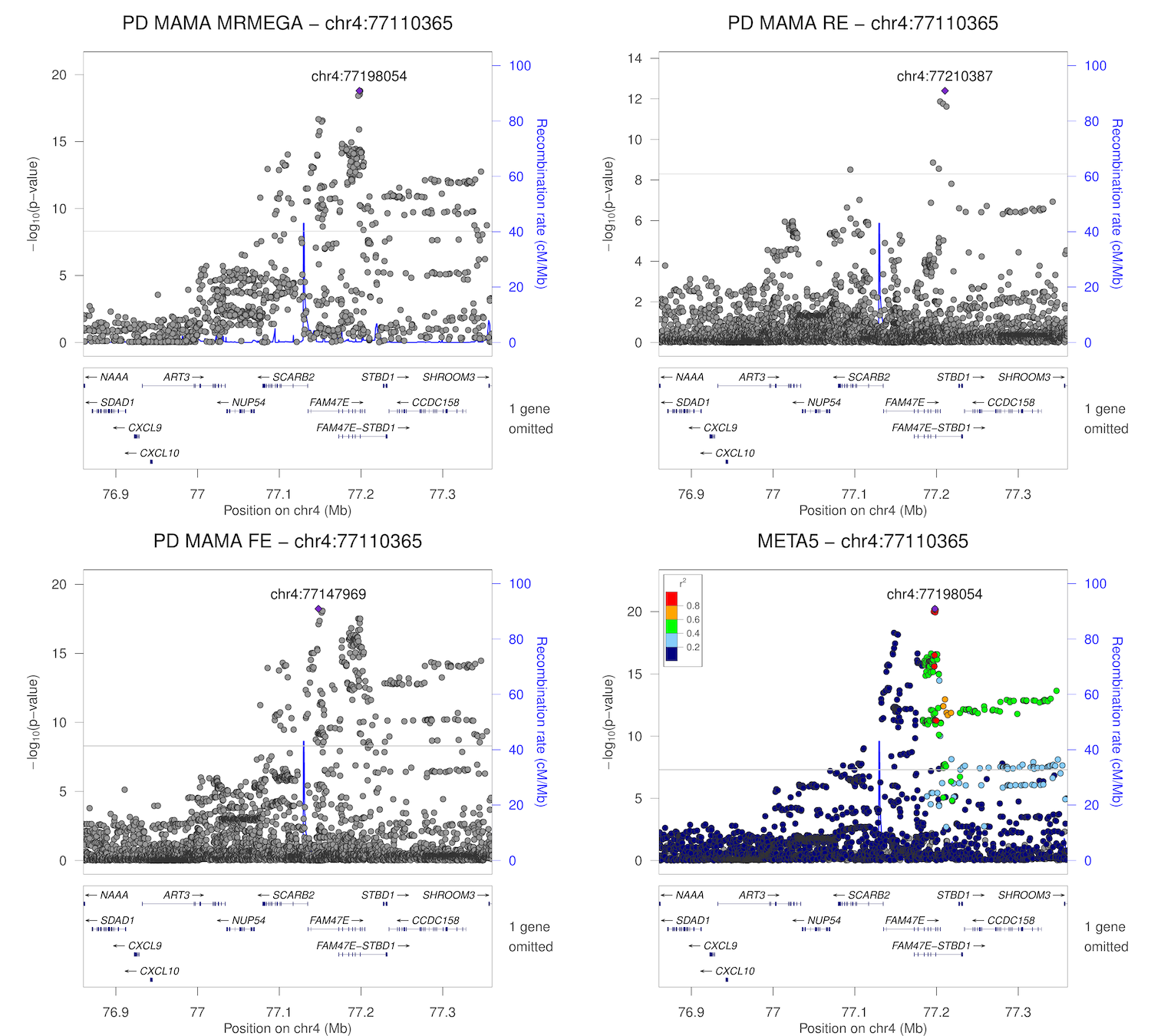

### chr12_123076598-123576598.png

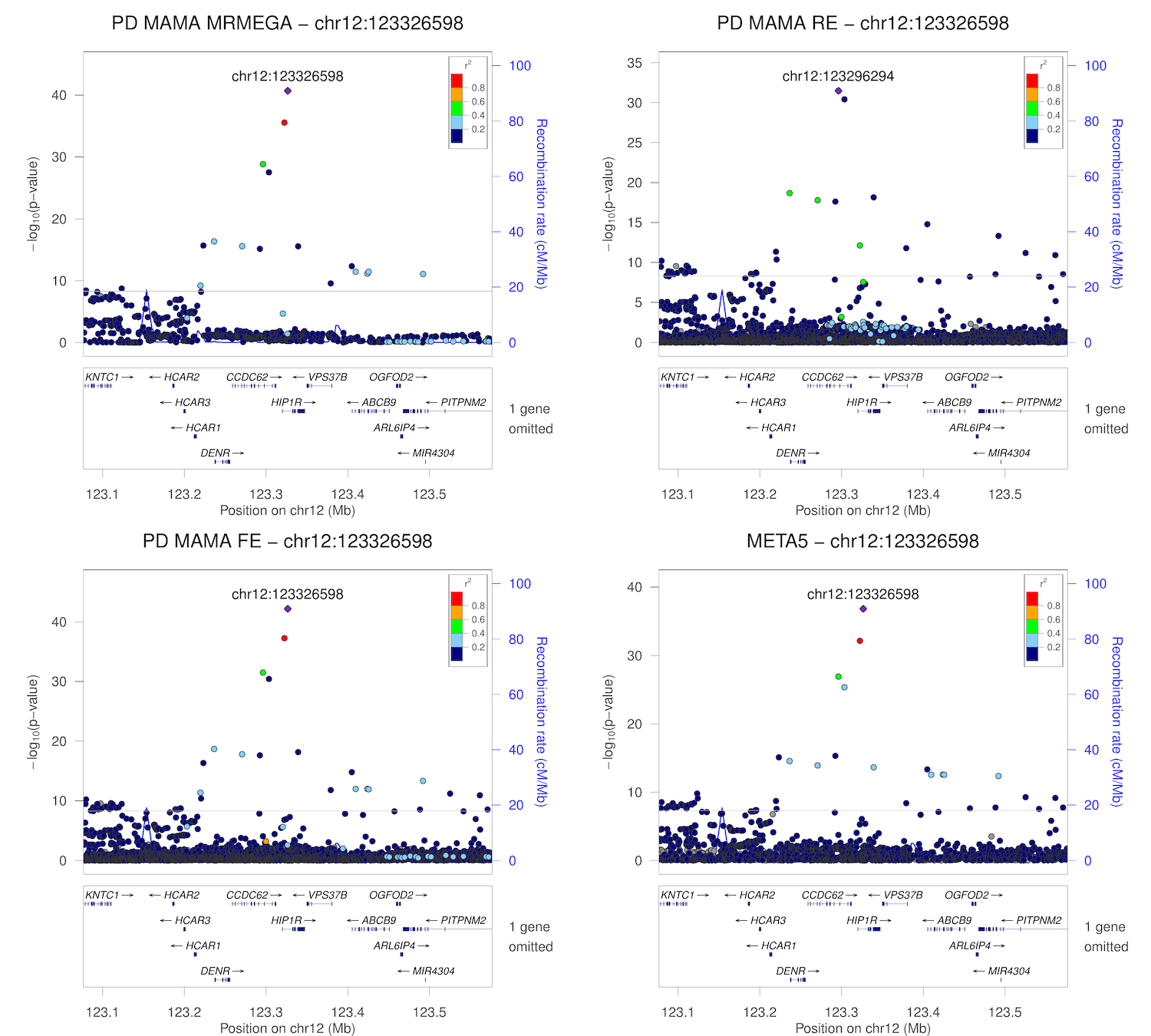

### chr13_97615021-98115021.png

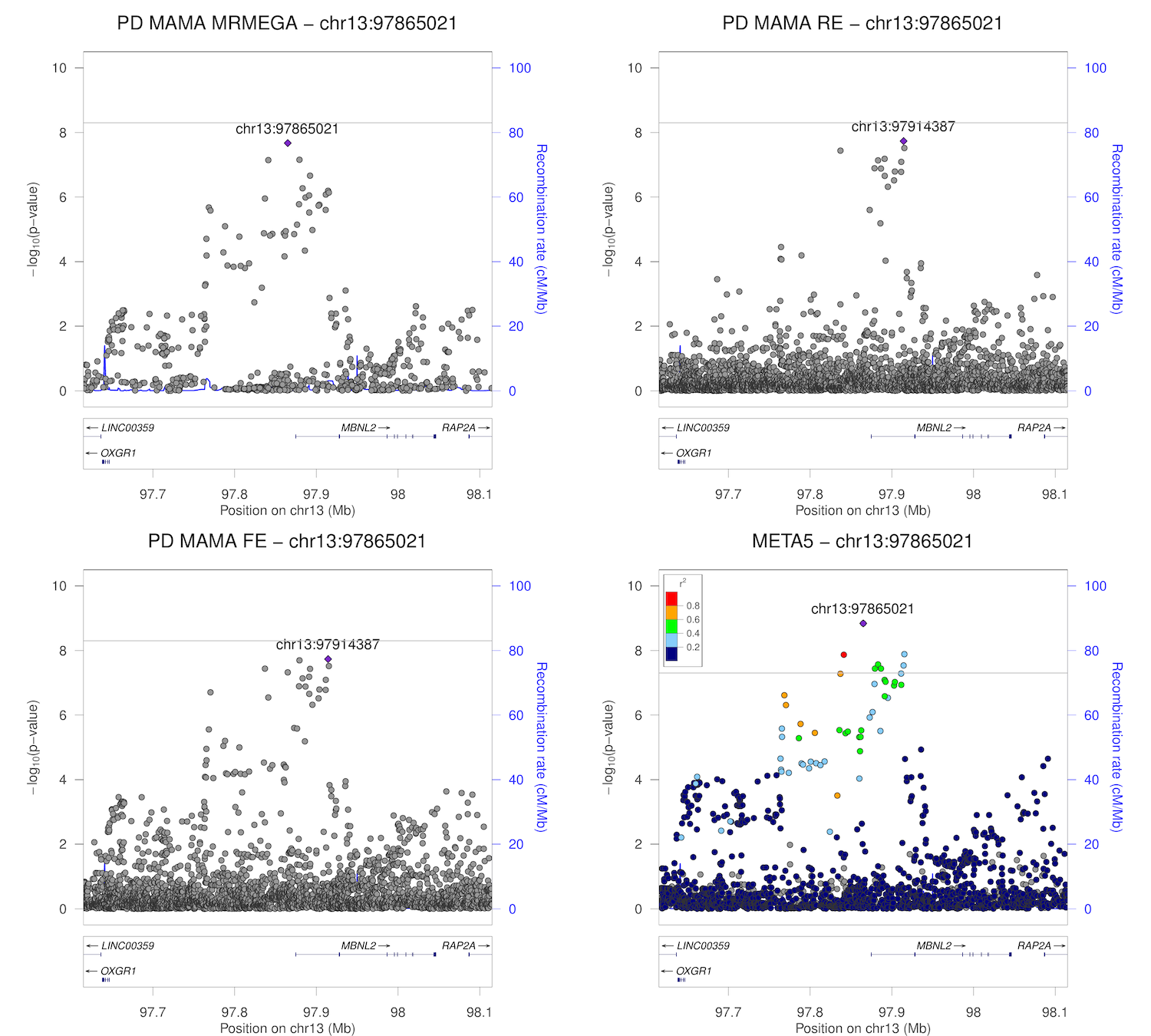

### chr14_75123034-75623034.png

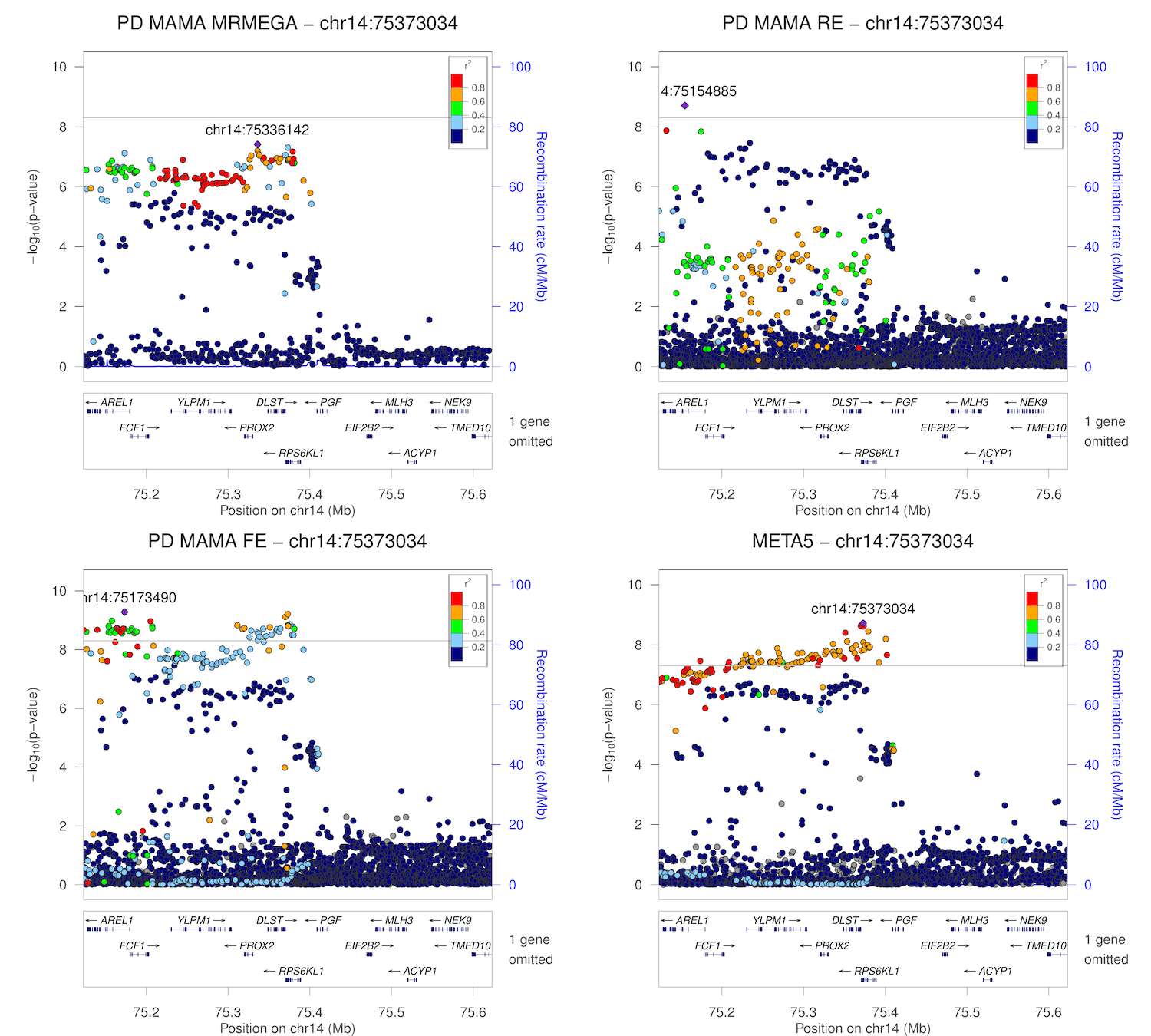

### chr16_28694396-29194396.png

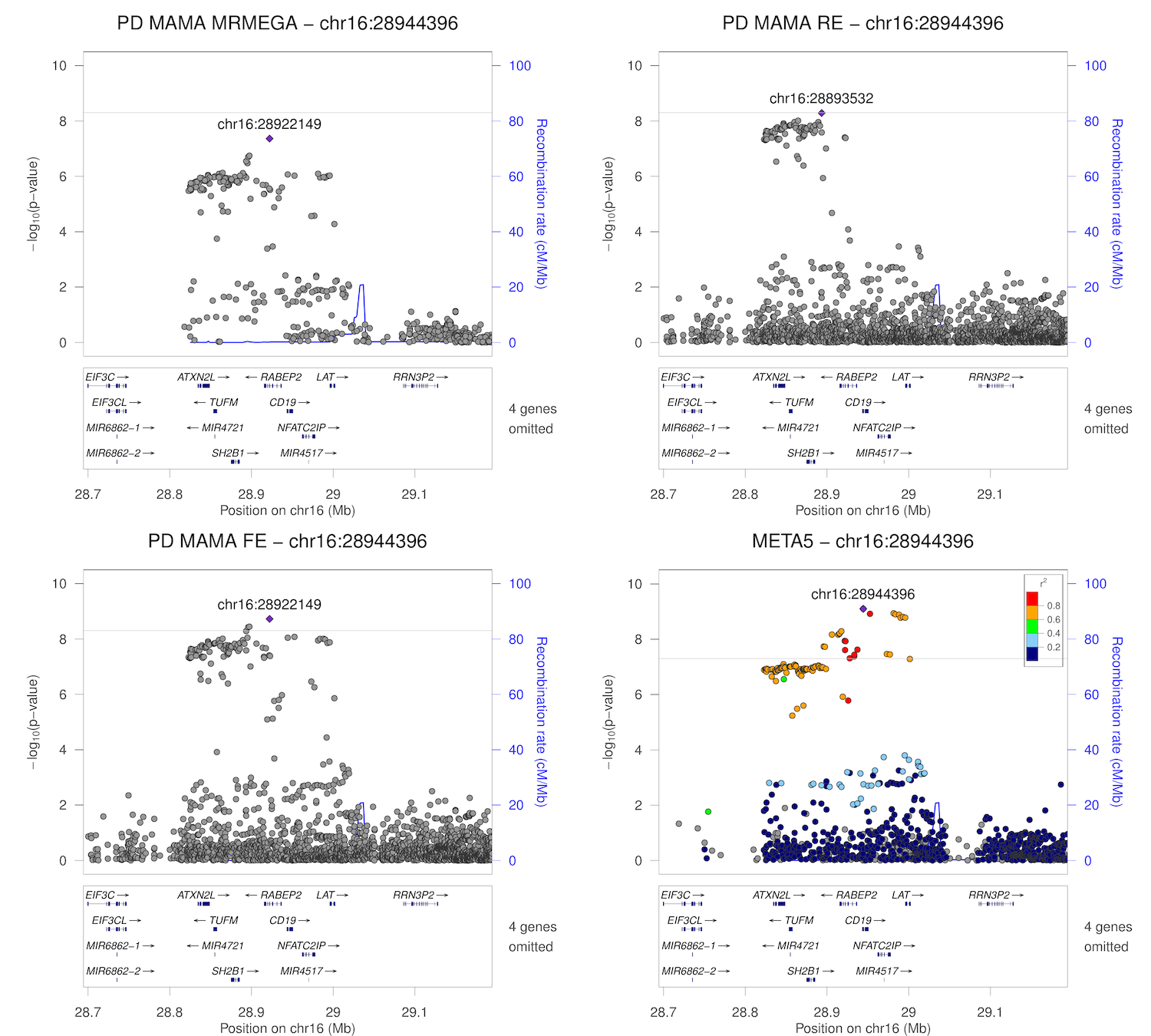

### chr17_42044337-42544337.png

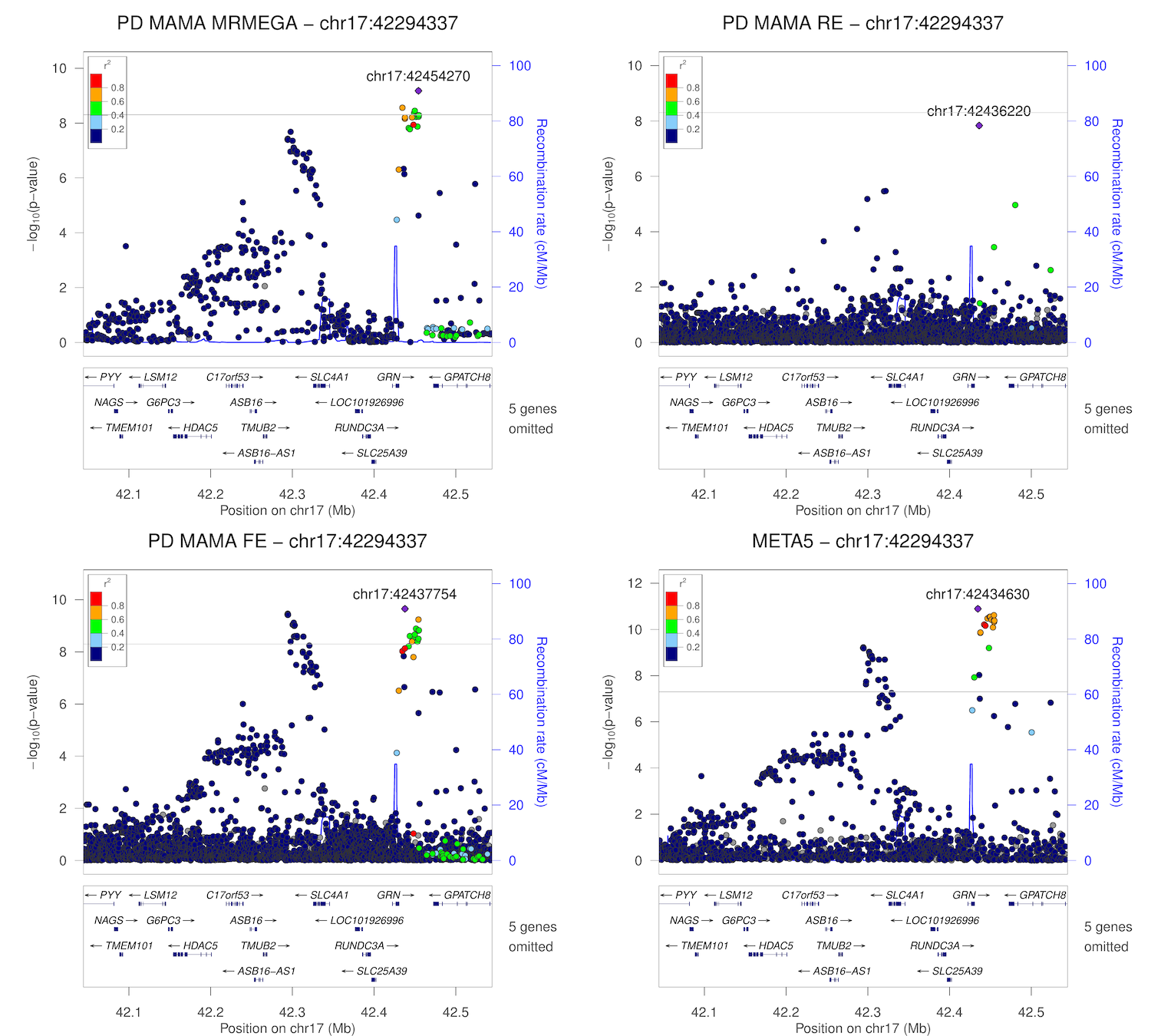

### chr17_43548308-44048308.png

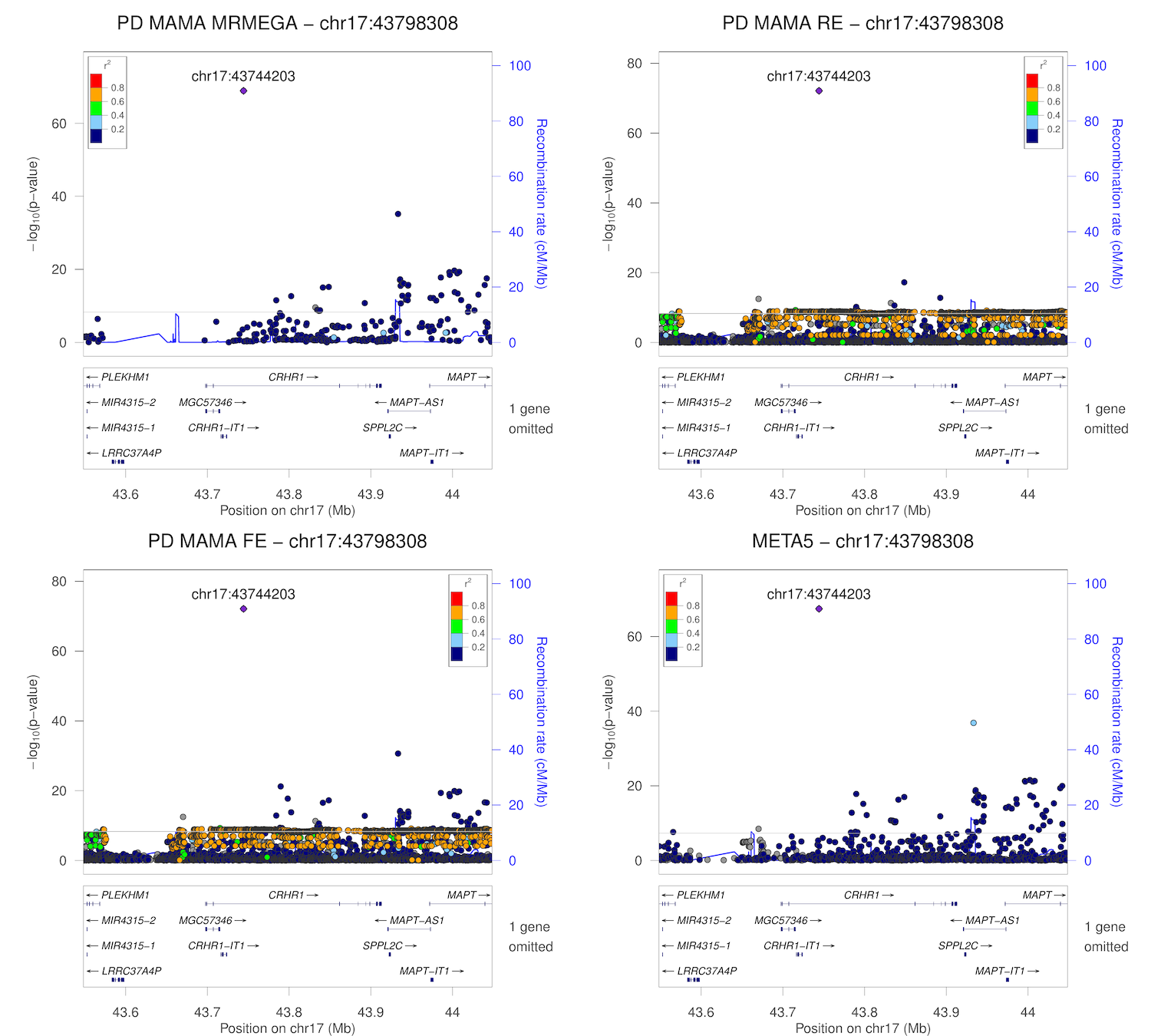

### chr19_2091047-2591047.png

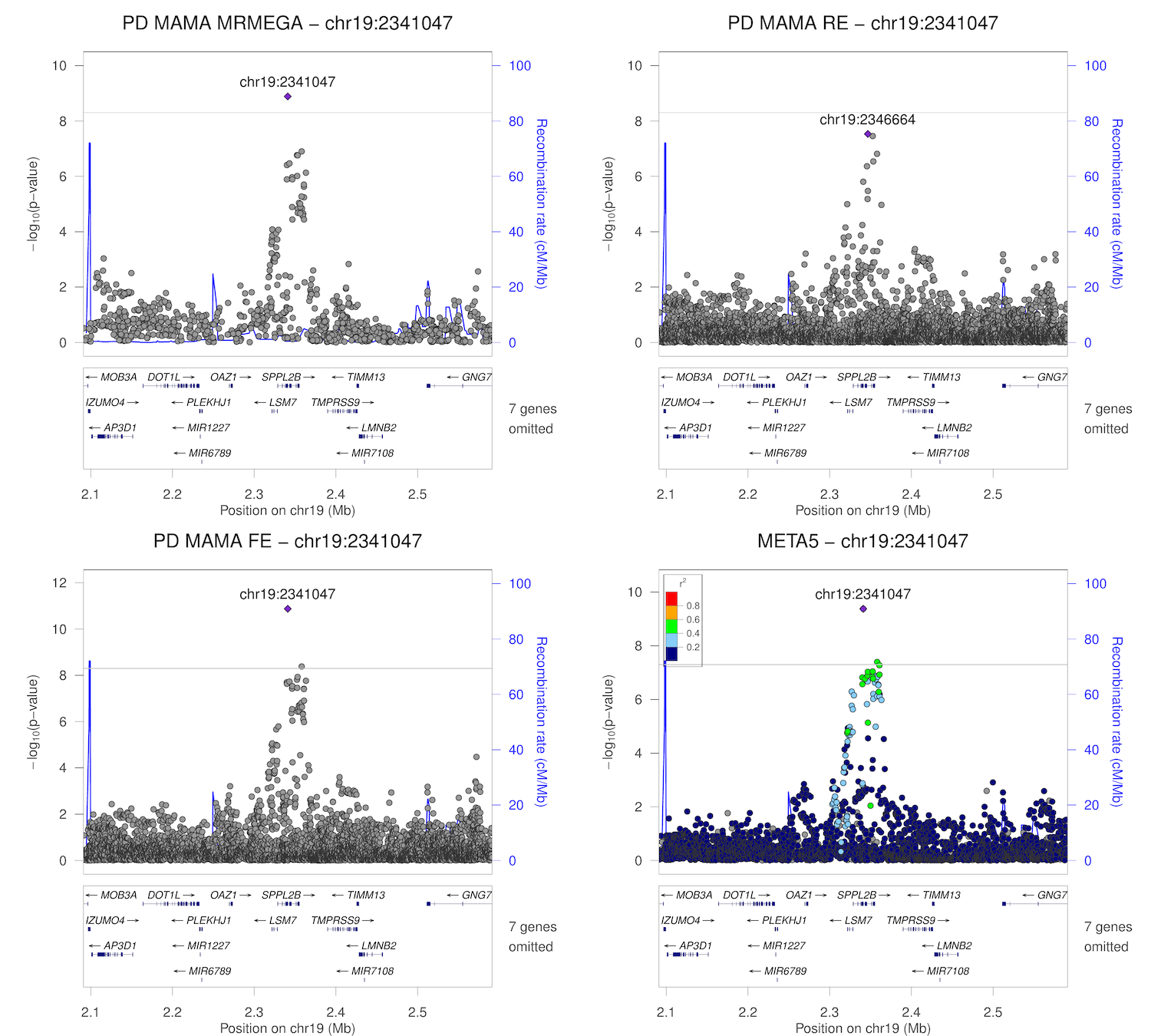

### COX4I2.png

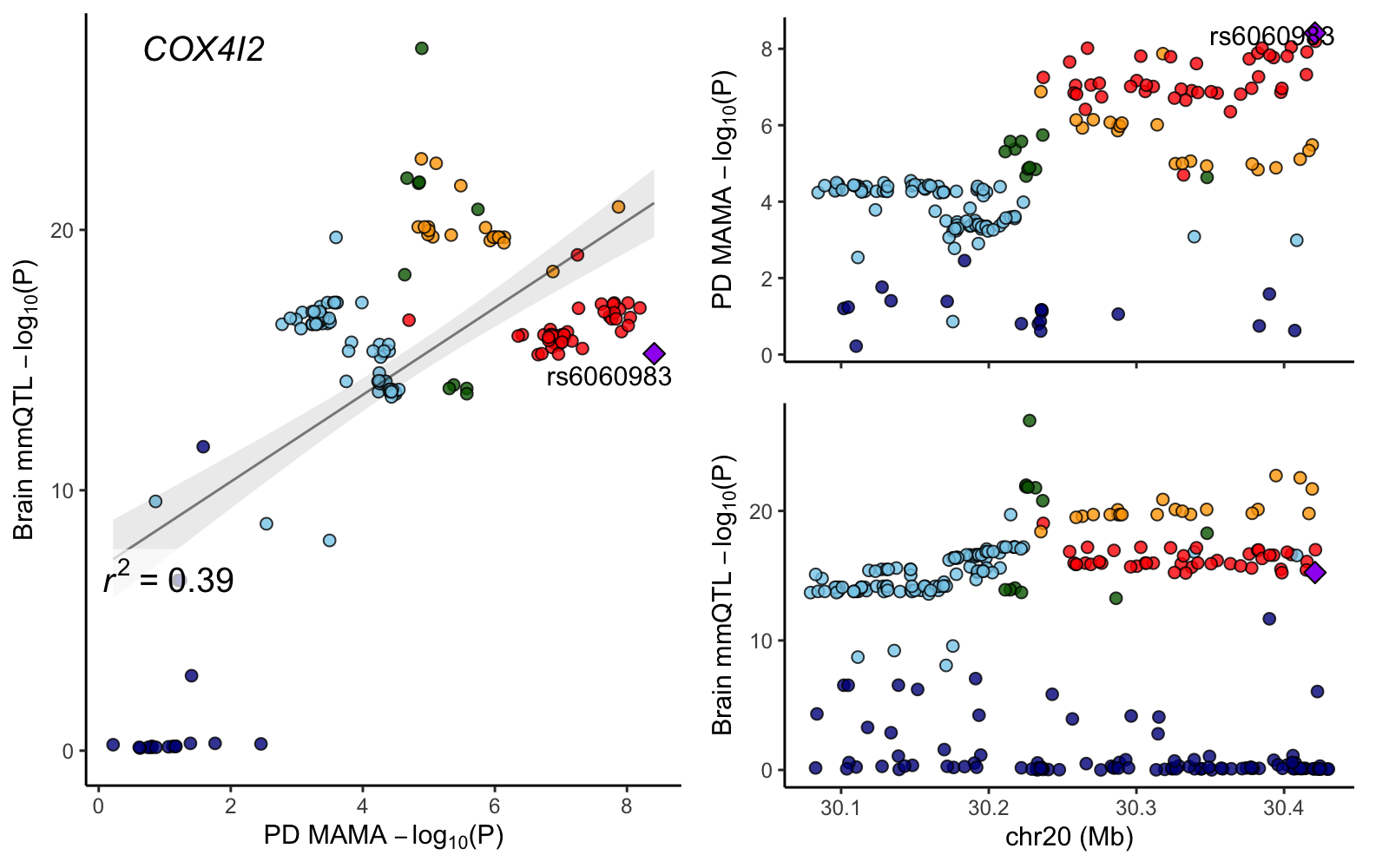

### CSDC2.png

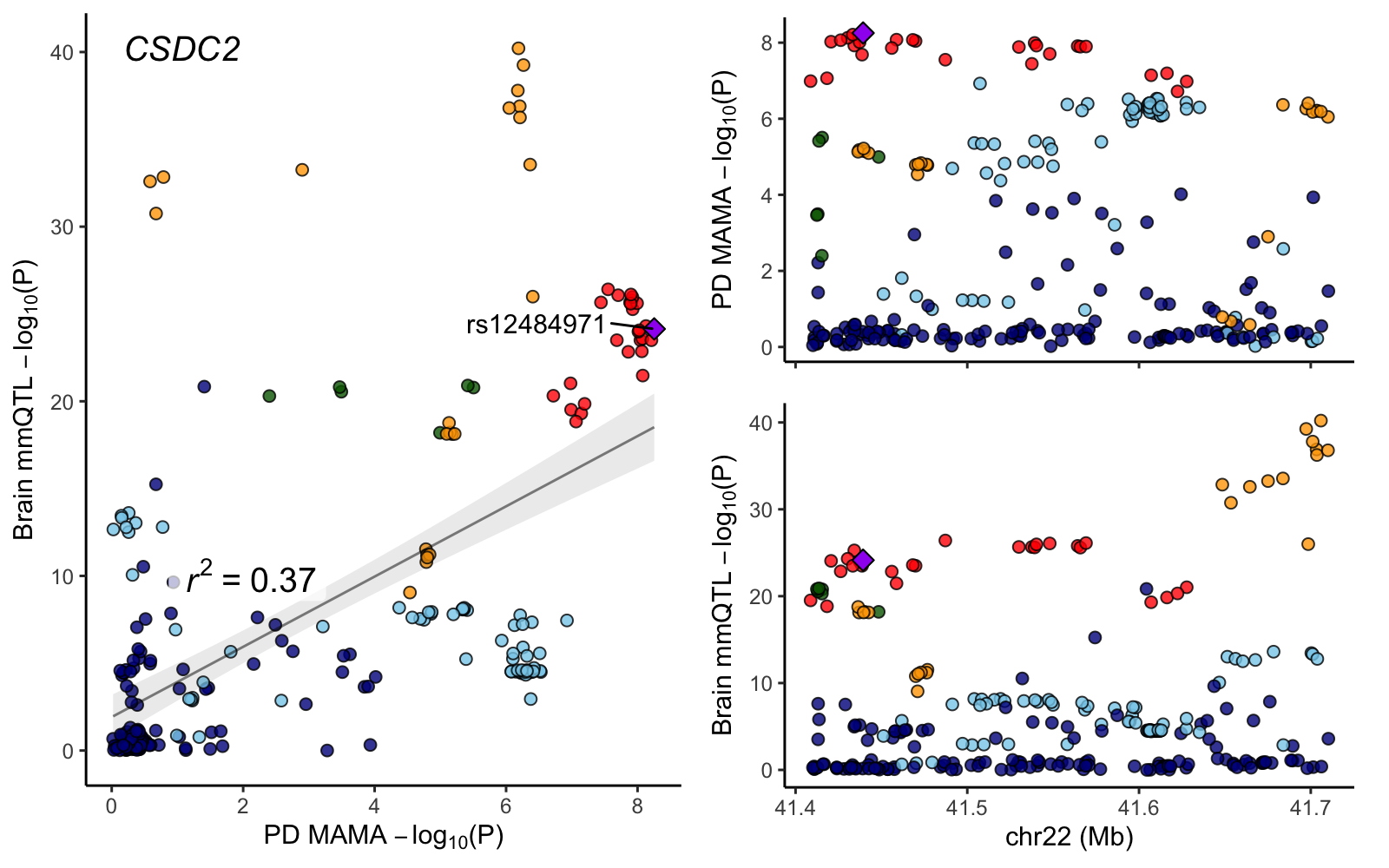

### DR1.png

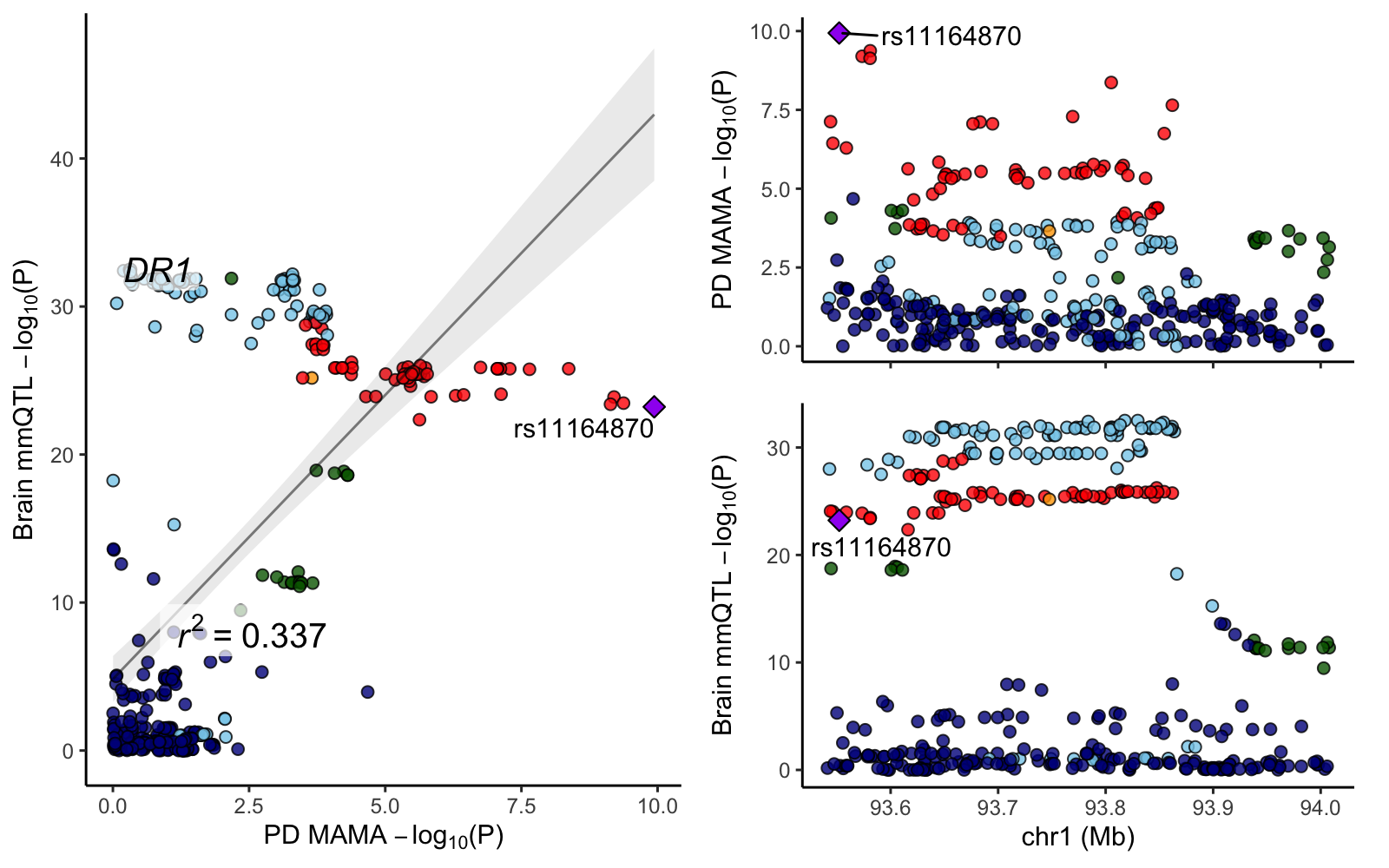

### EP300.png

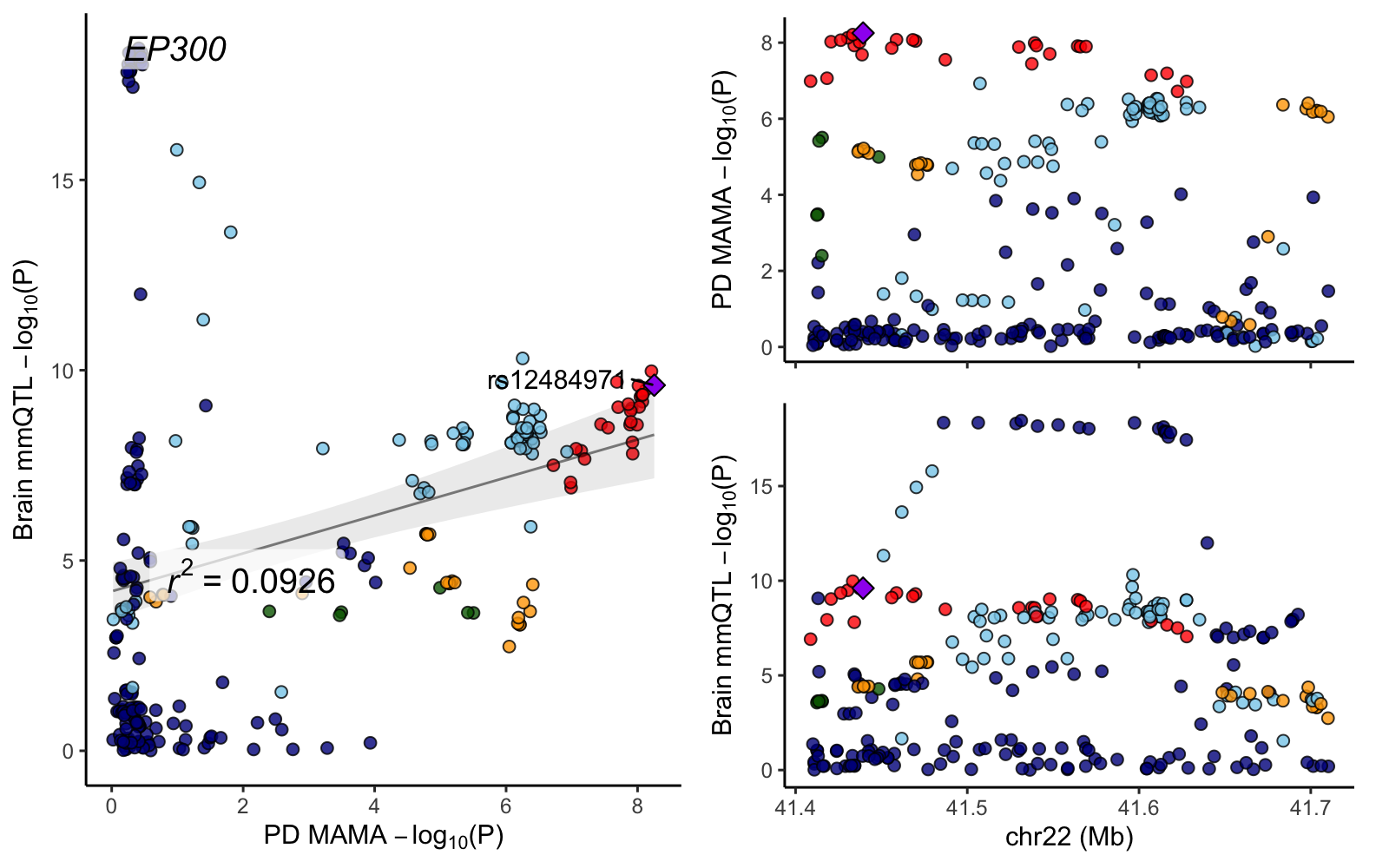

### FOXS1.png

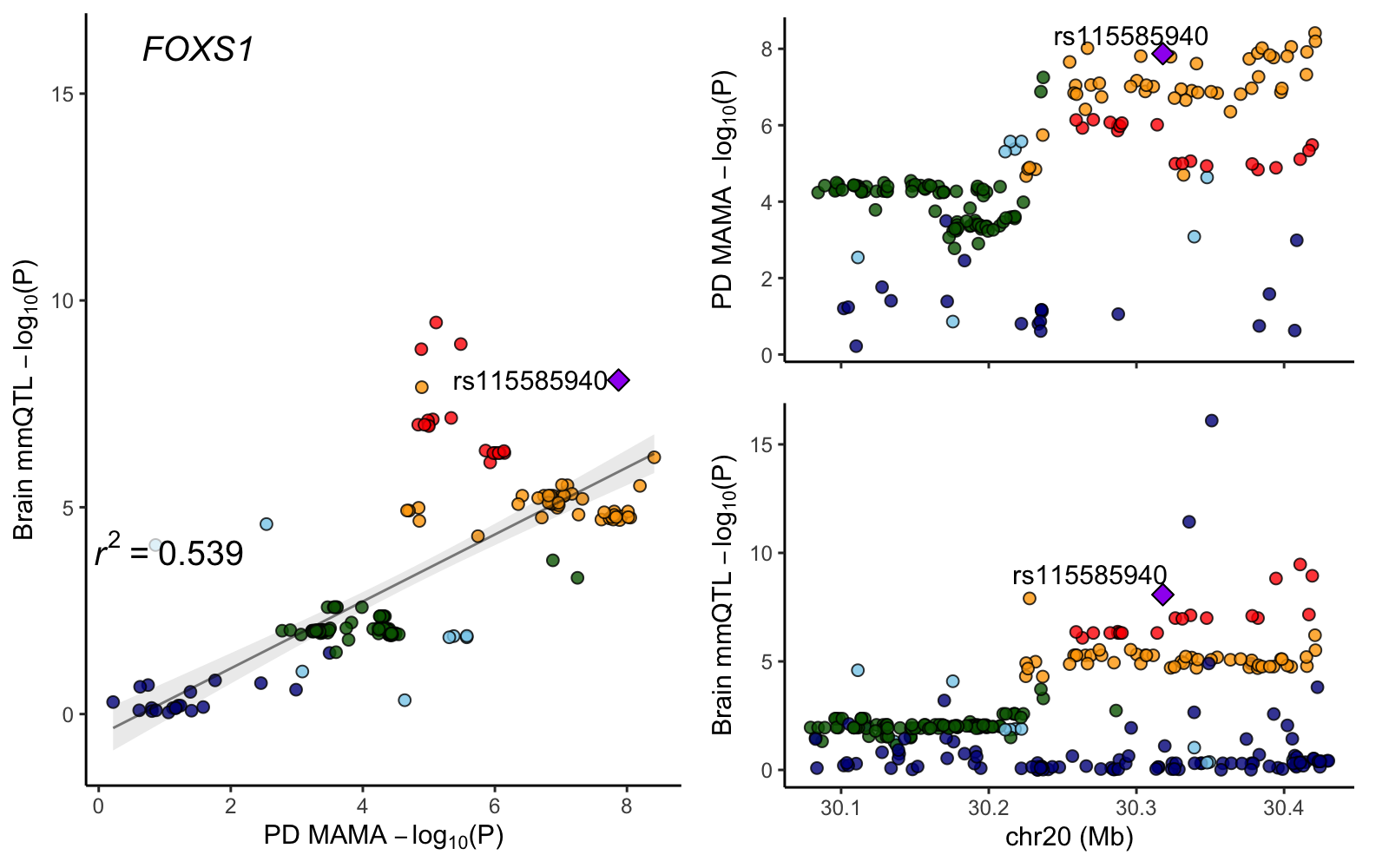

### HM13.png

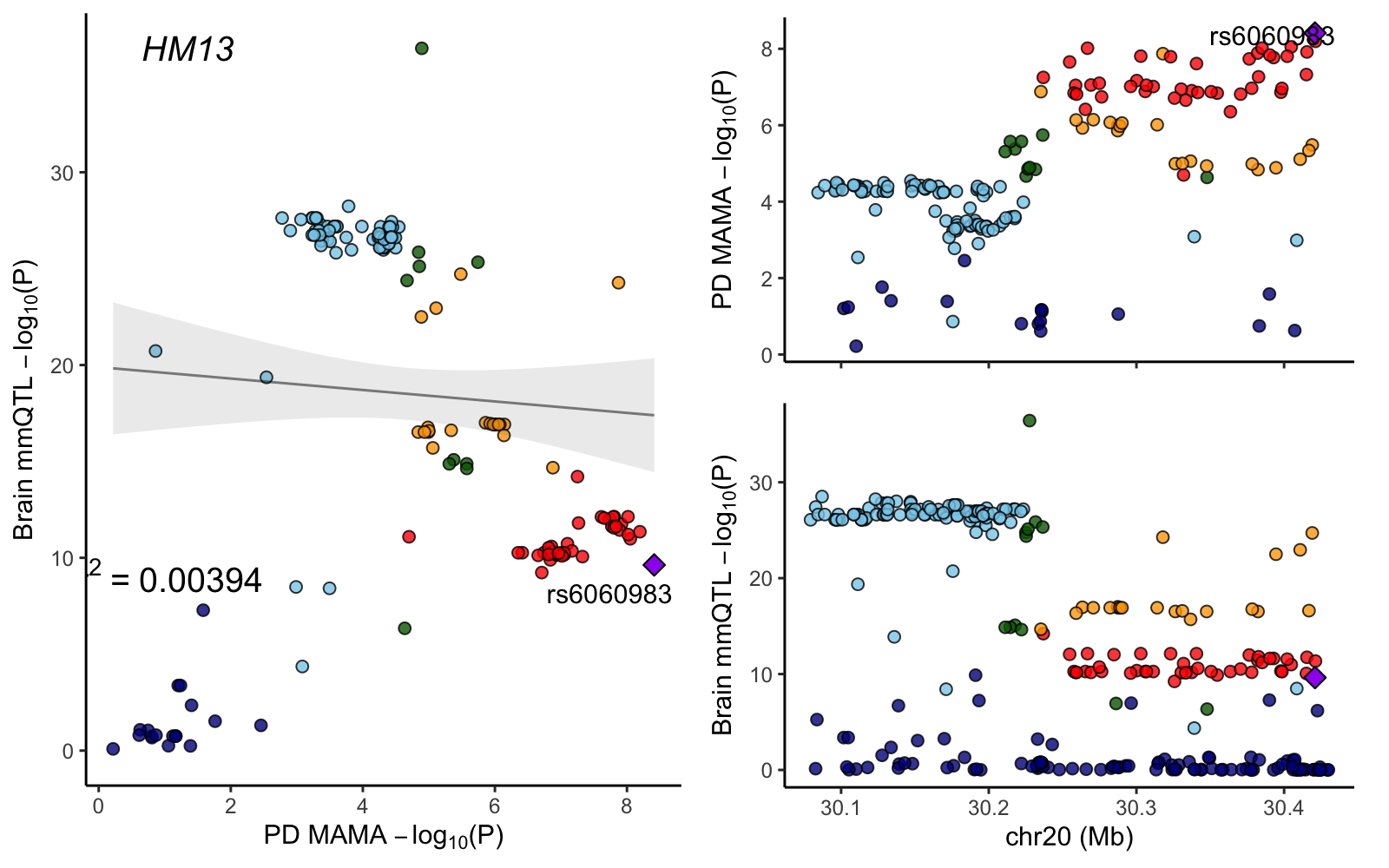

### KCNMB3.png

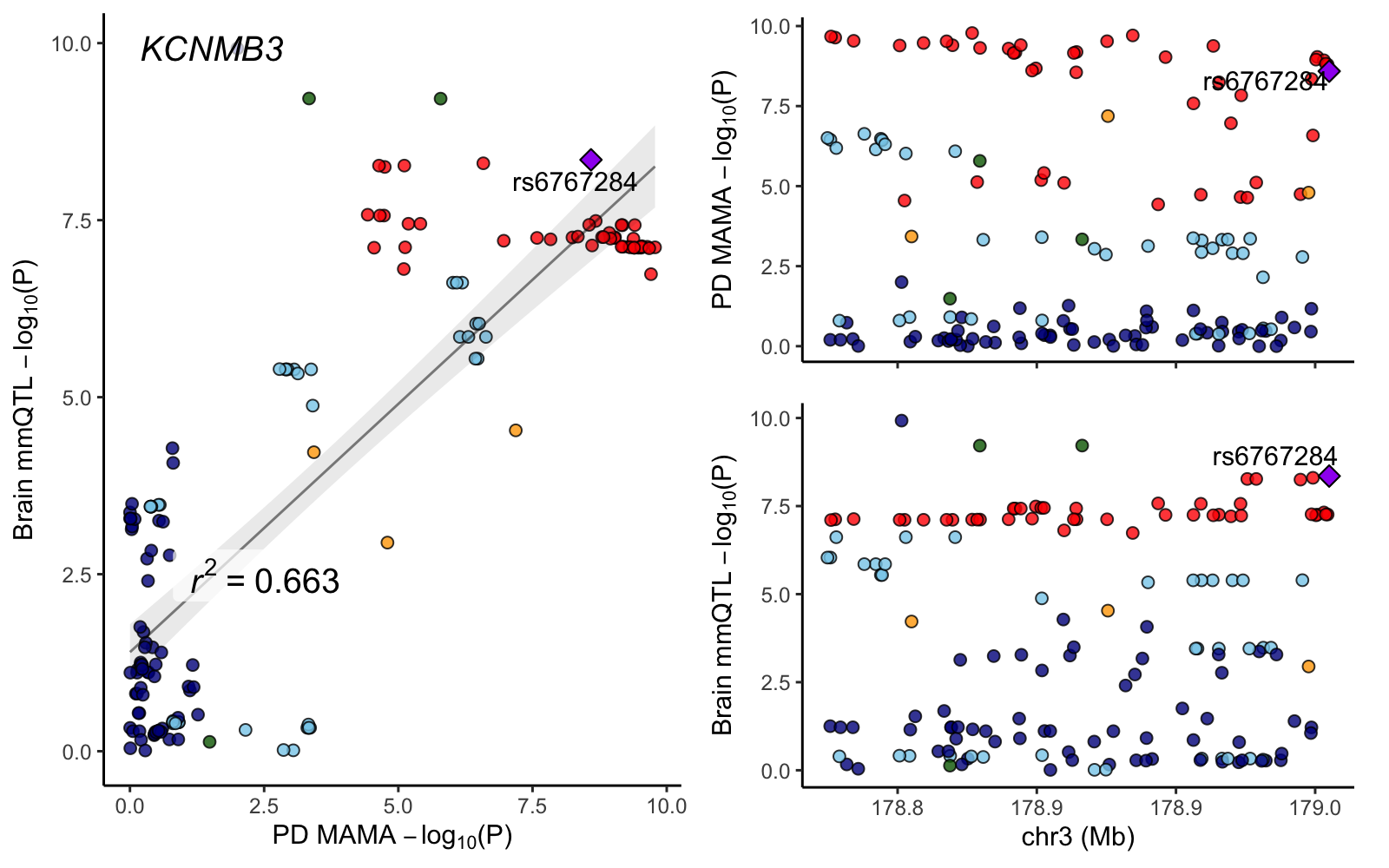

### L3MBTL2.png

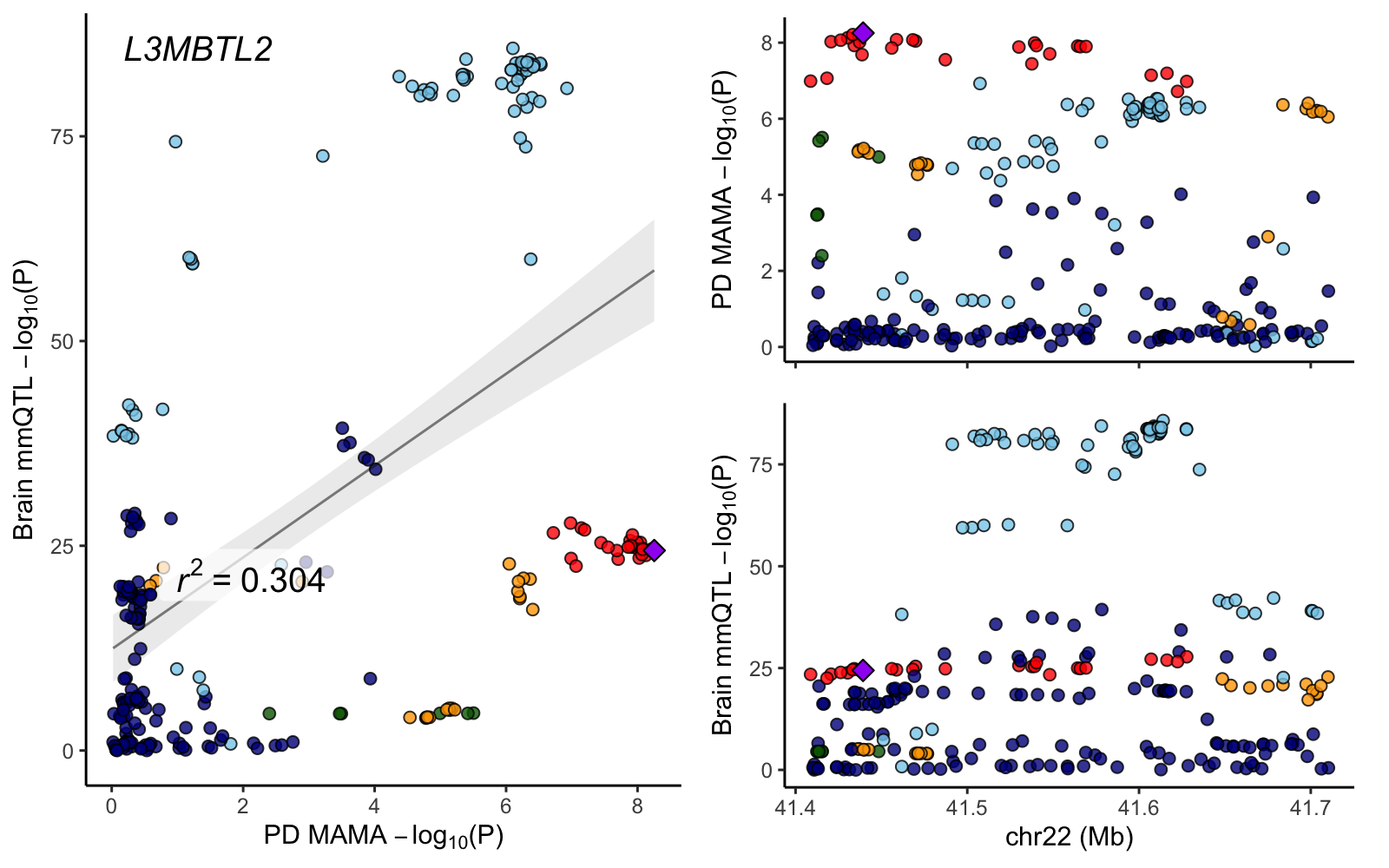

### MEI1.png

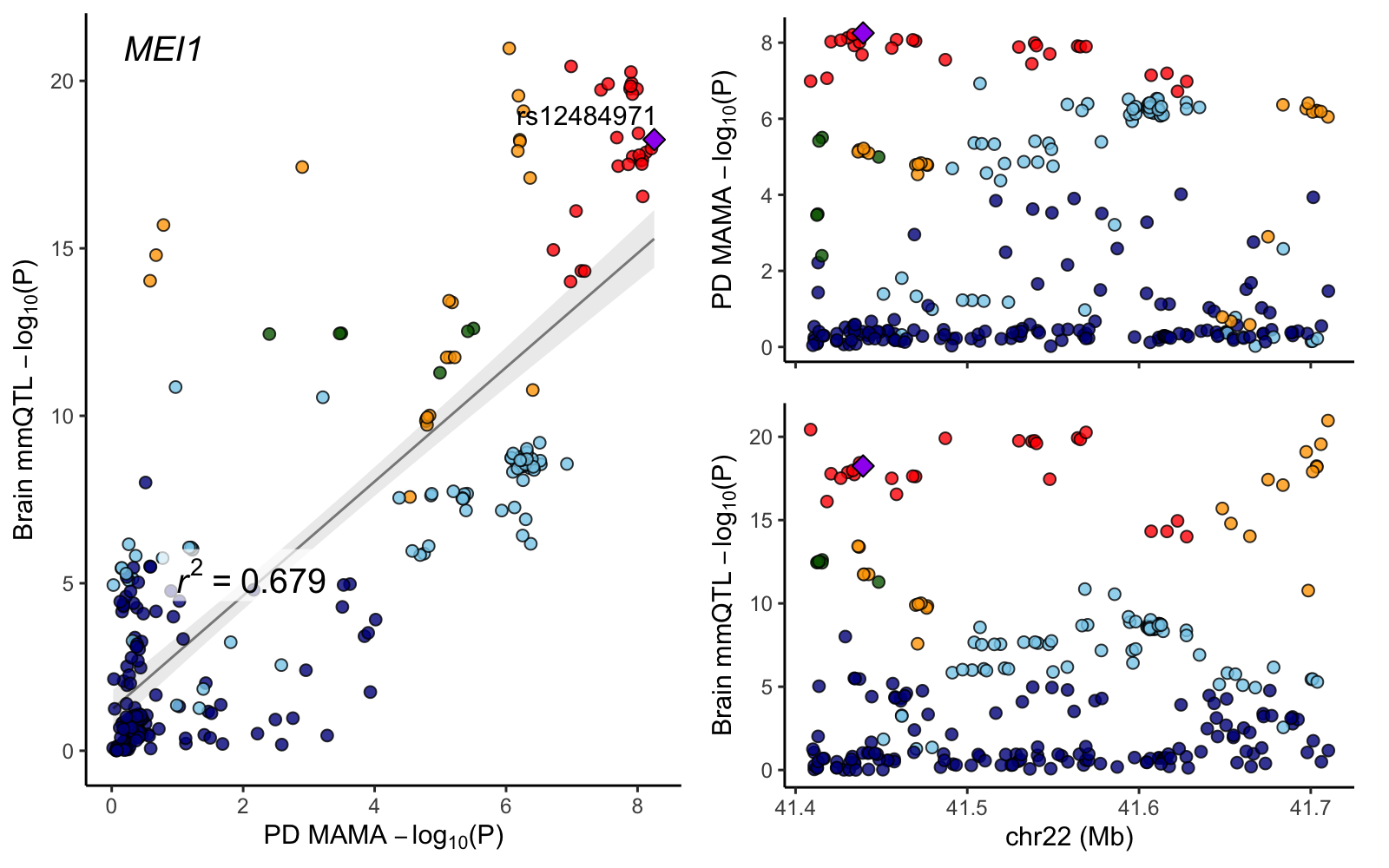

### NCOR1.png

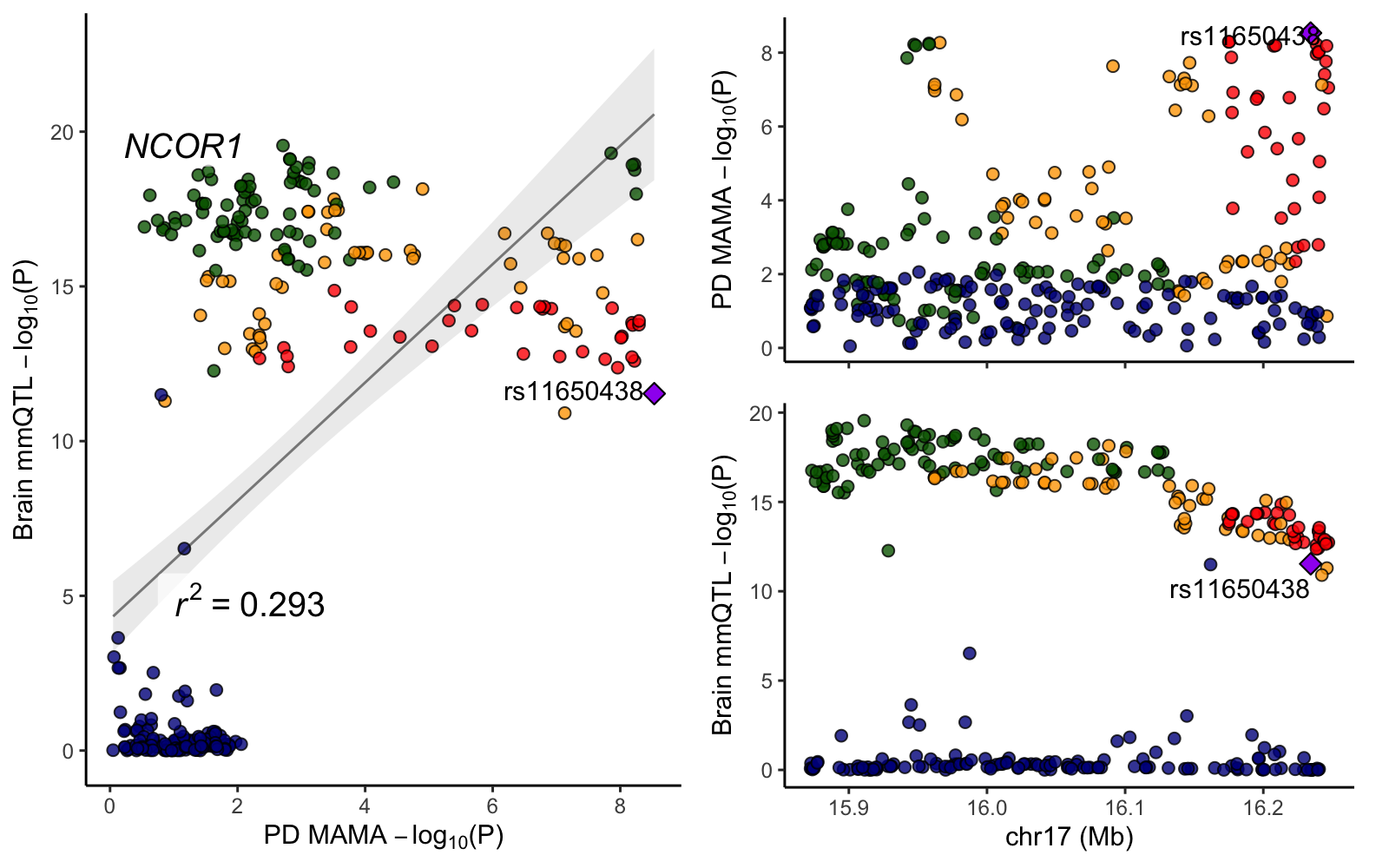

### NOP14-AS1.png

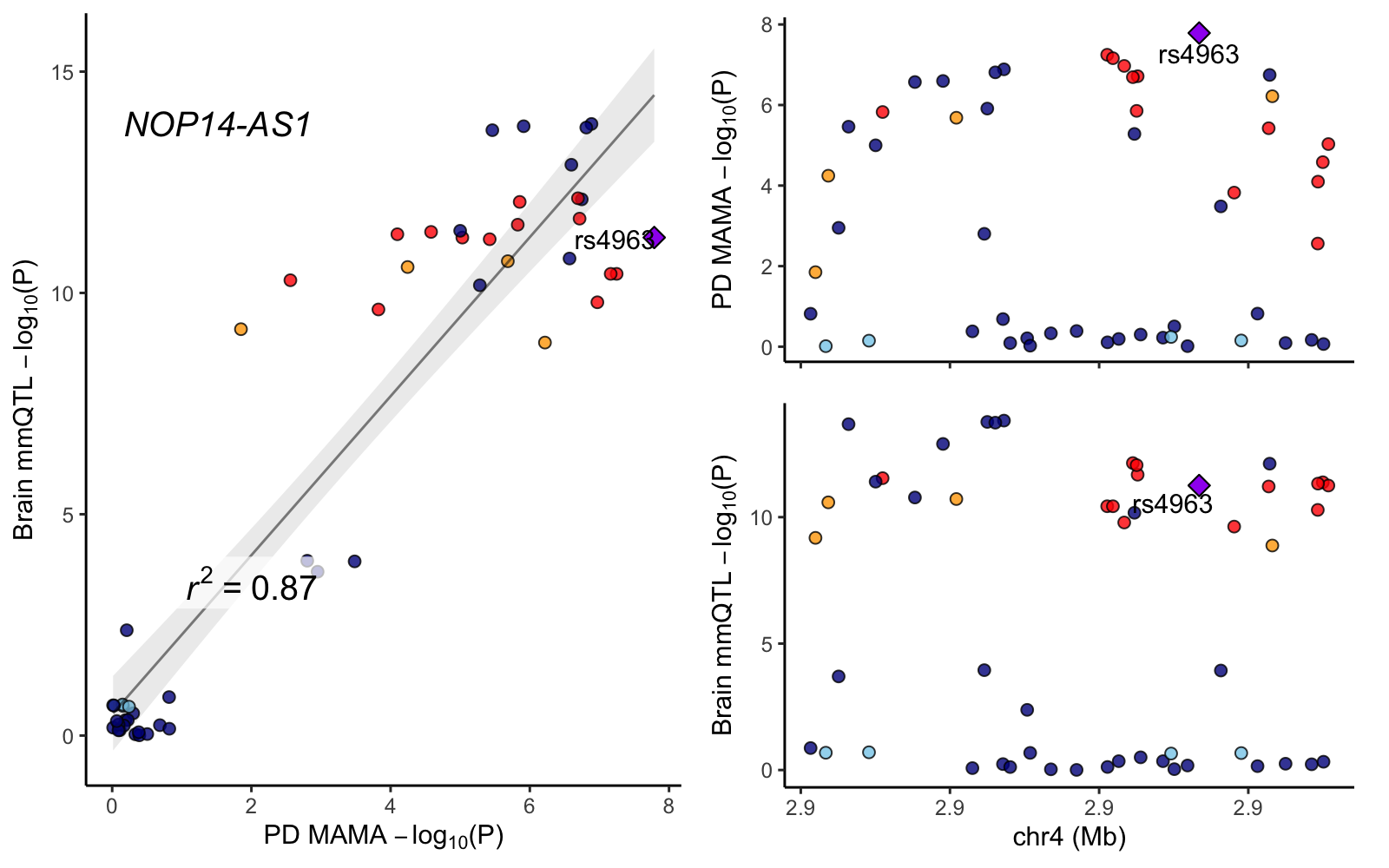

### PIGL.png

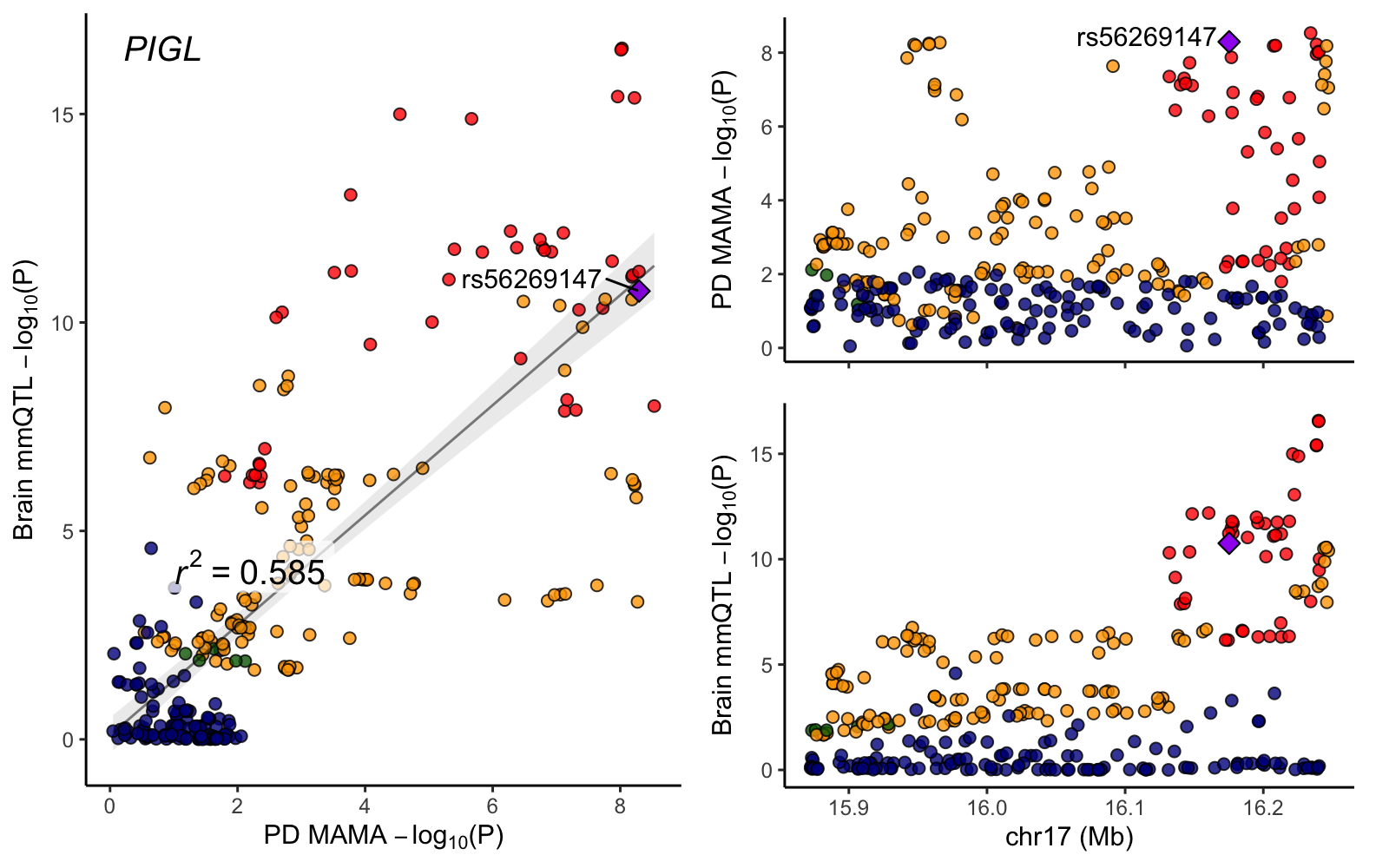

### PMM1.png

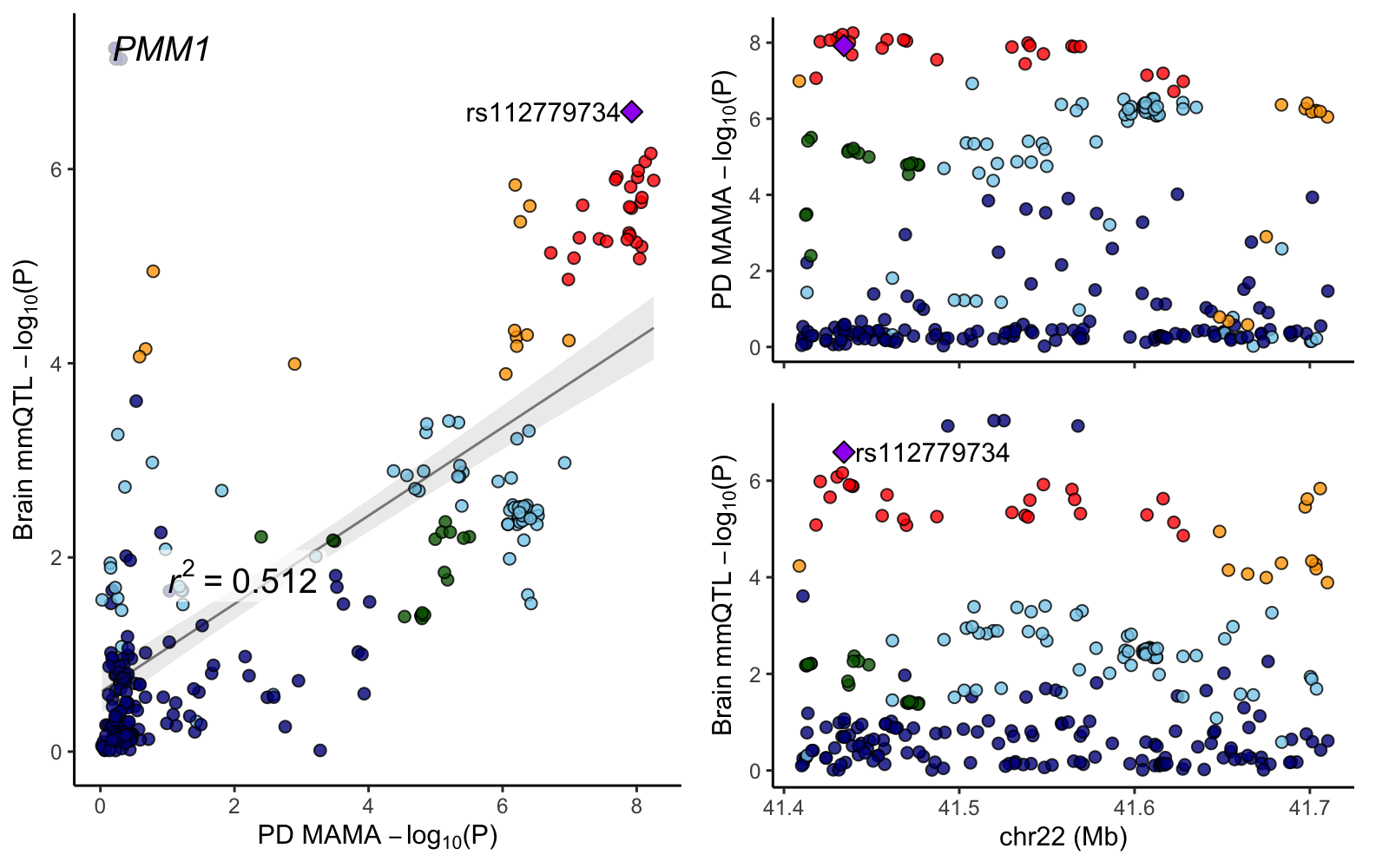

### POLR3H.png

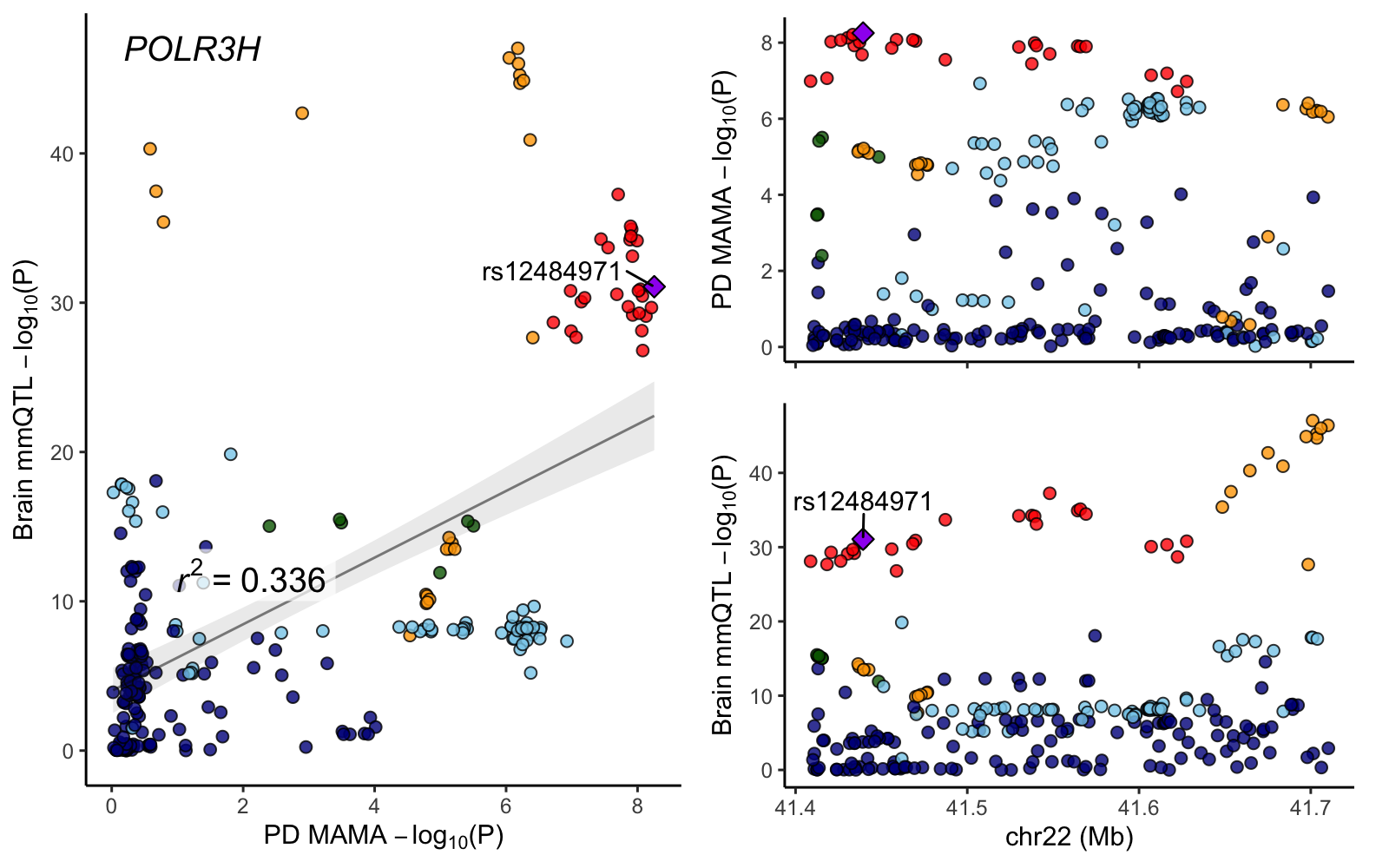

### PPP6R2.png

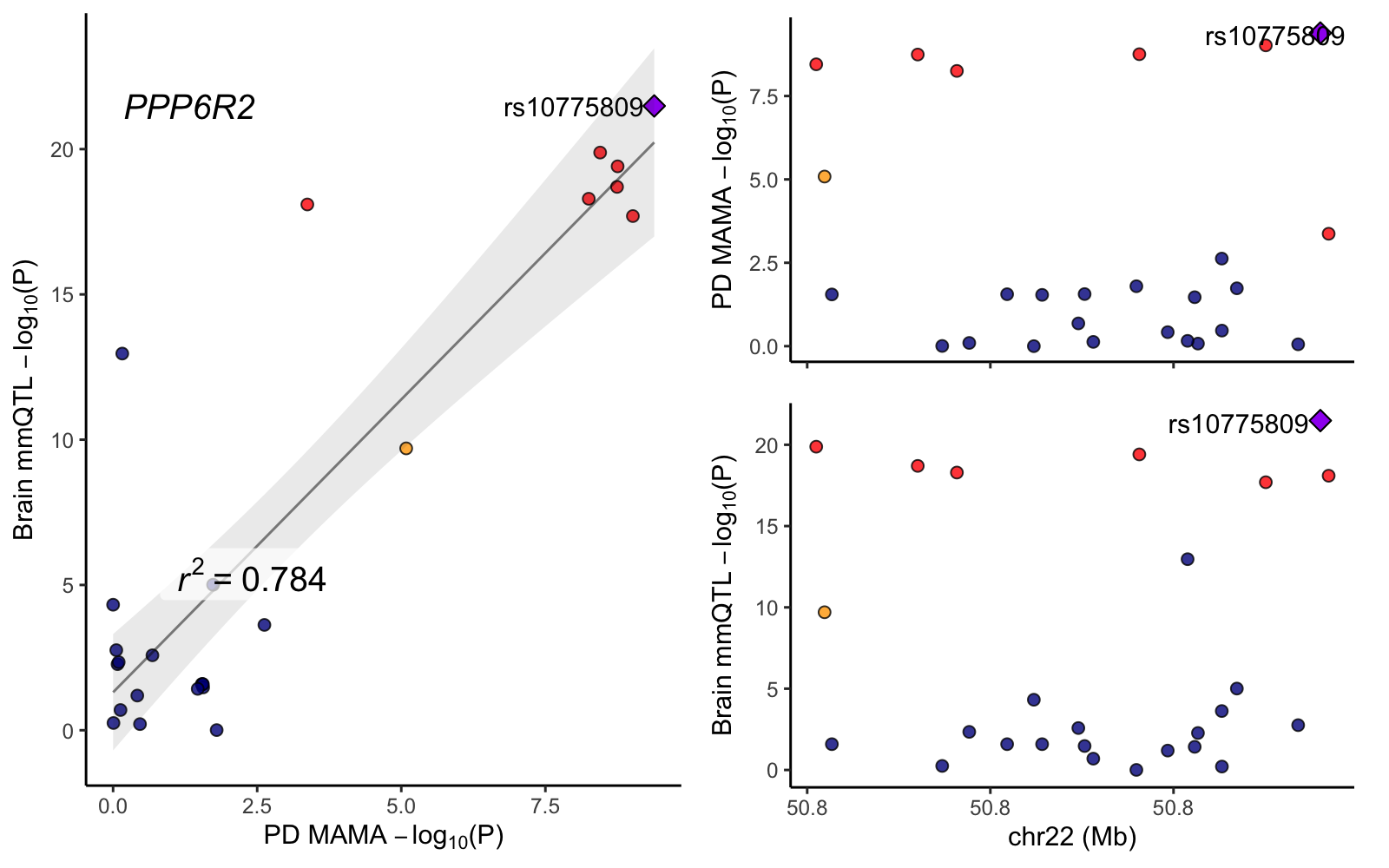

### SLC25A17.png

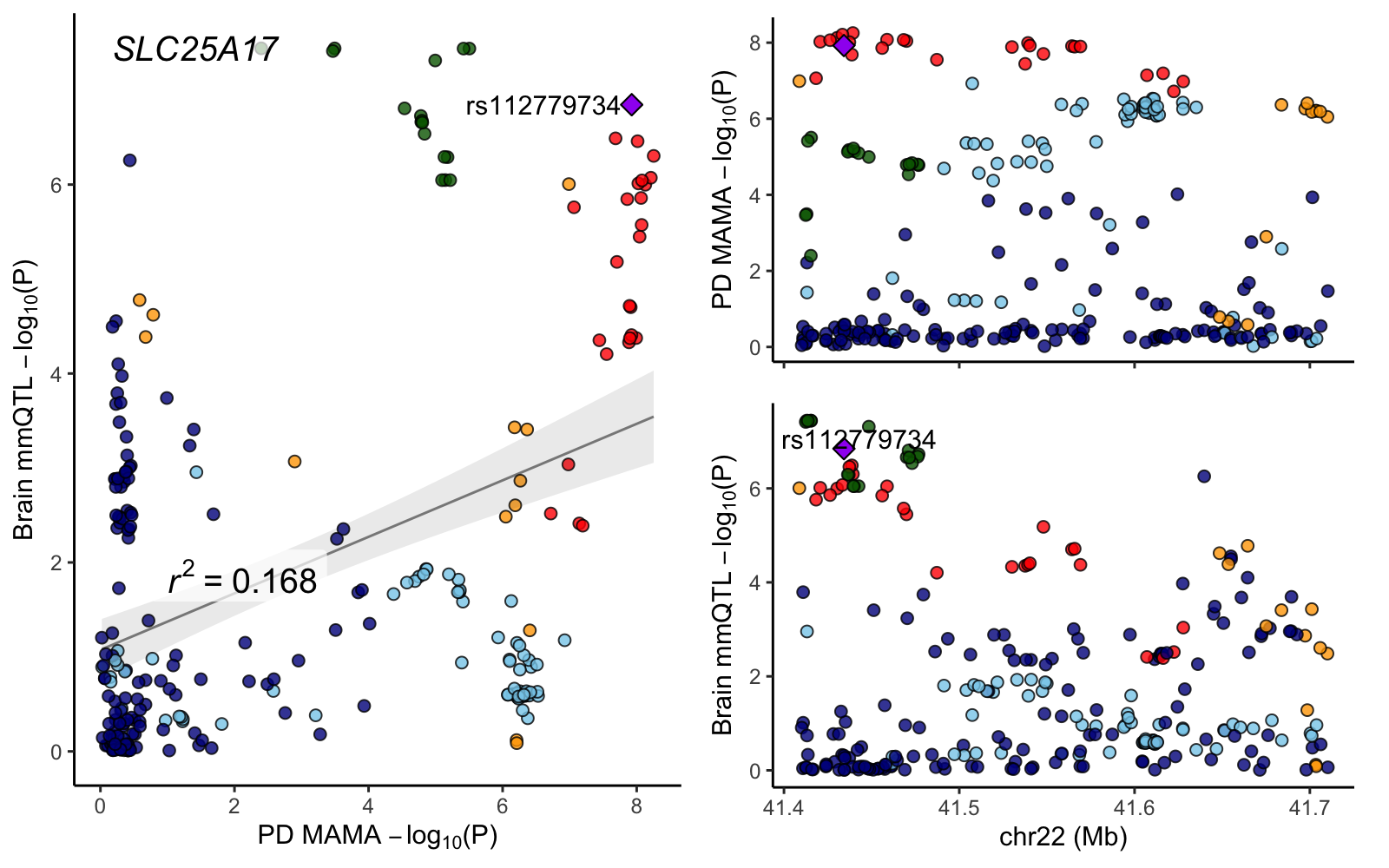

### SYBU.png

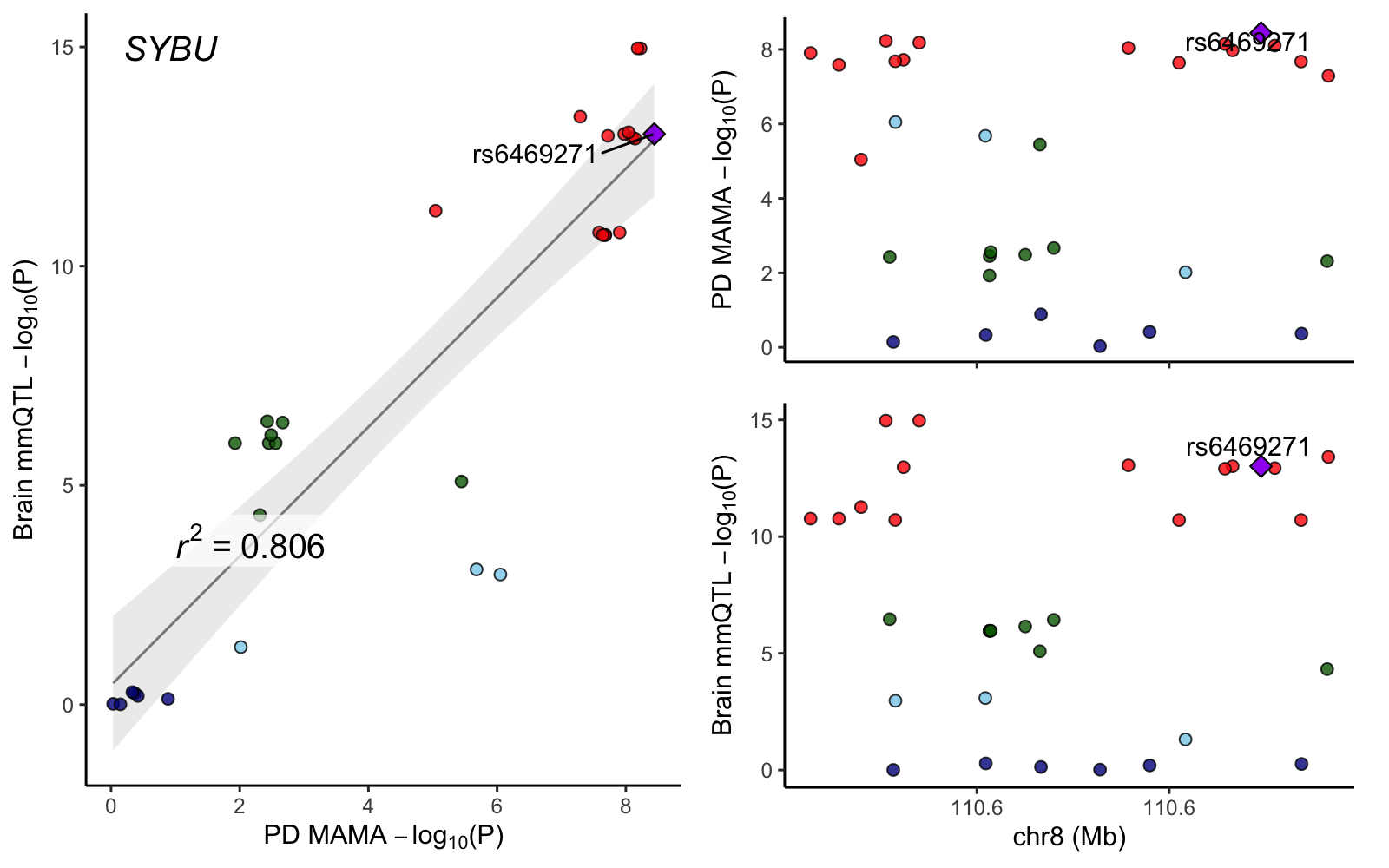

### TMED5.png

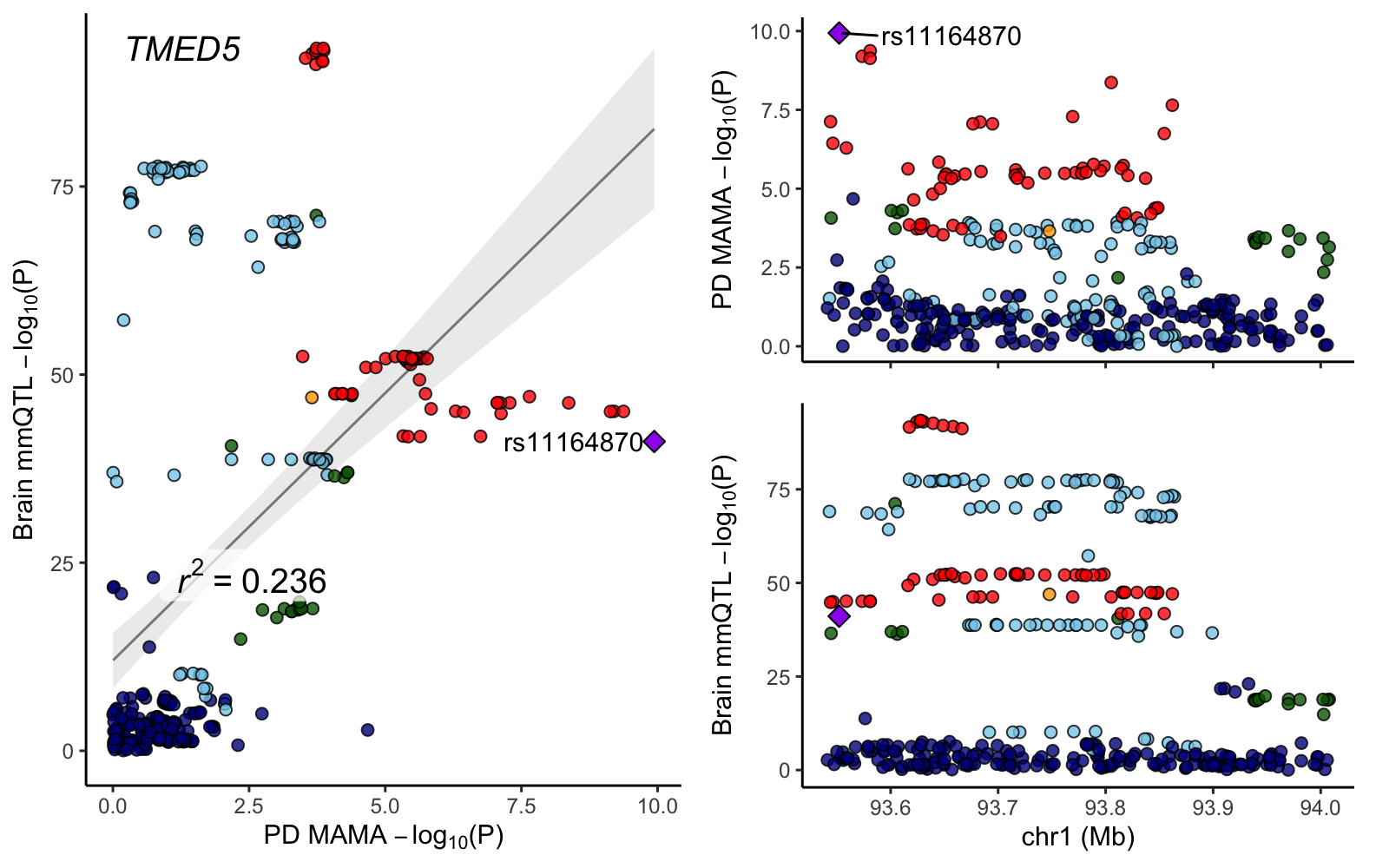

### TRPV2.png

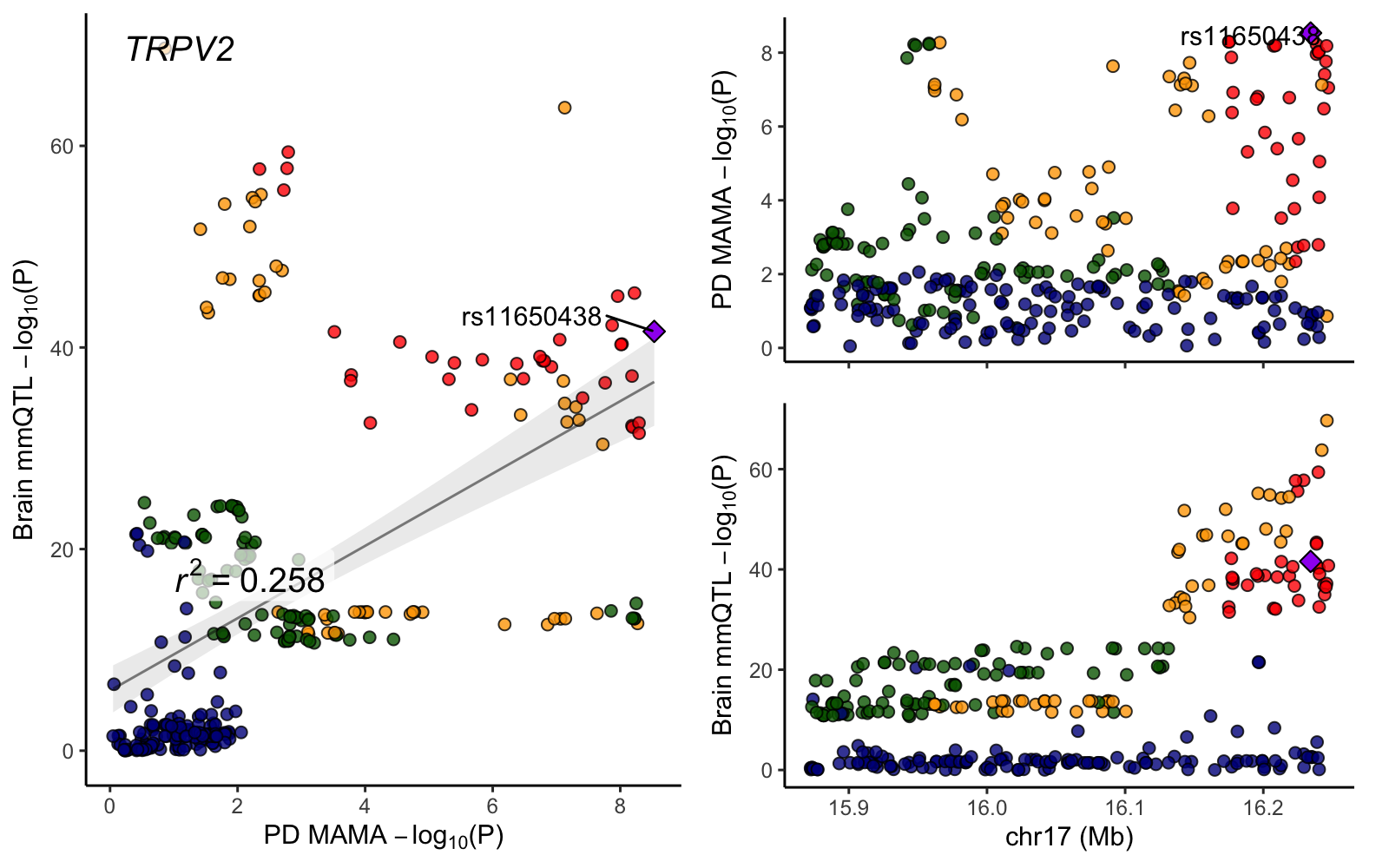

### TTC19.png

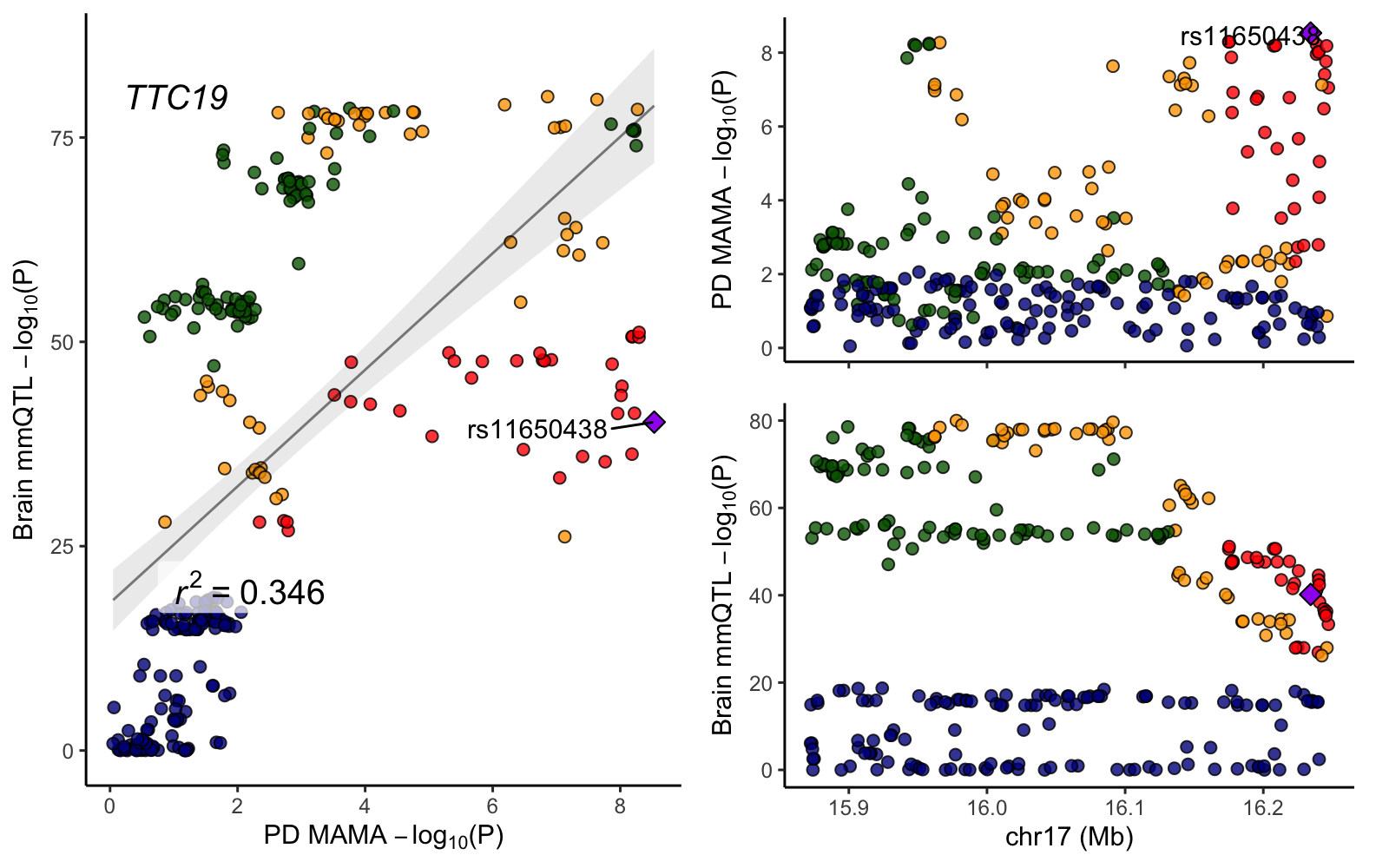

### XPNPEP3.png

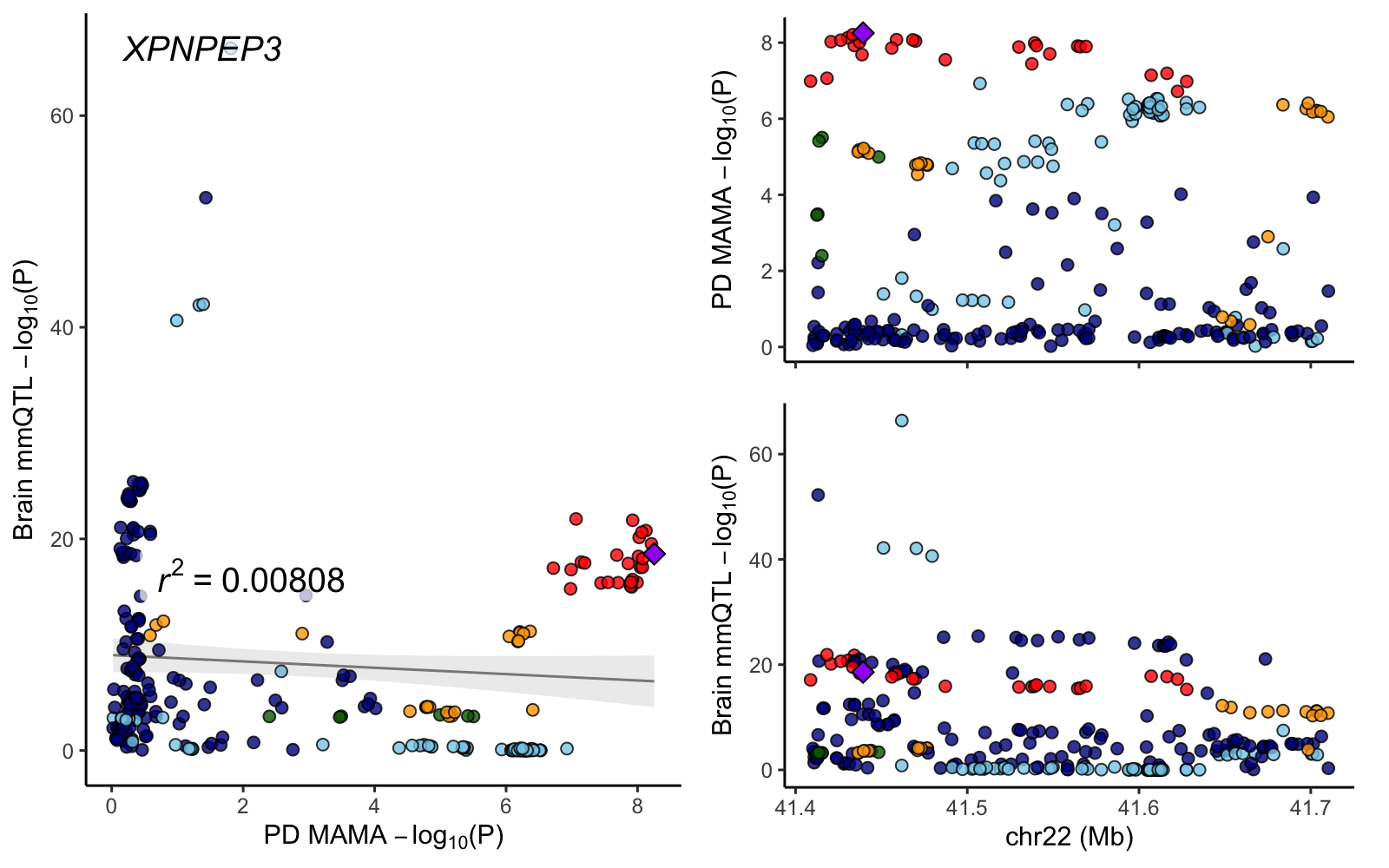

### ZC3H7B.png

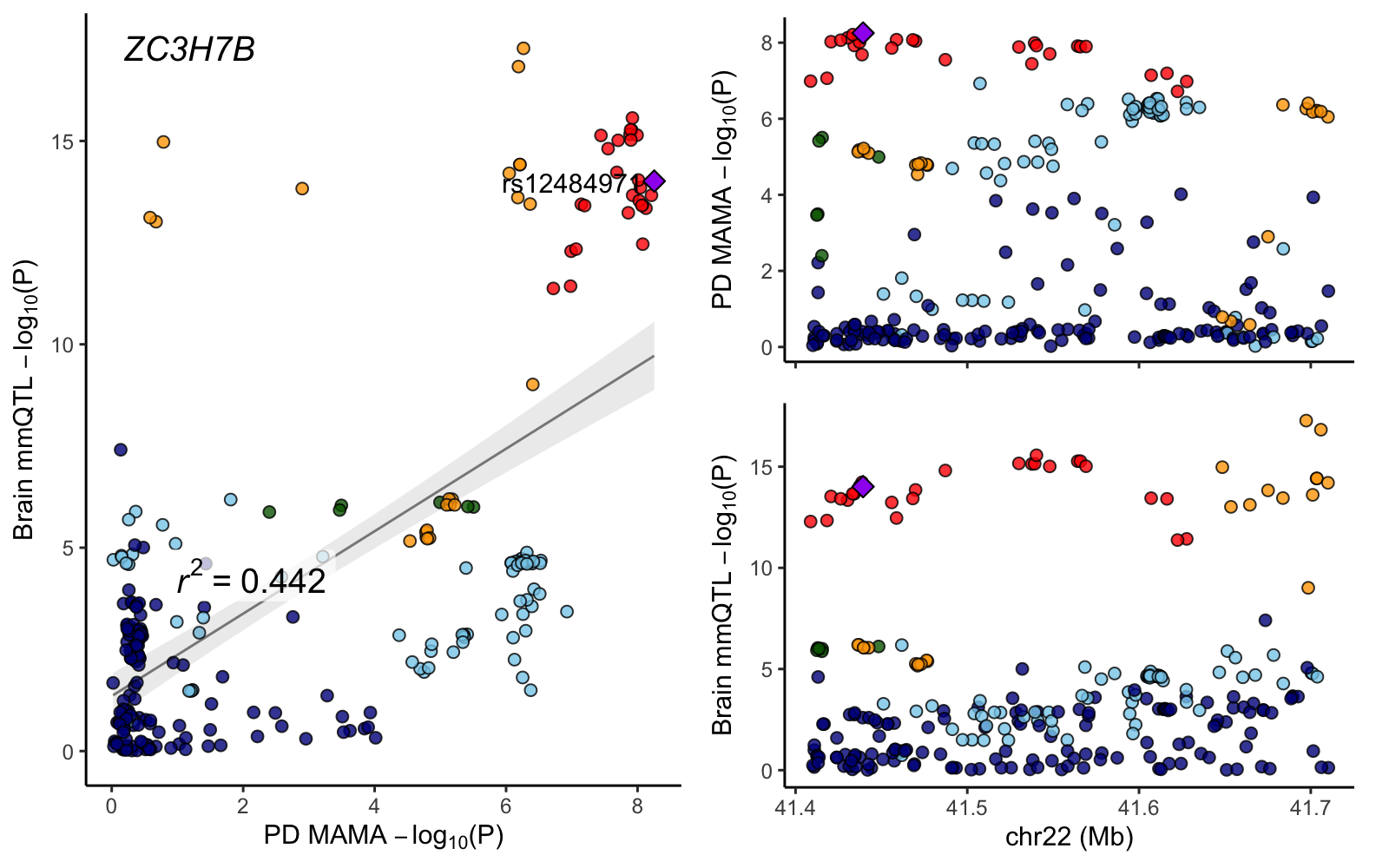

### ZSWIM7.png

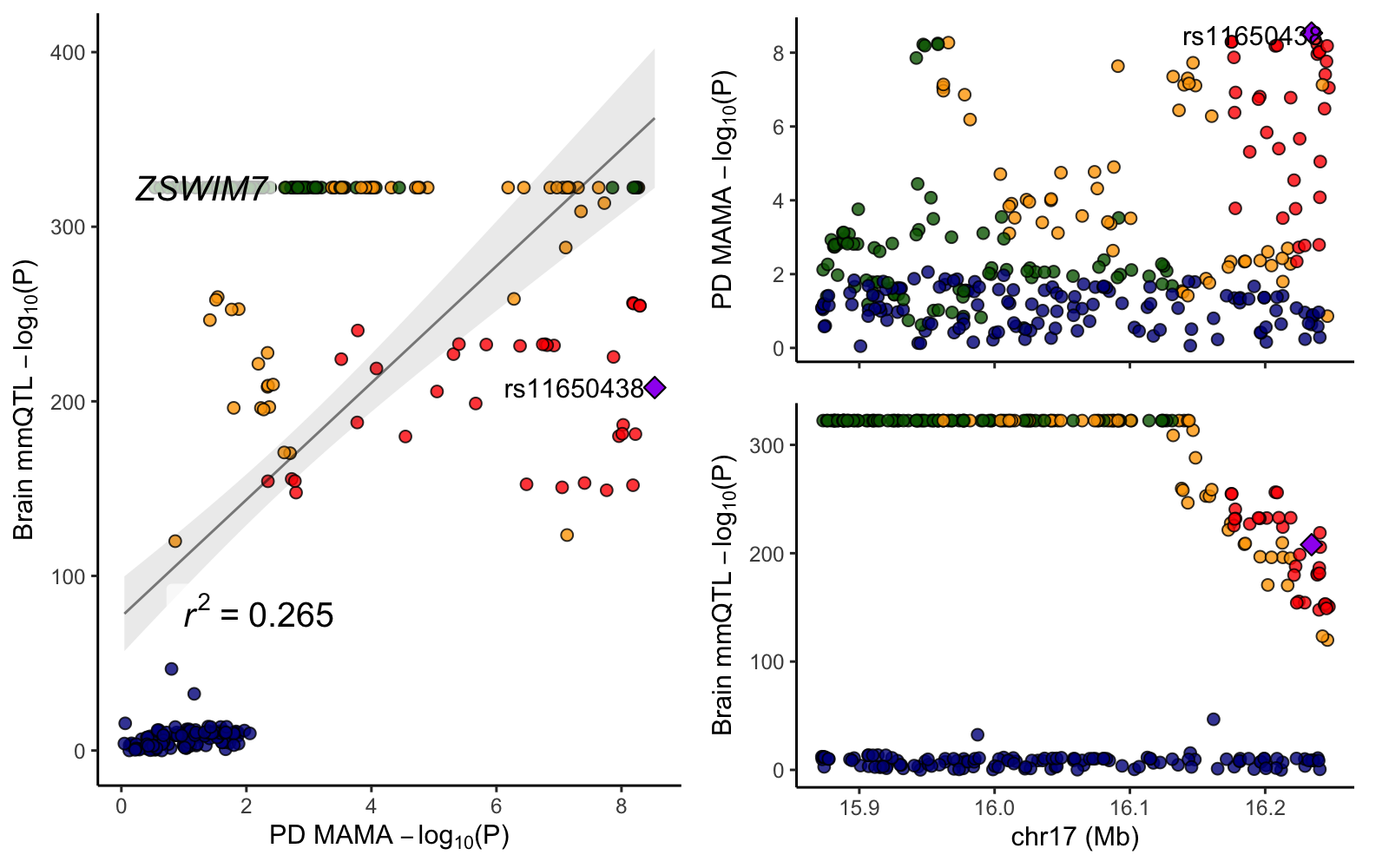
