## Supplementary figures and images for "Multi-ancestry genome-wide meta-analysis in Parkinson’s disease"

### chr1_93302187-93802187.png

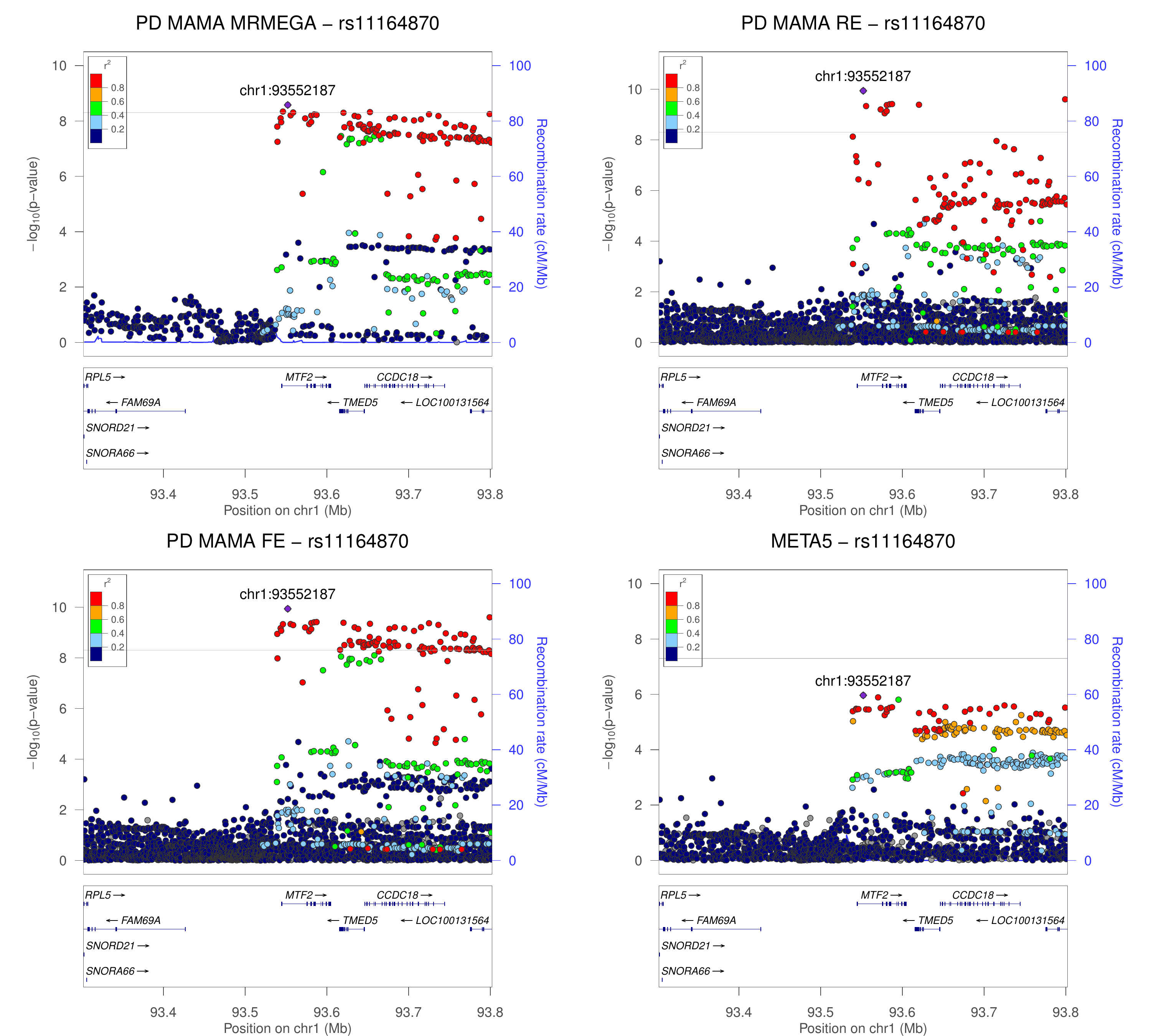

### chr1_154648185-155148185.png

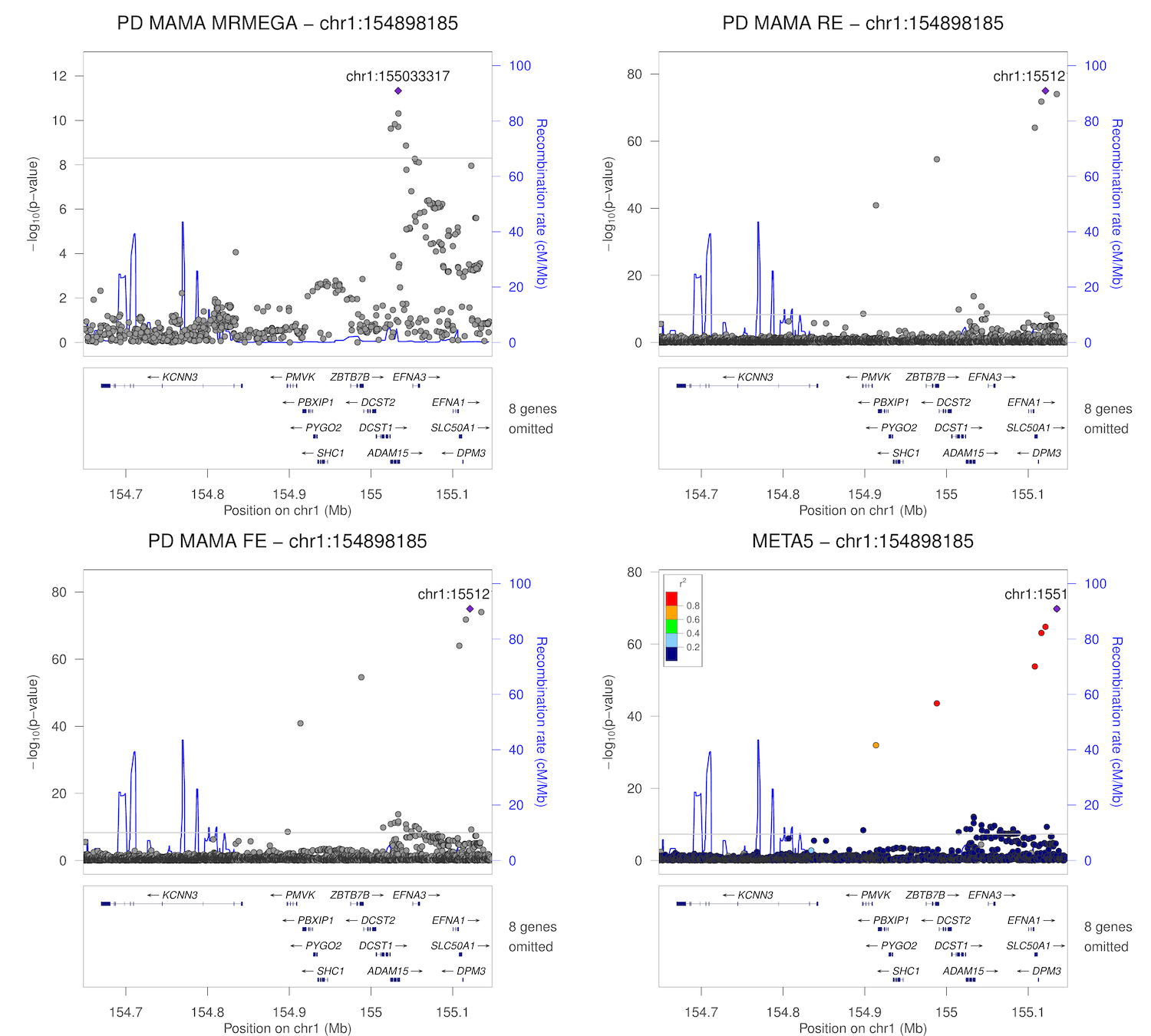

### chr1_154885036-155385036.png

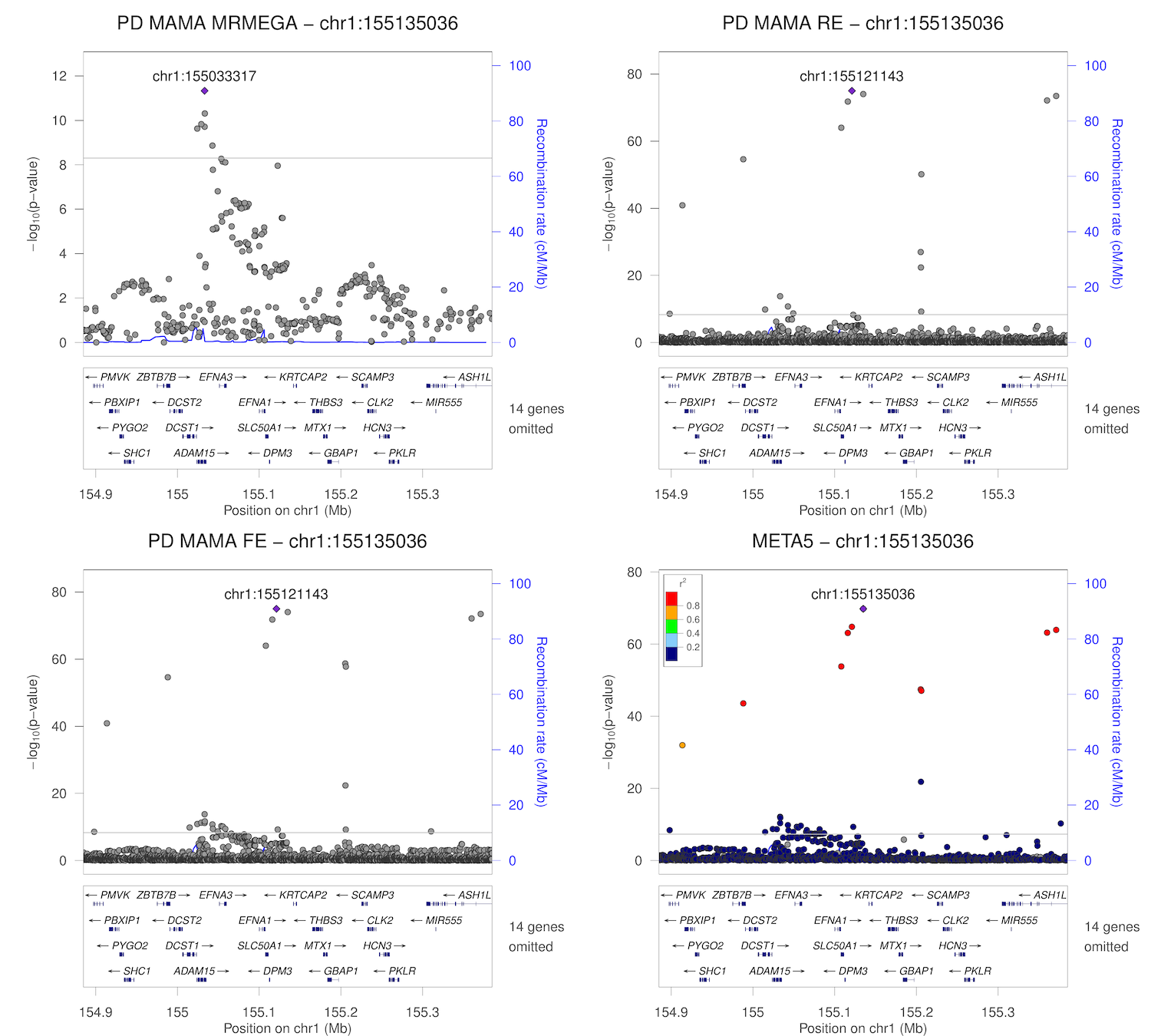

### chr1_154955634-155455634.png

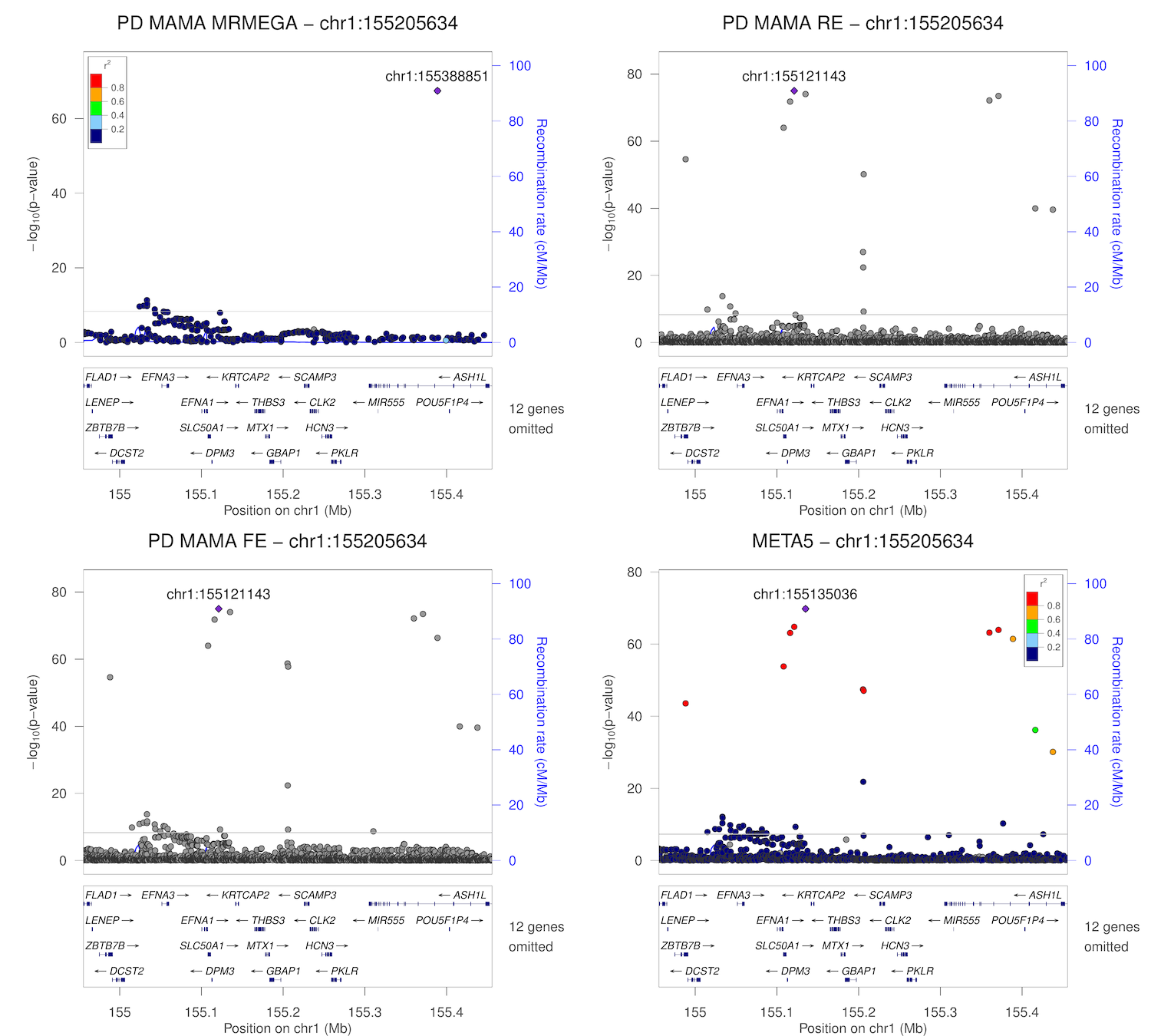

### chr1_161219054-161719054.png

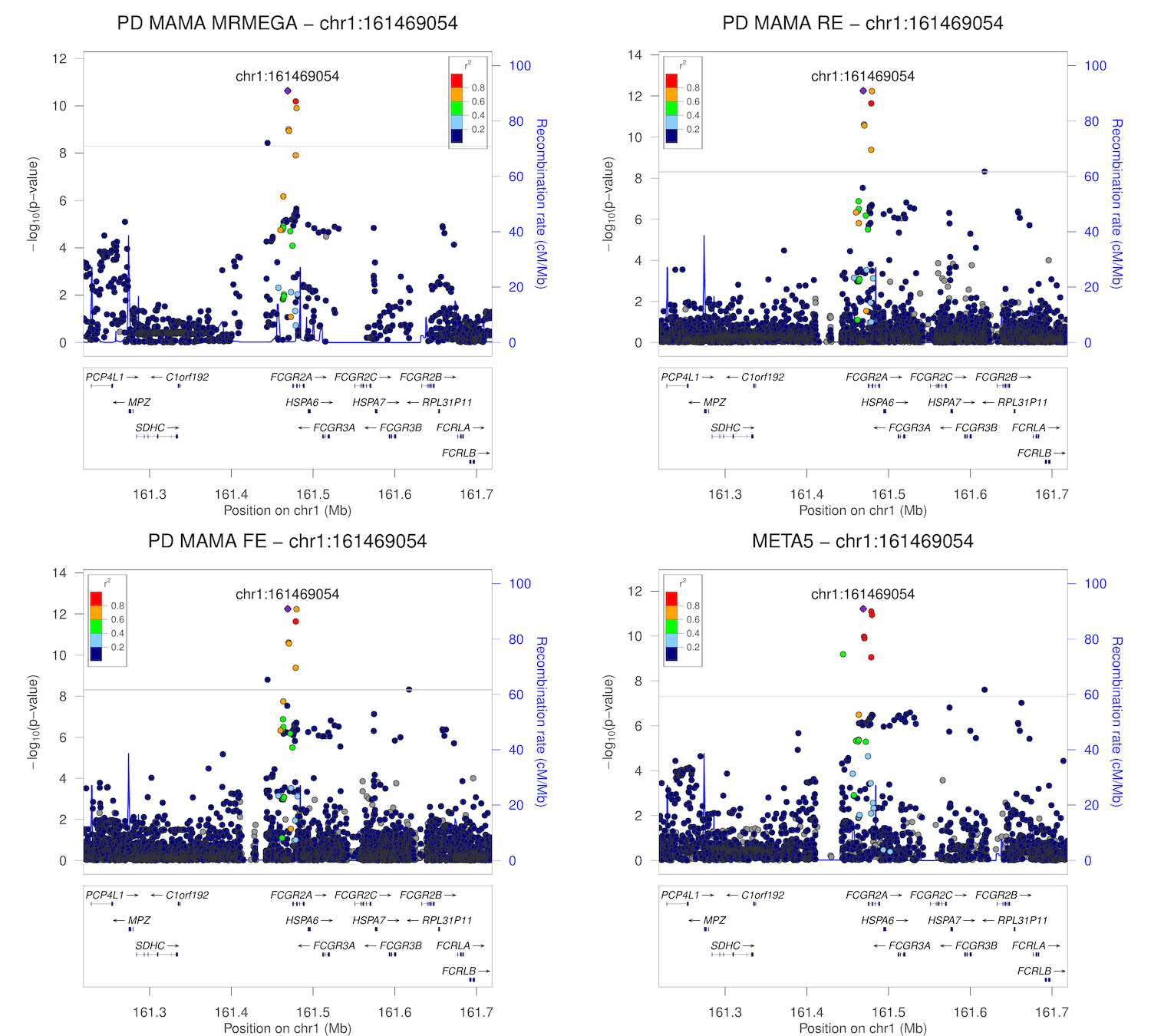

### chr1_171469769-171969769.png

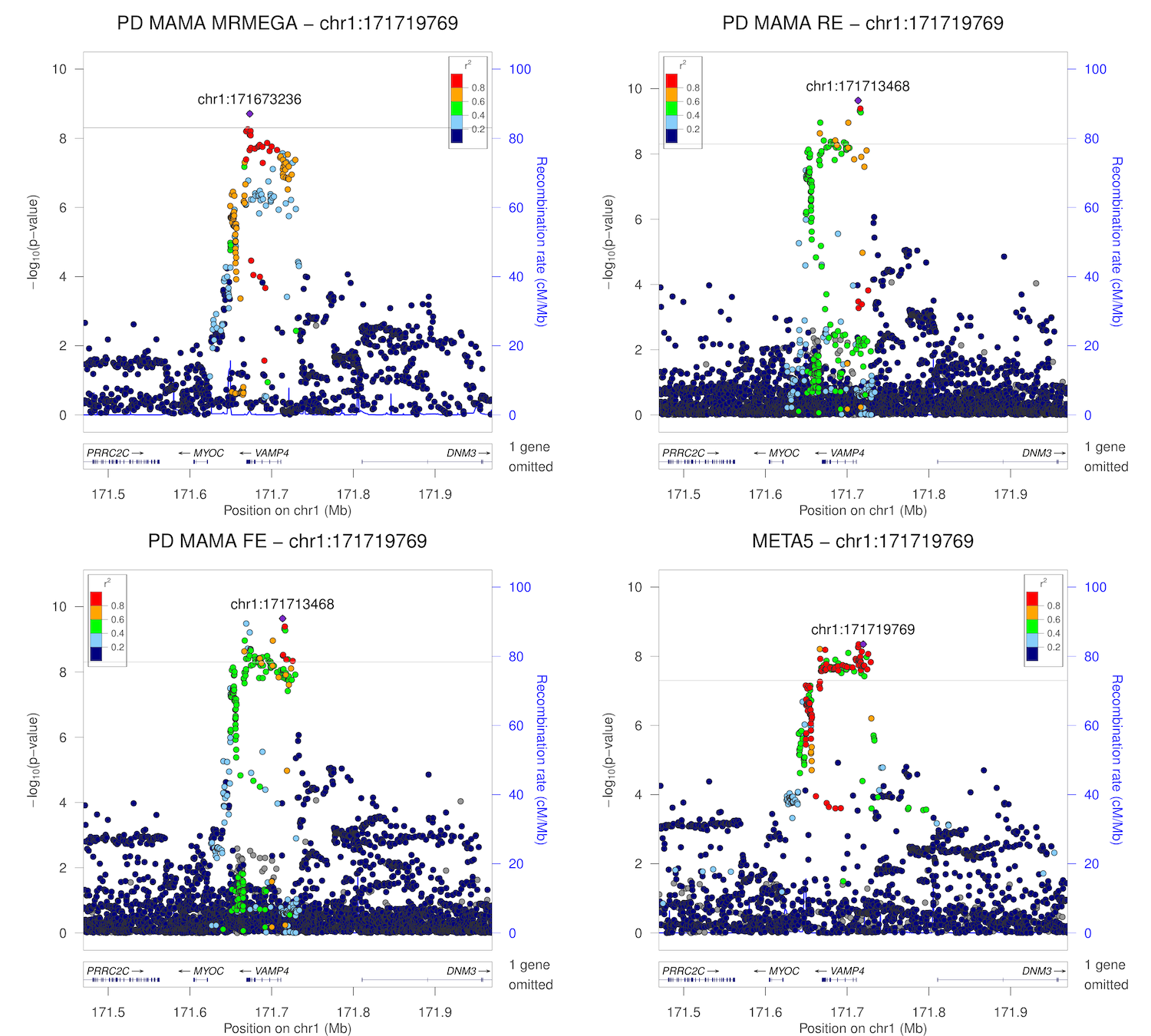

### chr1_205473572-205973572.png

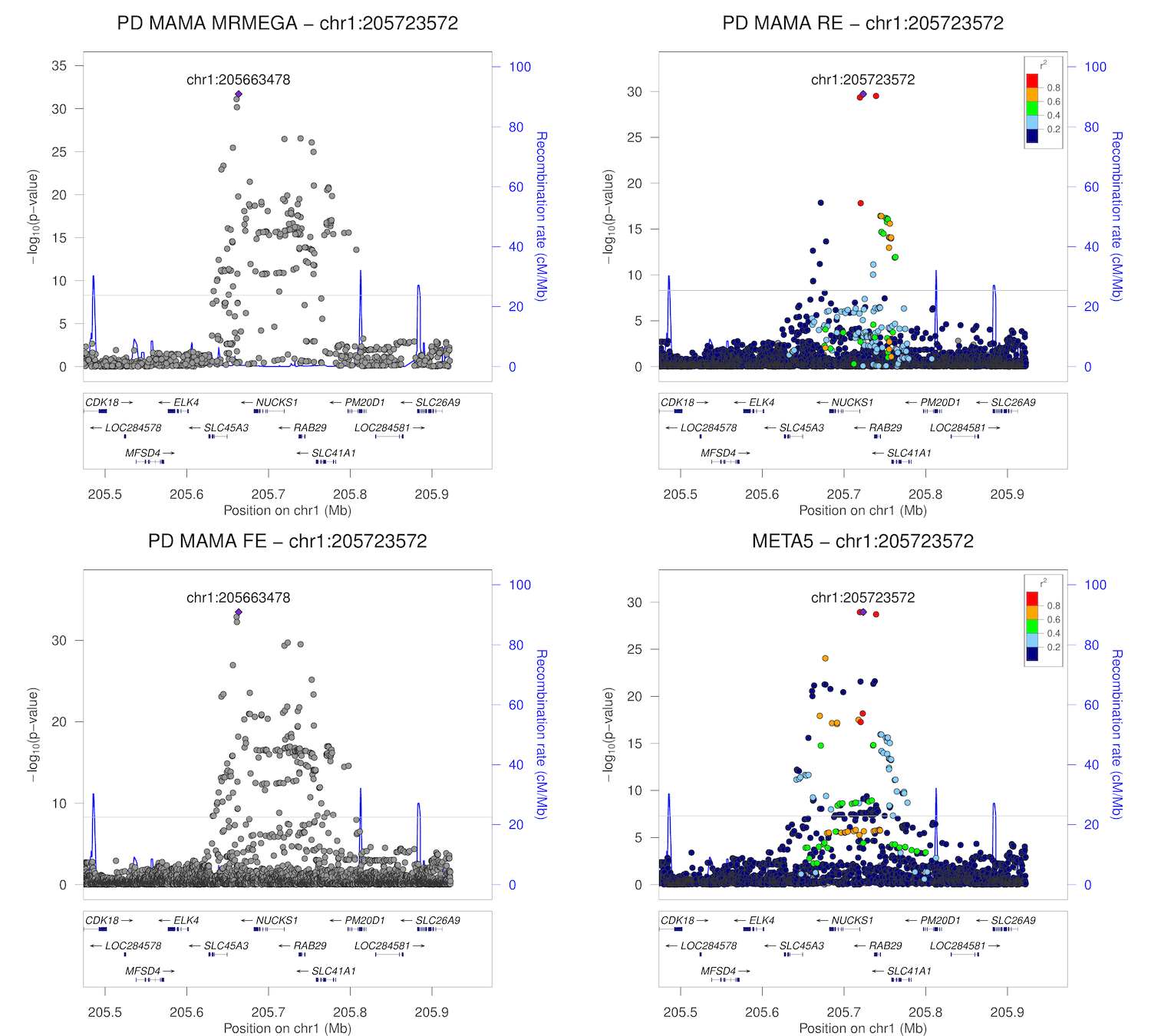

### chr1_205487739-205987739.png

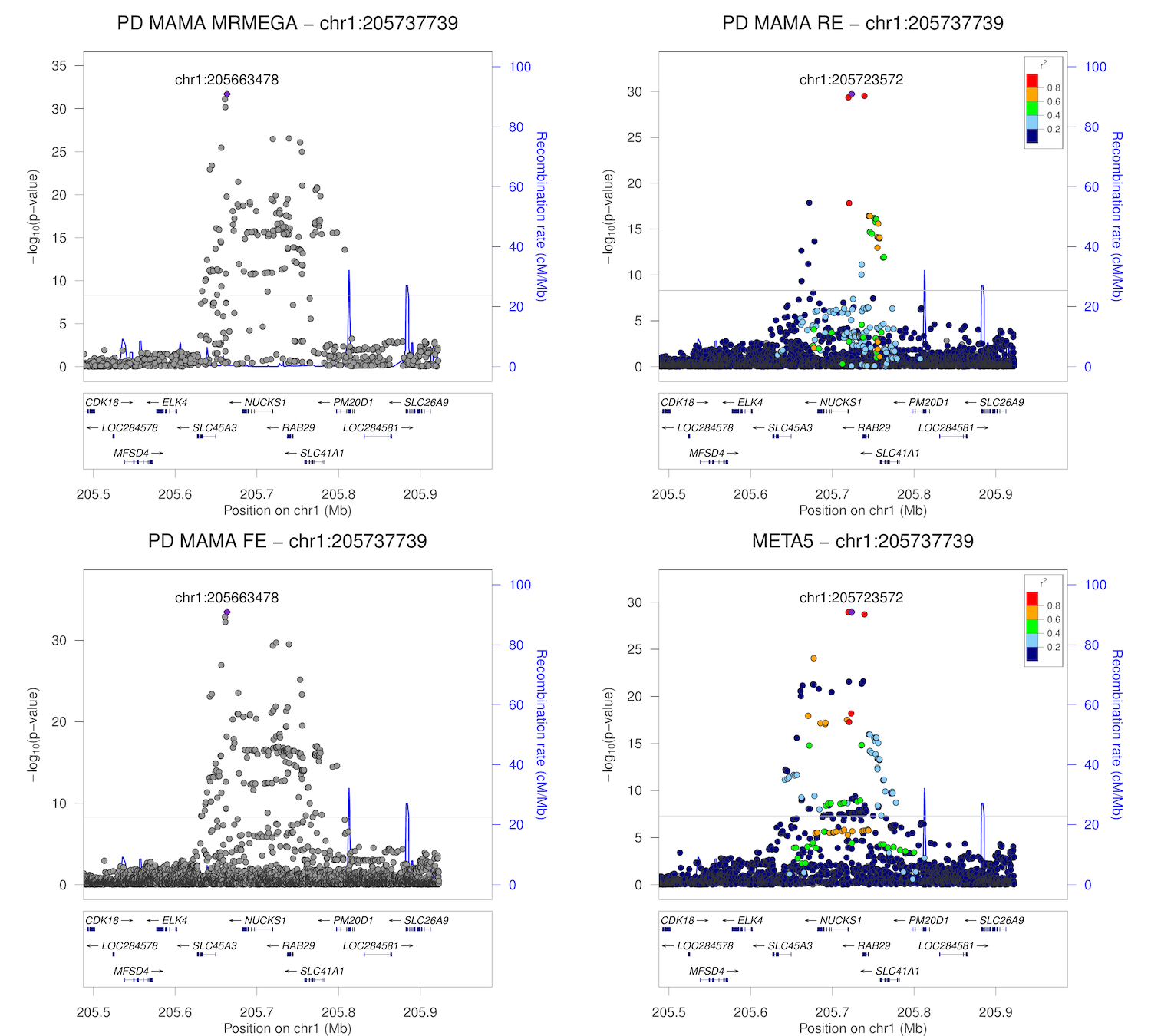

### chr1_226666078-227166078.png

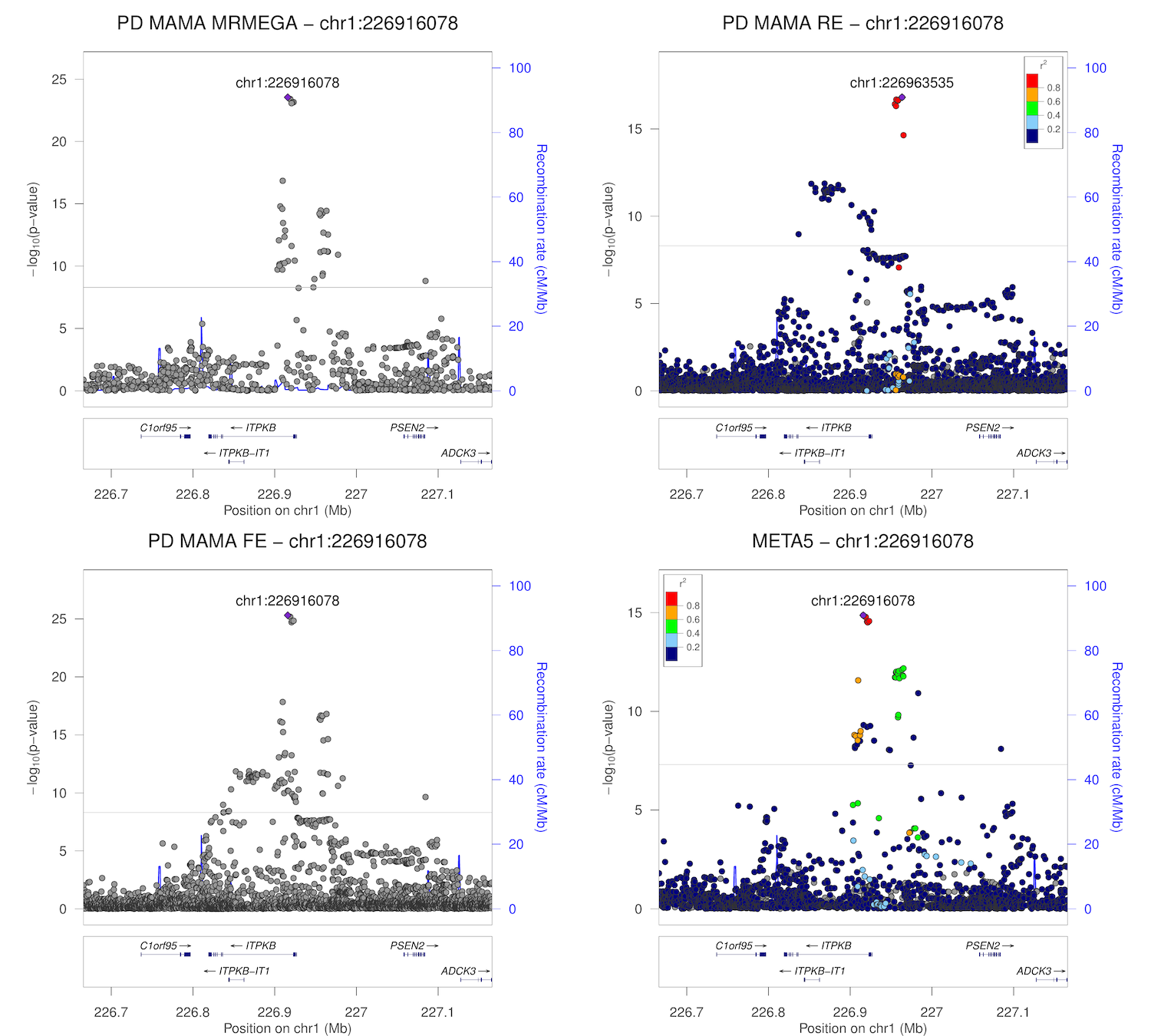

### chr1_232414611-232914611.png

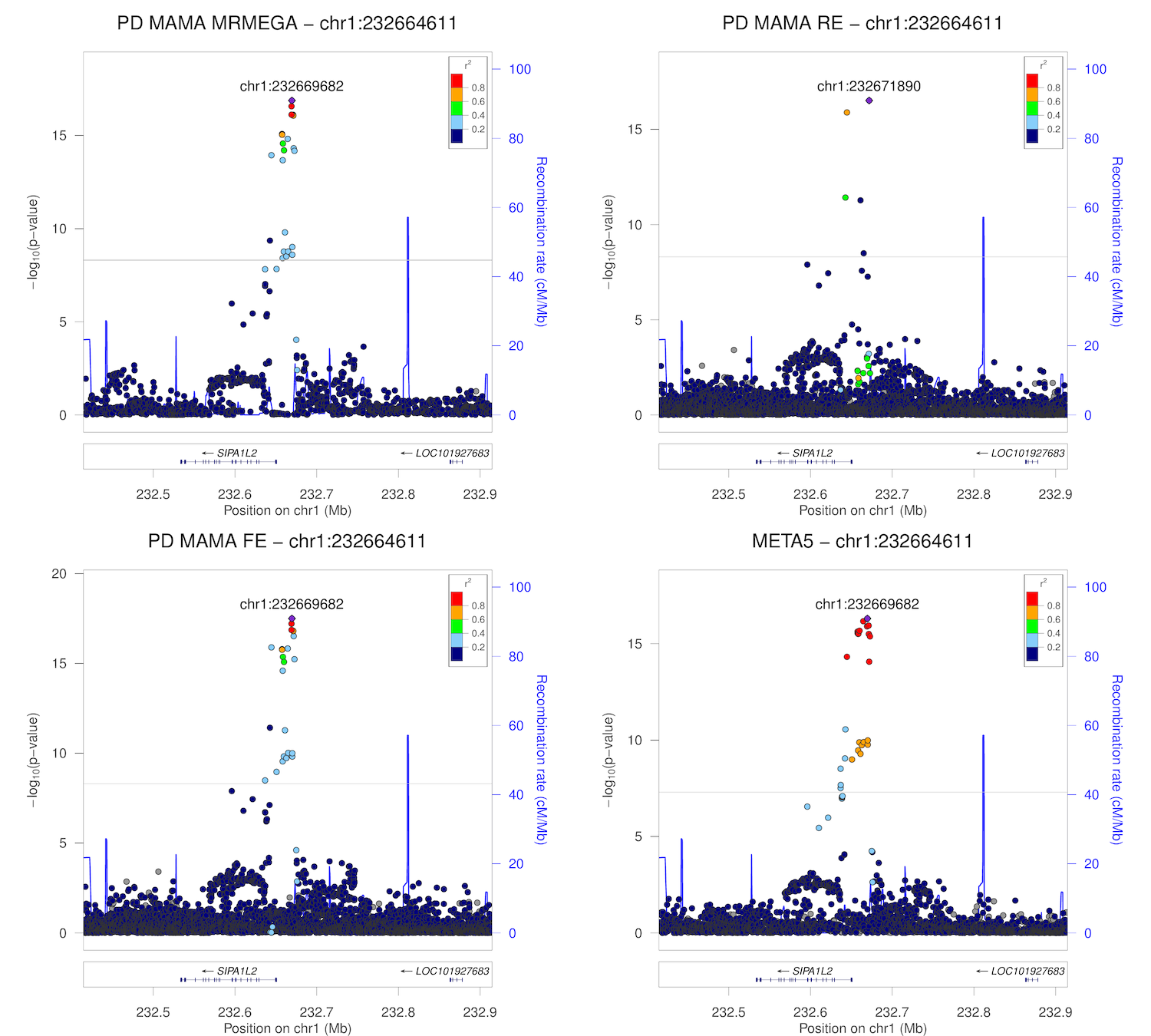

### chr2_17897848-18397848.png

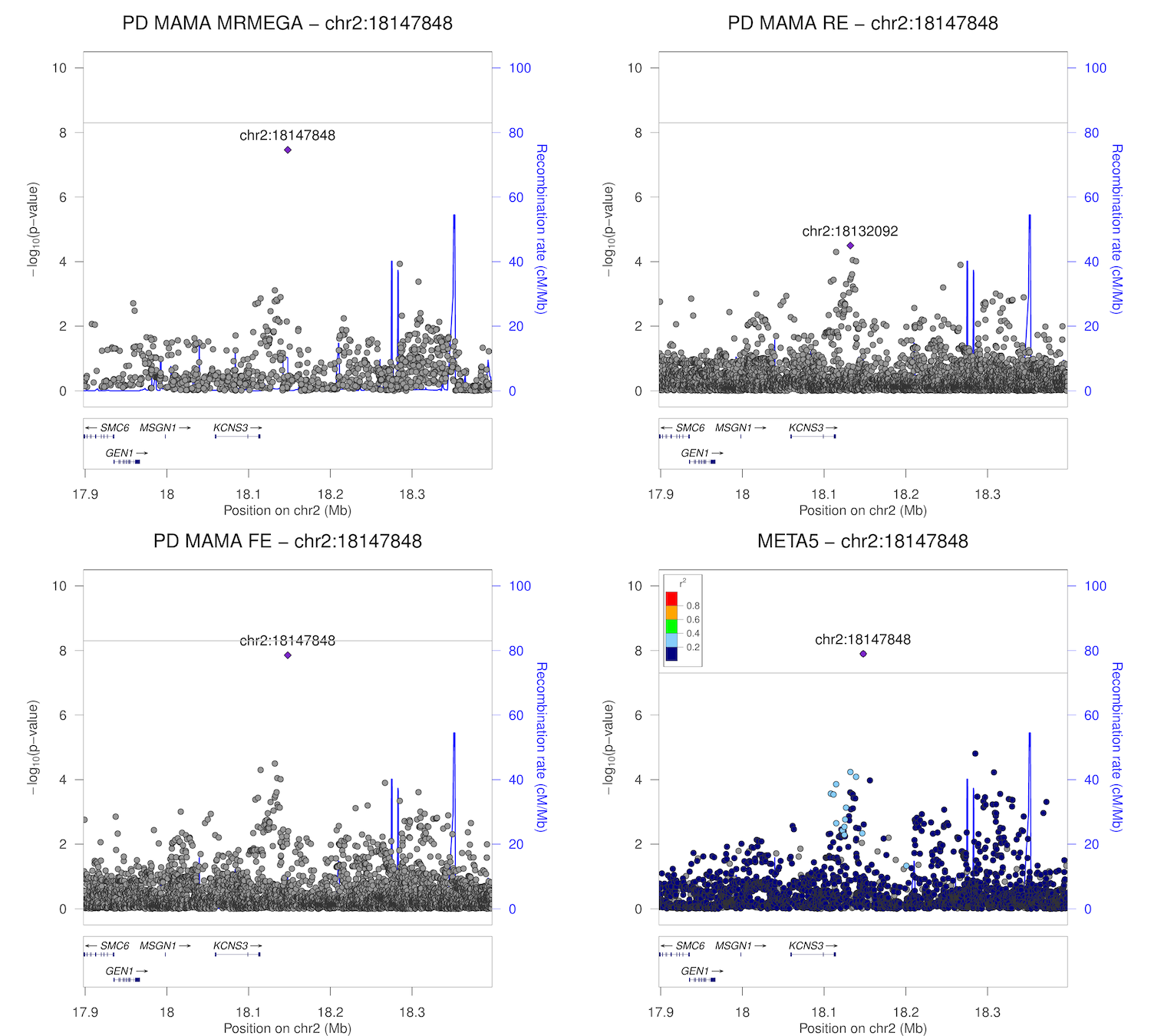

### chr2_95750943-96250943.png

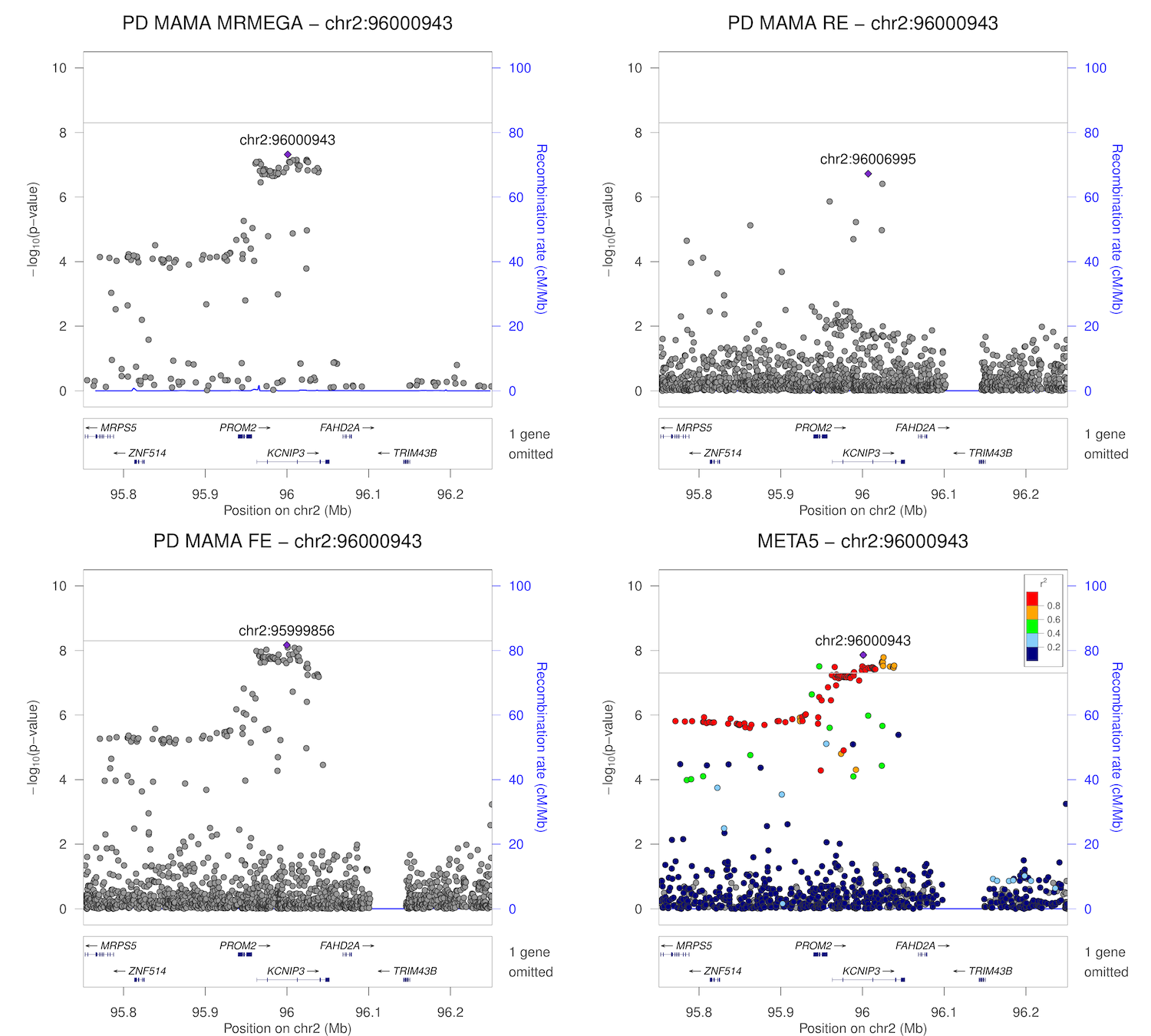

### chr2_102146963-102646963.png

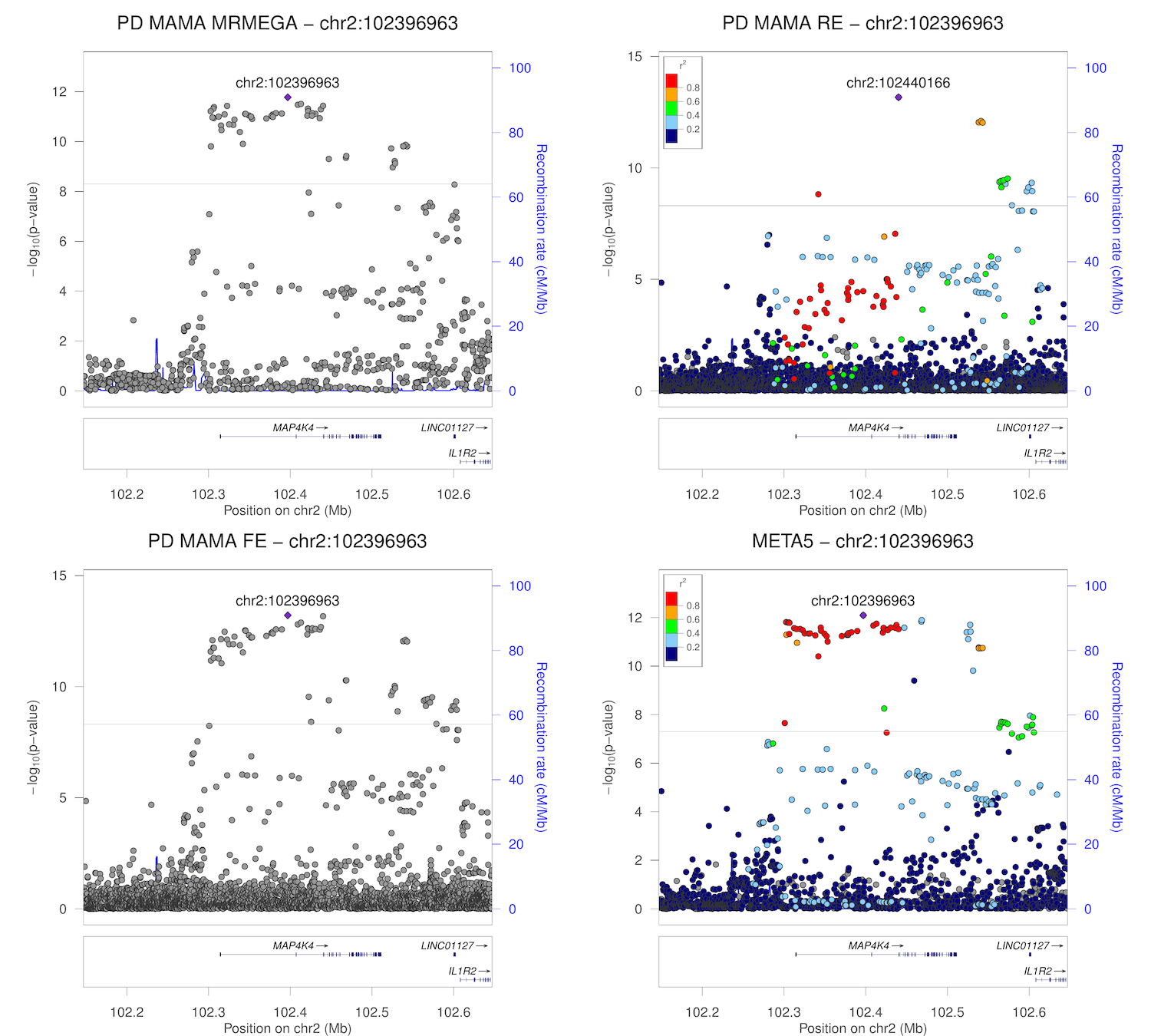

### chr2_135214616-135714616.png

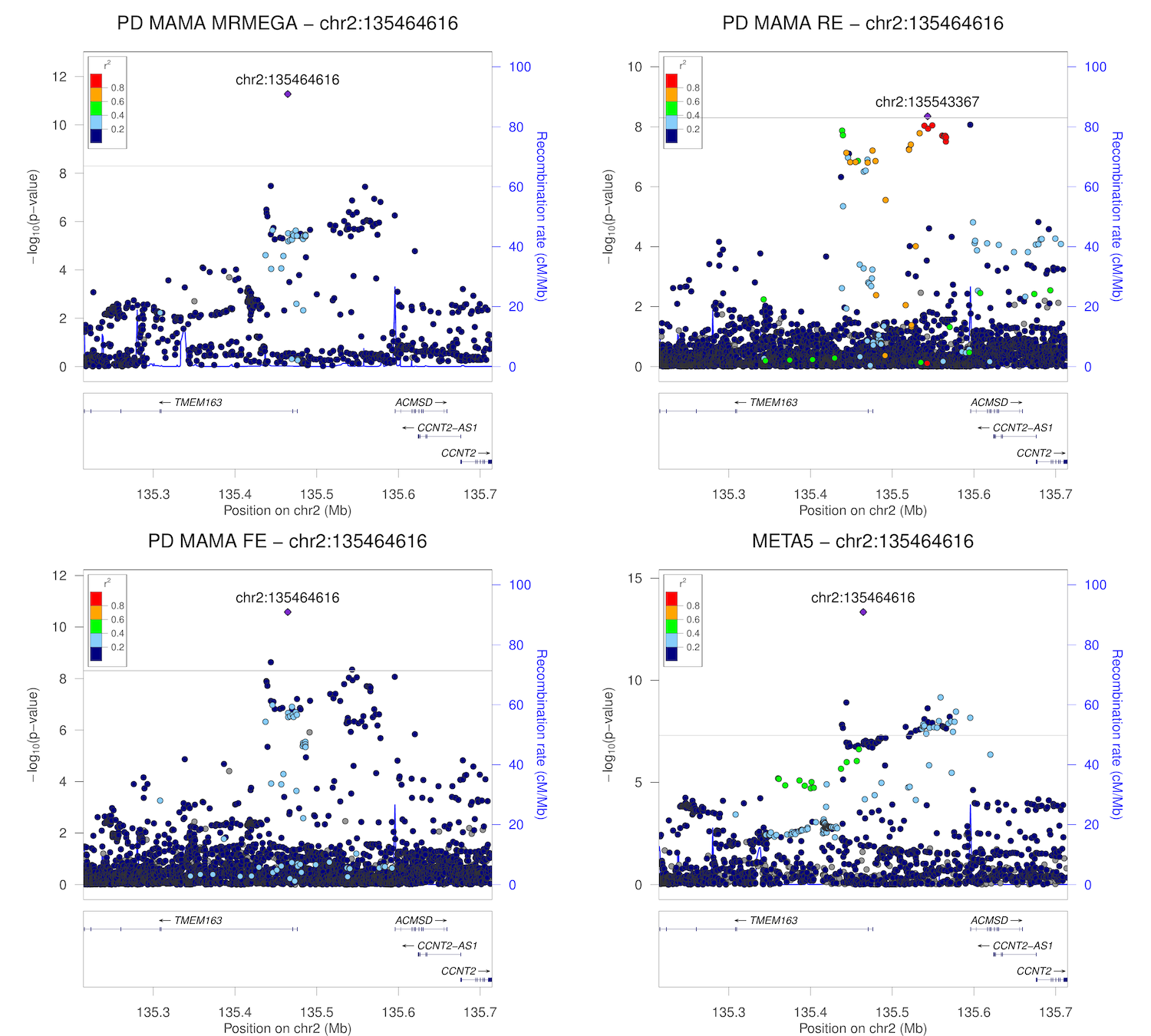

### chr3_28455690-28955690.png

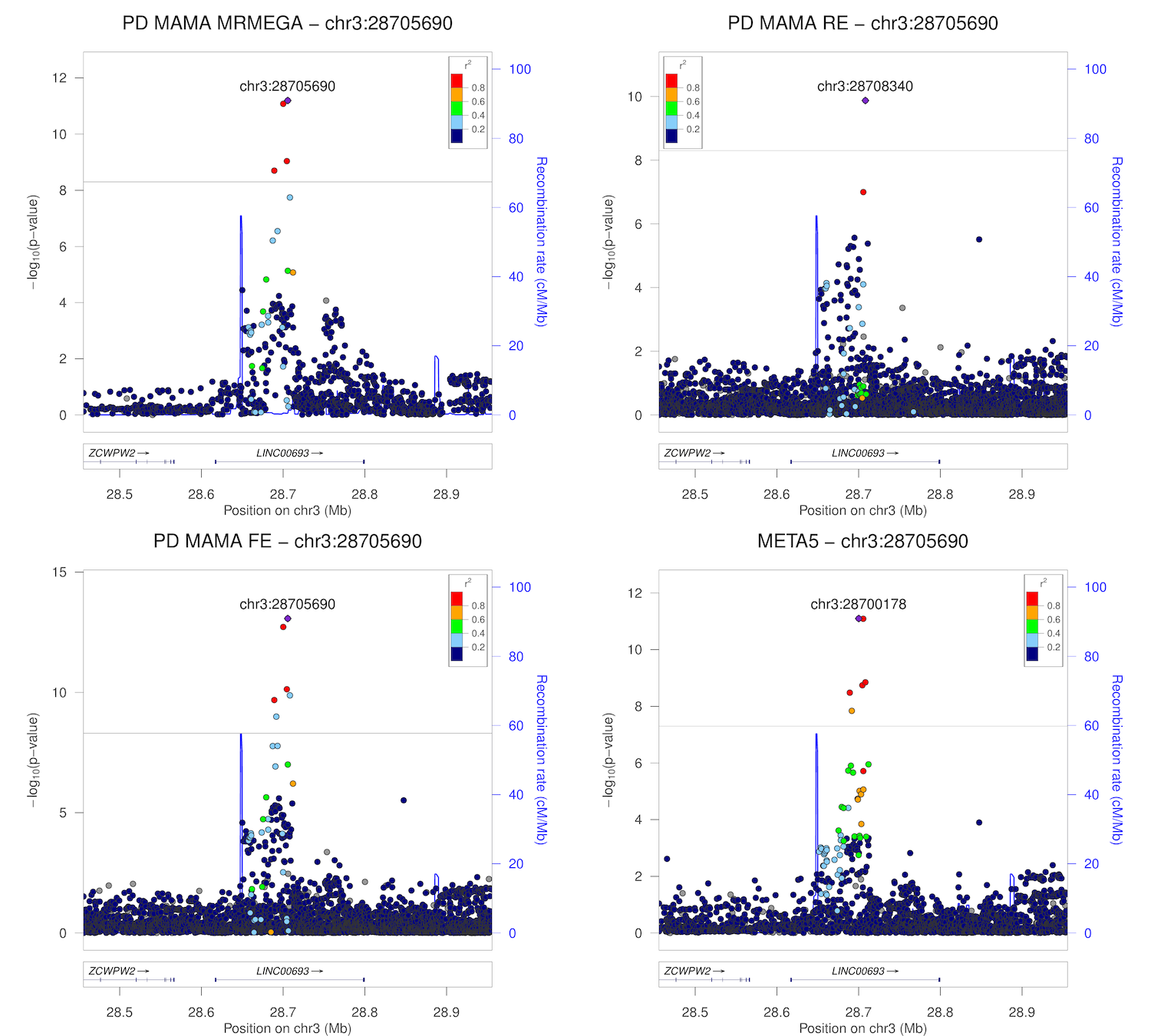

### chr3_48498989-48998989.png

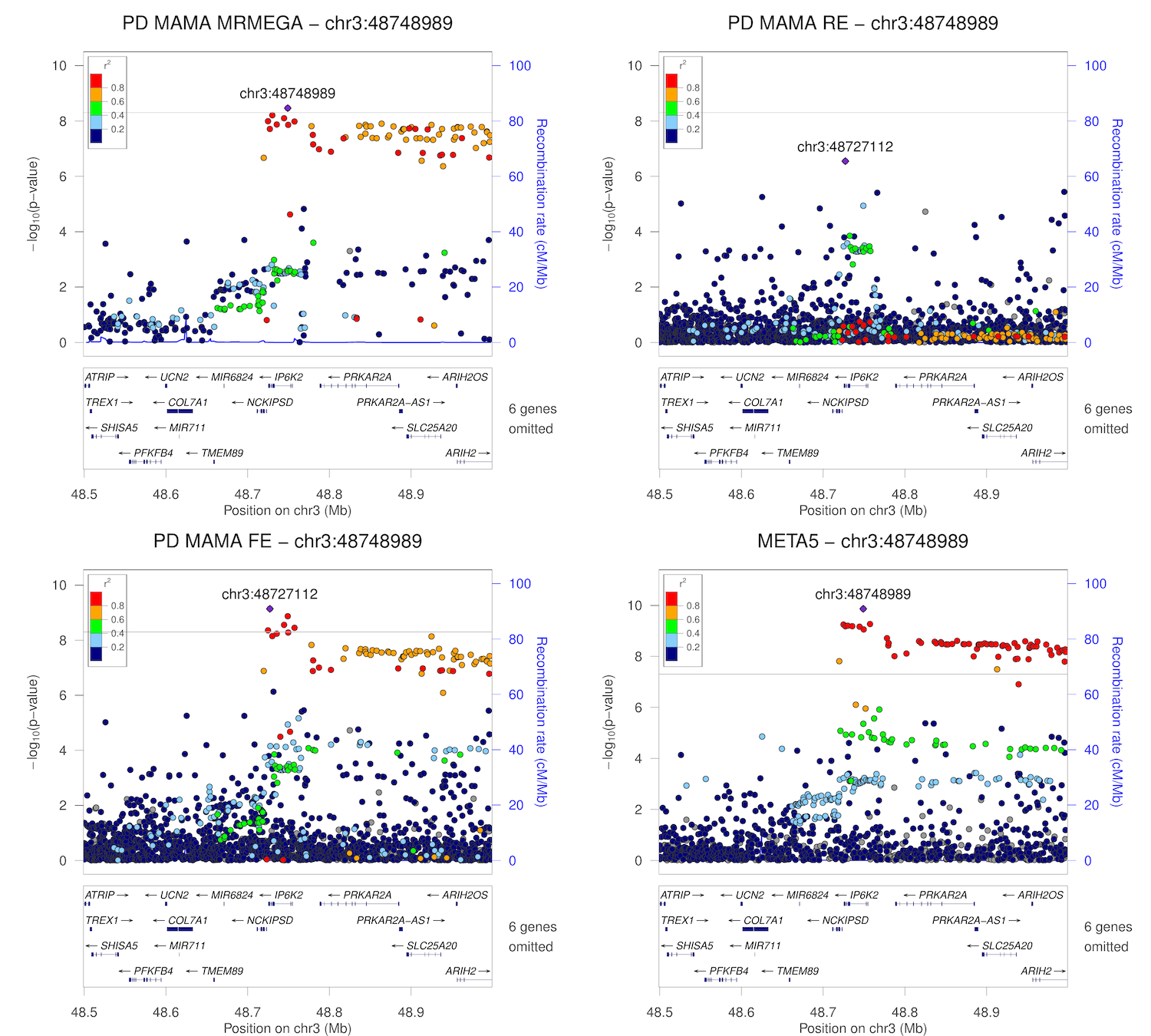

### chr3_121946892-122446892.png

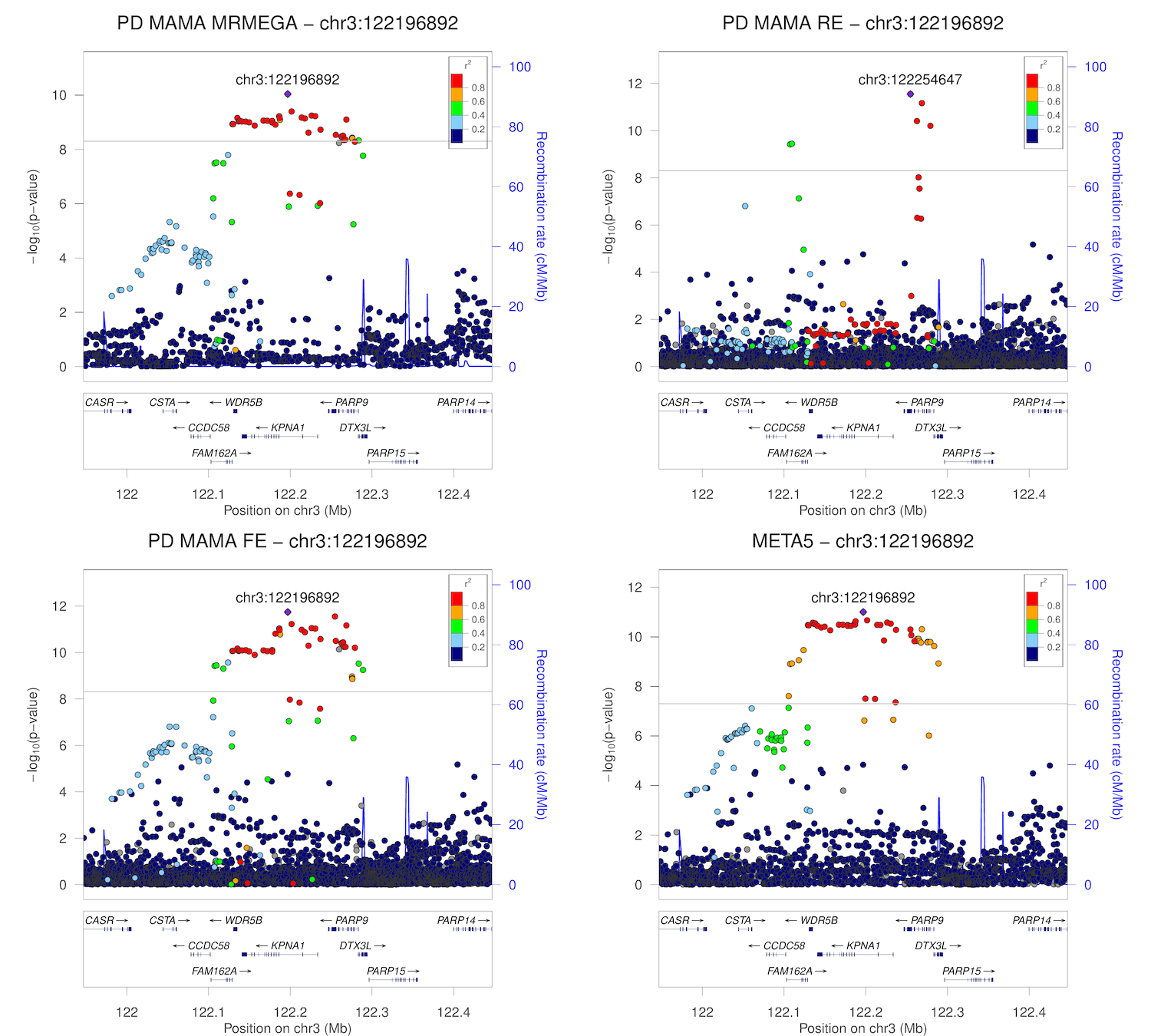

### chr3_150858965-151358965.png

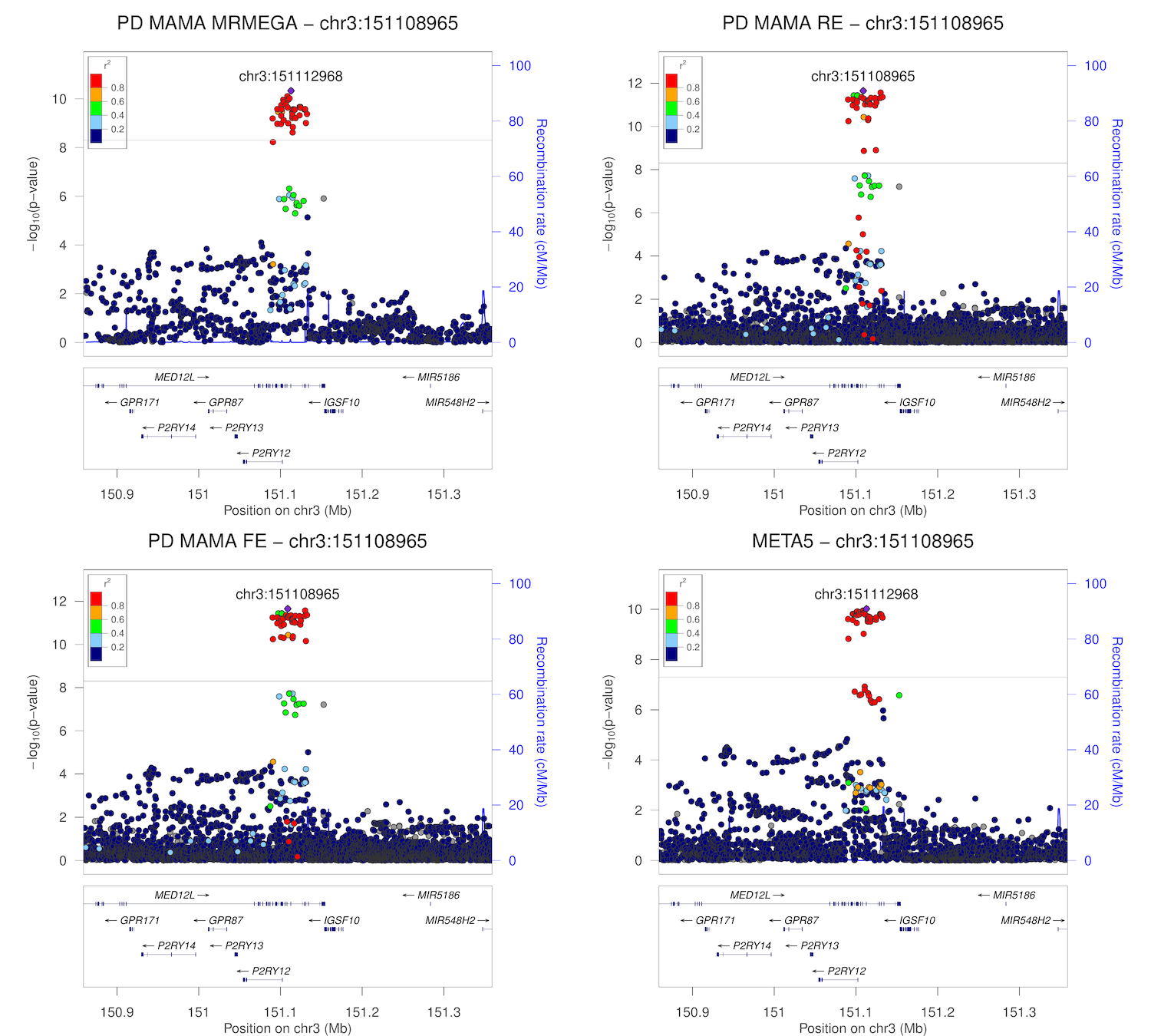

### chr3_160827630-161327630.png

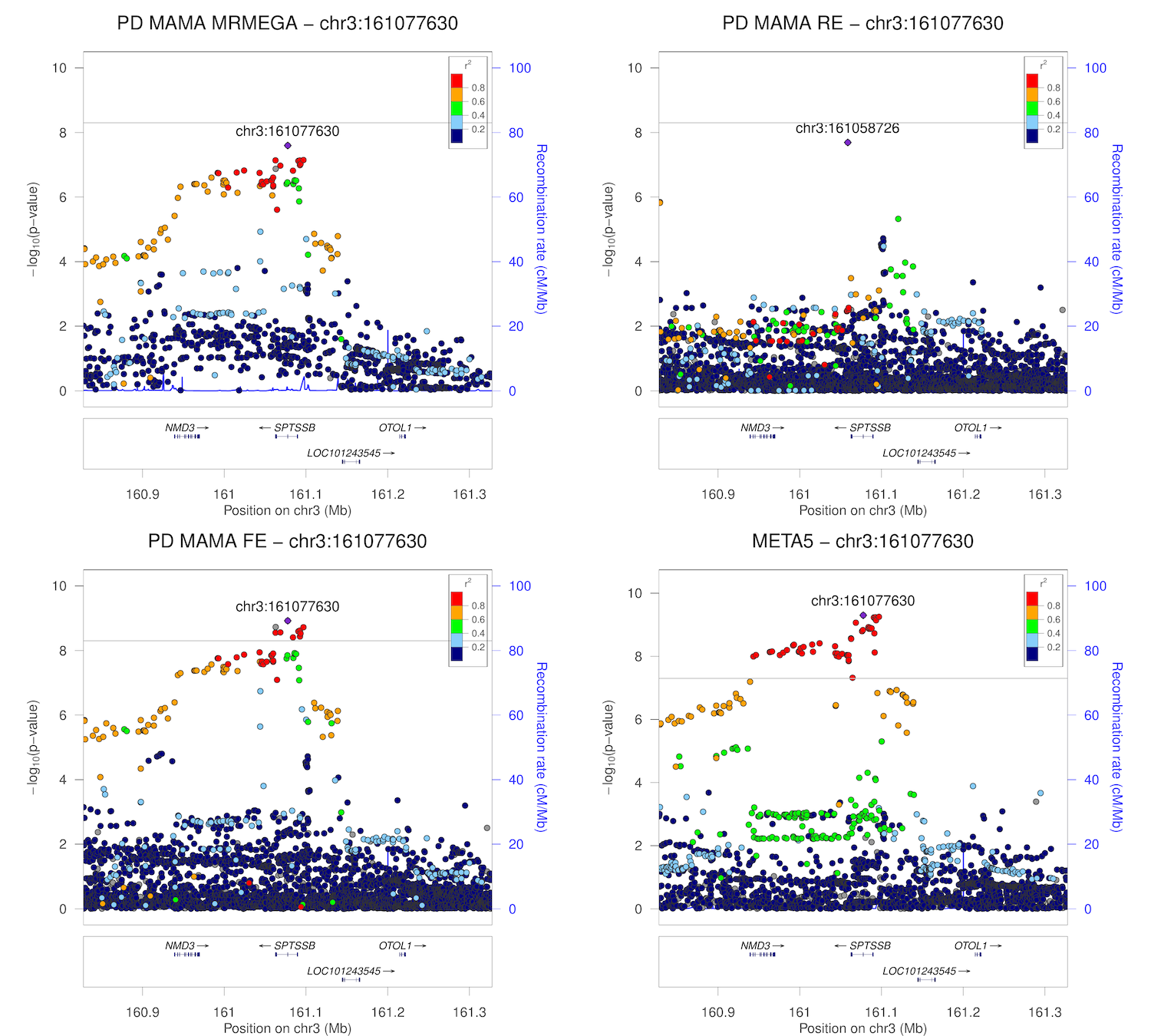

### chr3_178611417-179111417.png

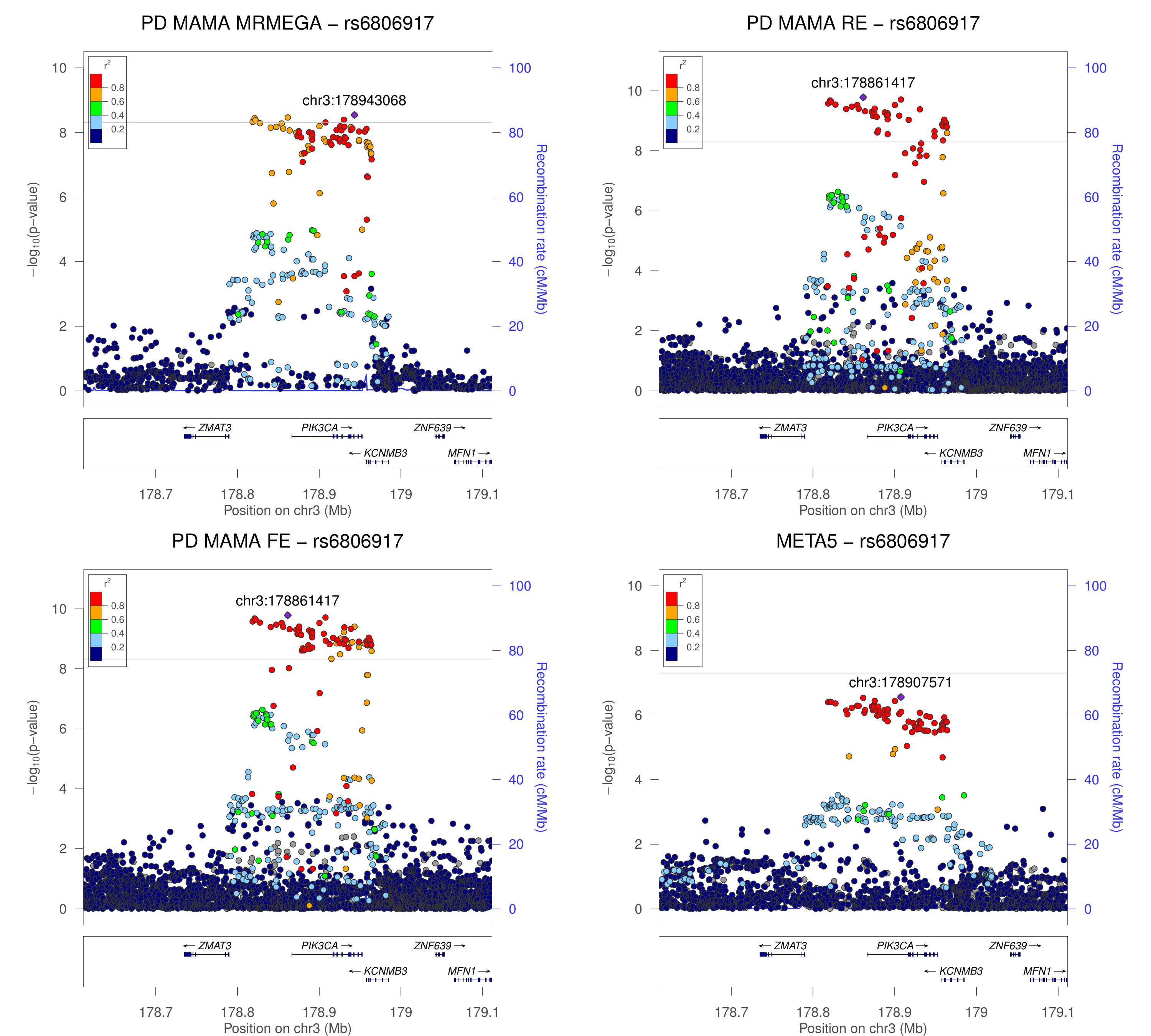

### chr3_182510073-183010073.png

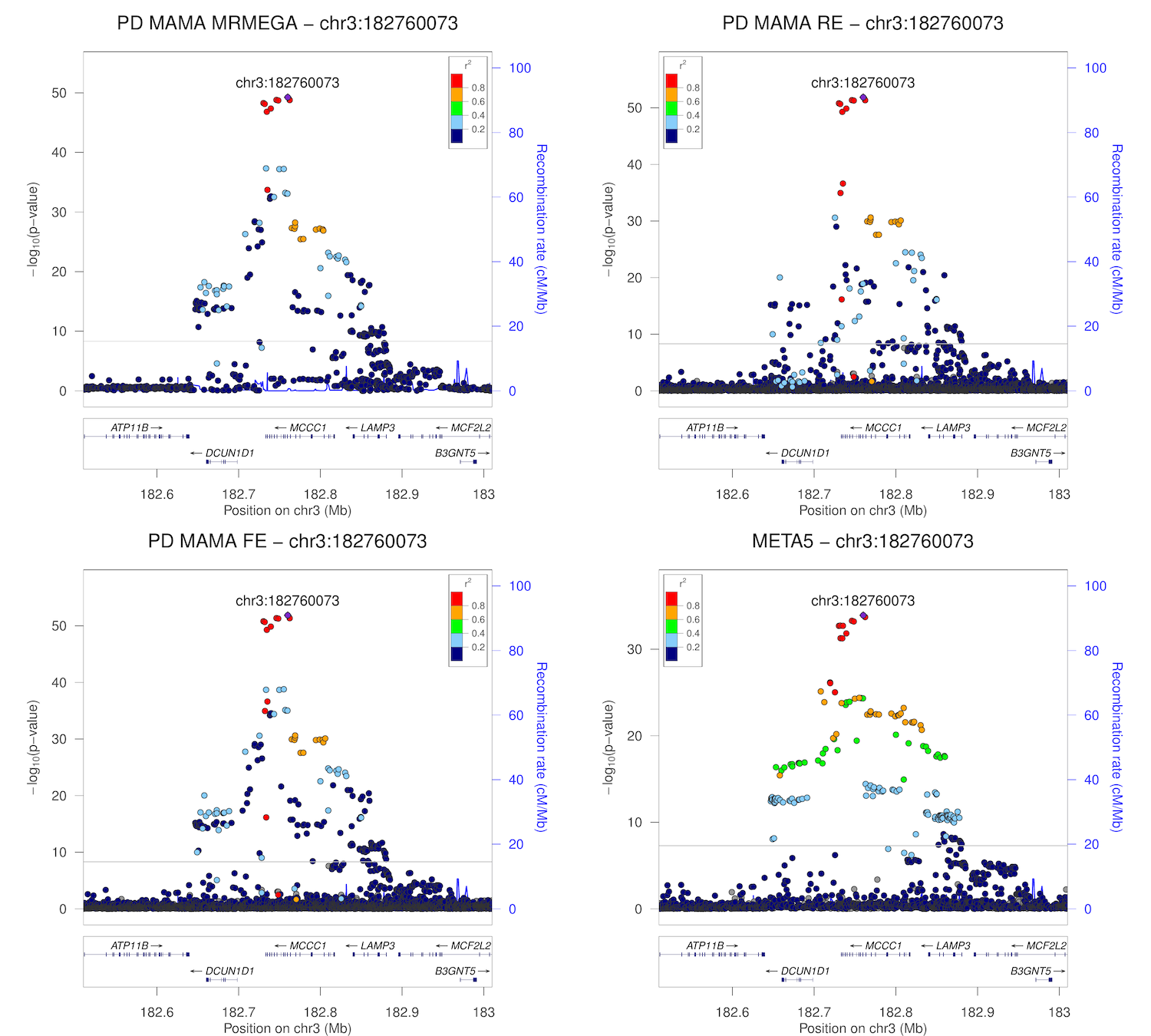

### chr4_675376-1175376.png

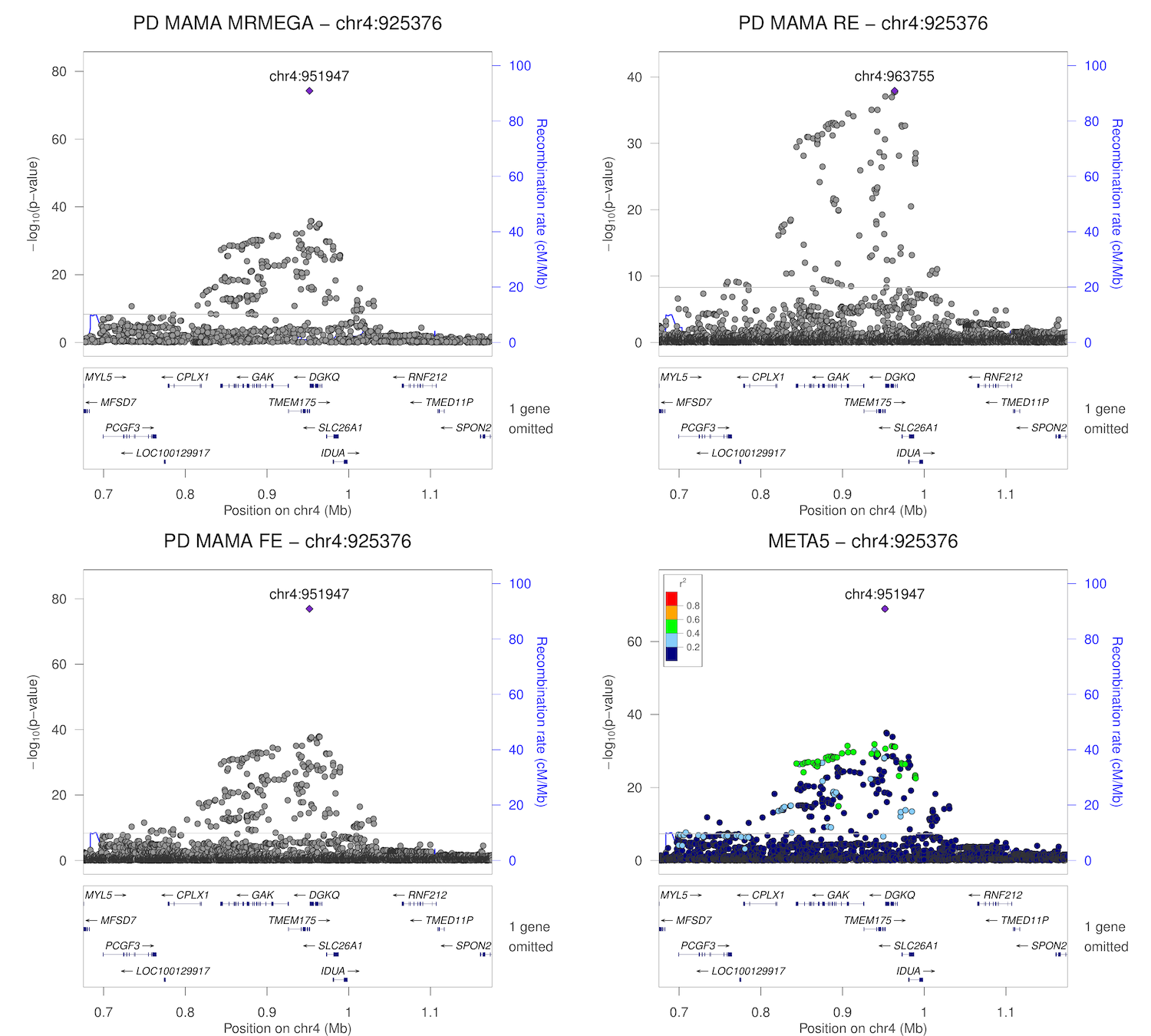

### chr4_701947-1201947.png

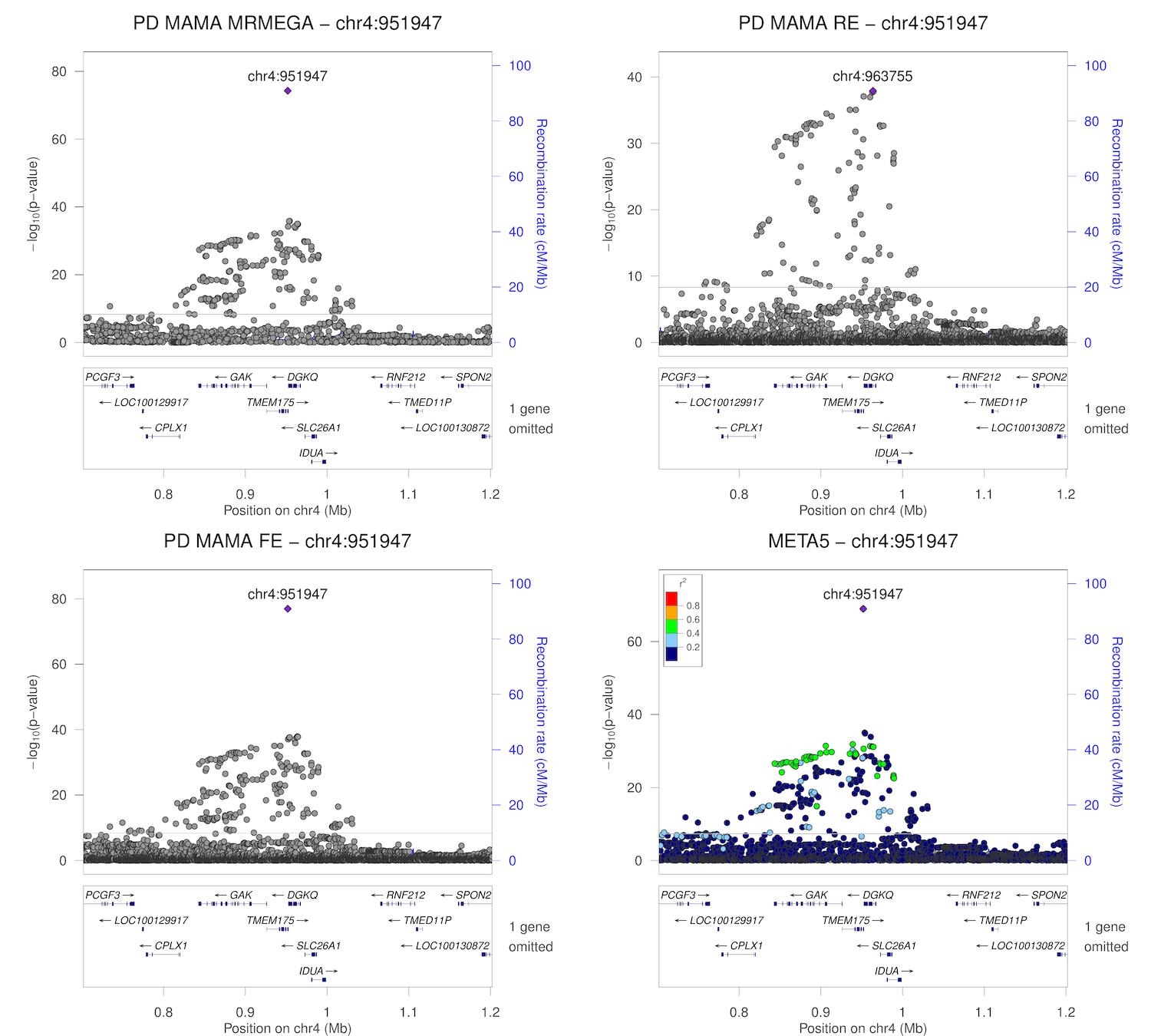

### chr4_2599168-3099168.png

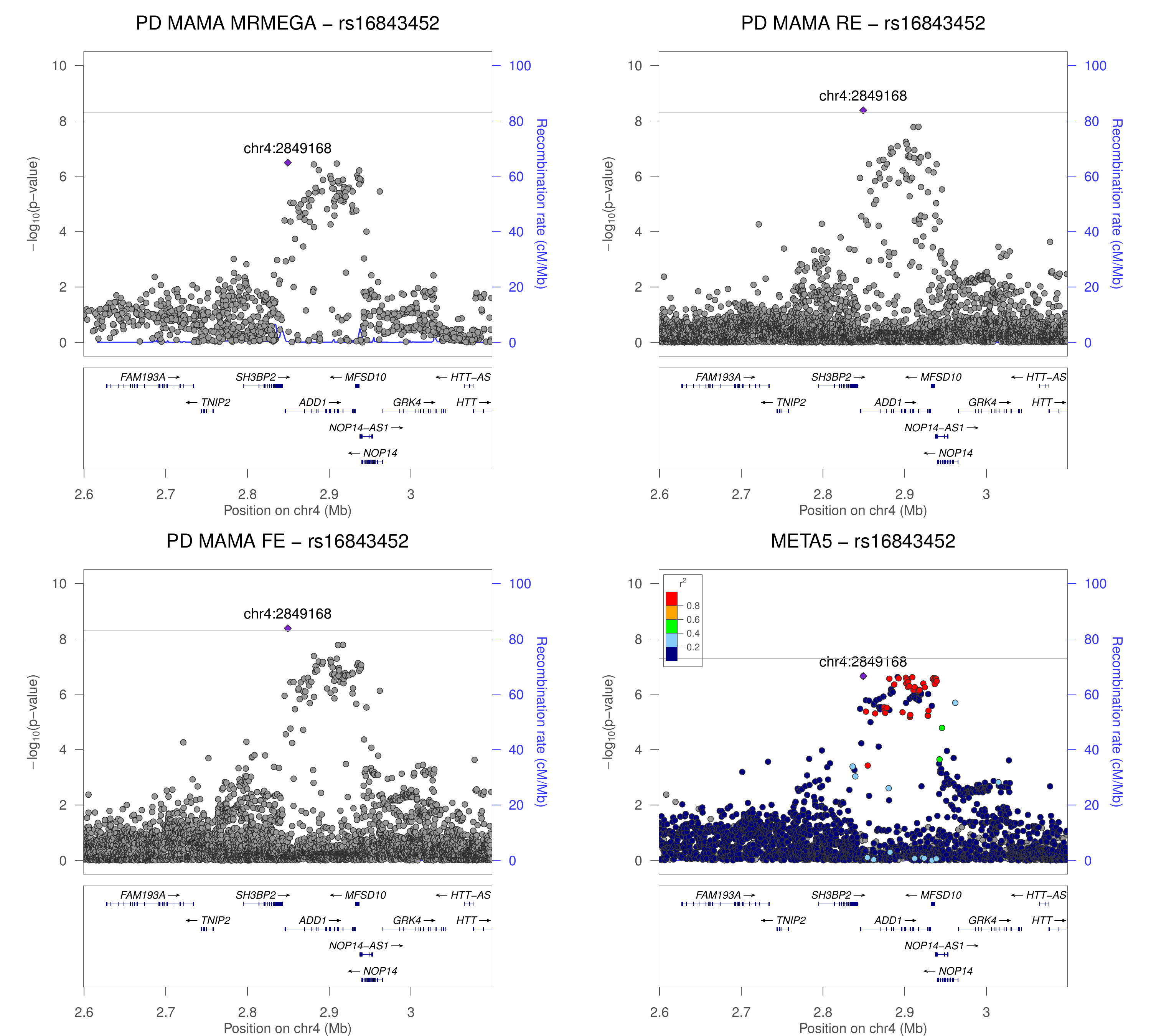

### chr4_15487348-15987348.png

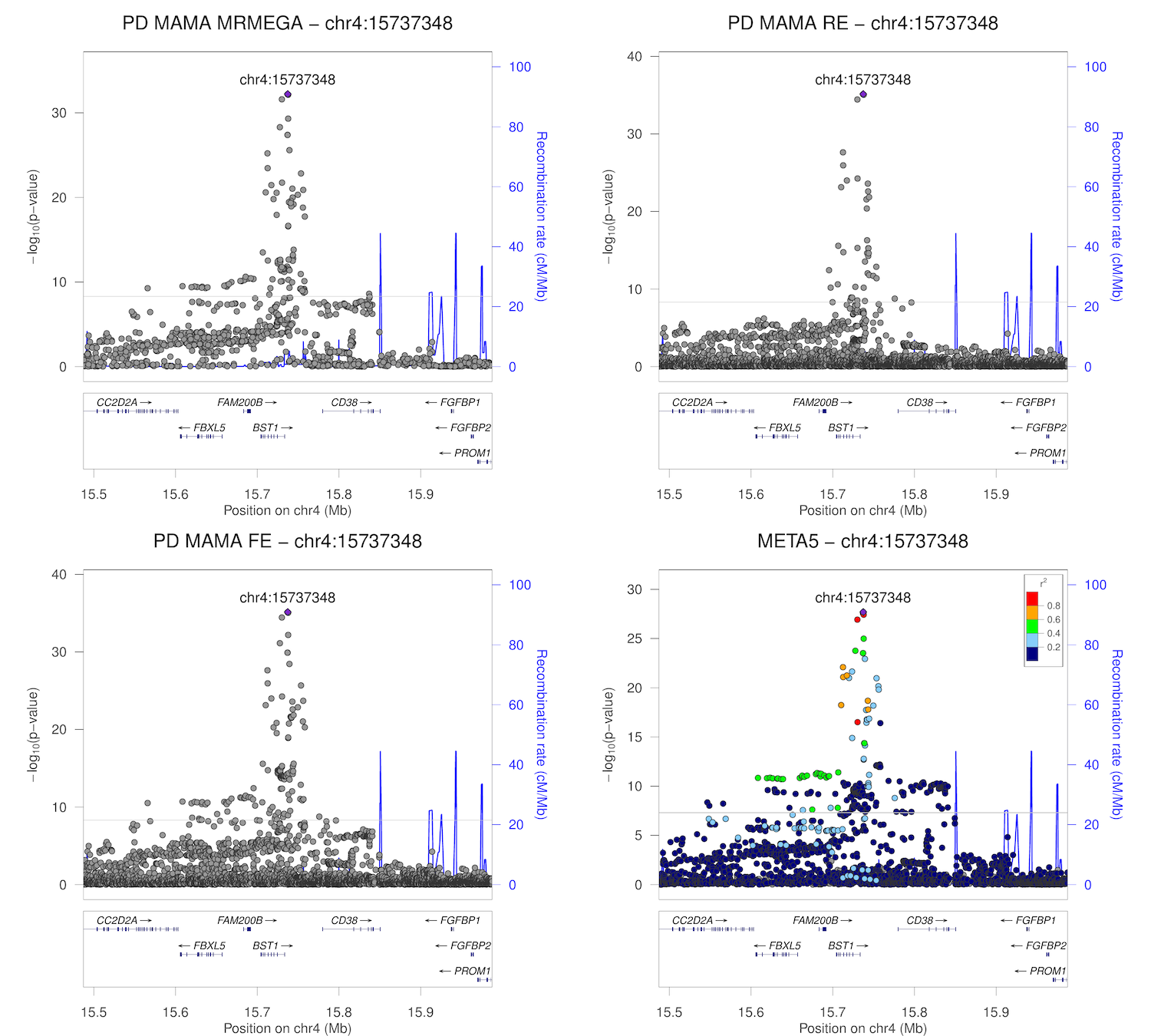

### chr4_17718811-18218811.png

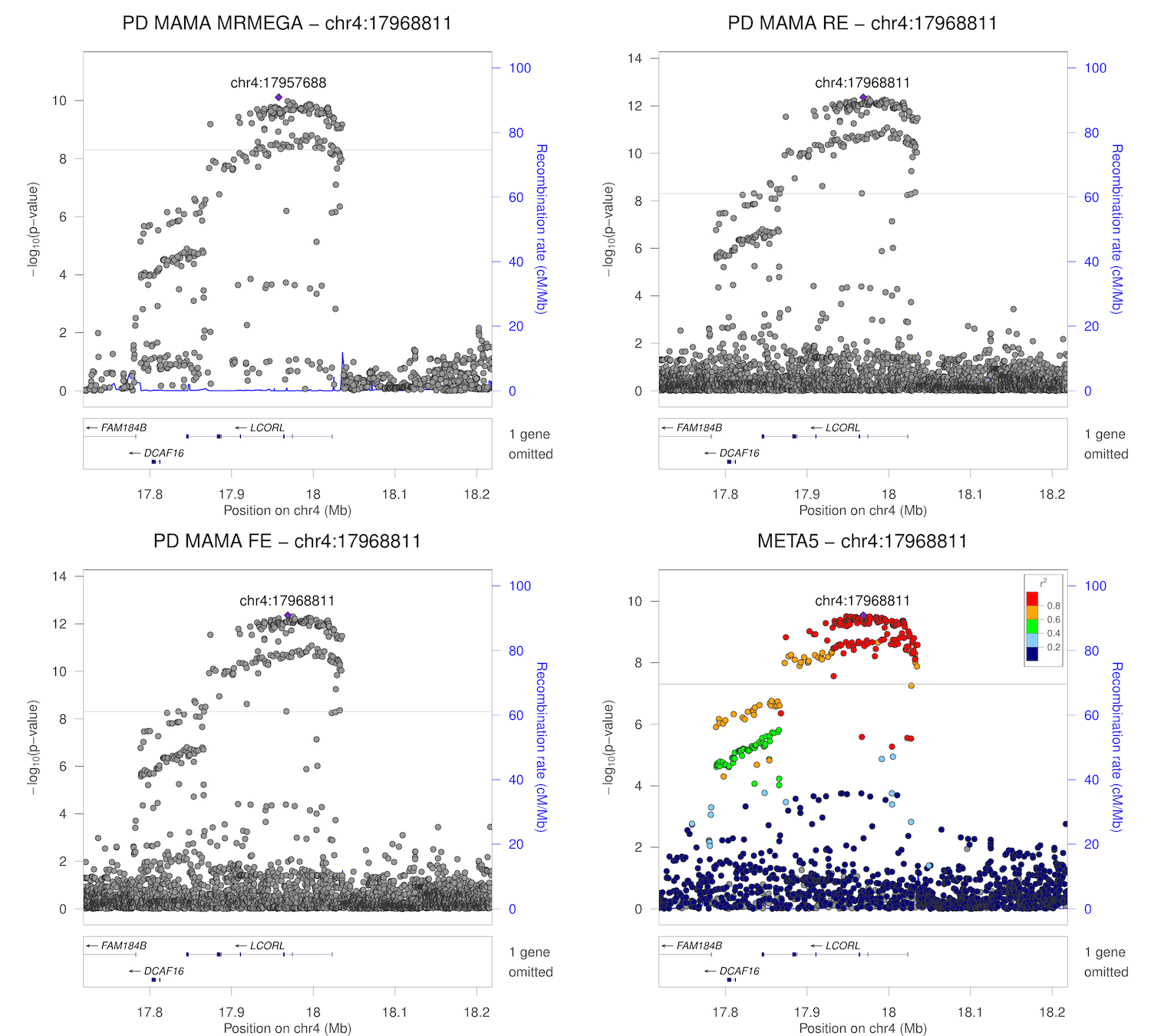

### chr4_76897969-77397969.png

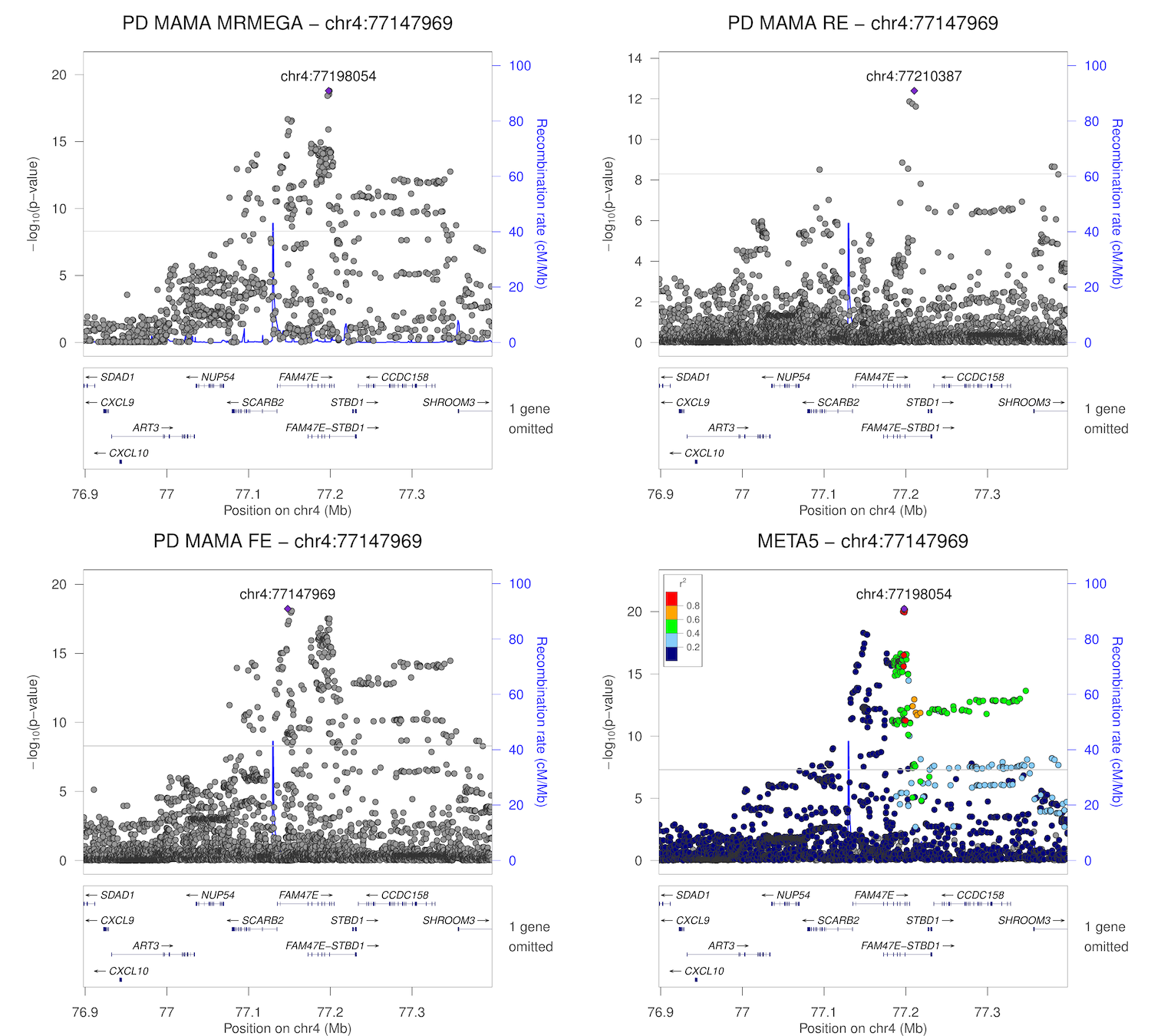

### chr4_76948054-77448054.png

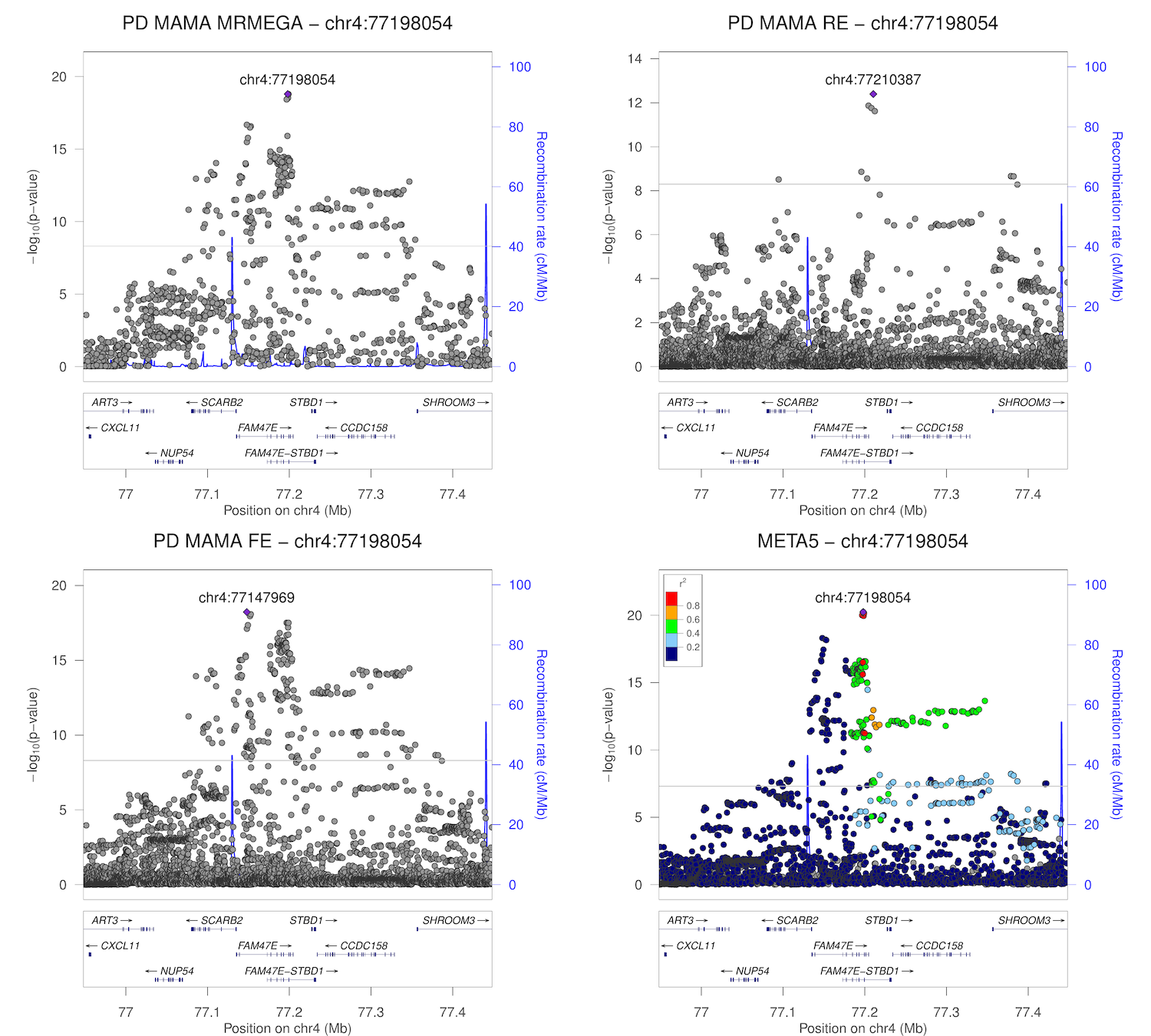

### chr4_90376111-90876111.png

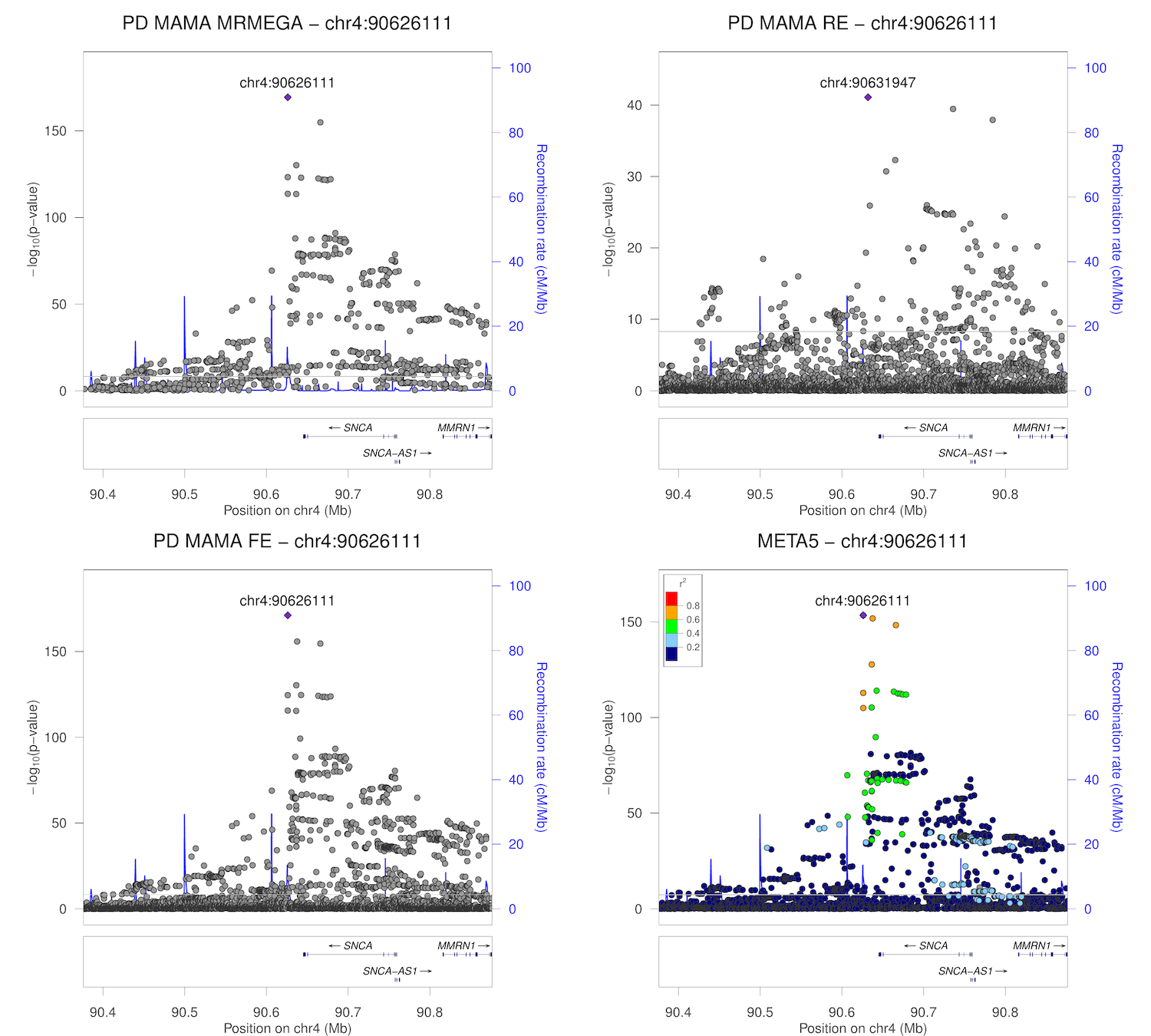

### chr4_90386630-90886630.png

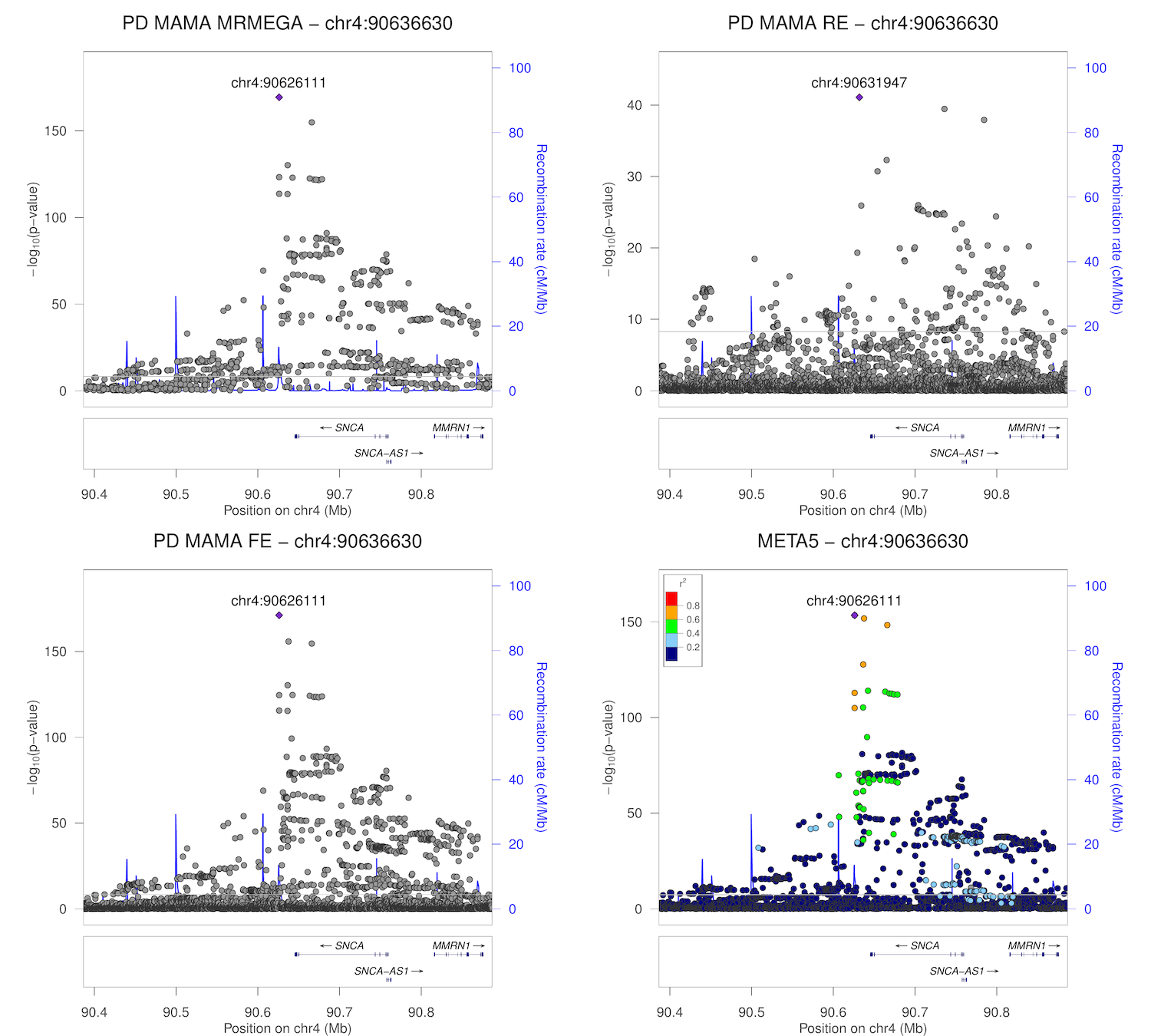

### chr4_114119065-114619065.png

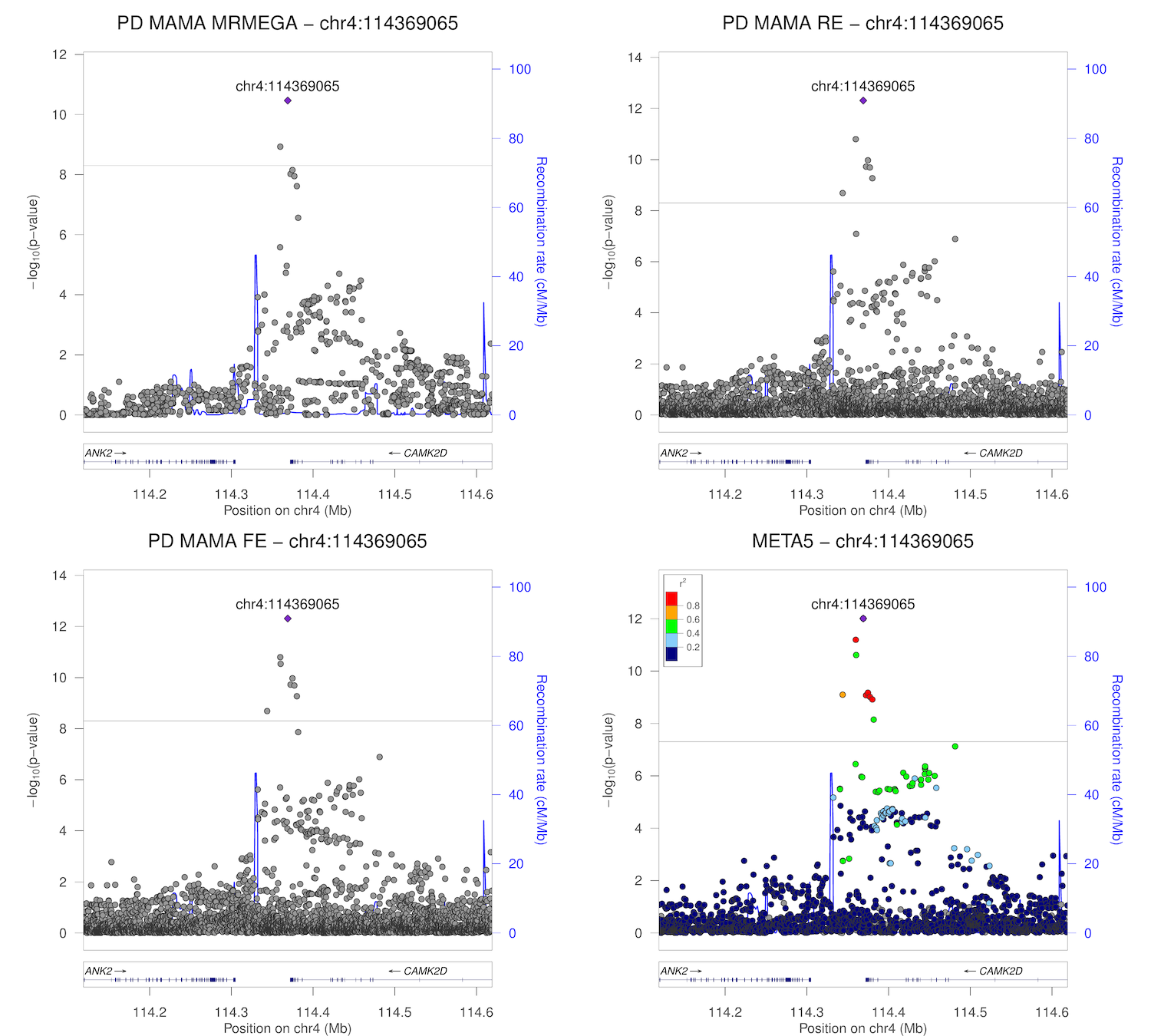

### chr4_170333157-170833157.png

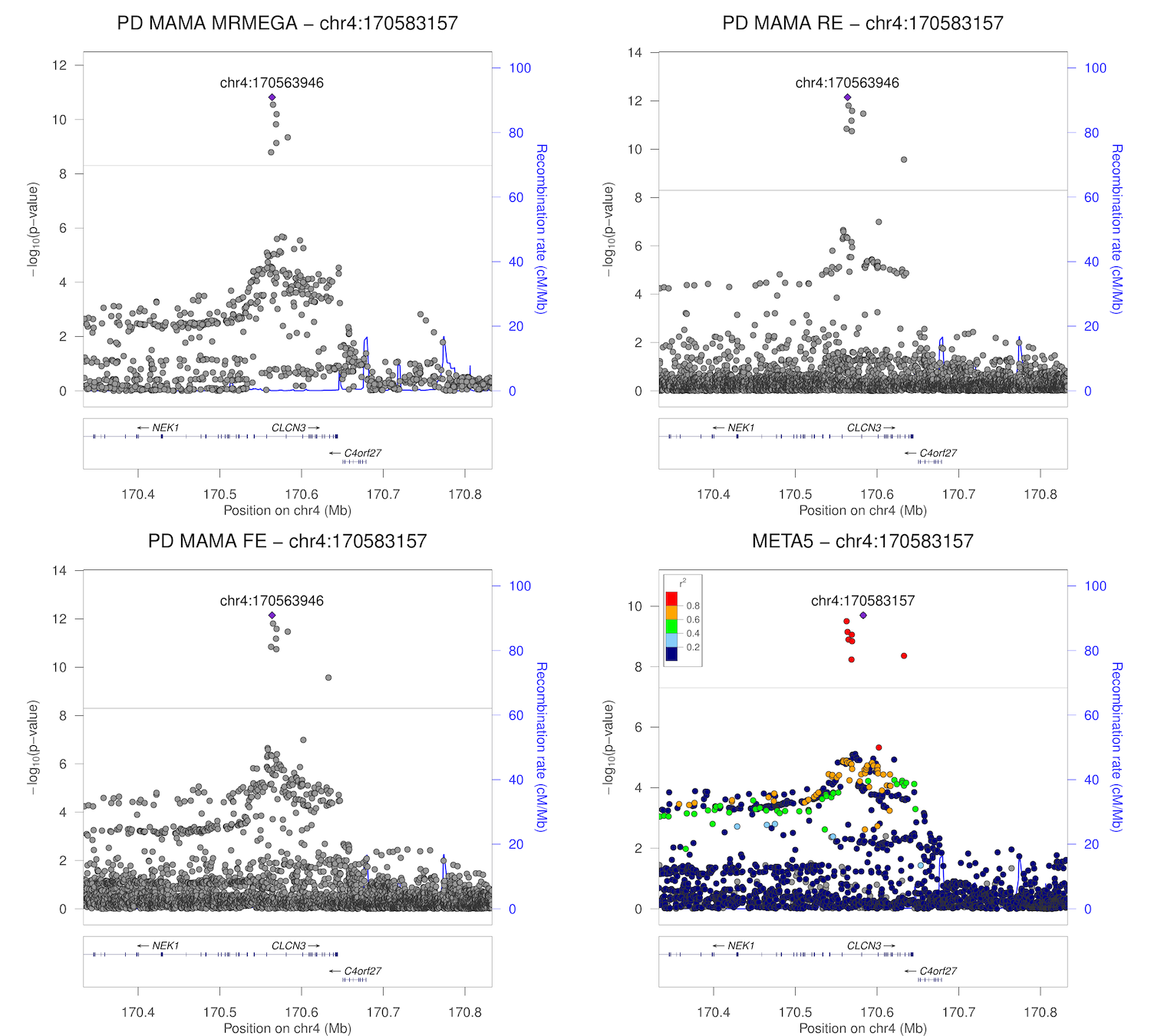

### chr5_59887959-60387959.png

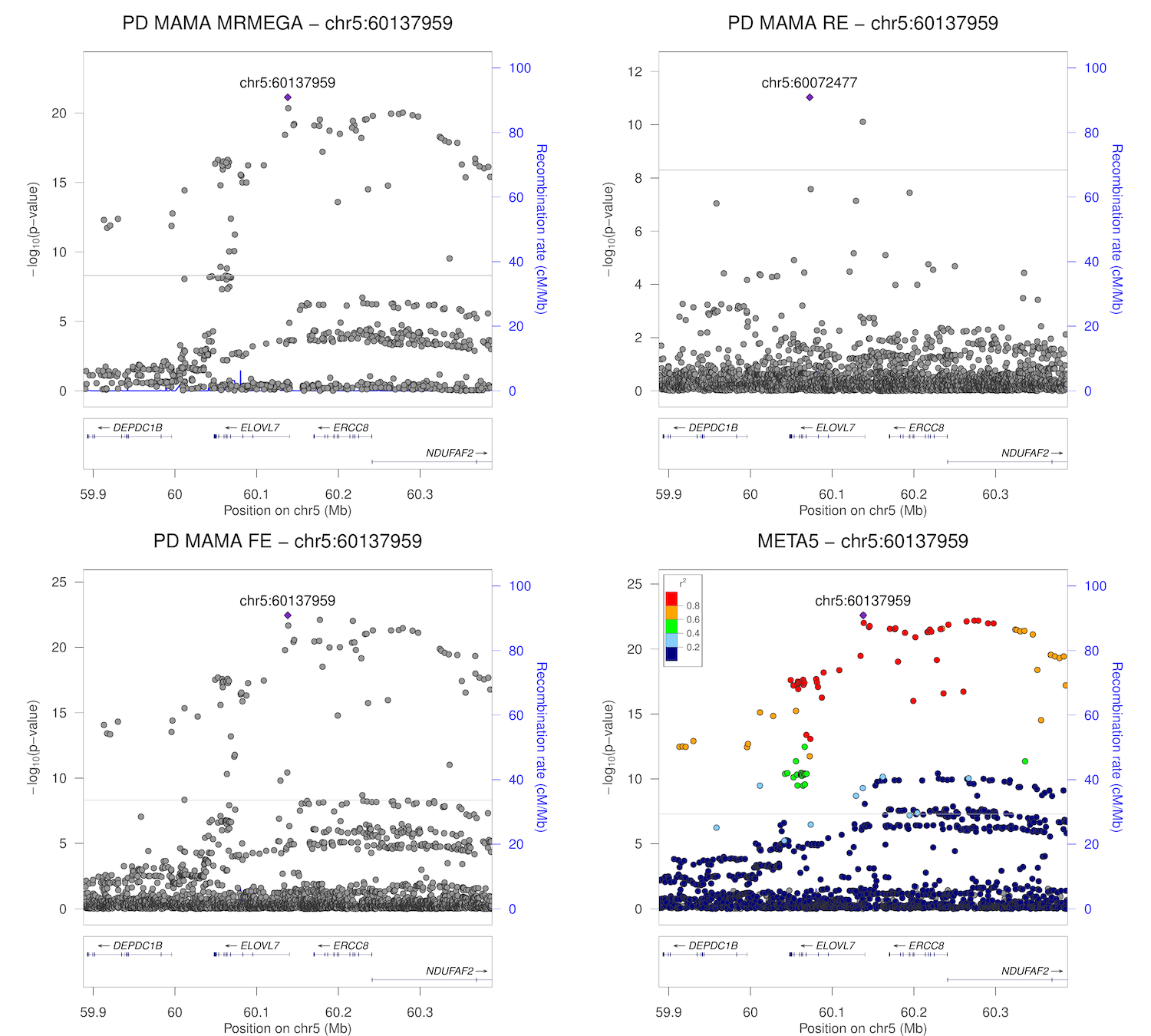

### chr5_102115794-102615794.png

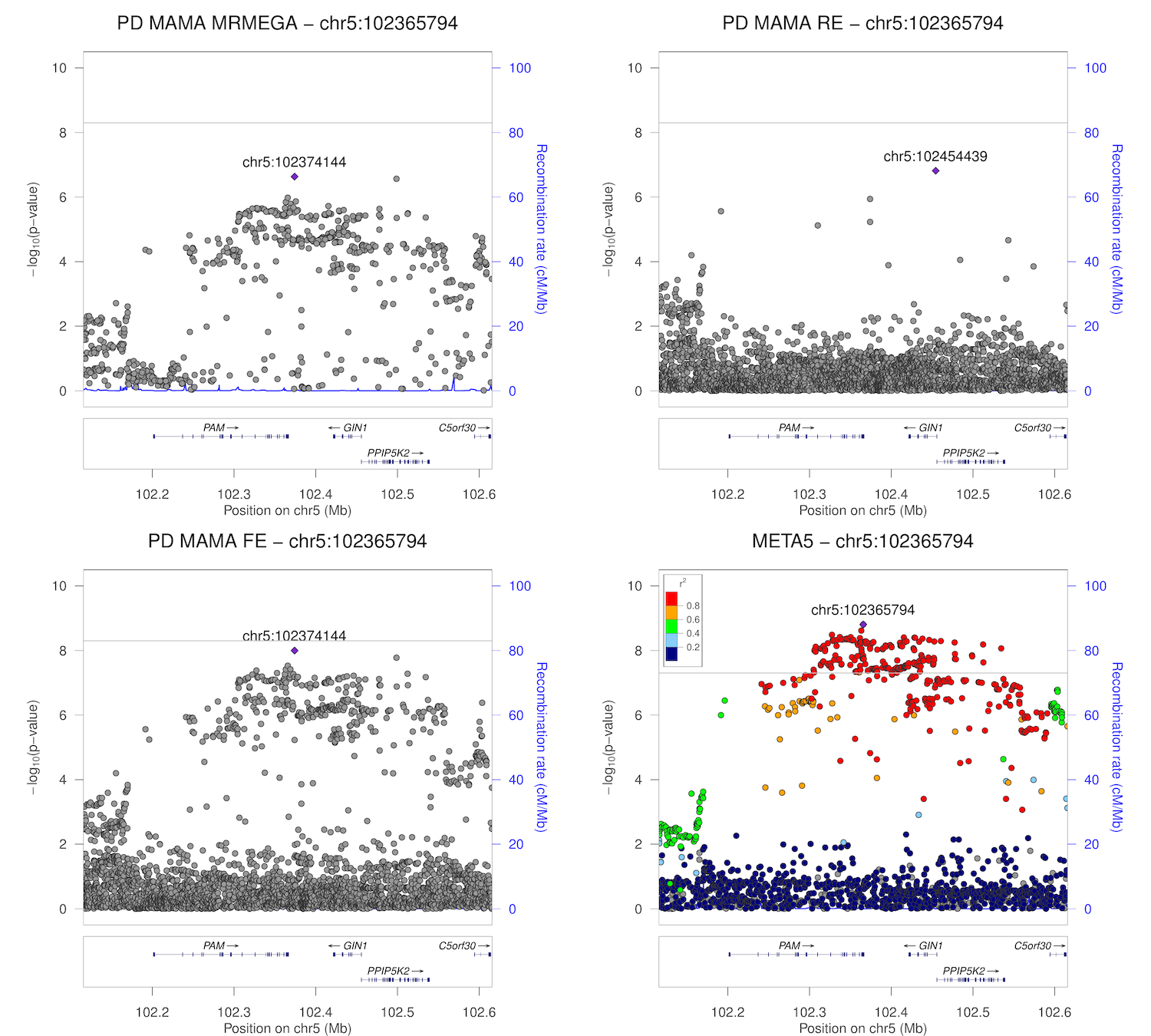

### chr5_133949105-134449105.png

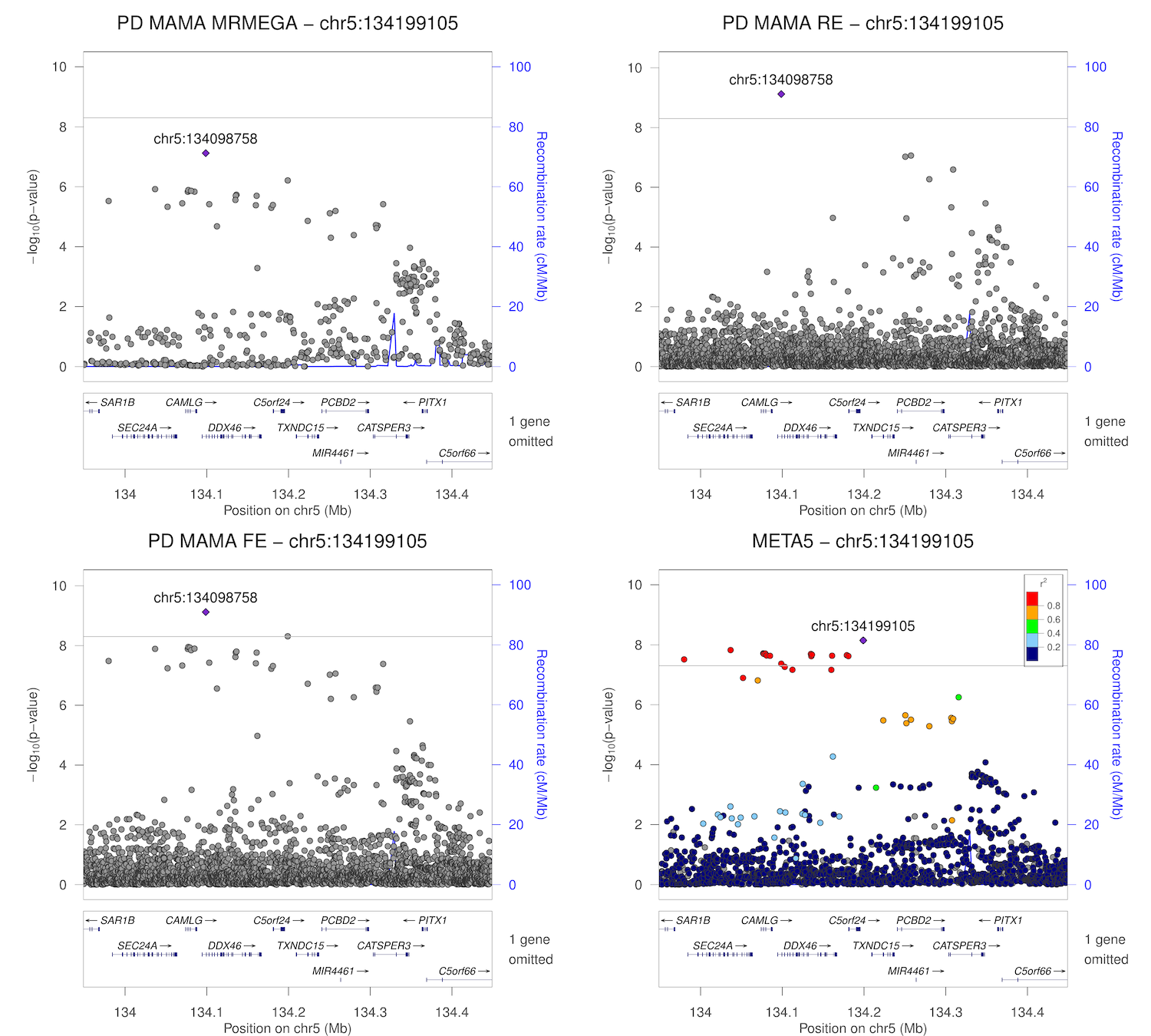

### chr6_27488801-27988801.png

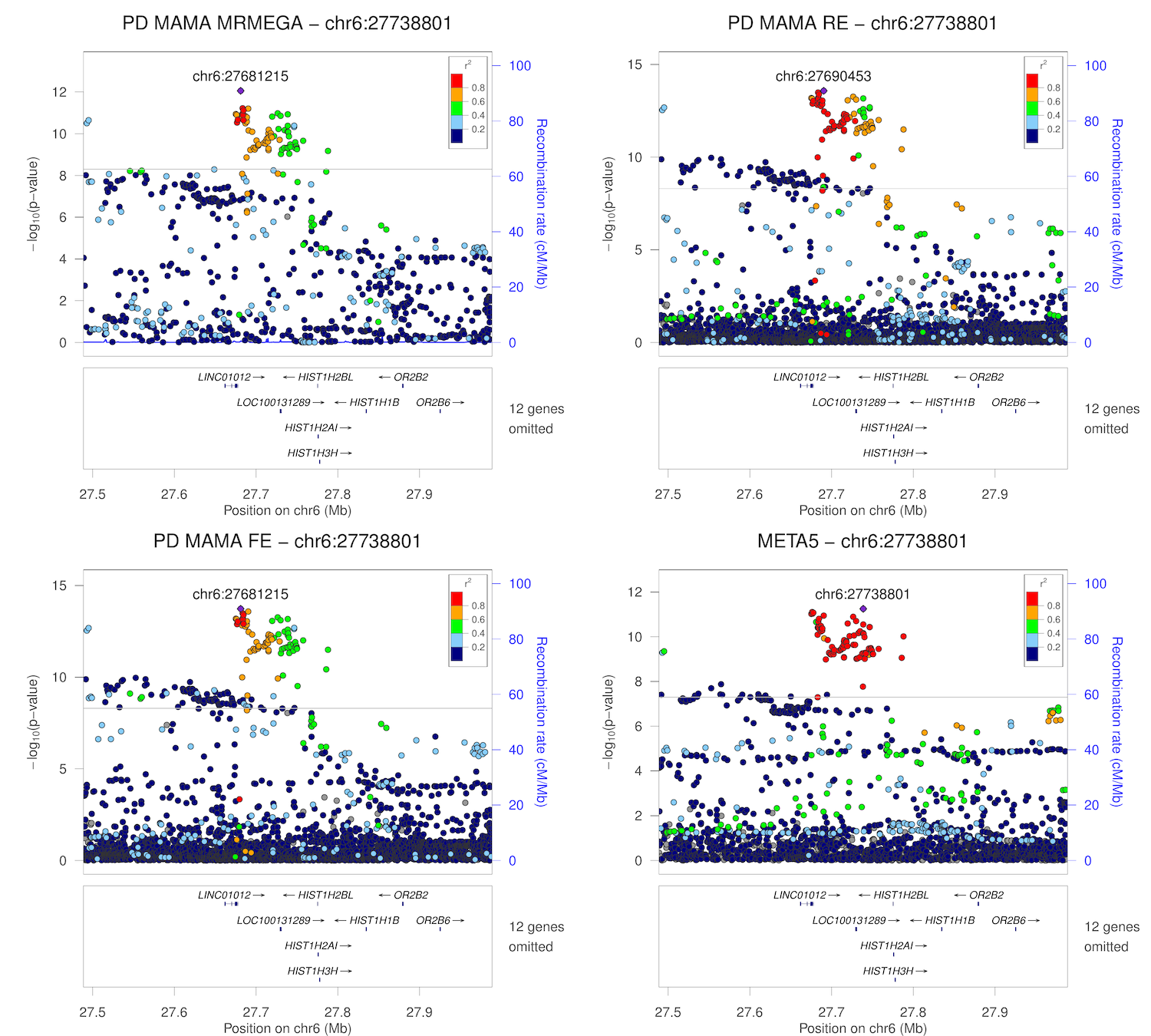

### chr6_29858683-30358683.png

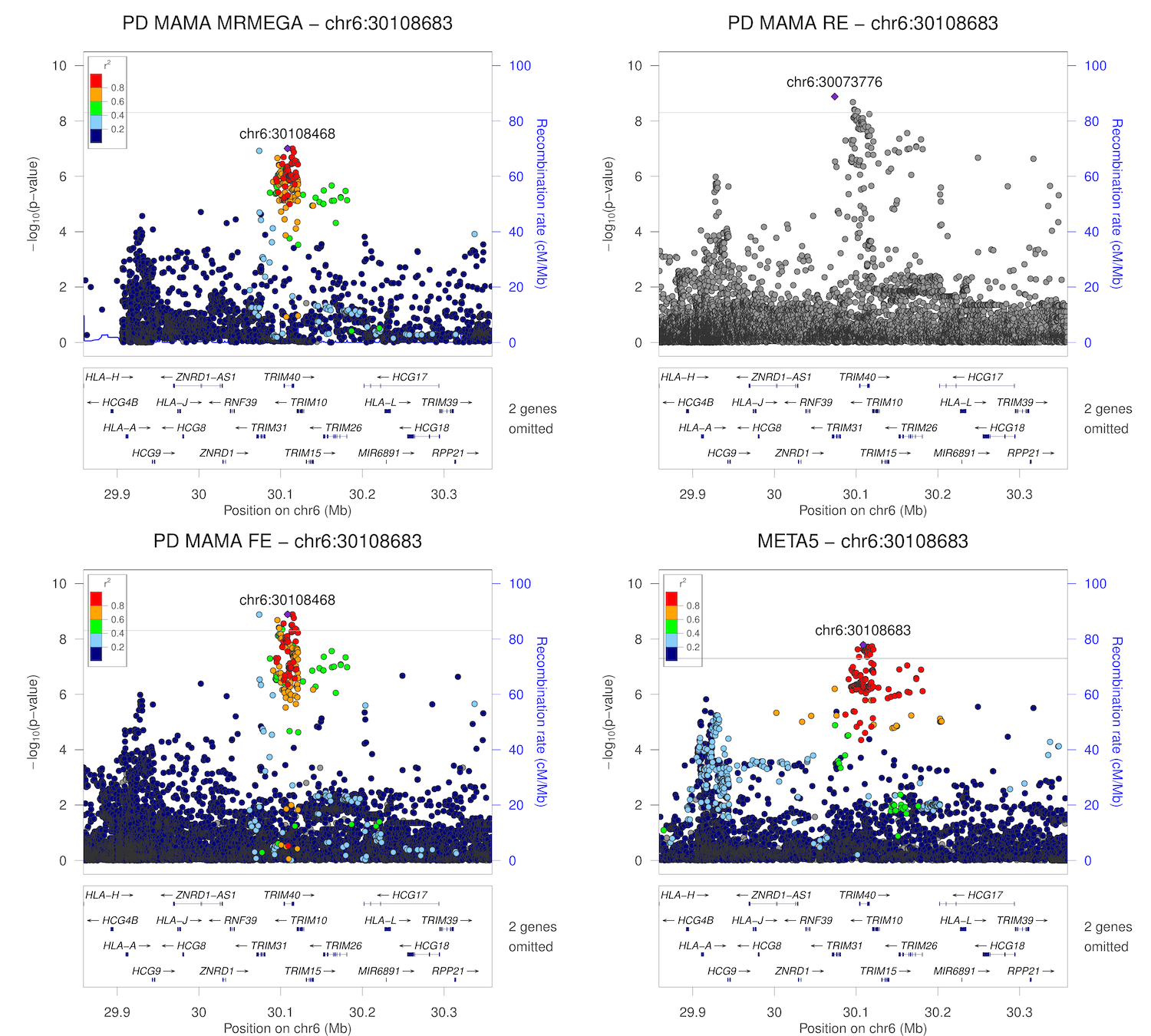

### chr6_32328772-32828772.png

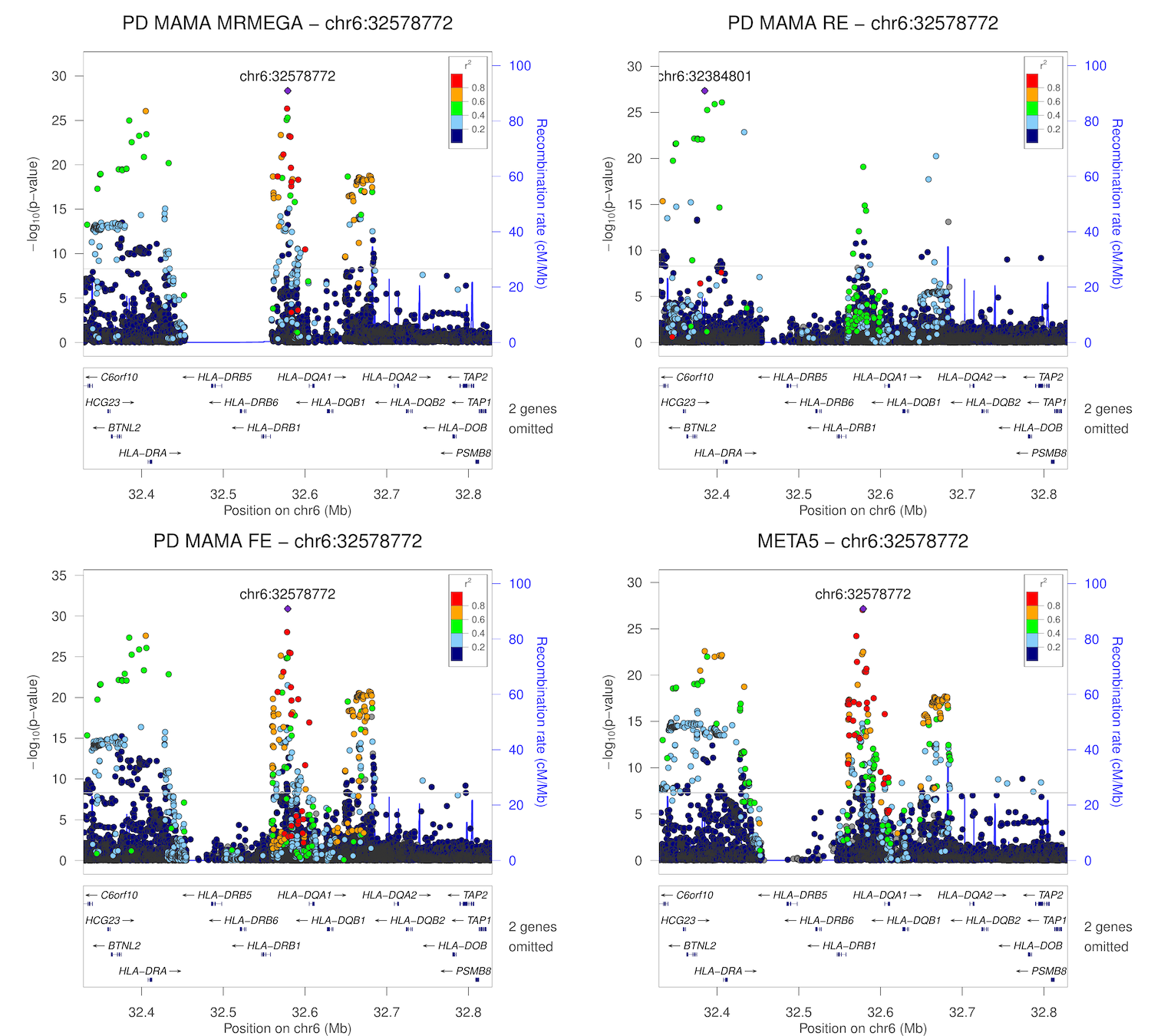

### chr6_72237762-72737762.png

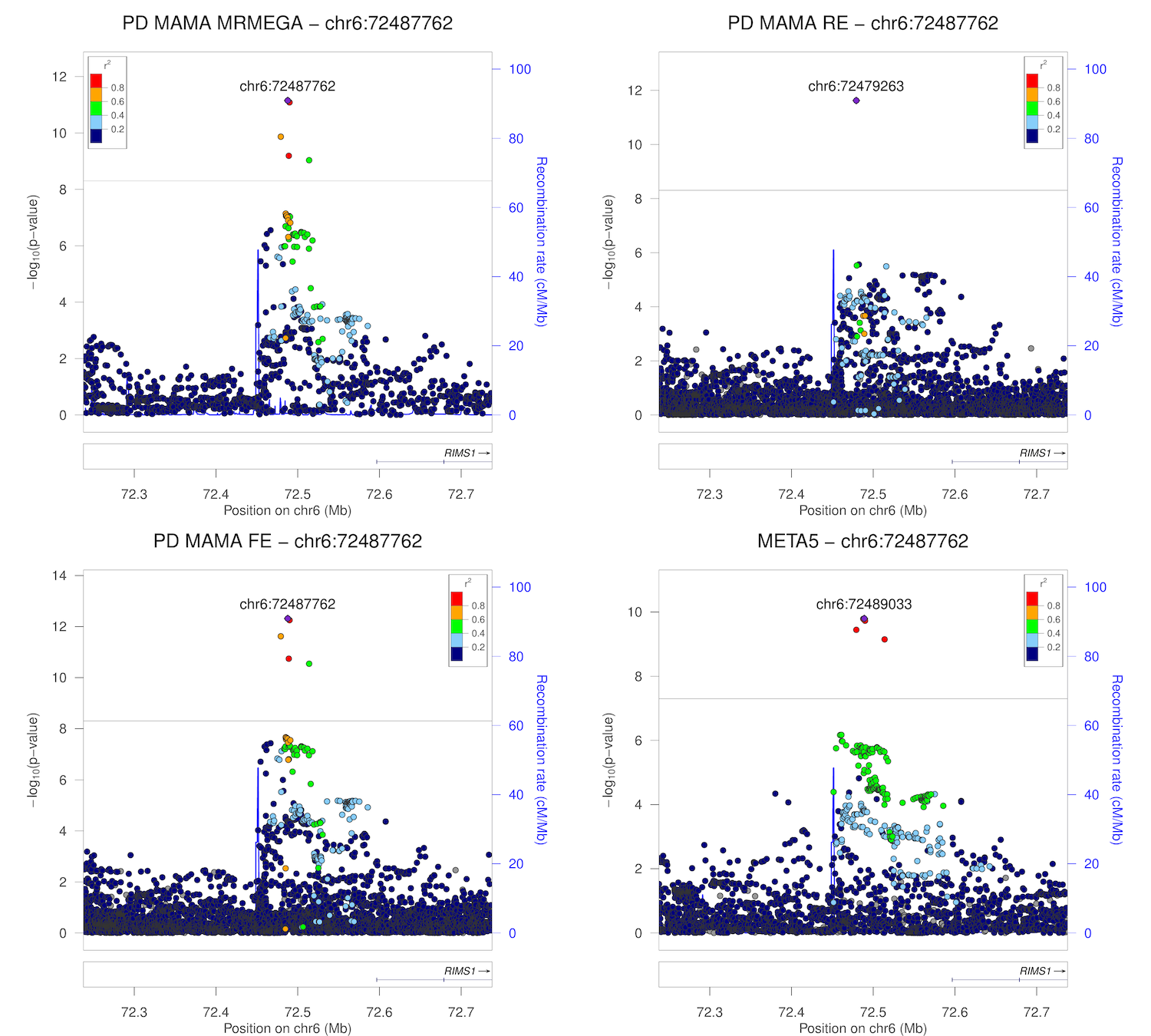

### chr6_111993291-112493291.png

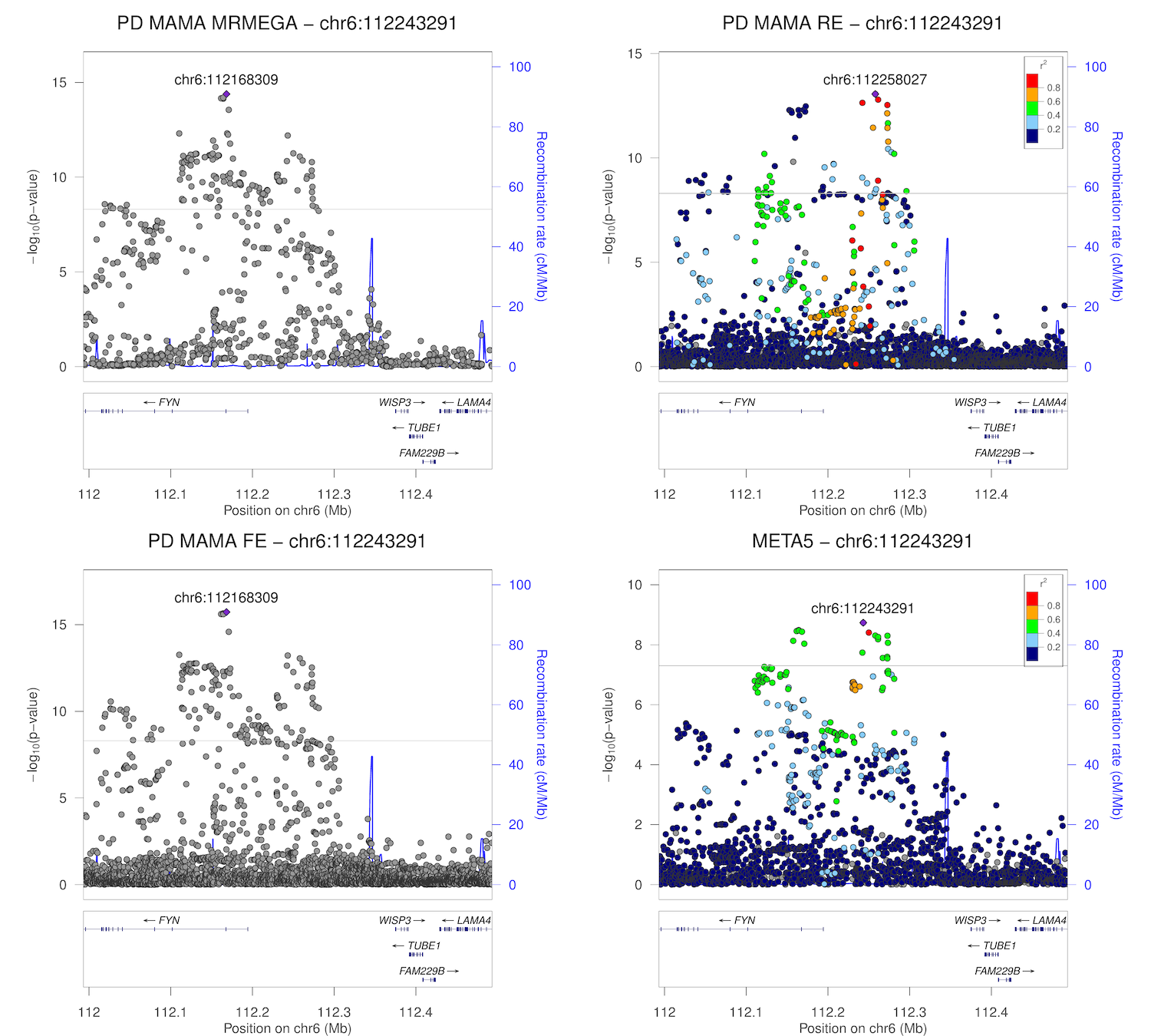

### chr6_132960361-133460361.png

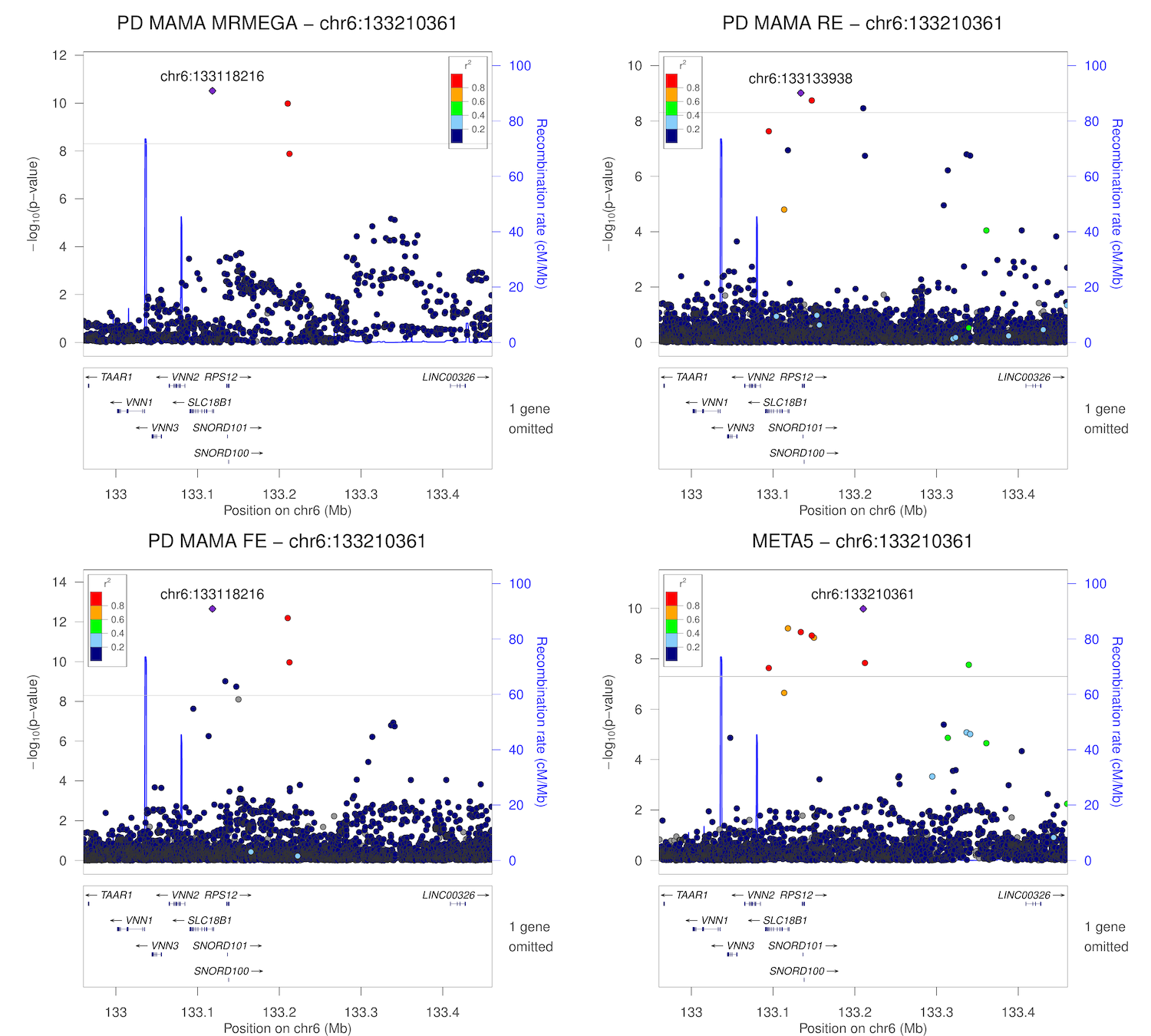

### chr7_23050049-23550049.png

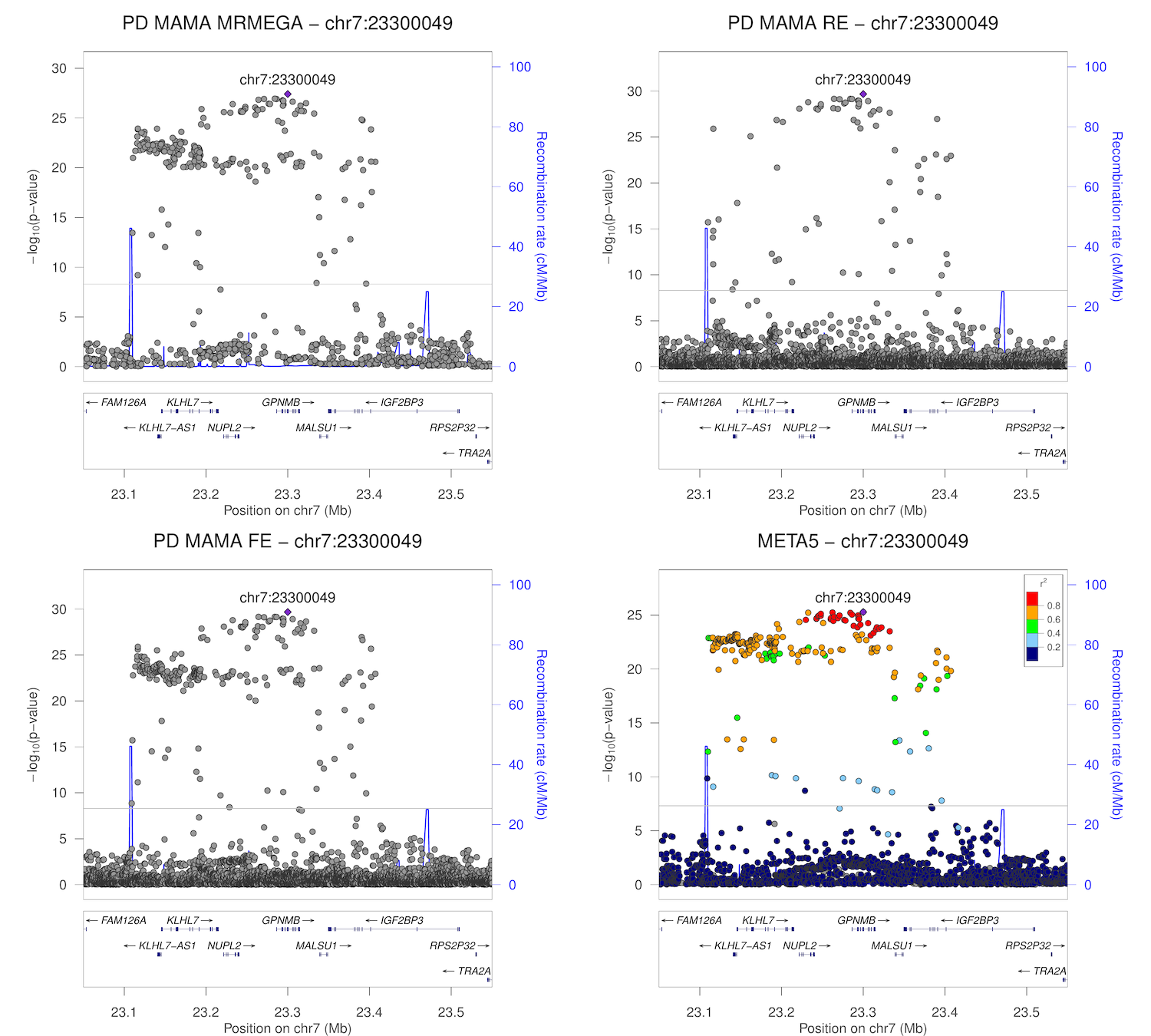

### chr7_65759851-66259851.png

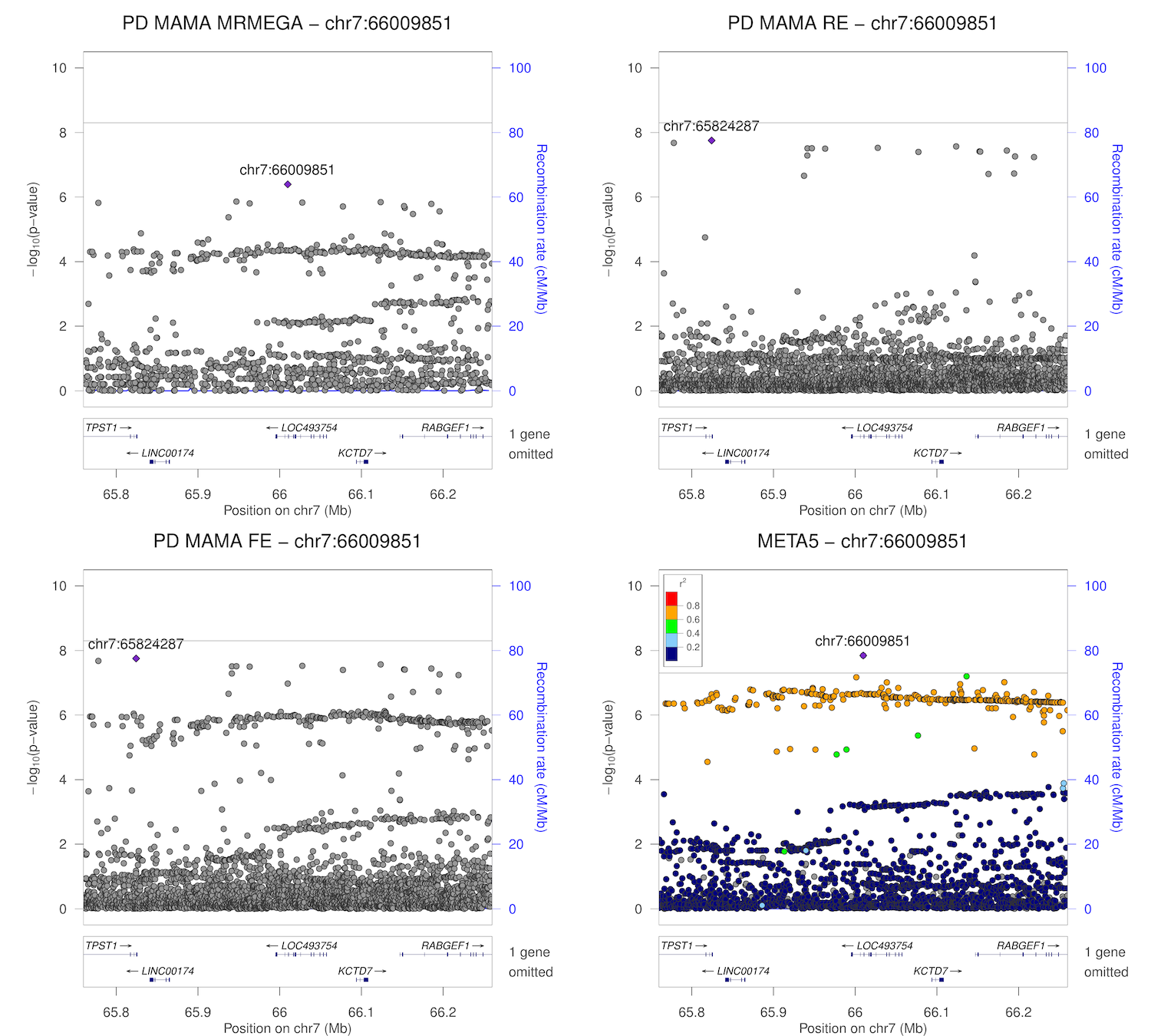

### chr8_11462443-11962443.png

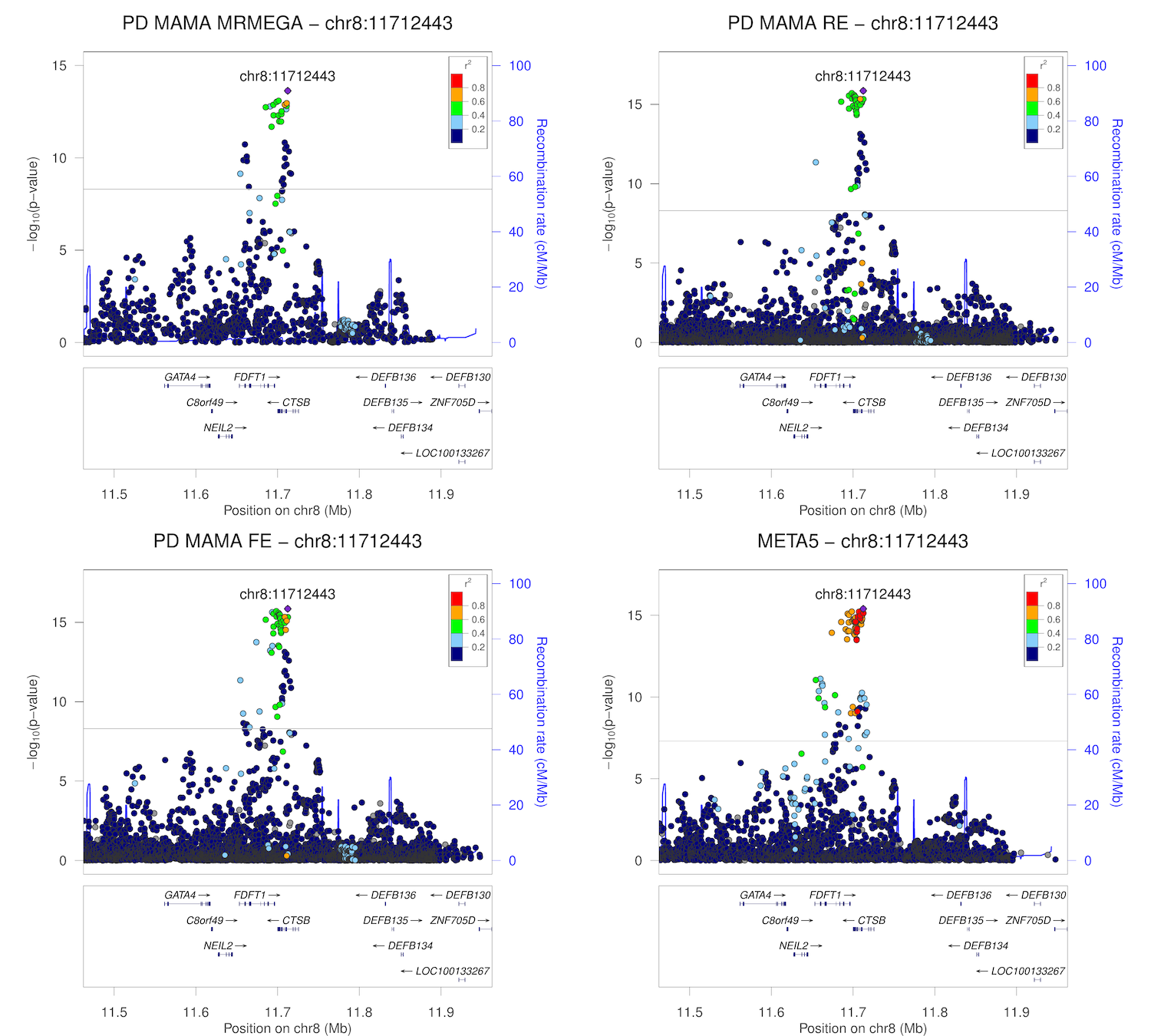

### chr8_16447593-16947593.png

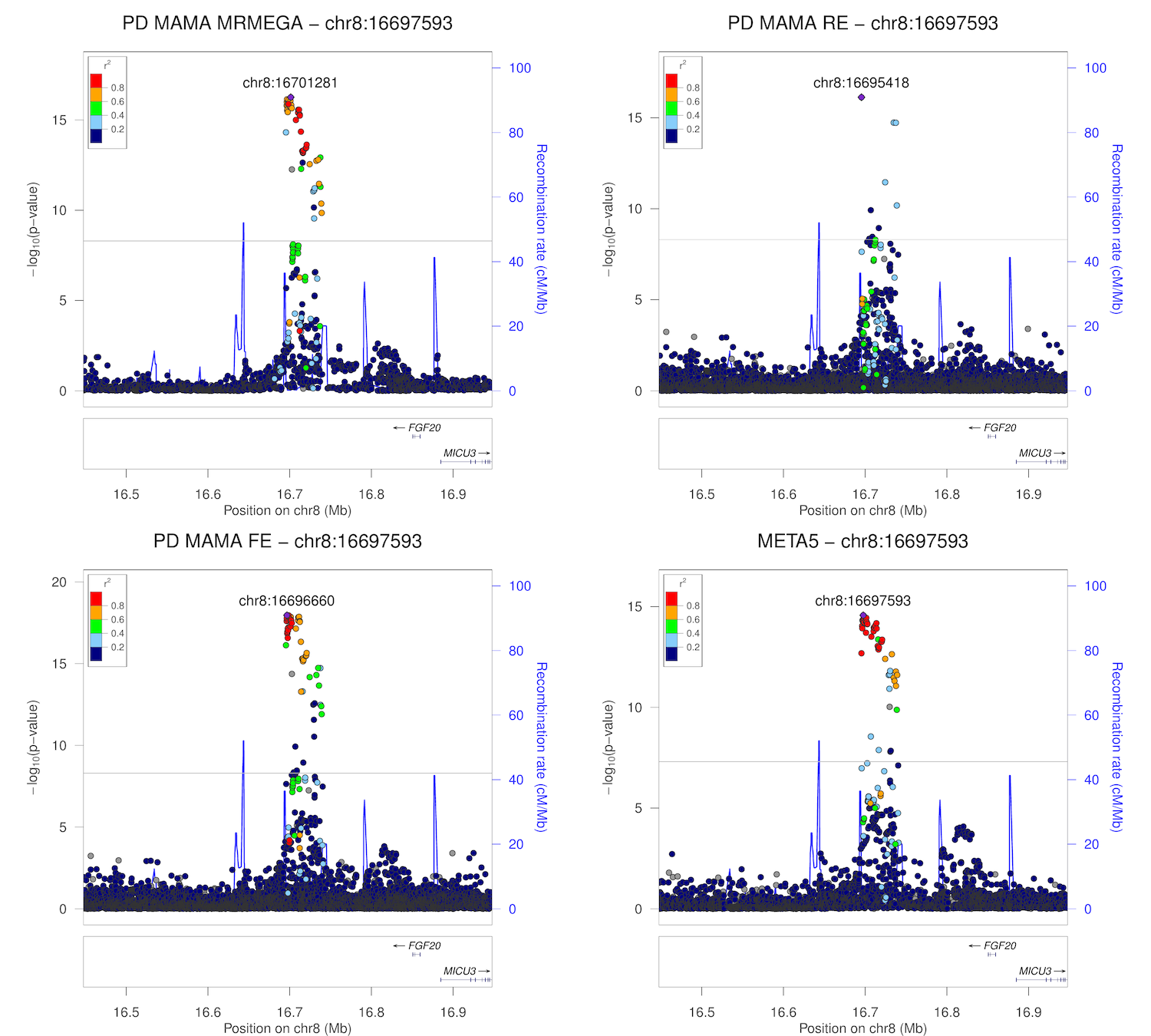

### chr8_22275980-22775980.png

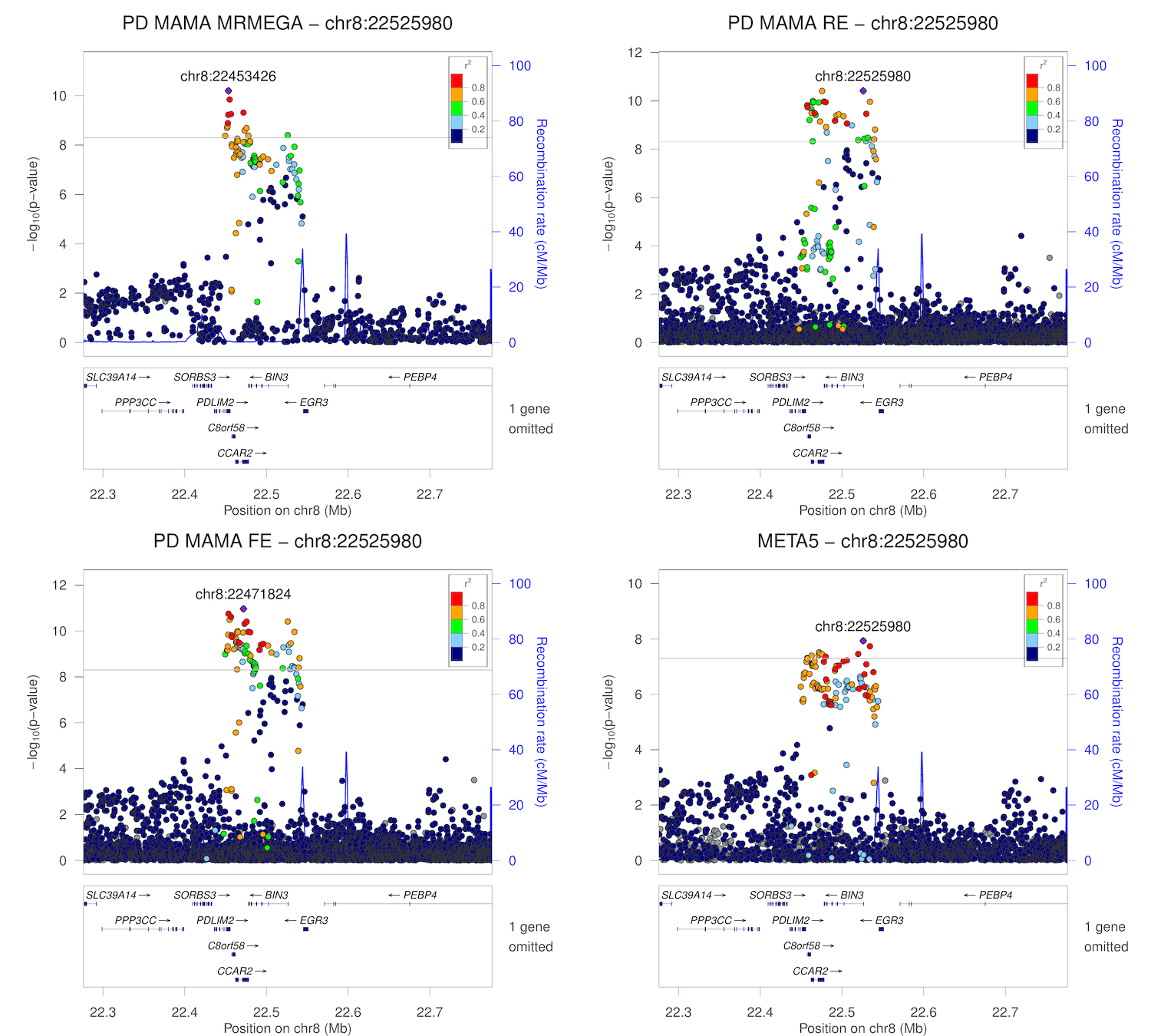

### chr8_110394774-110894774.png

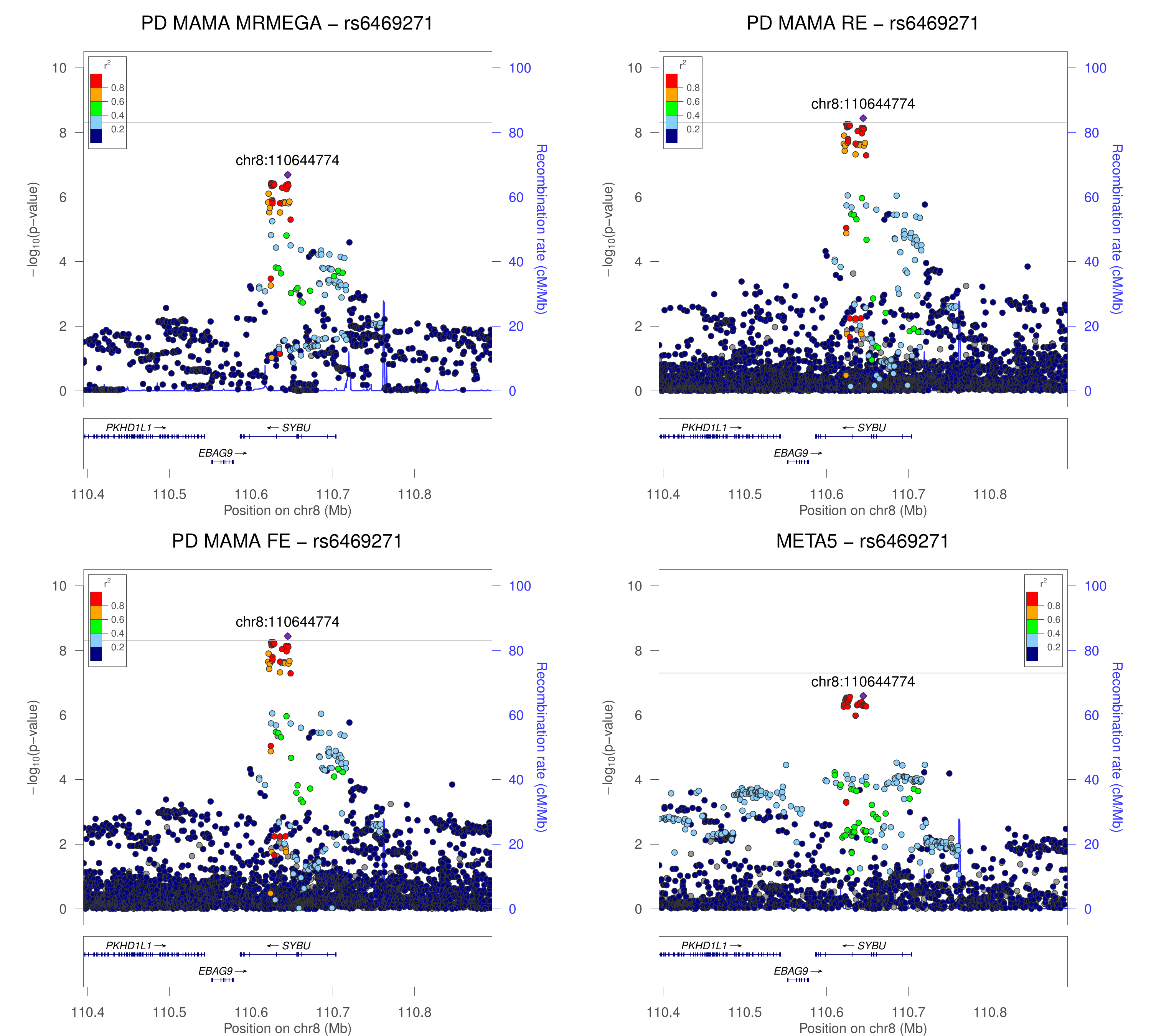

### chr8_130651909-131151909.png

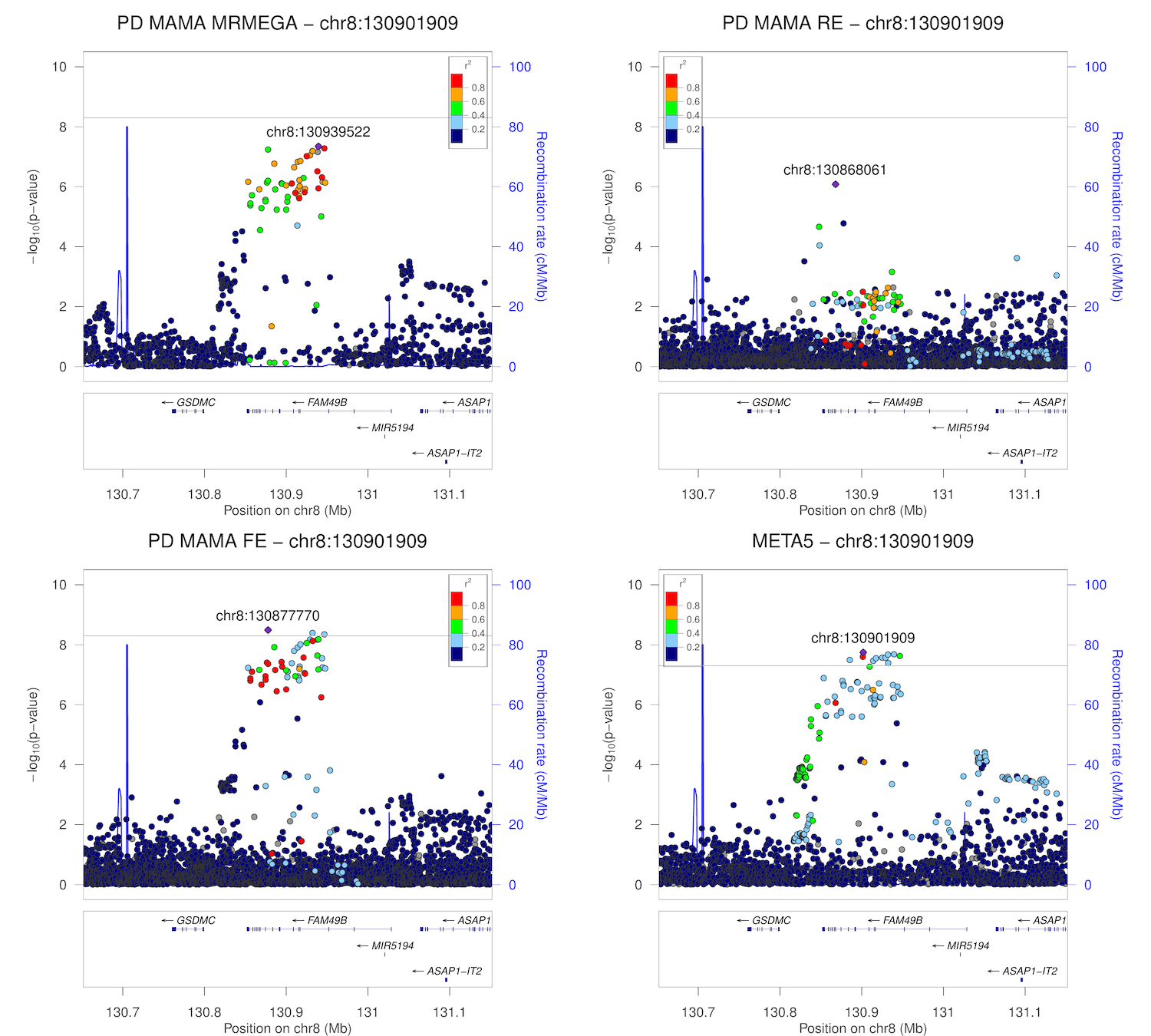

### chr9_17329690-17829690.png

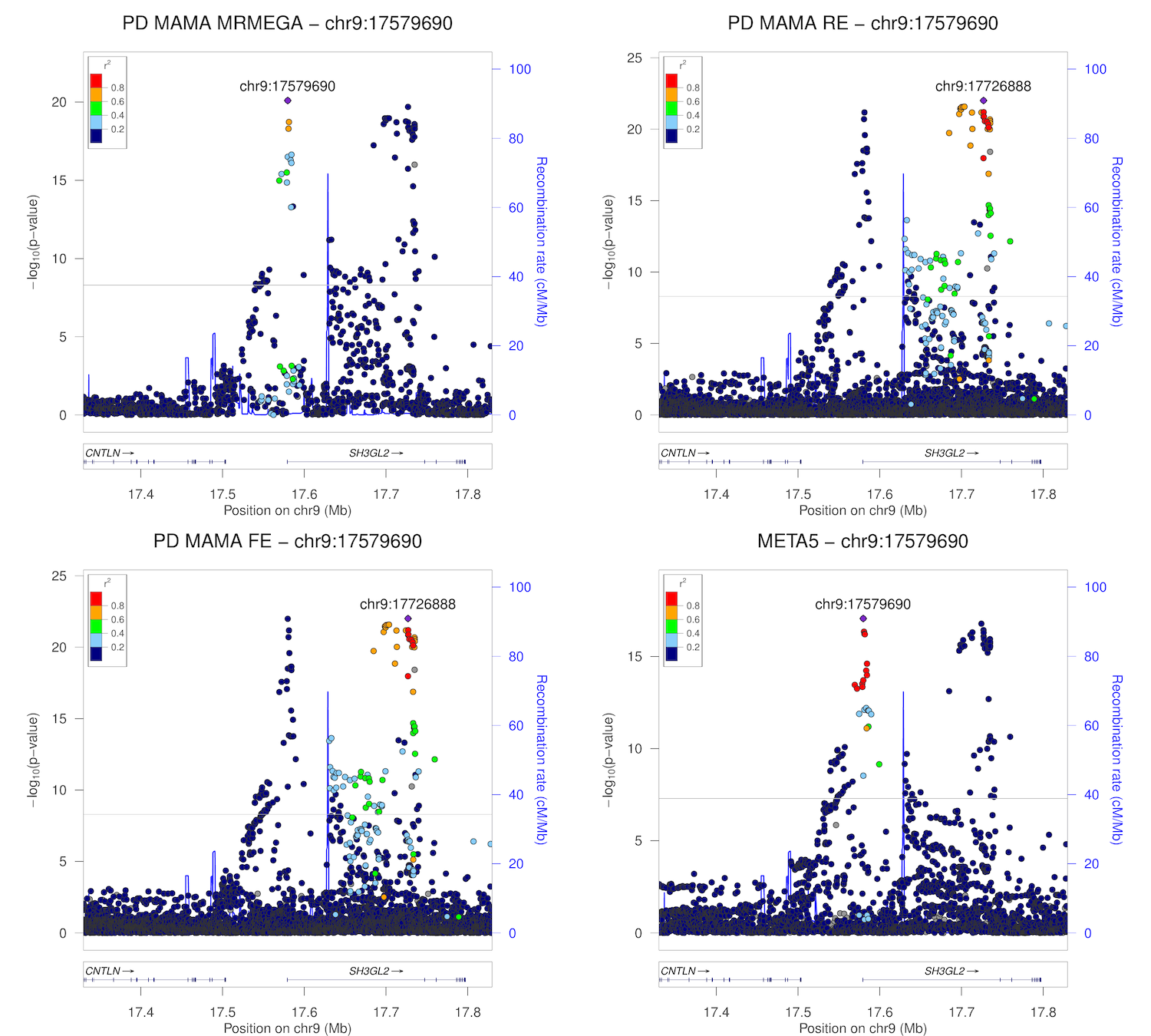

### chr9_17477065-17977065.png

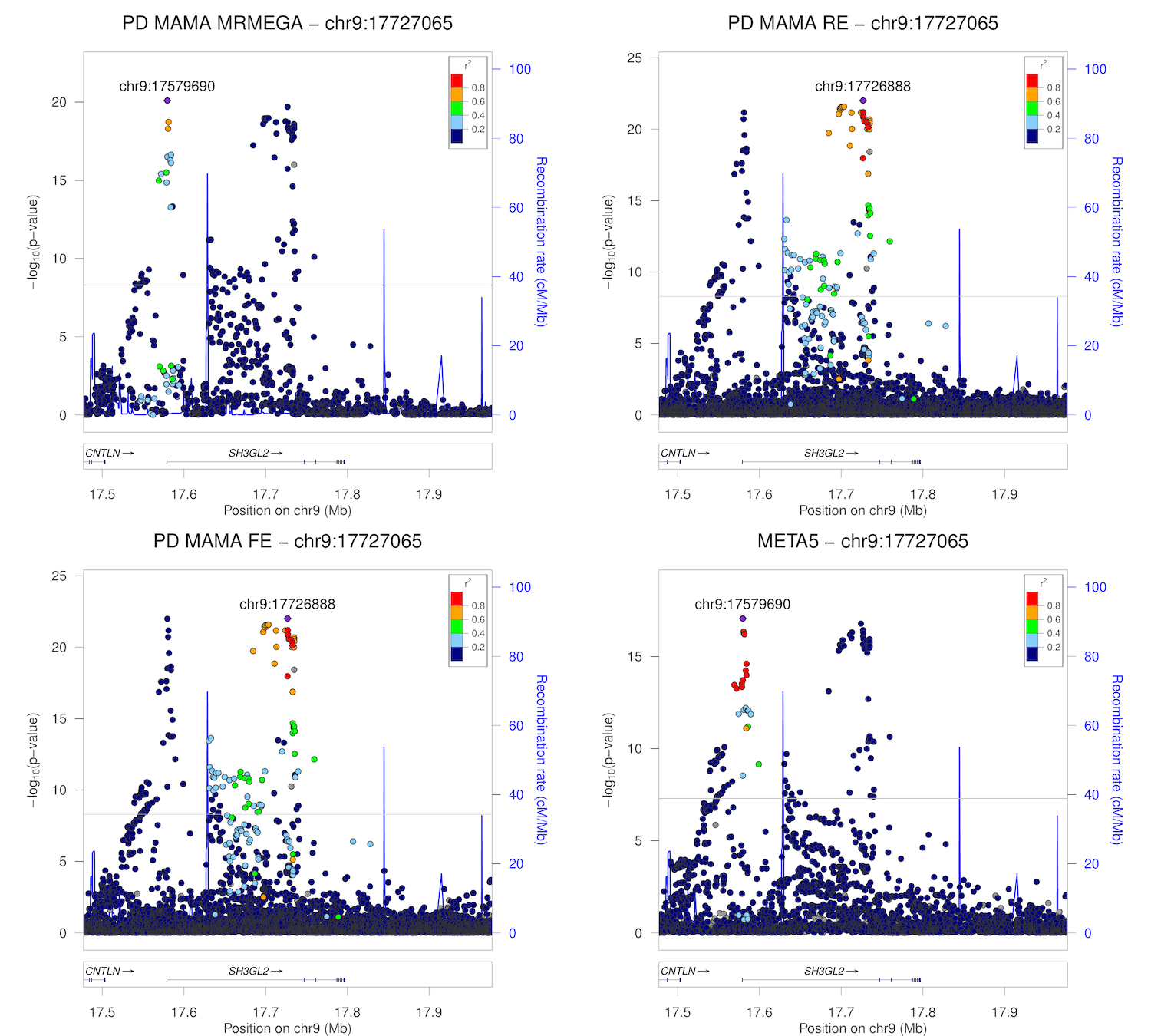

### chr9_33796391-34296391.png

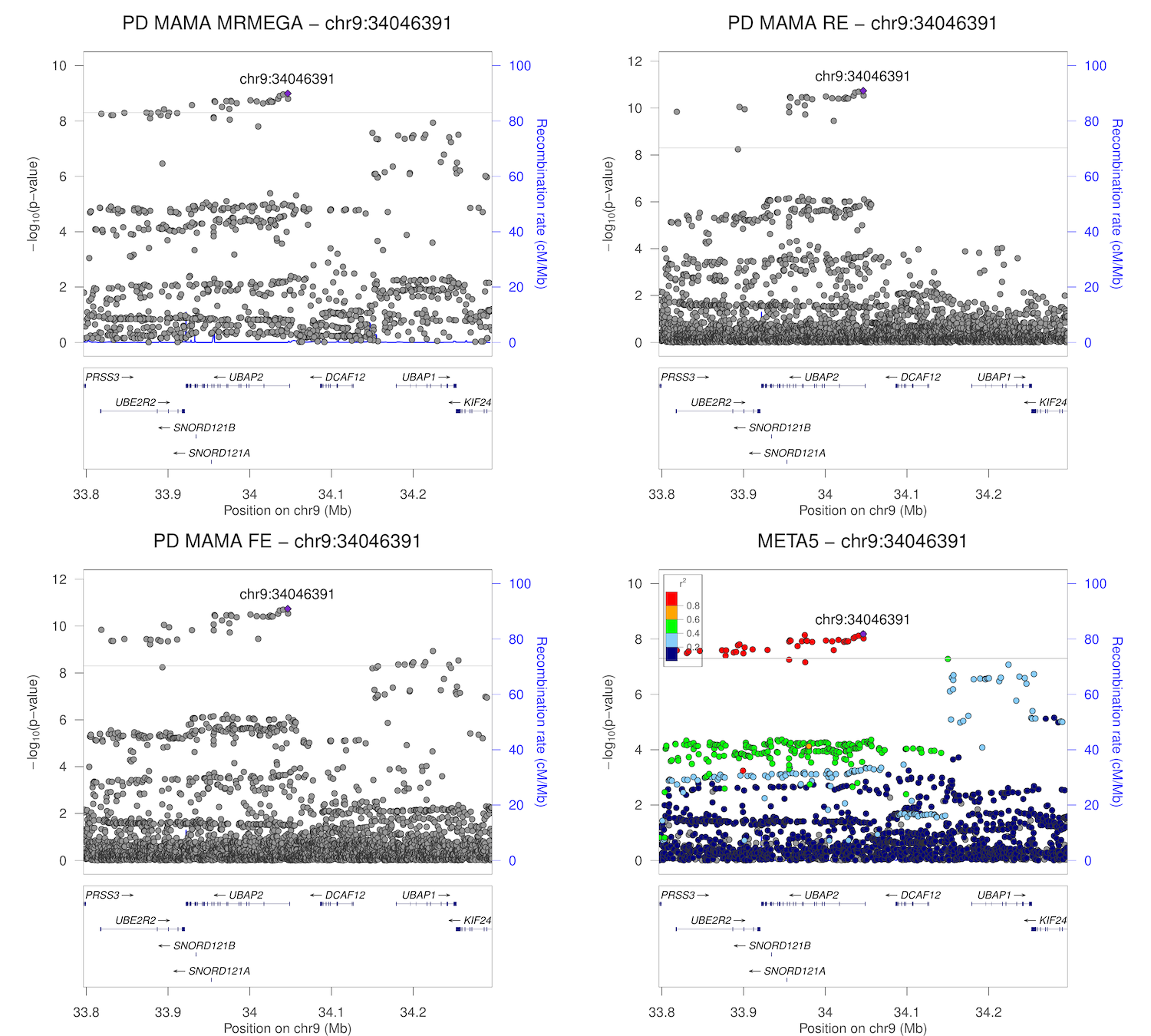

### chr10_15307406-15807406.png

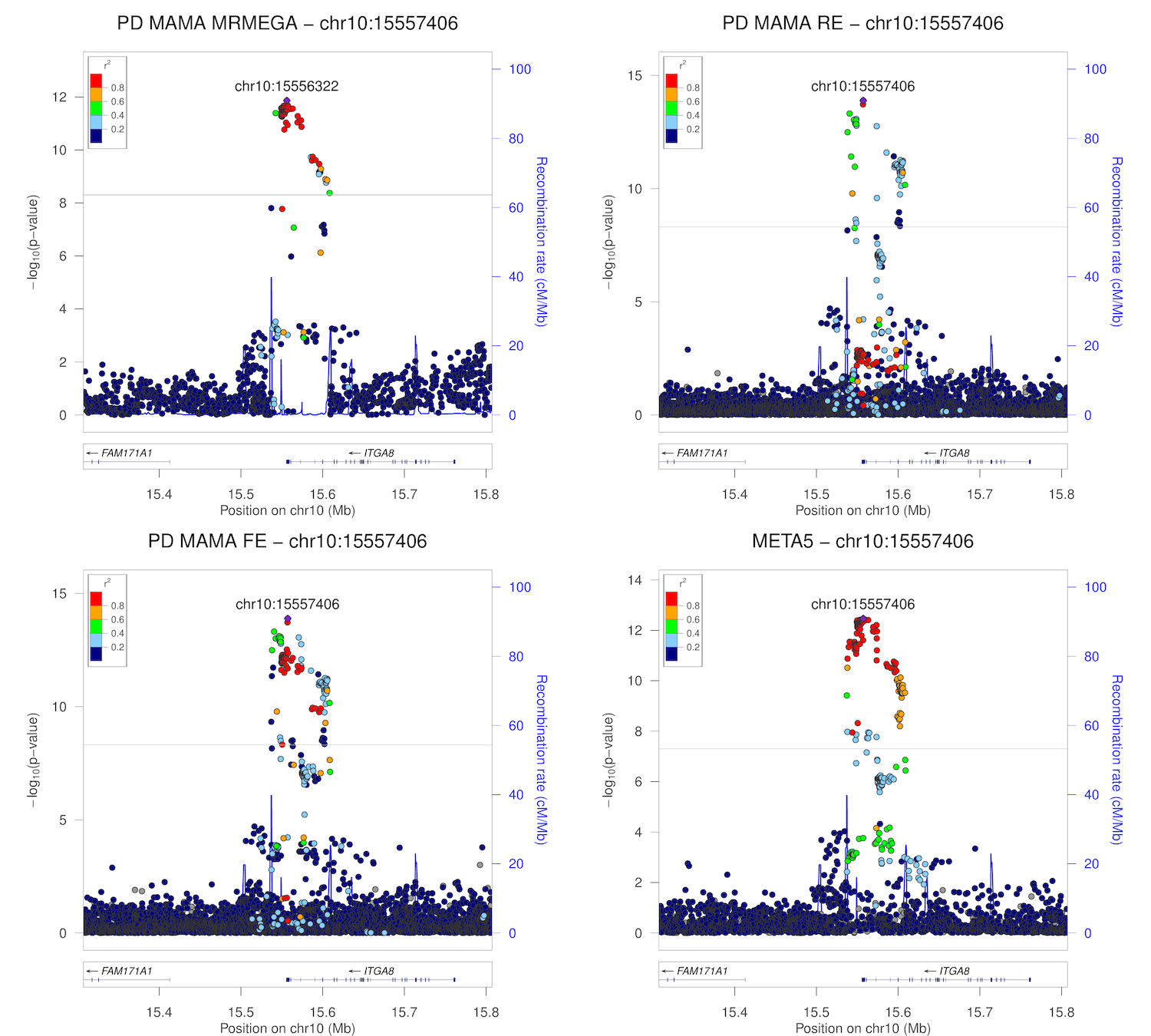

### chr10_103765279-104265279.png

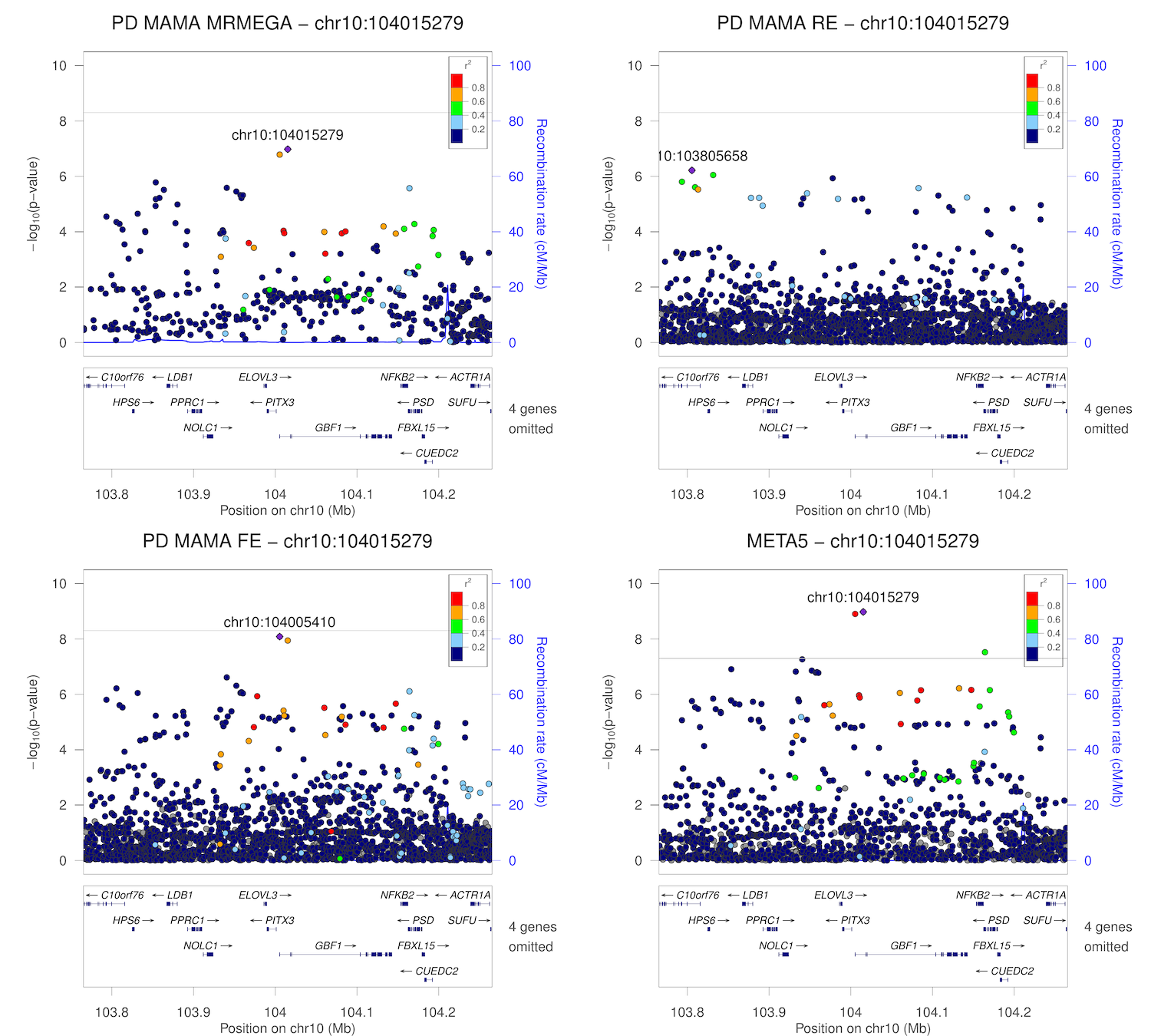

### chr10_121165685-121665685.png

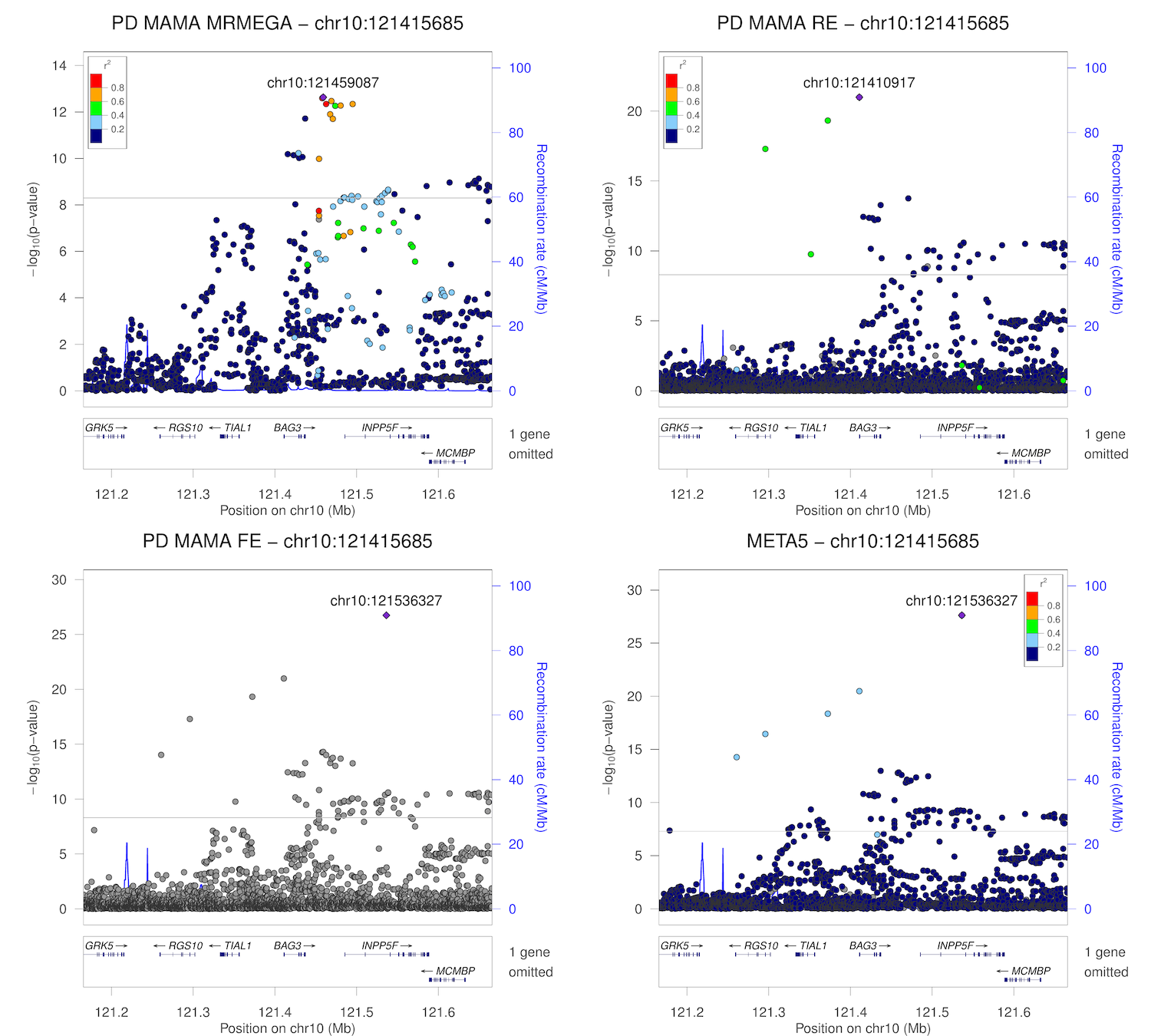

### chr10_121286327-121786327.png

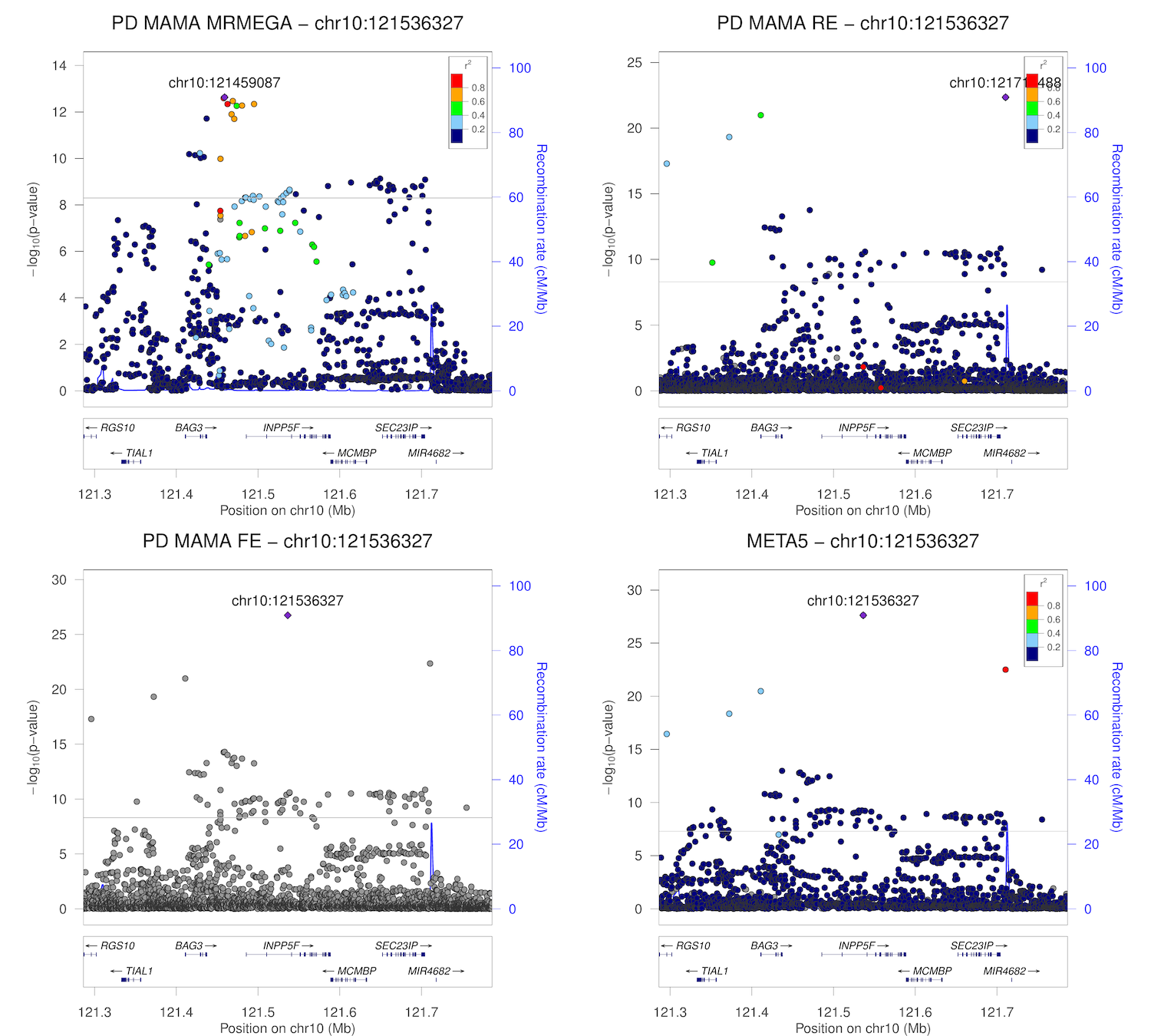

### chr11_10308777-10808777.png

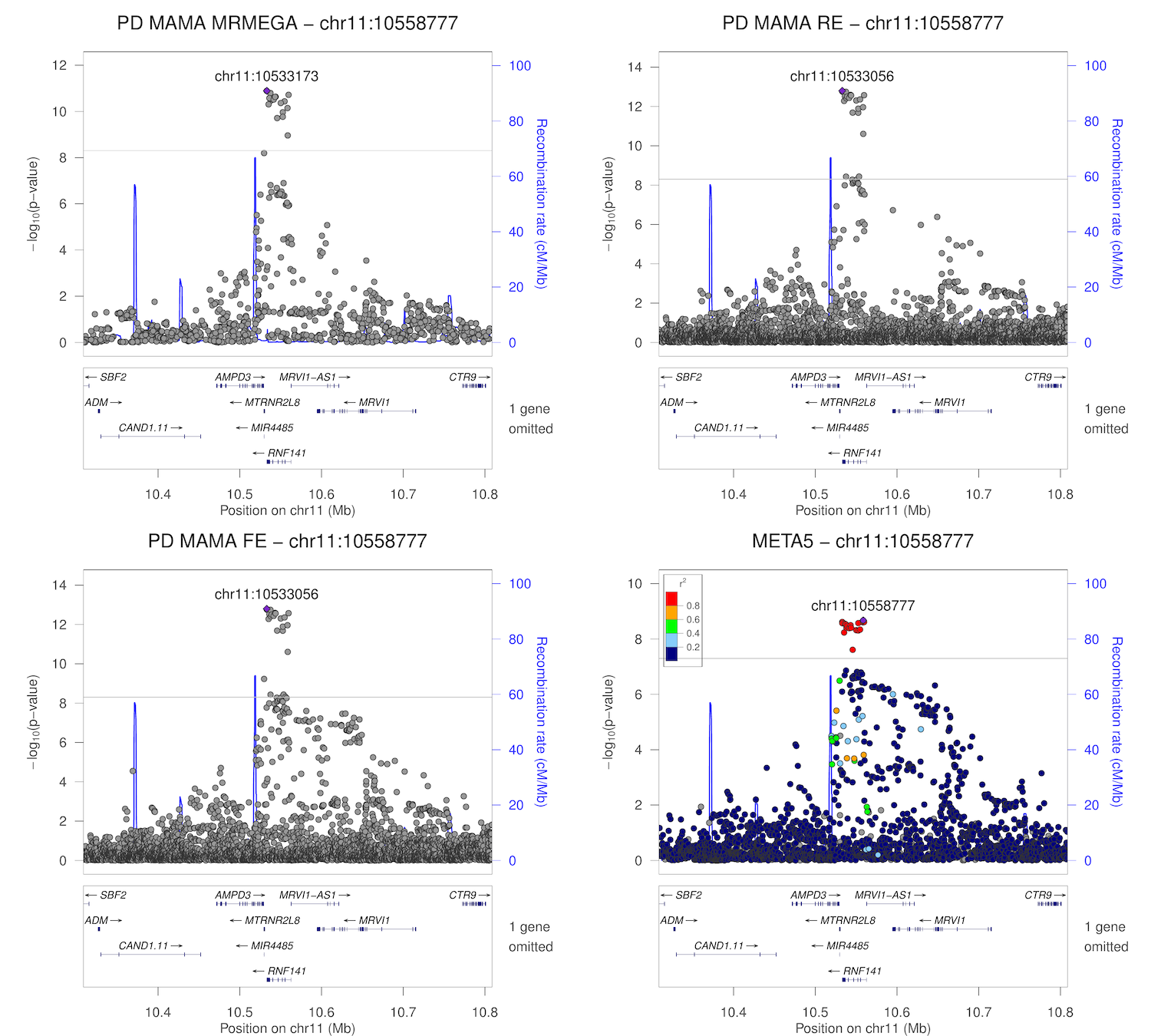

### chr11_83237277-83737277.png

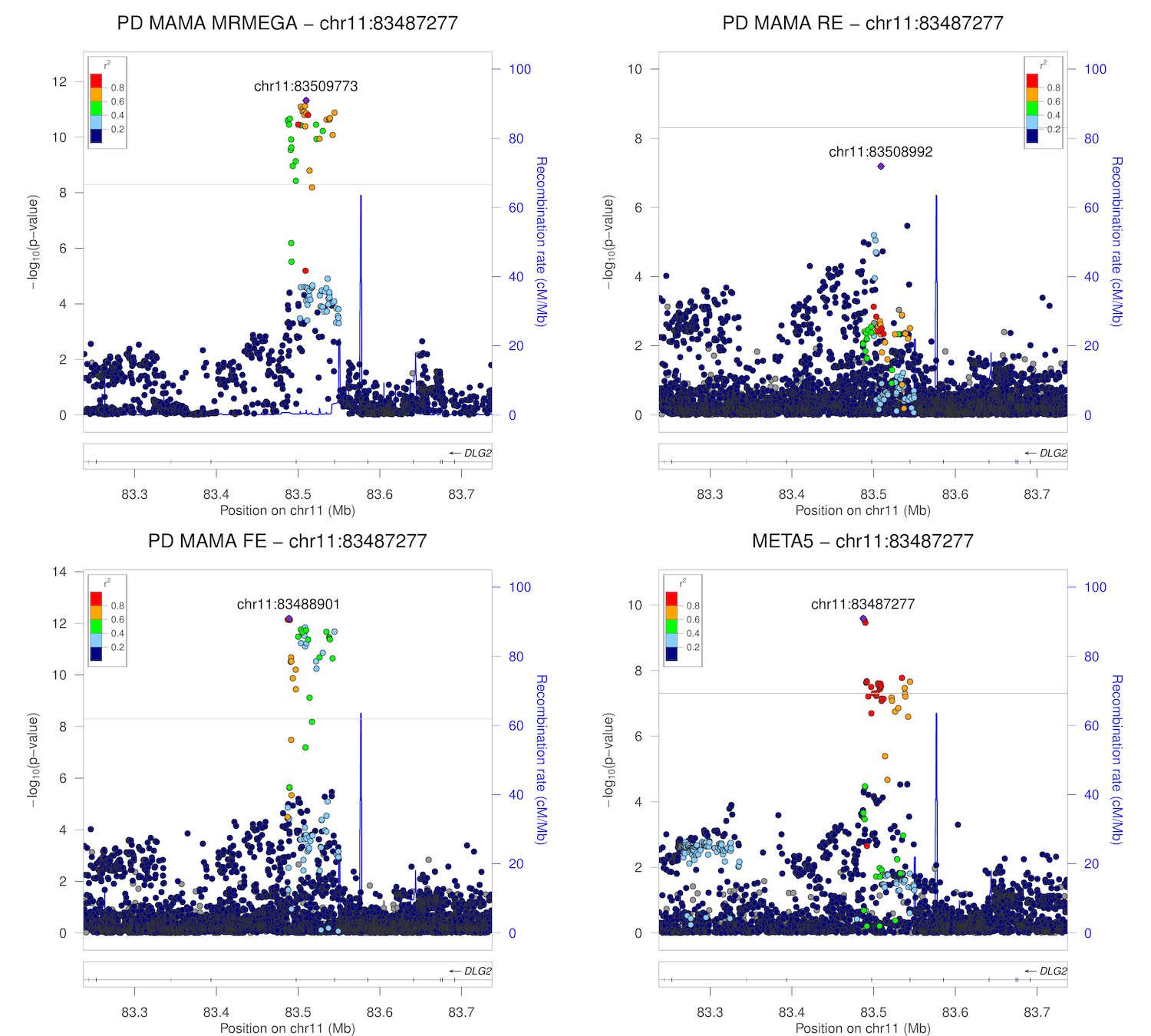

### chr11_133537001-134037001.png

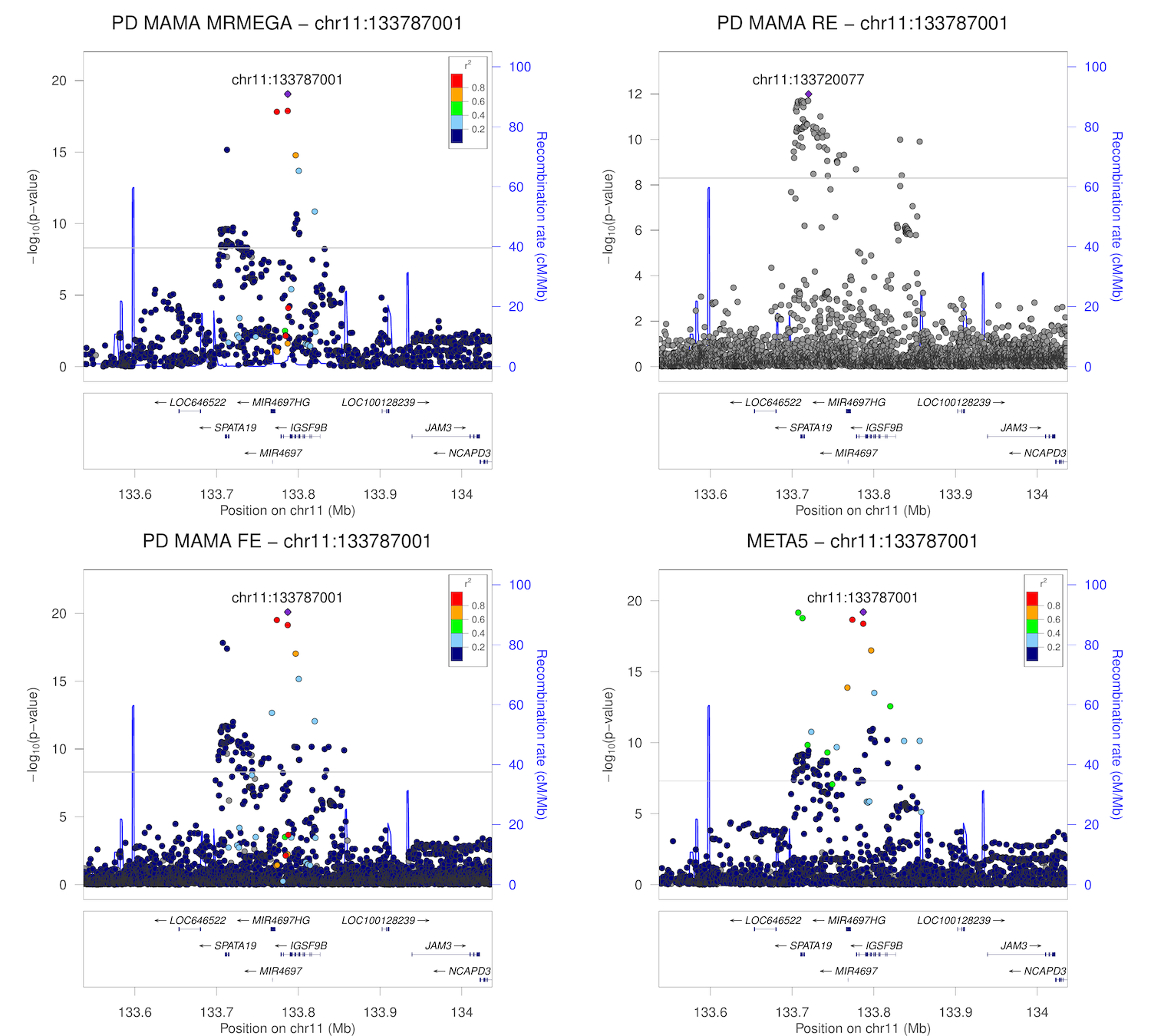

### chr12_40364434-40864434.png

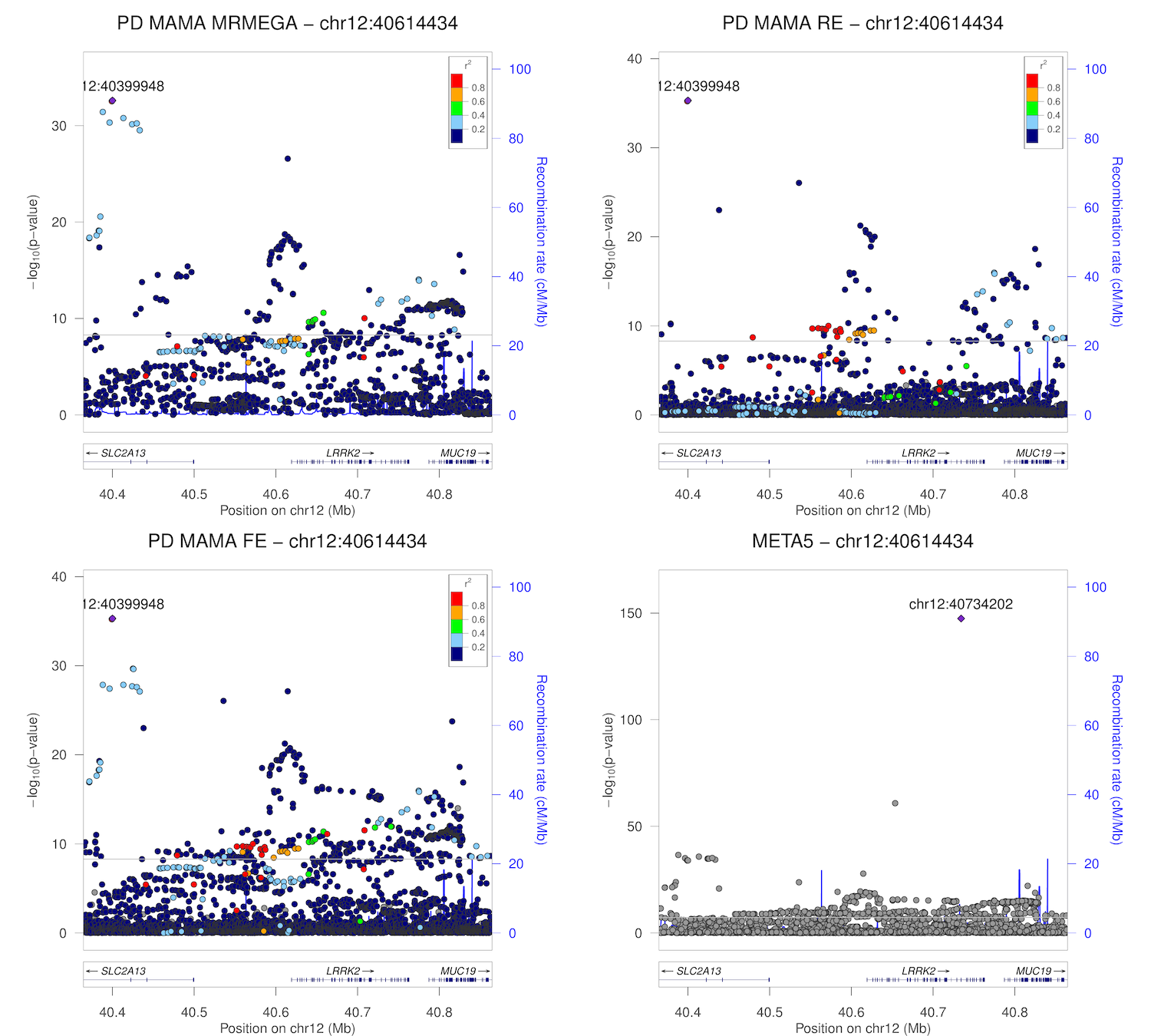

### chr12_40484202-40984202.png

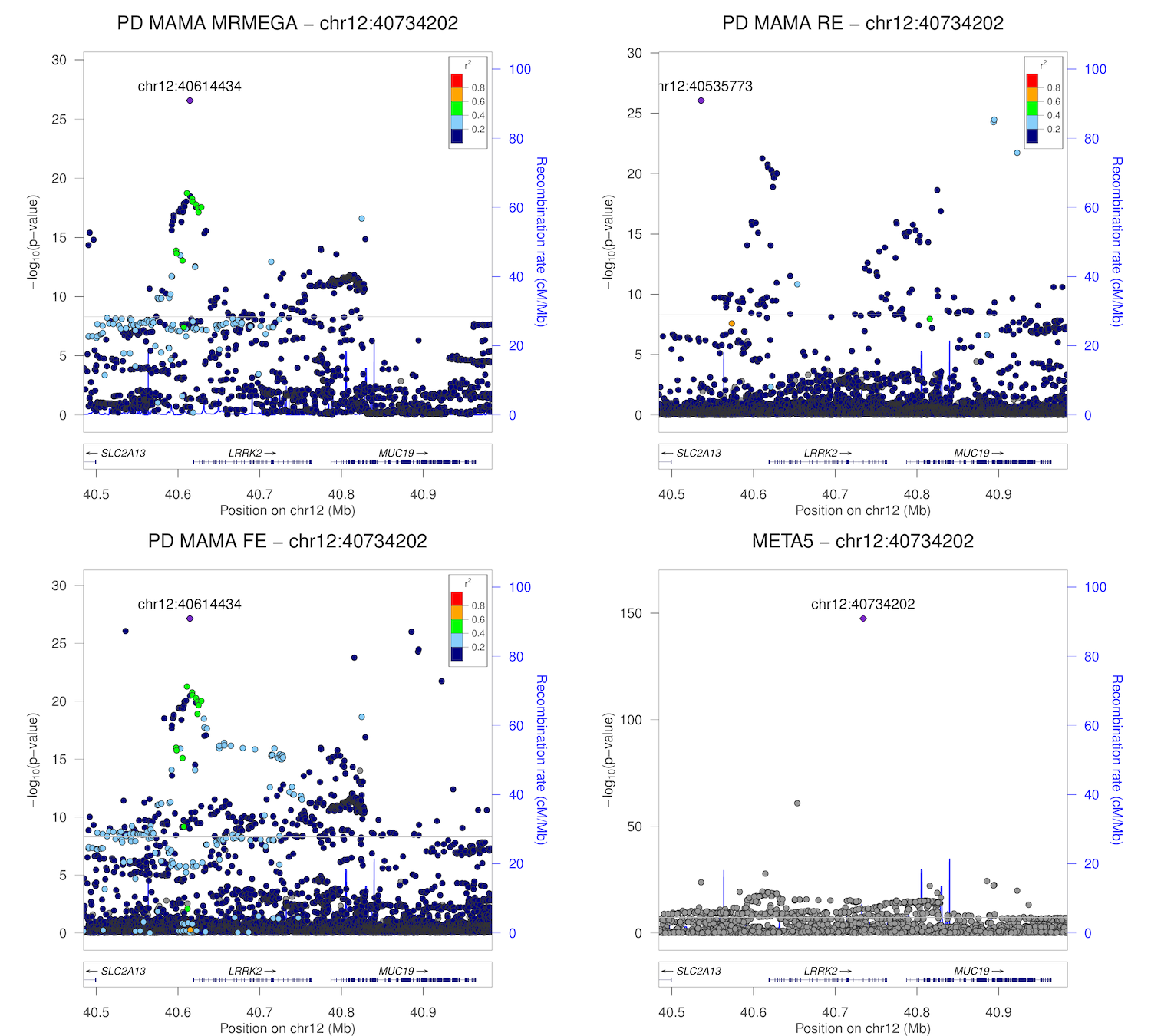

### chr12_46169086-46669086.png

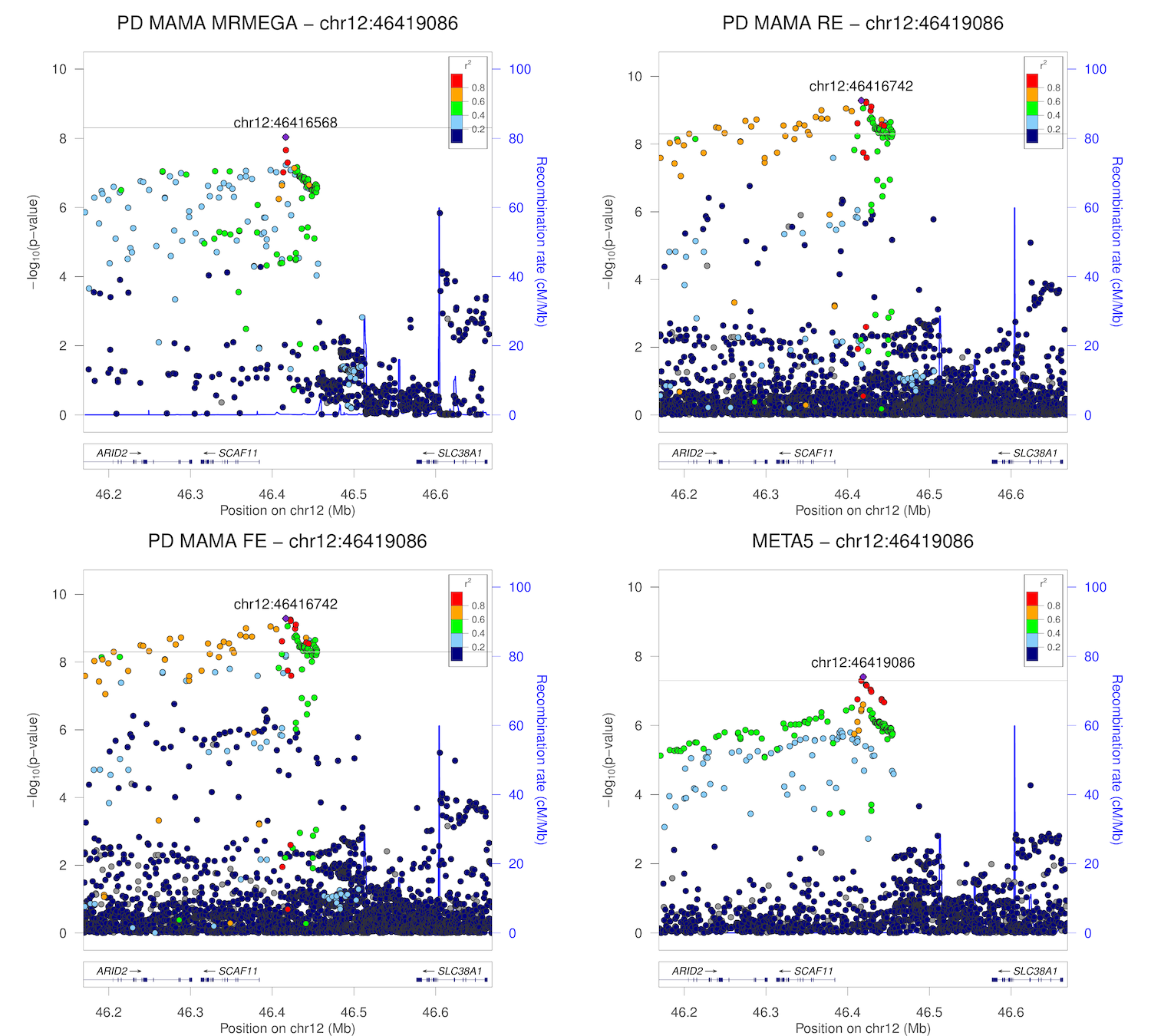

### chr12_132813768-133313768.png

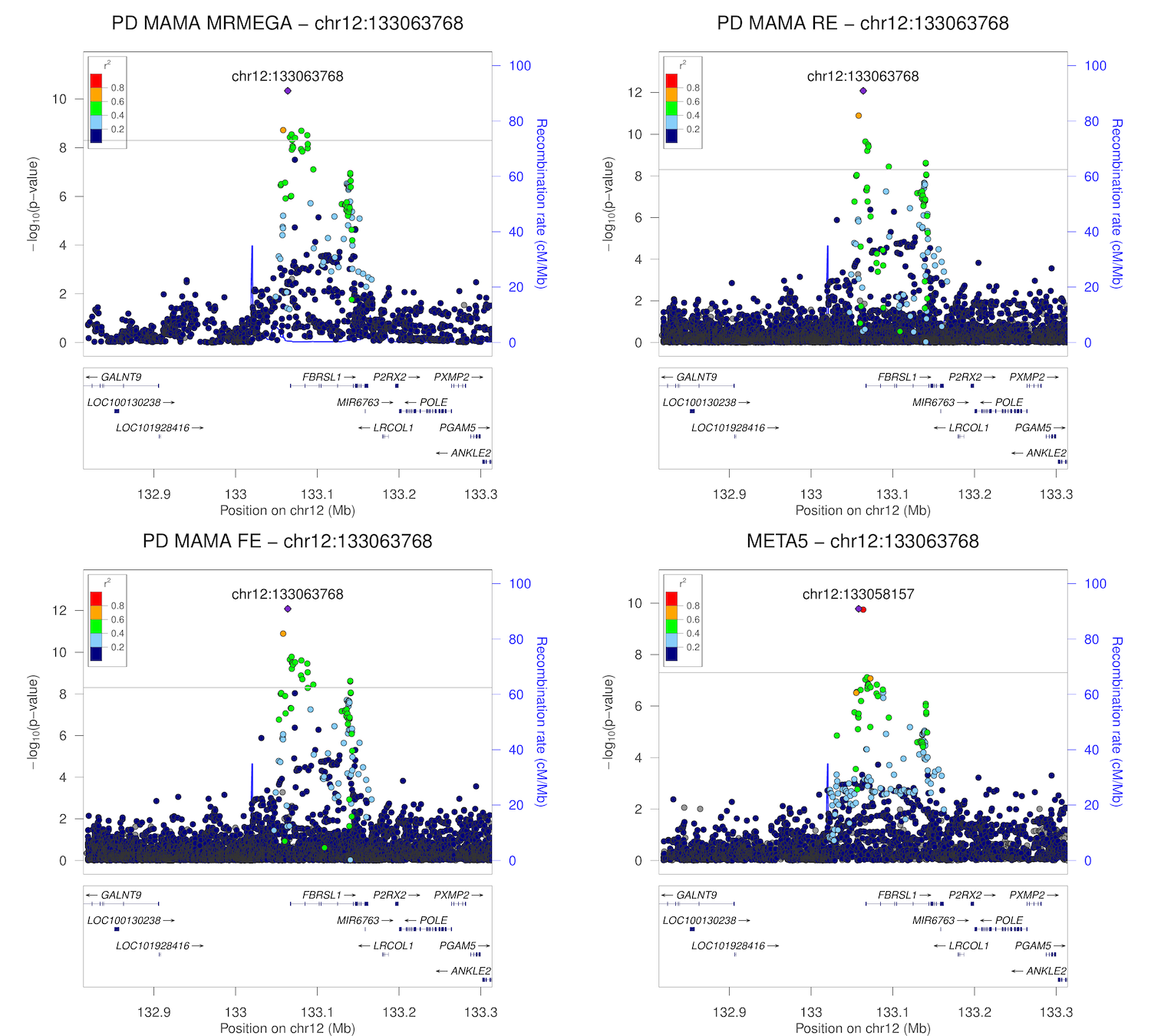

### chr13_49677732-50177732.png

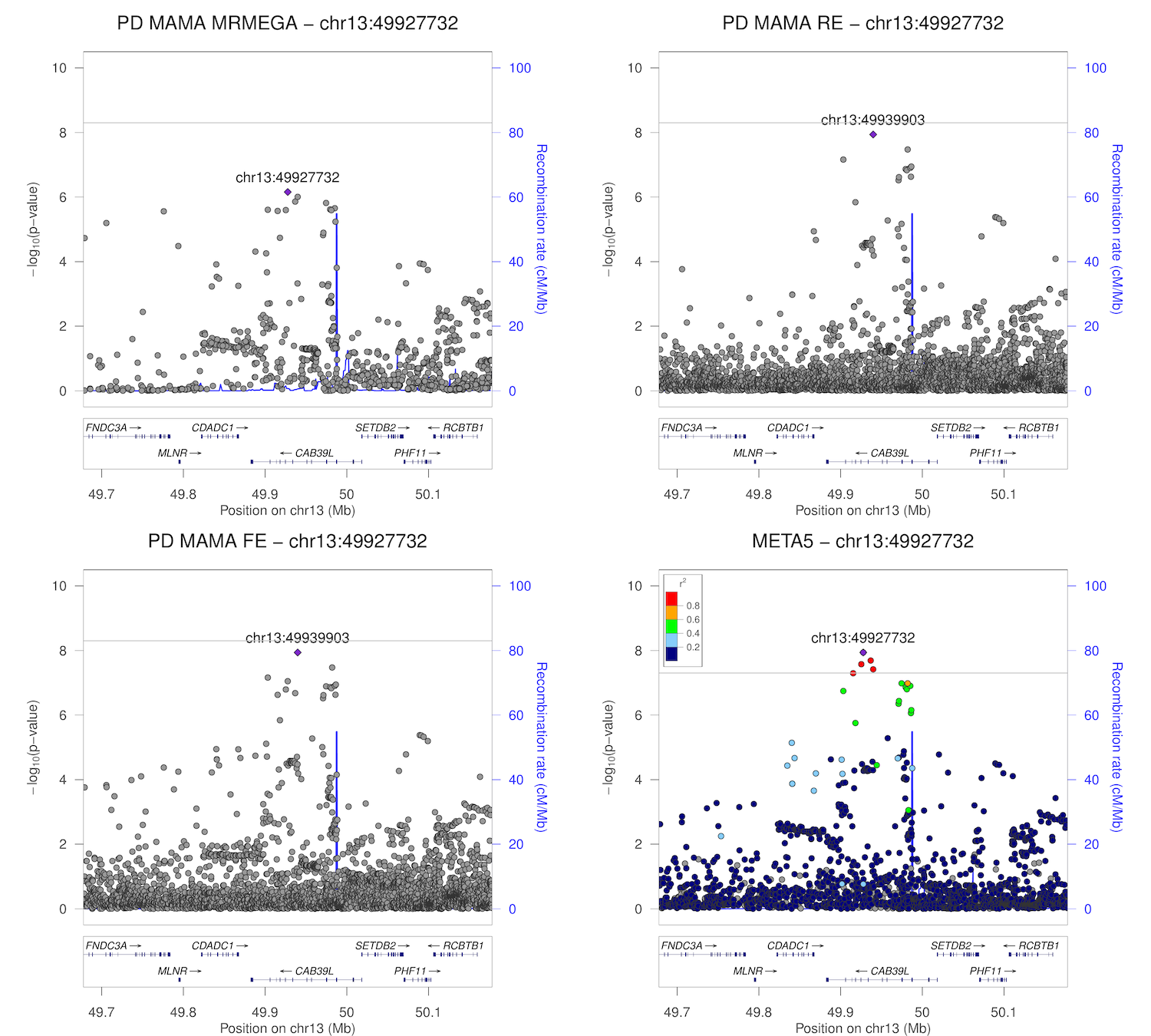

### chr13_110213168-110713168.png

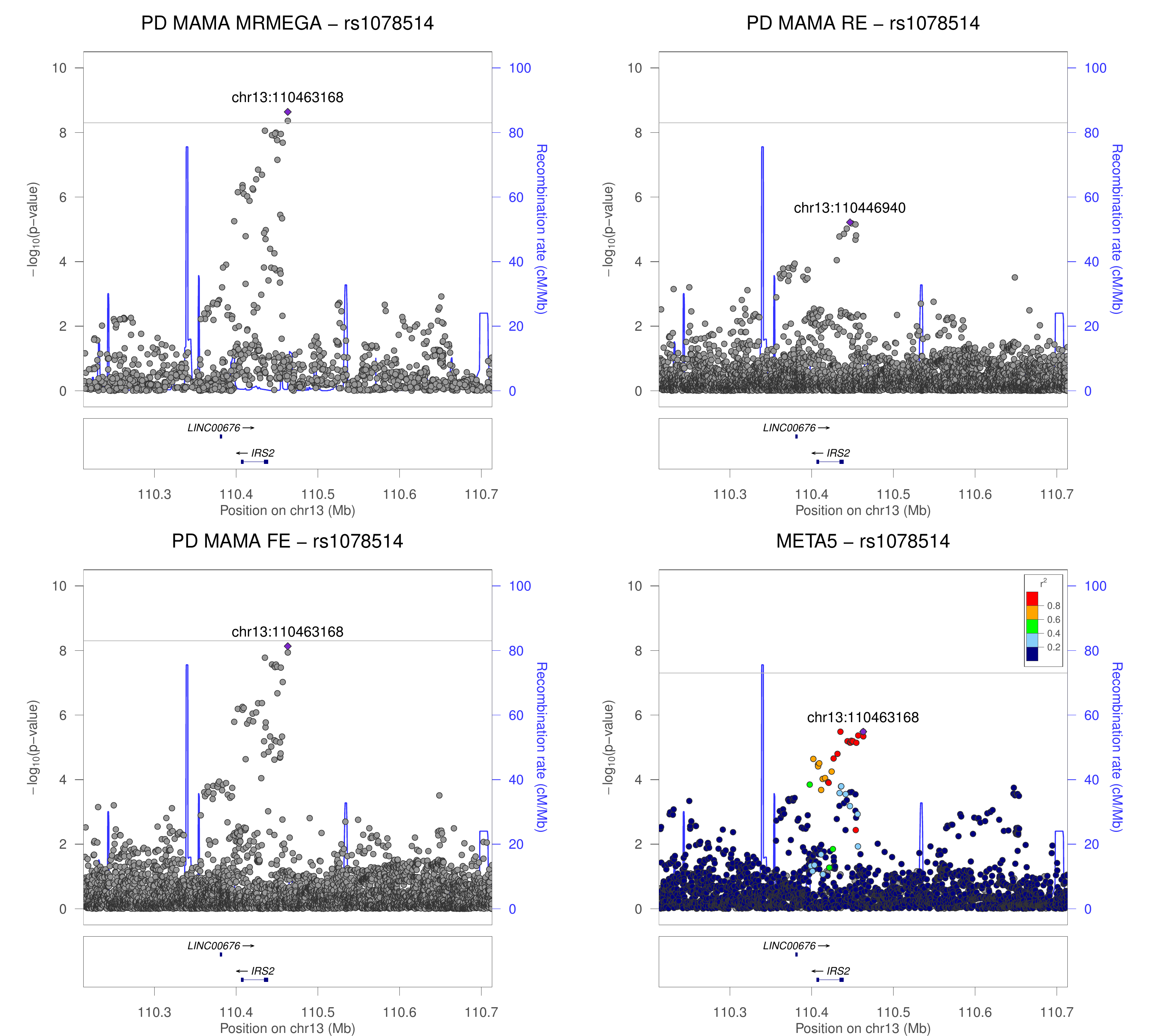

### chr14_37739270-38239270.png

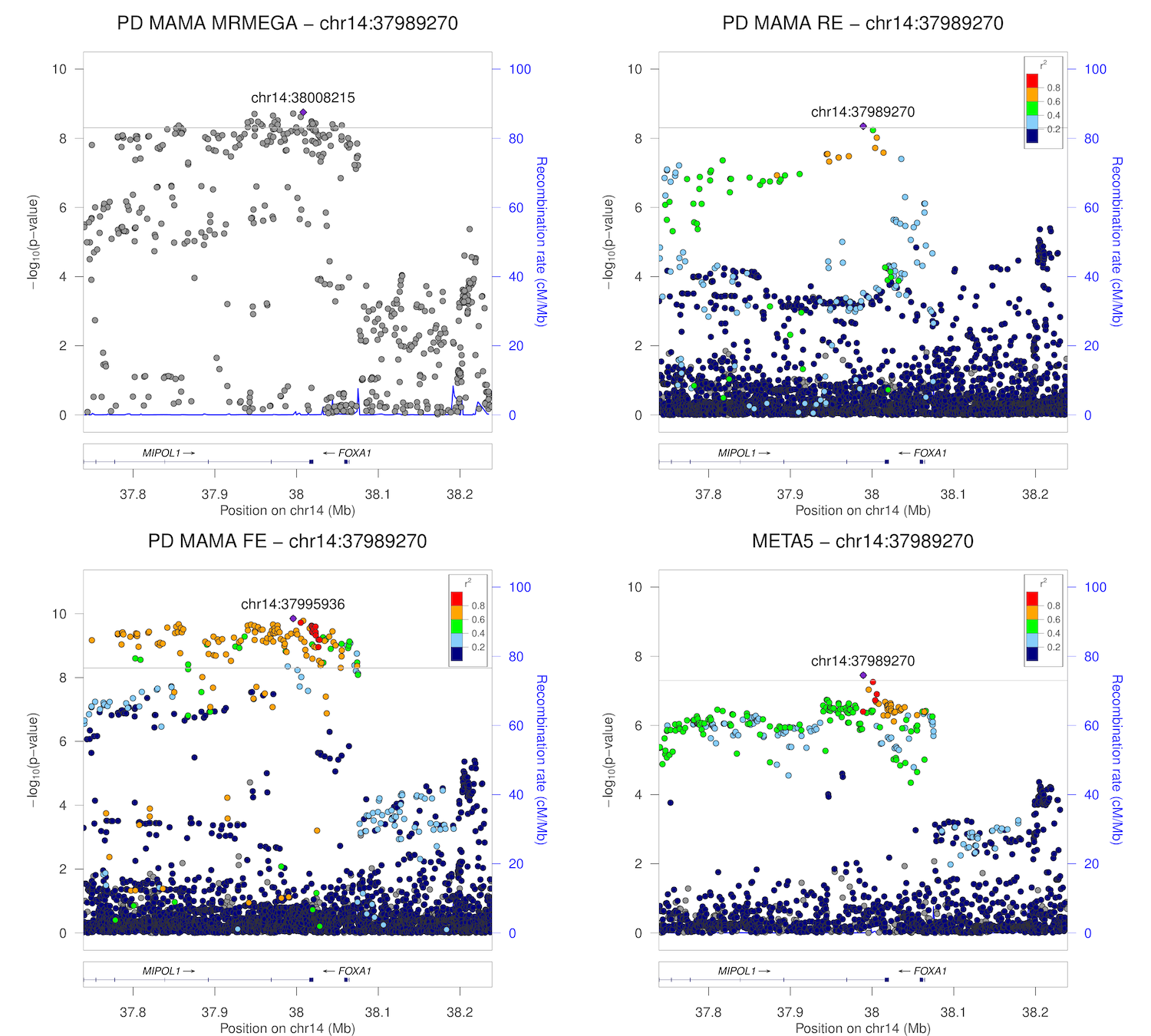

### chr14_55098869-55598869.png

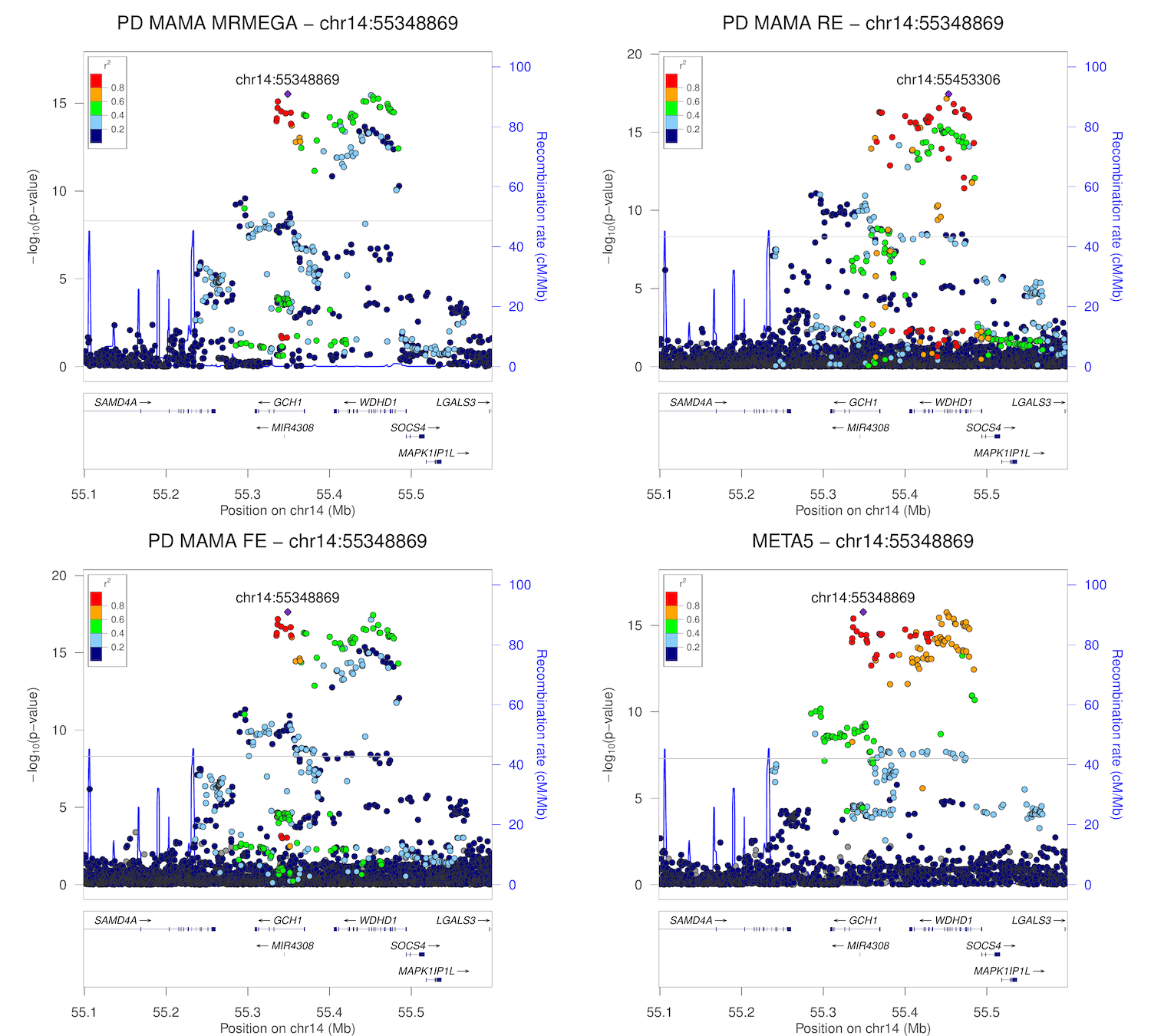

### chr14_88214264-88714264.png

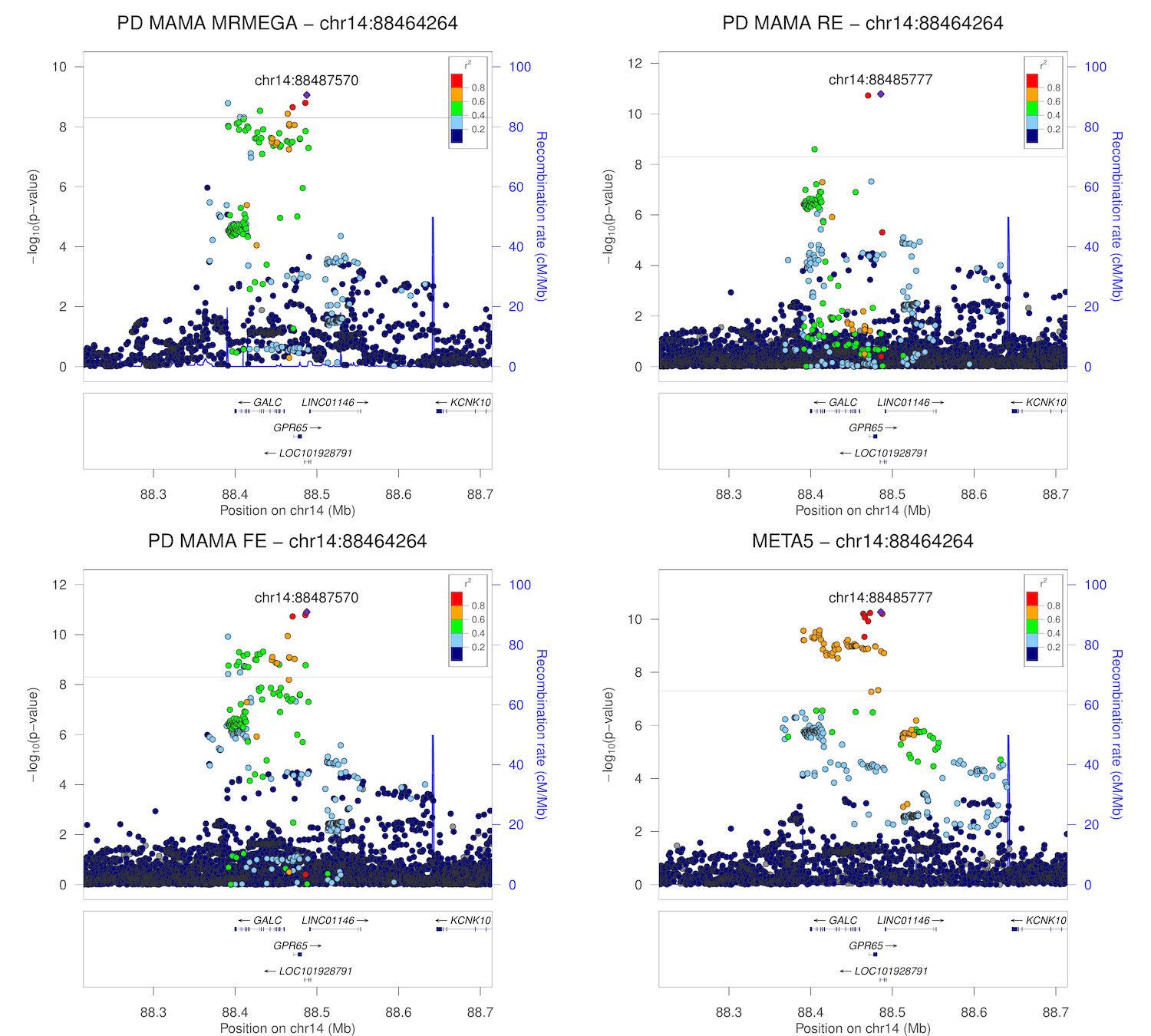

### chr15_50537409-51037409.png

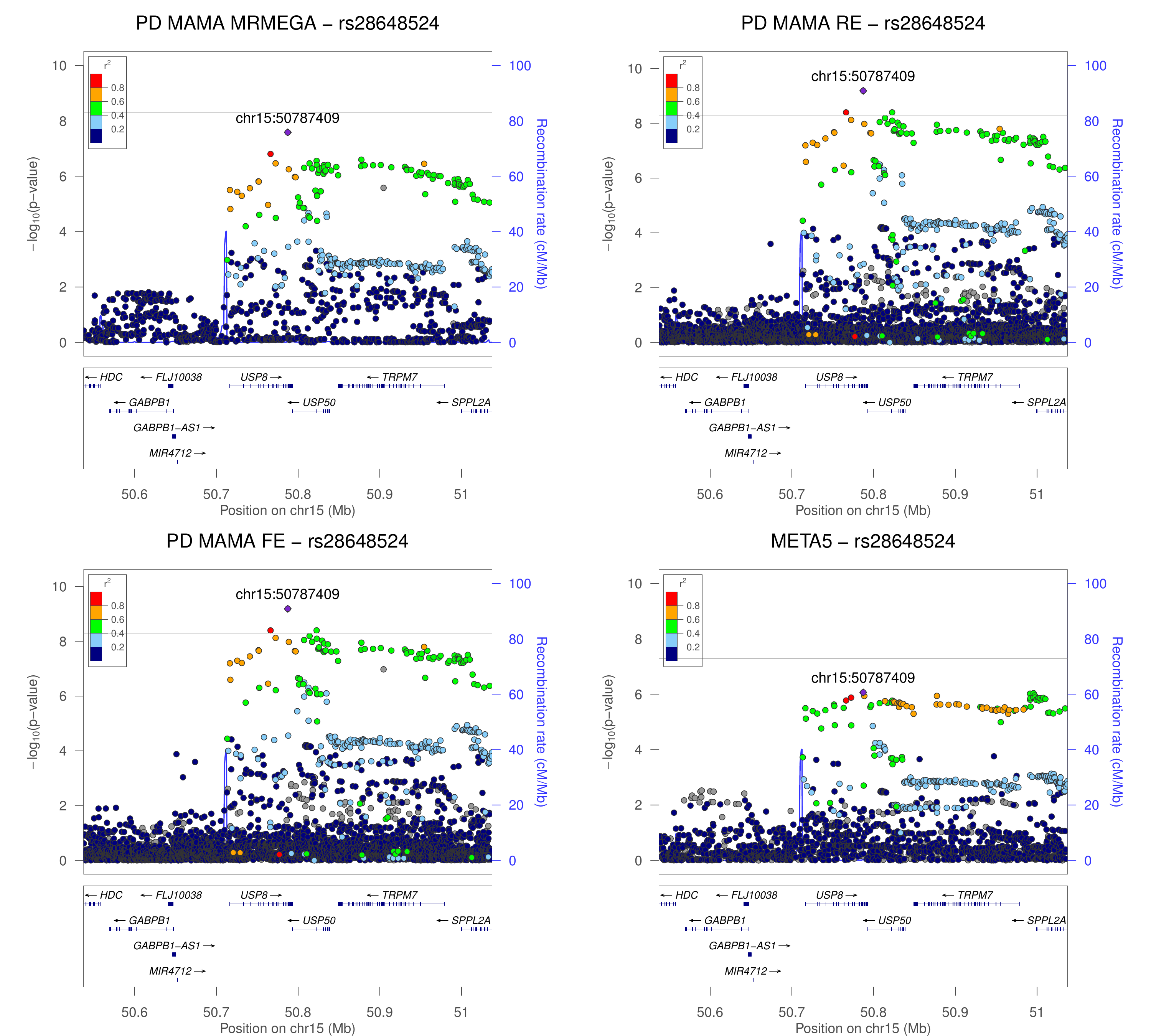

### chr15_61747385-62247385.png

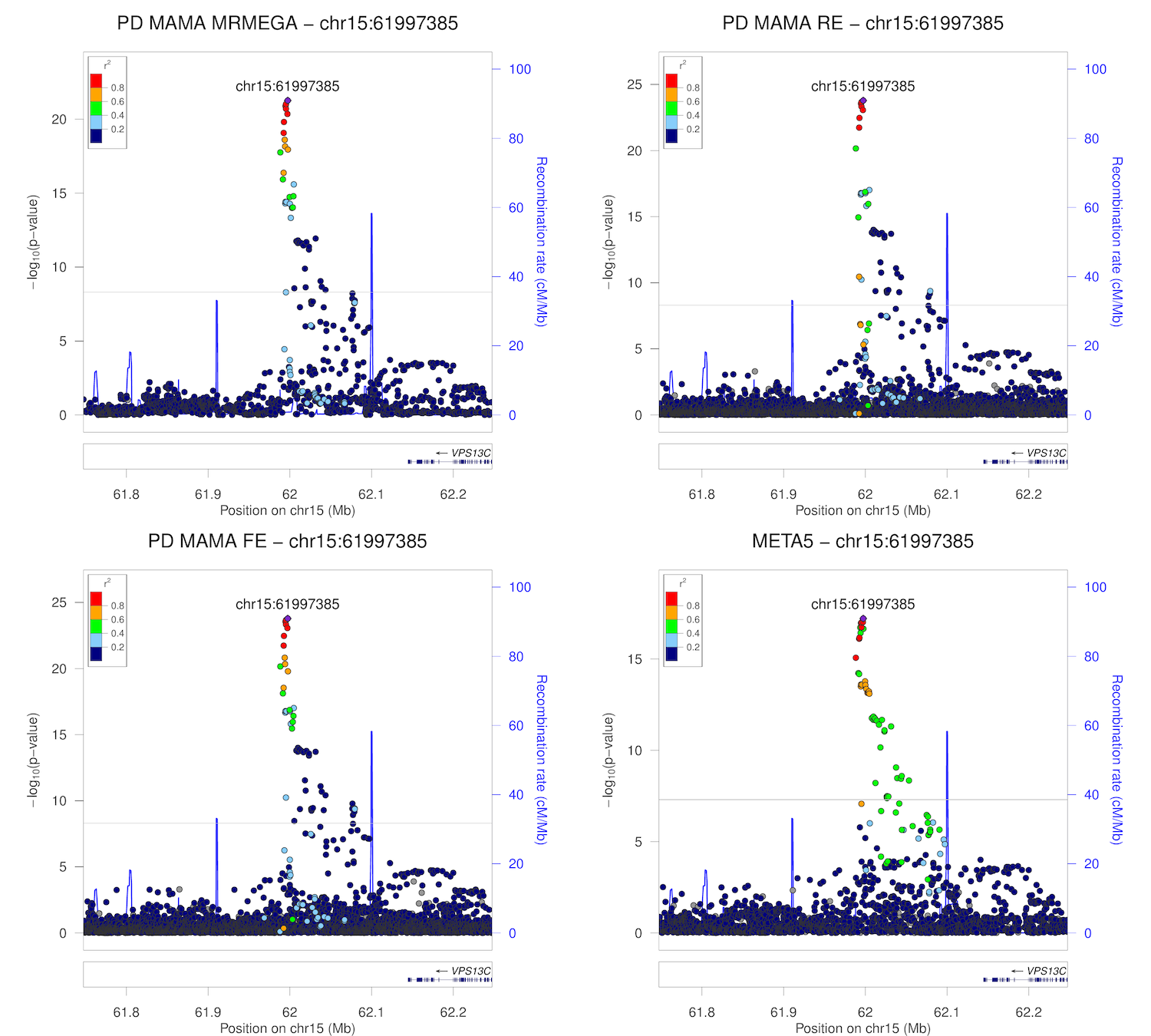

### chr16_19027493-19527493.png

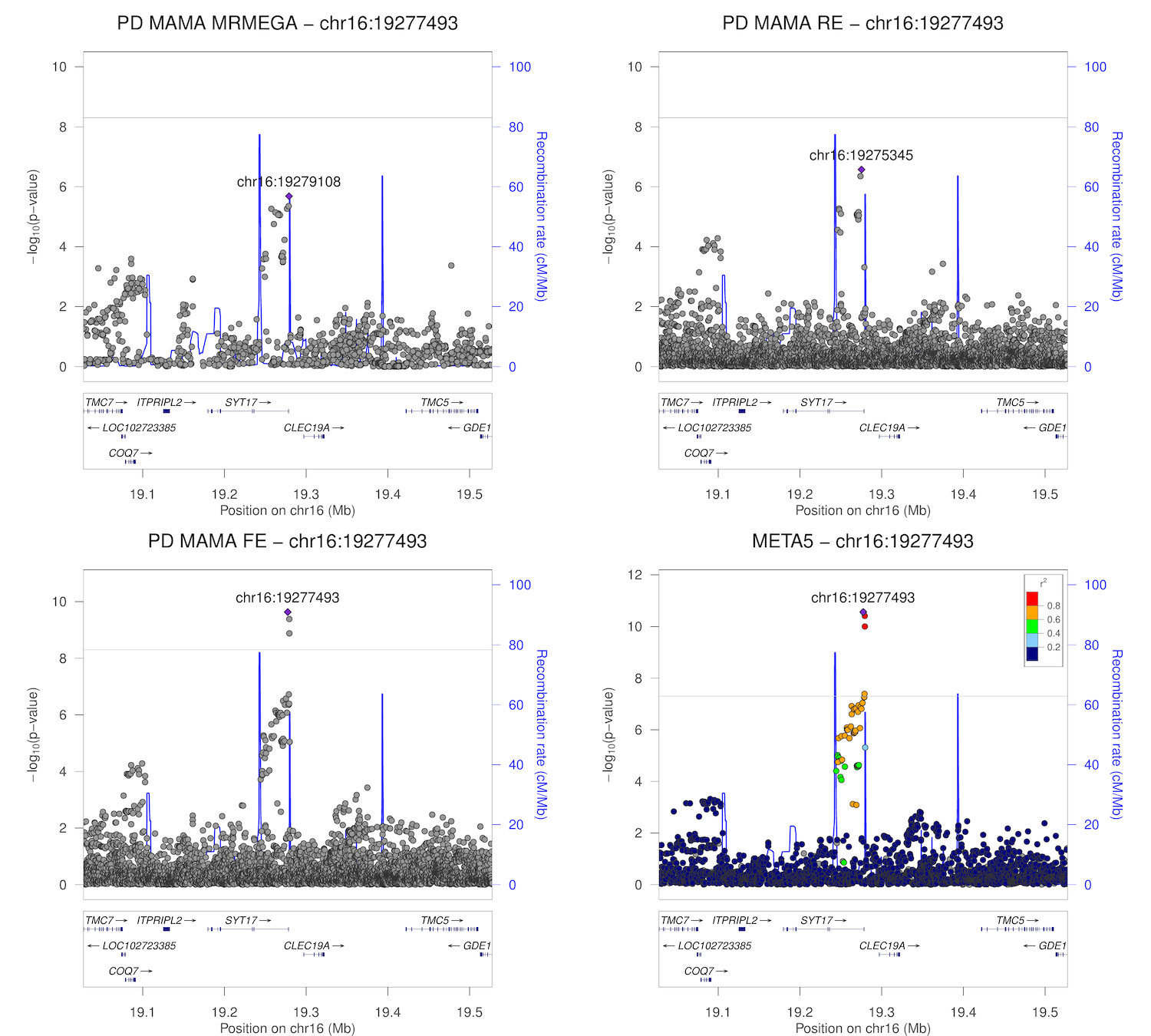

### chr16_30727799-31227799.png

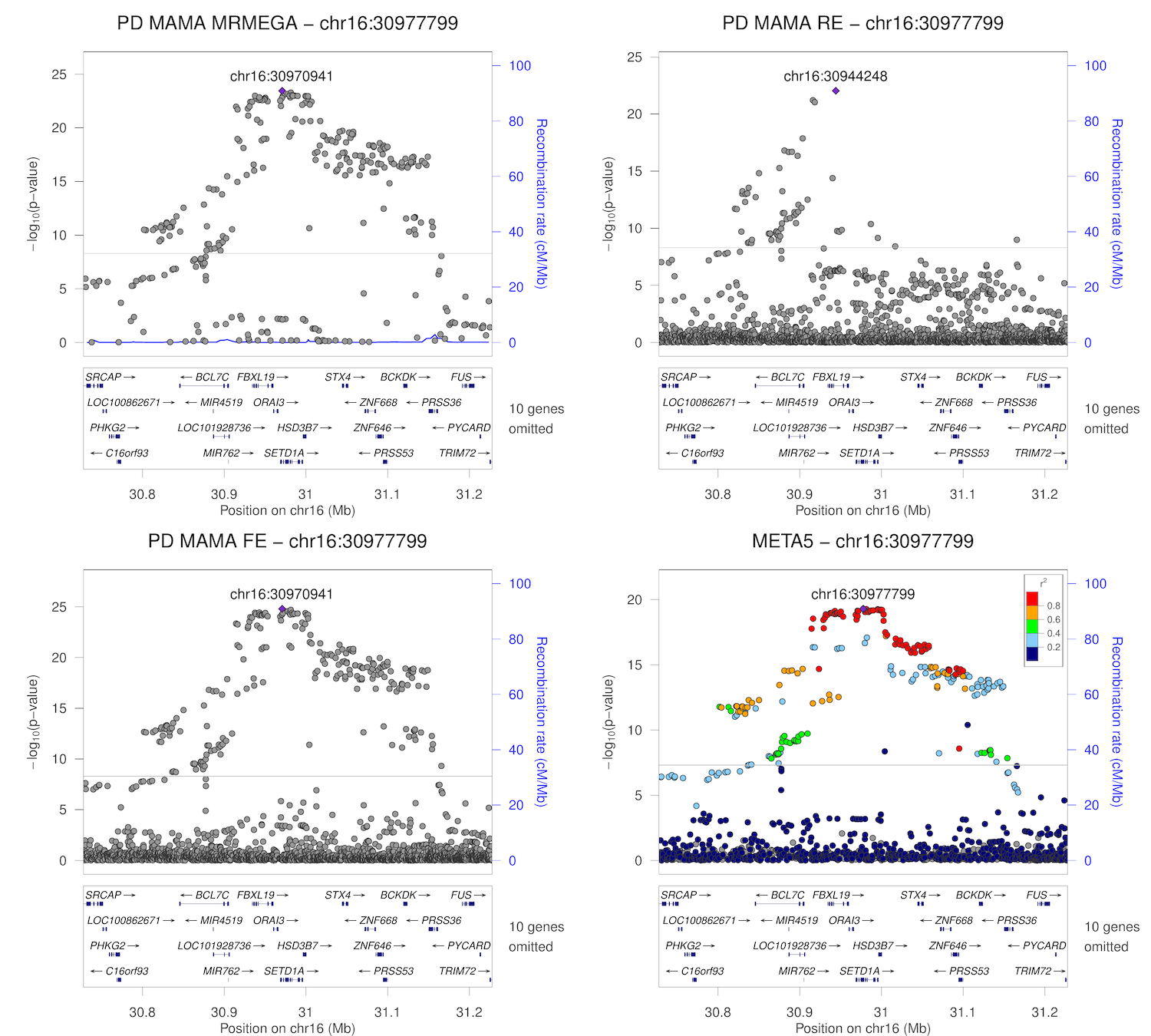

### chr16_50486656-50986656.png

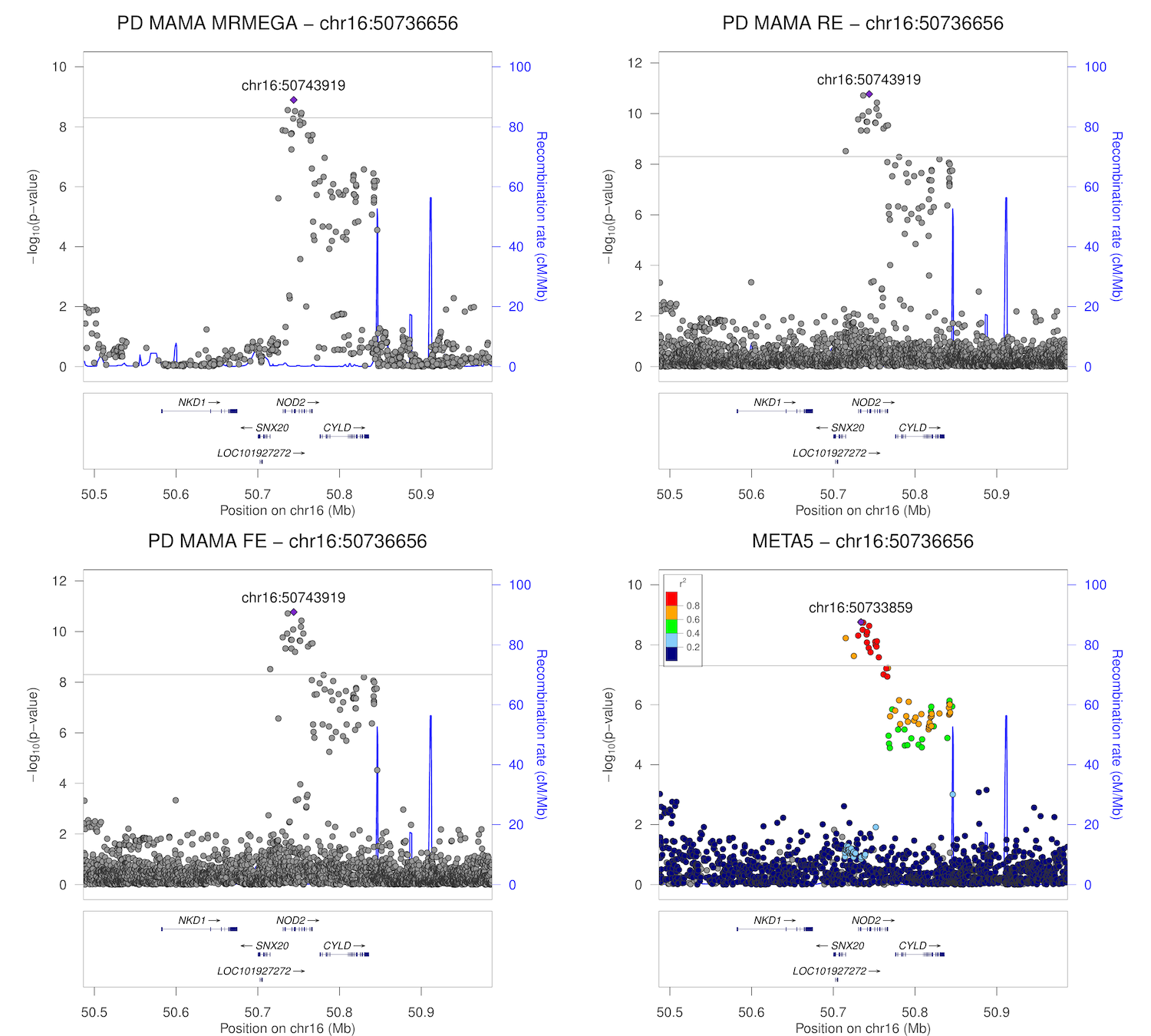

### chr16_52386242-52886242.png

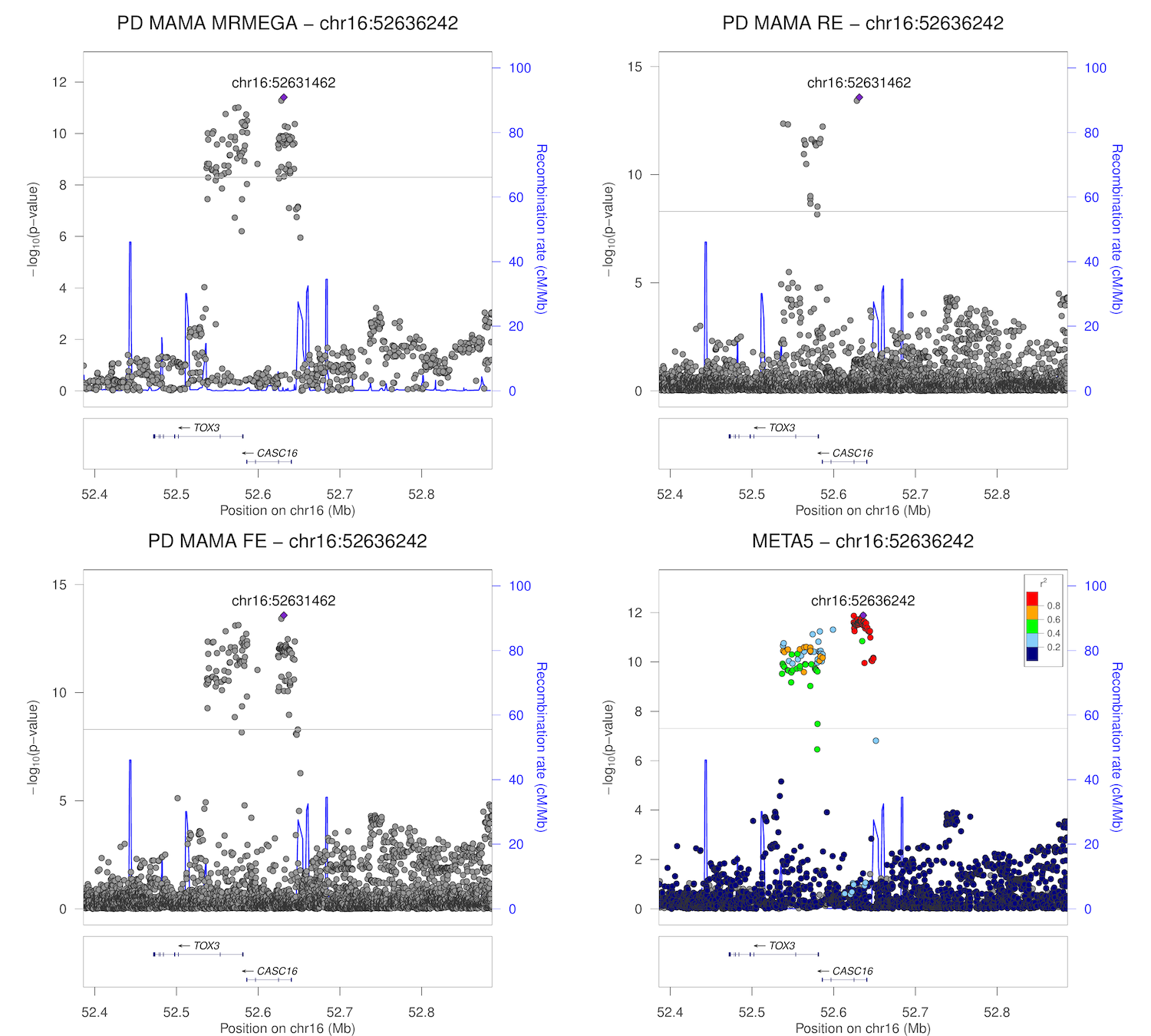

### chr16_52719426-53219426.png

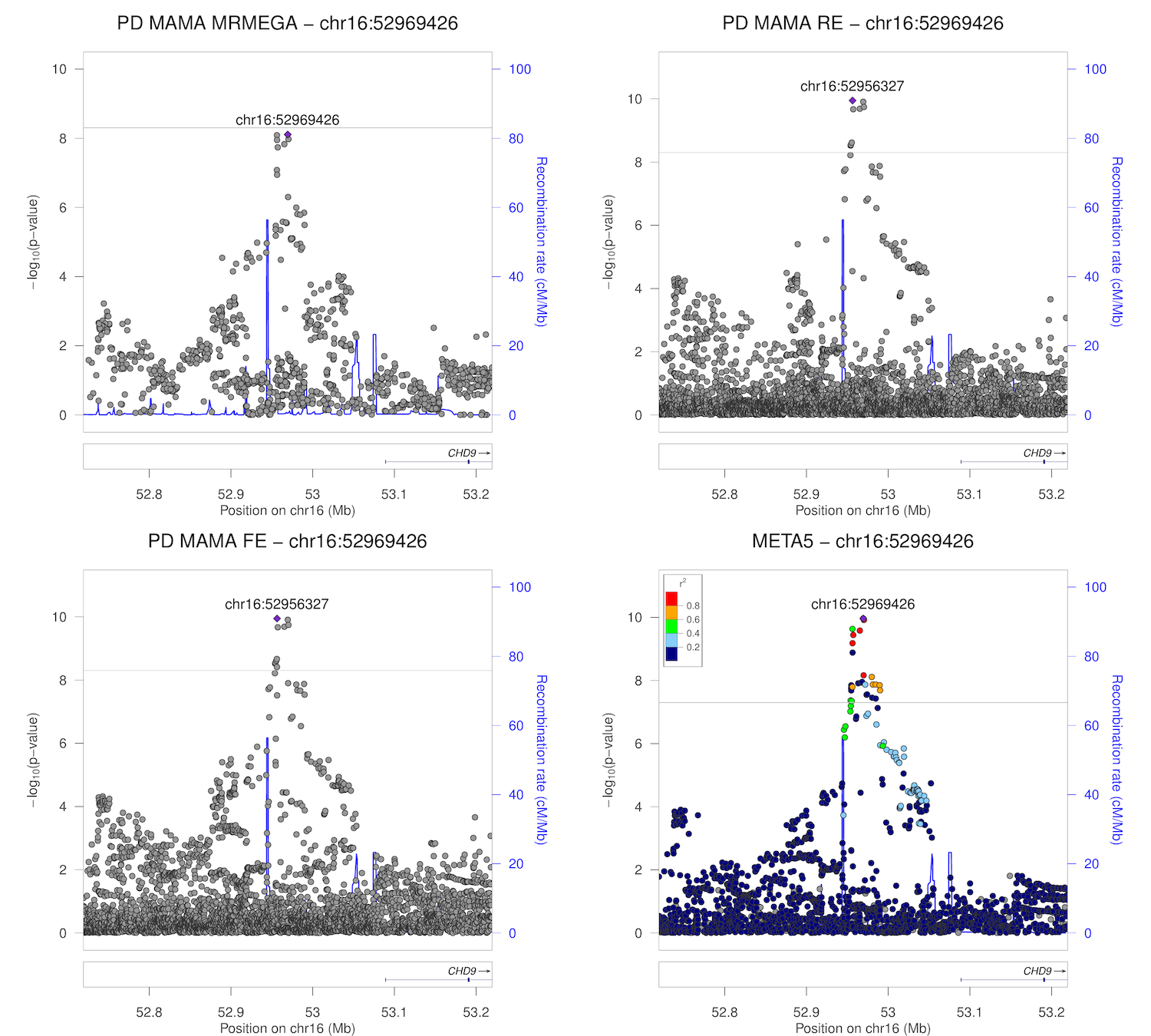

### chr17_7105621-7605621.png

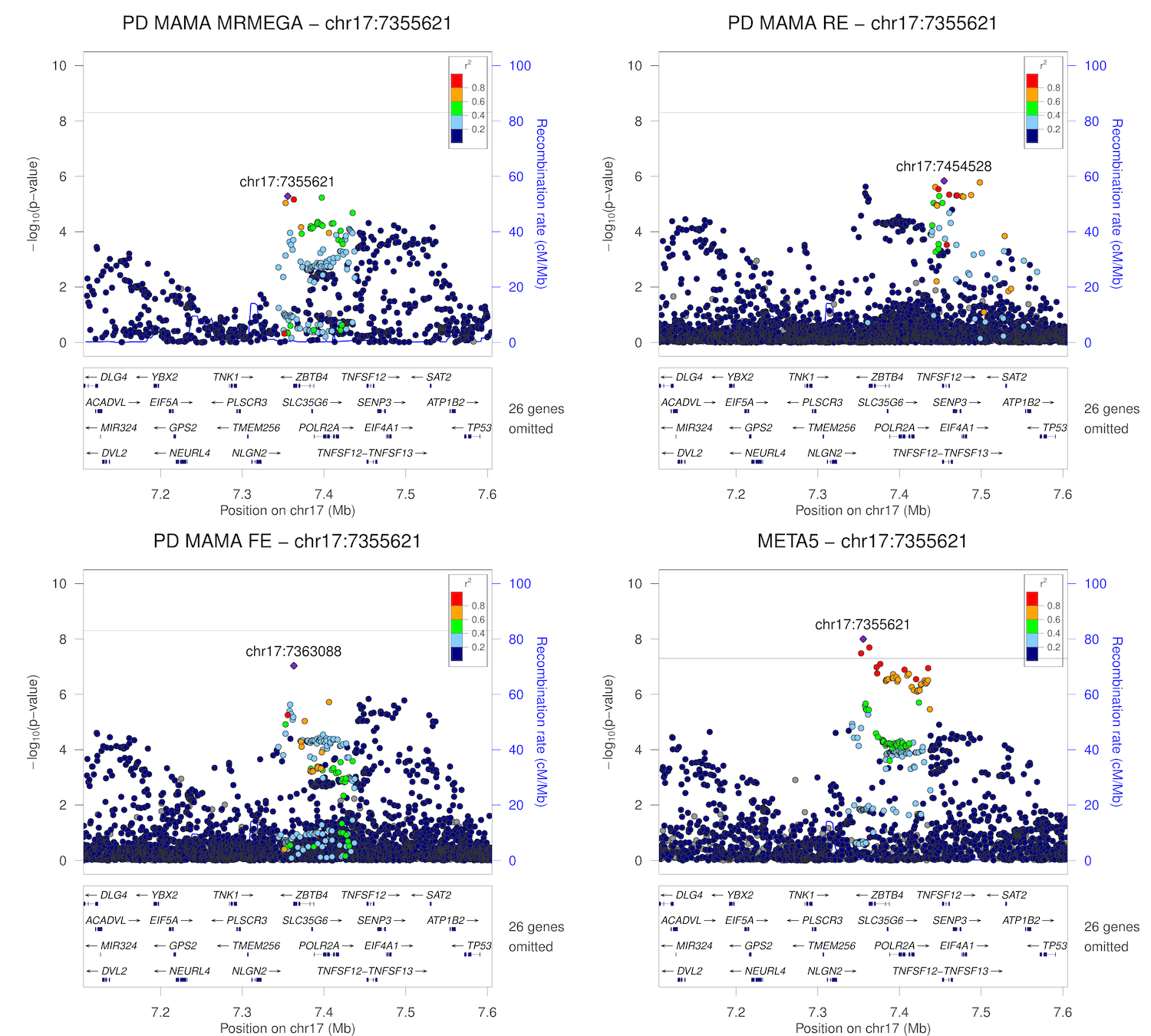

### chr17_15984260-16484260.png

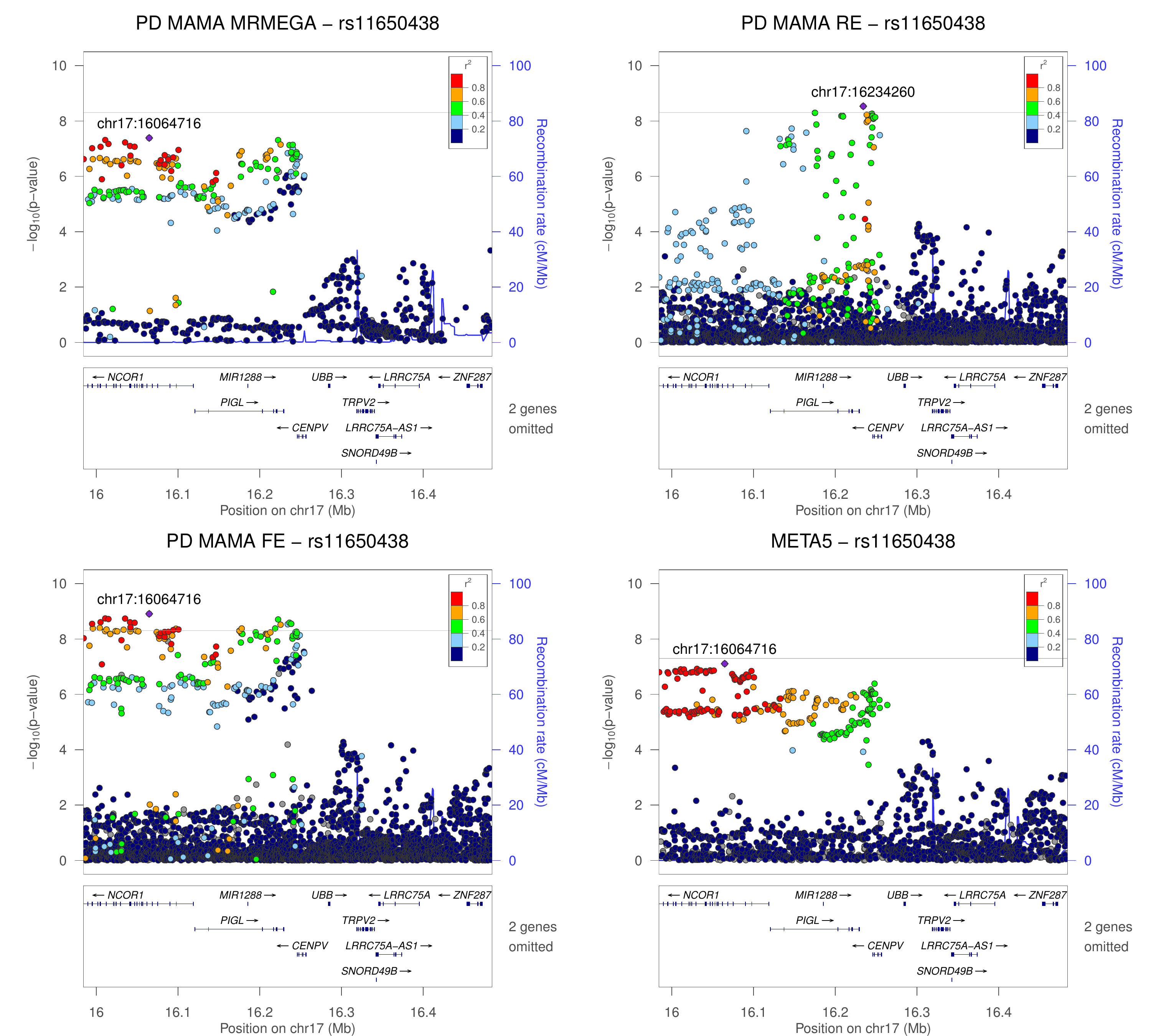

### chr17_40491013-40991013.png

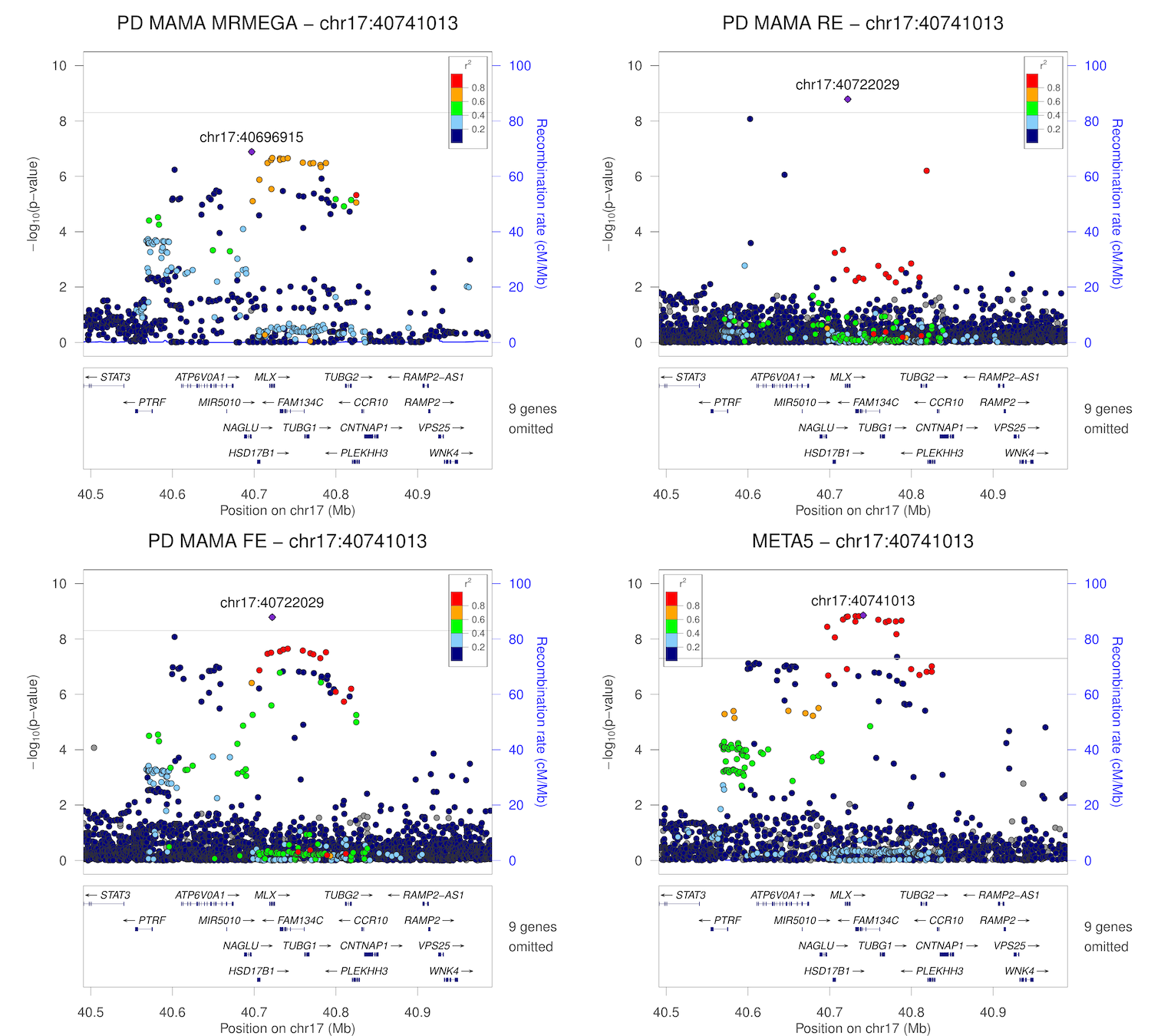

### chr17_42184630-42684630.png

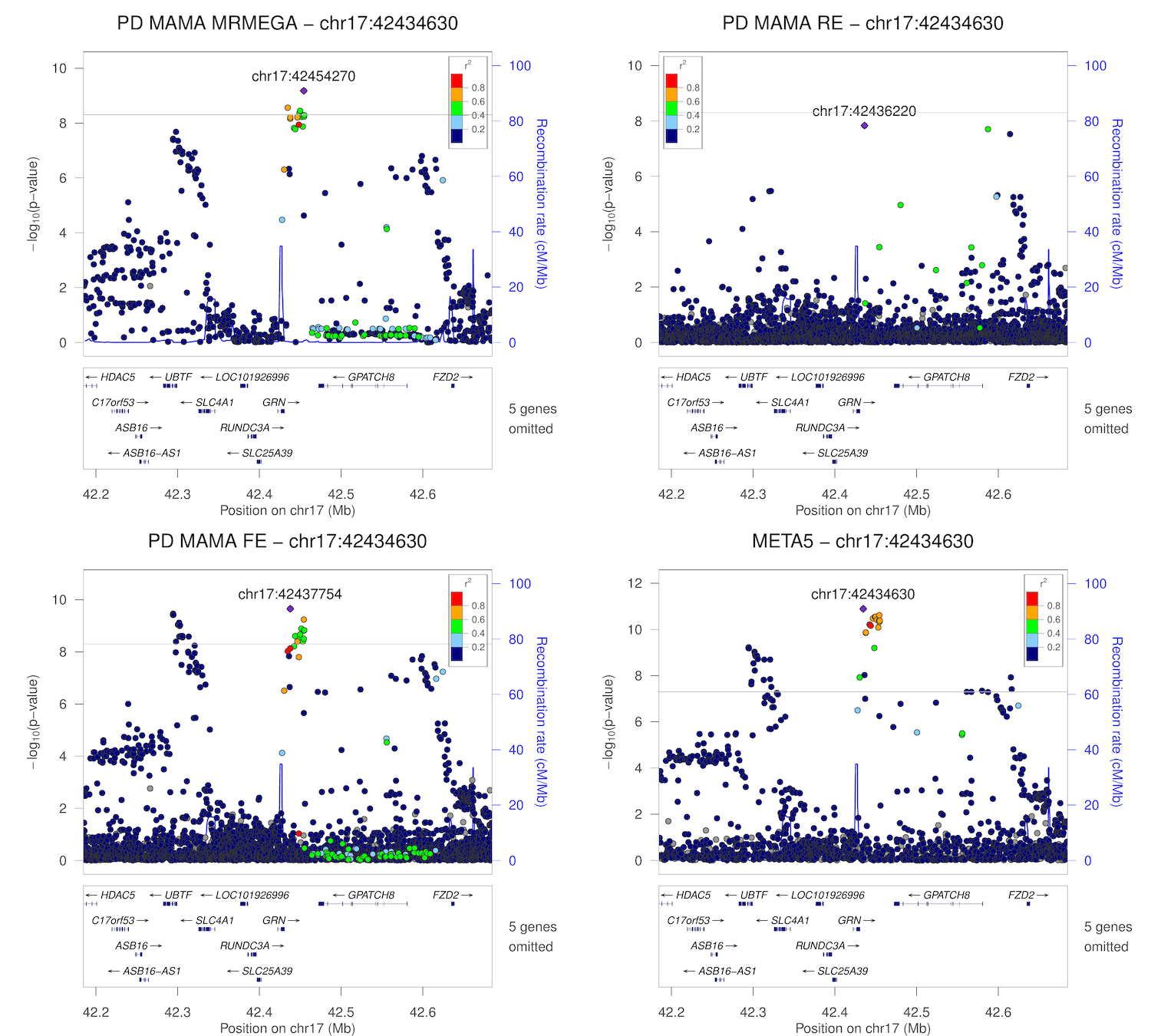

### chr17_43494203-43994203.png

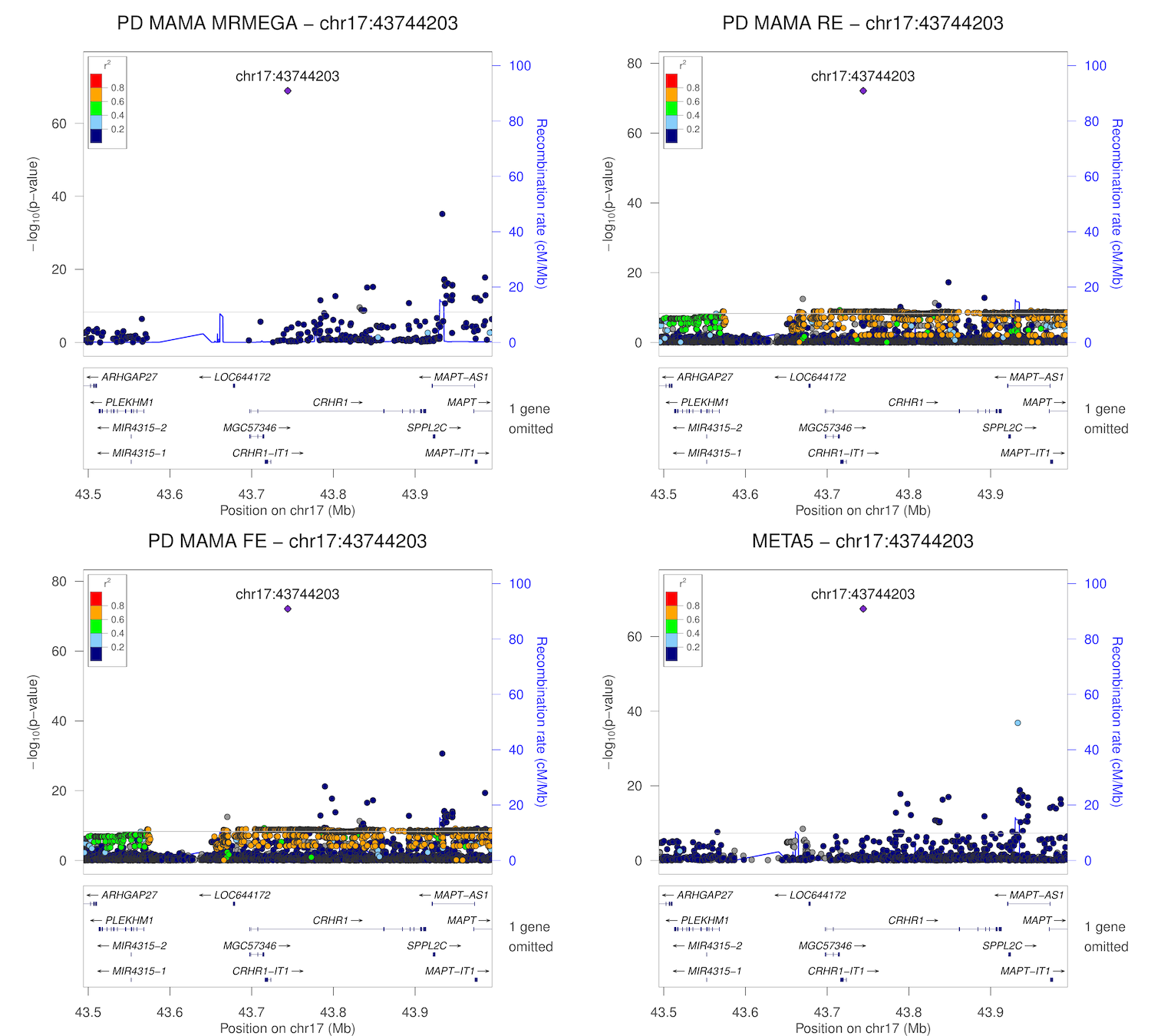

### chr17_44616805-45116805.png

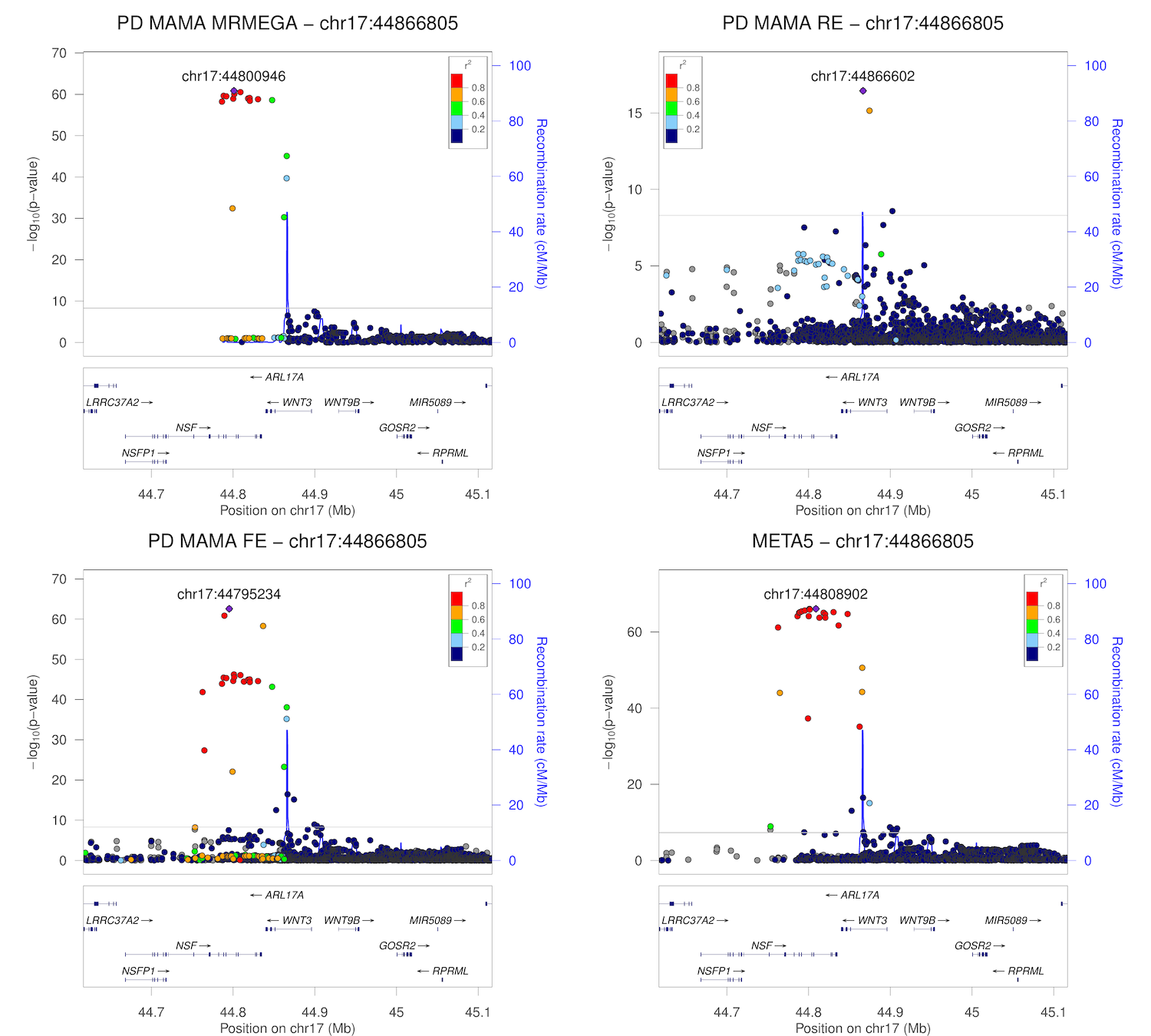

### chr17_59667366-60167366.png

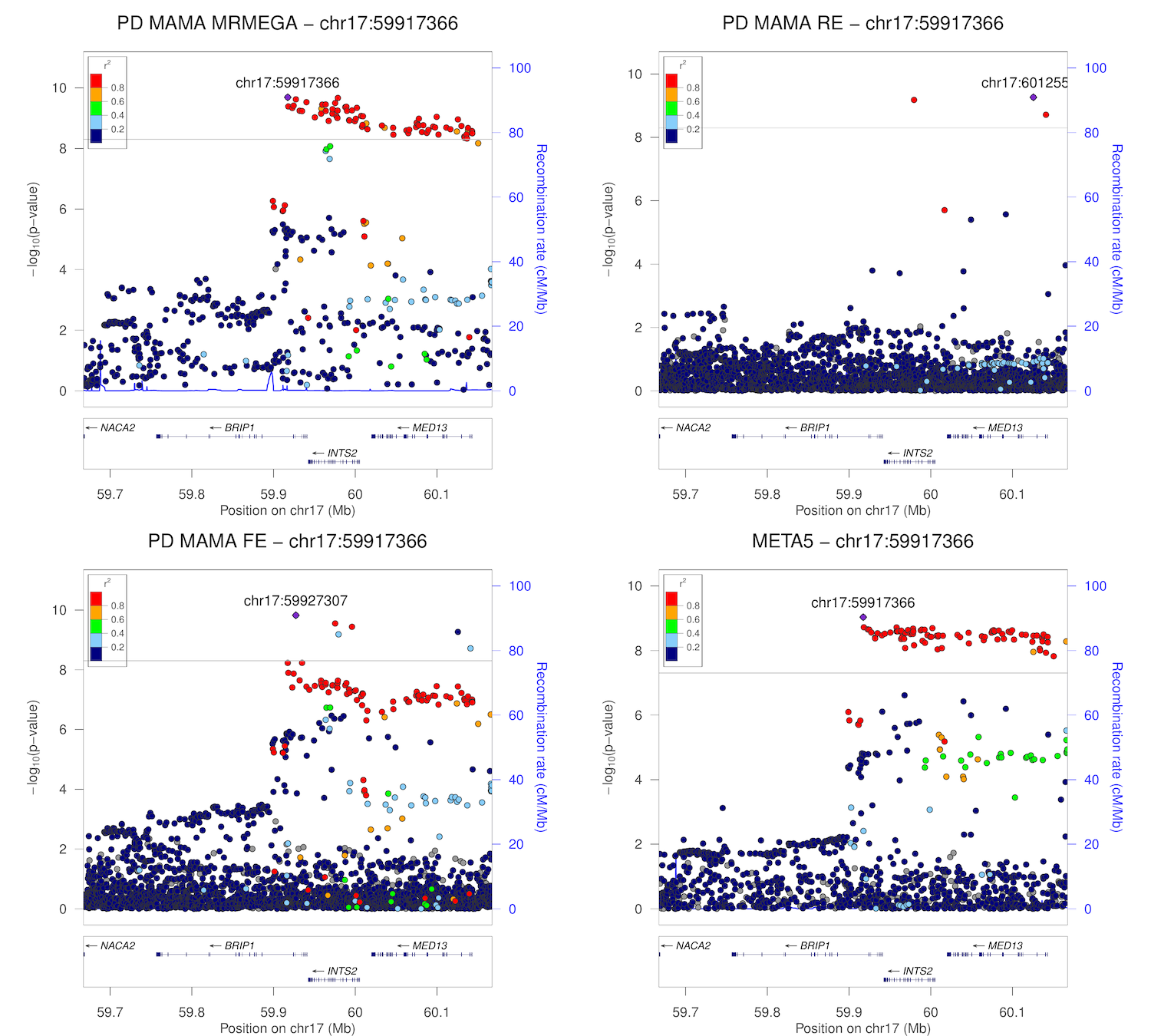

### chr17_76175480-76675480.png

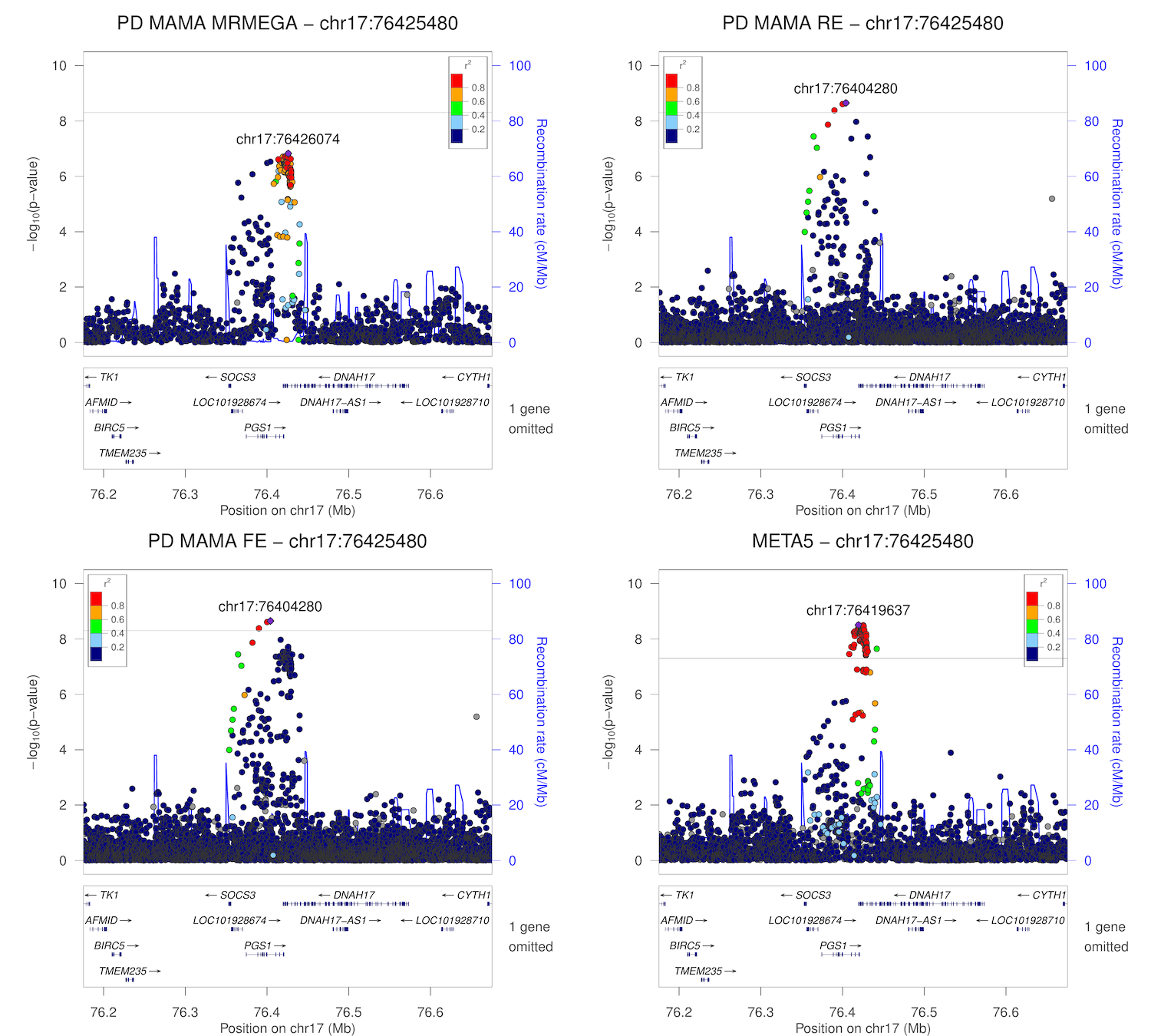

### chr17_79795086-80295086.png

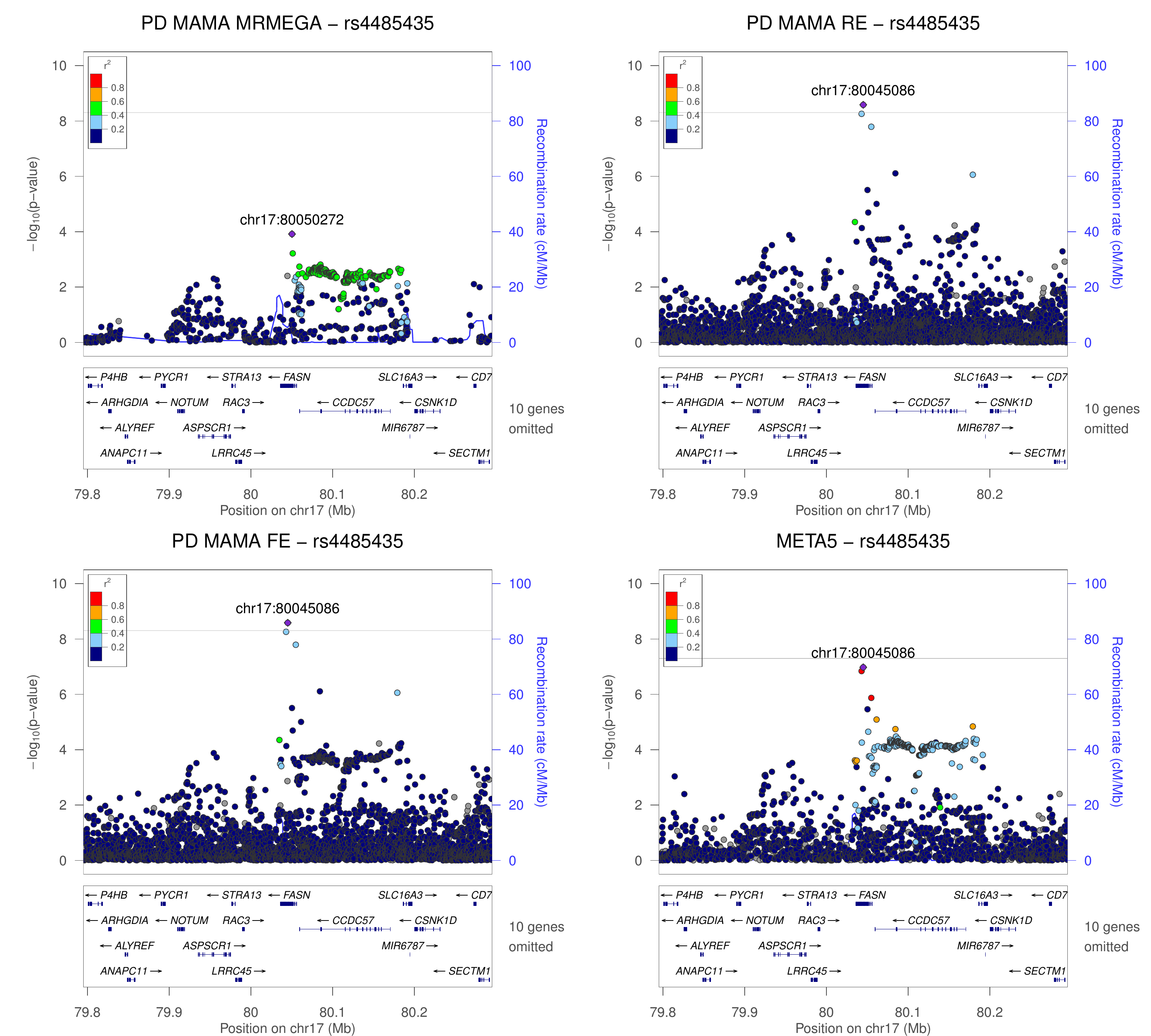

### chr18_31054318-31554318.png

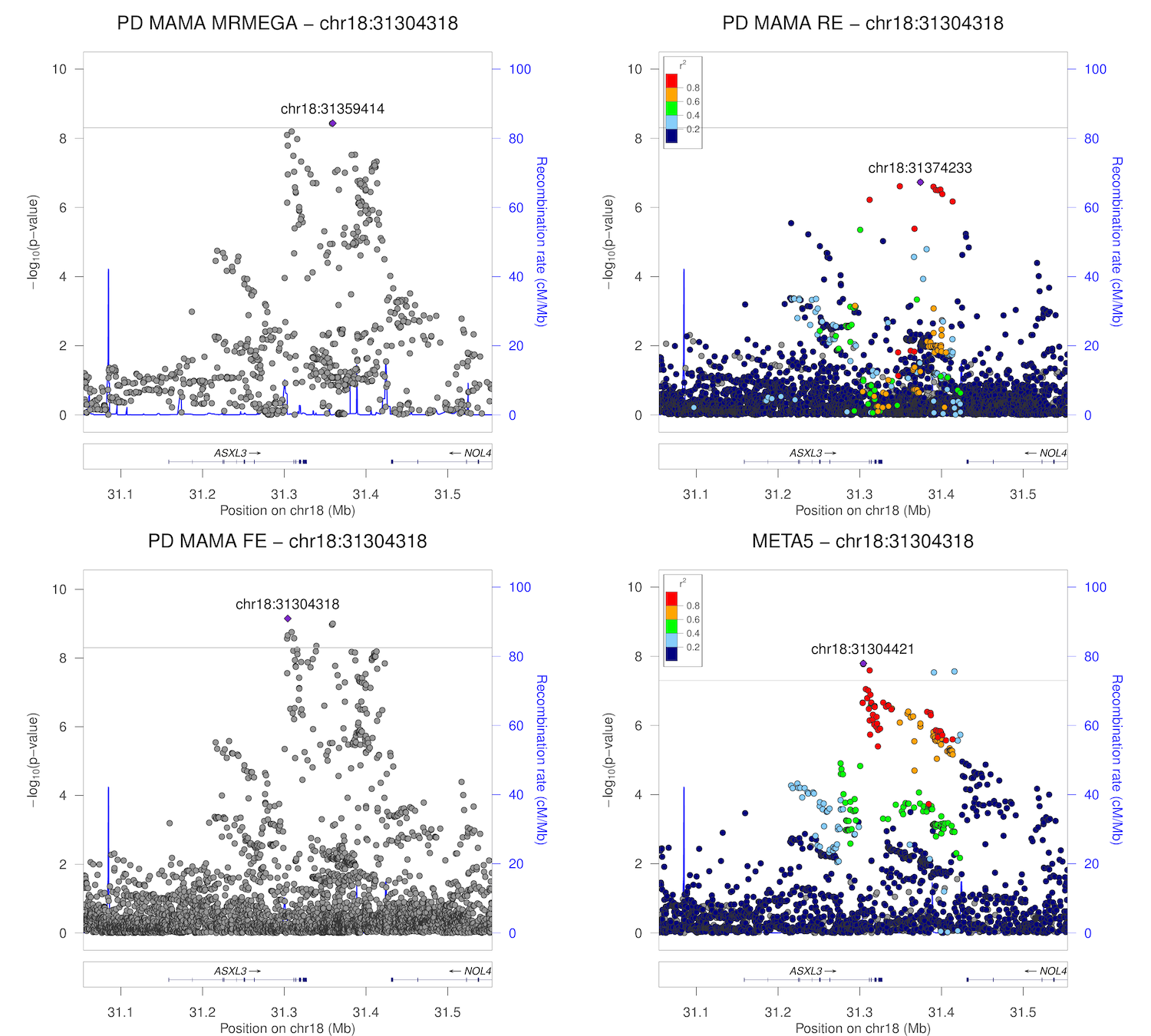

### chr18_40423380-40923380.png

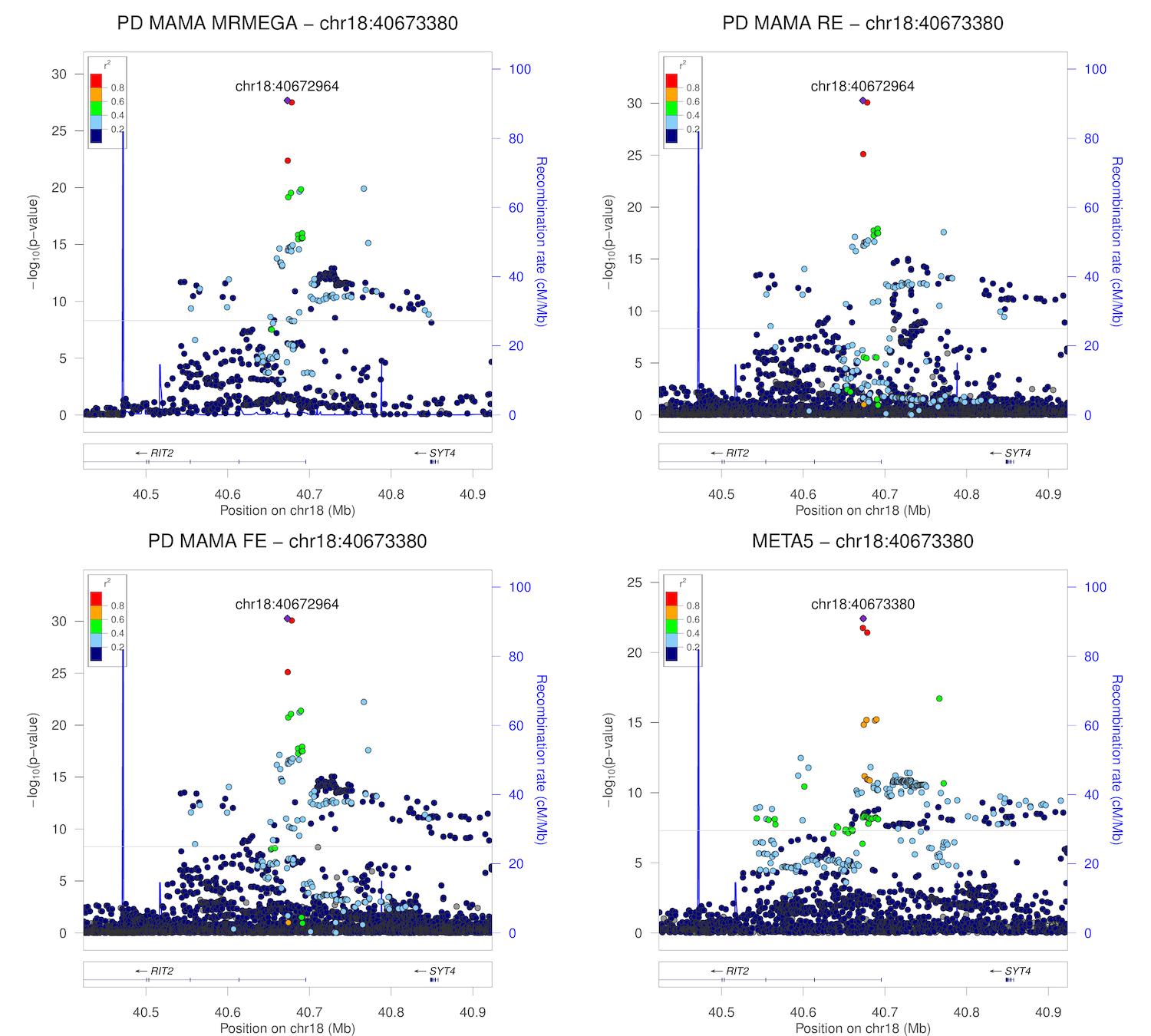

### chr18_48433589-48933589.png

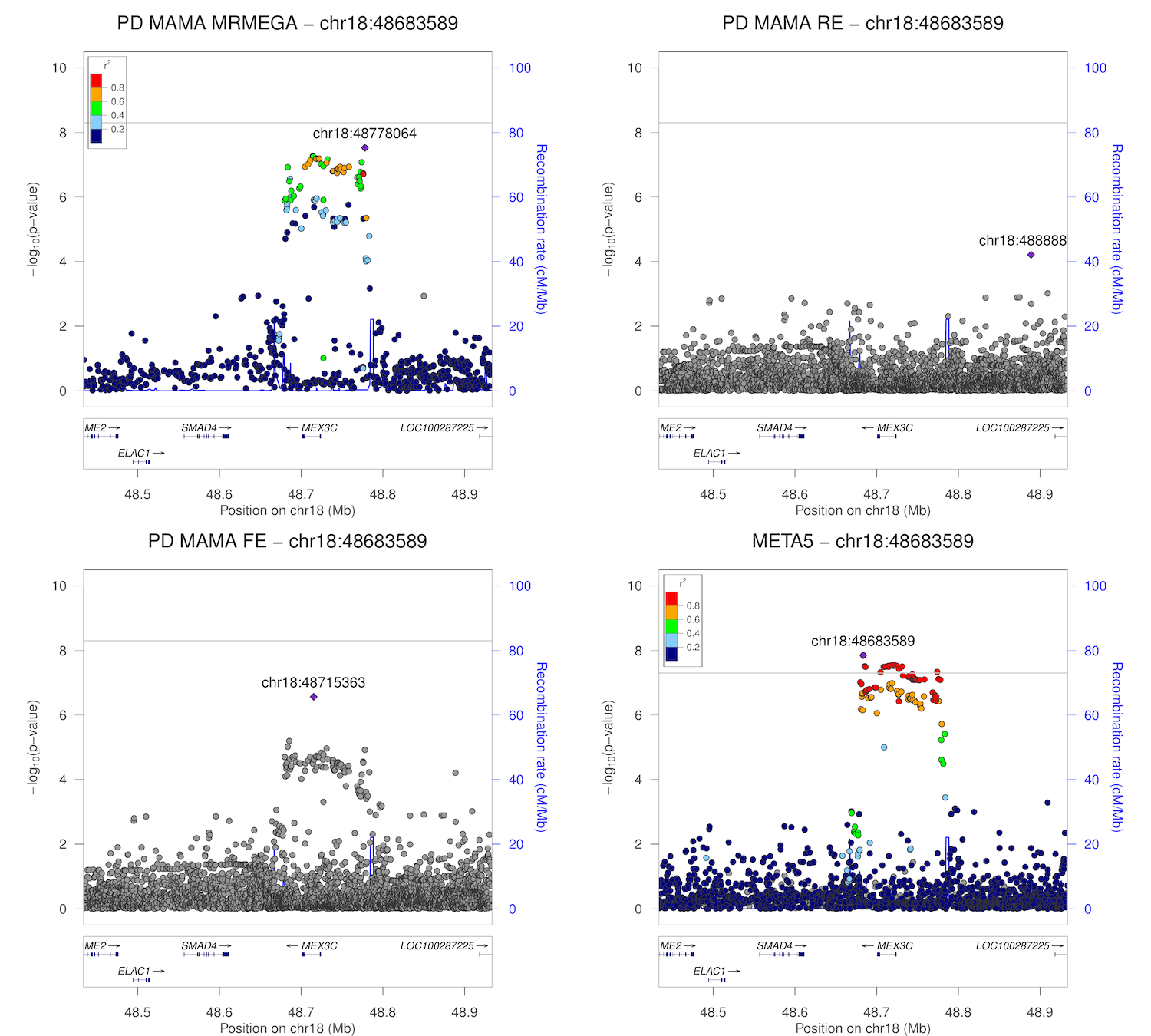

### chr20_5756041-6256041.png

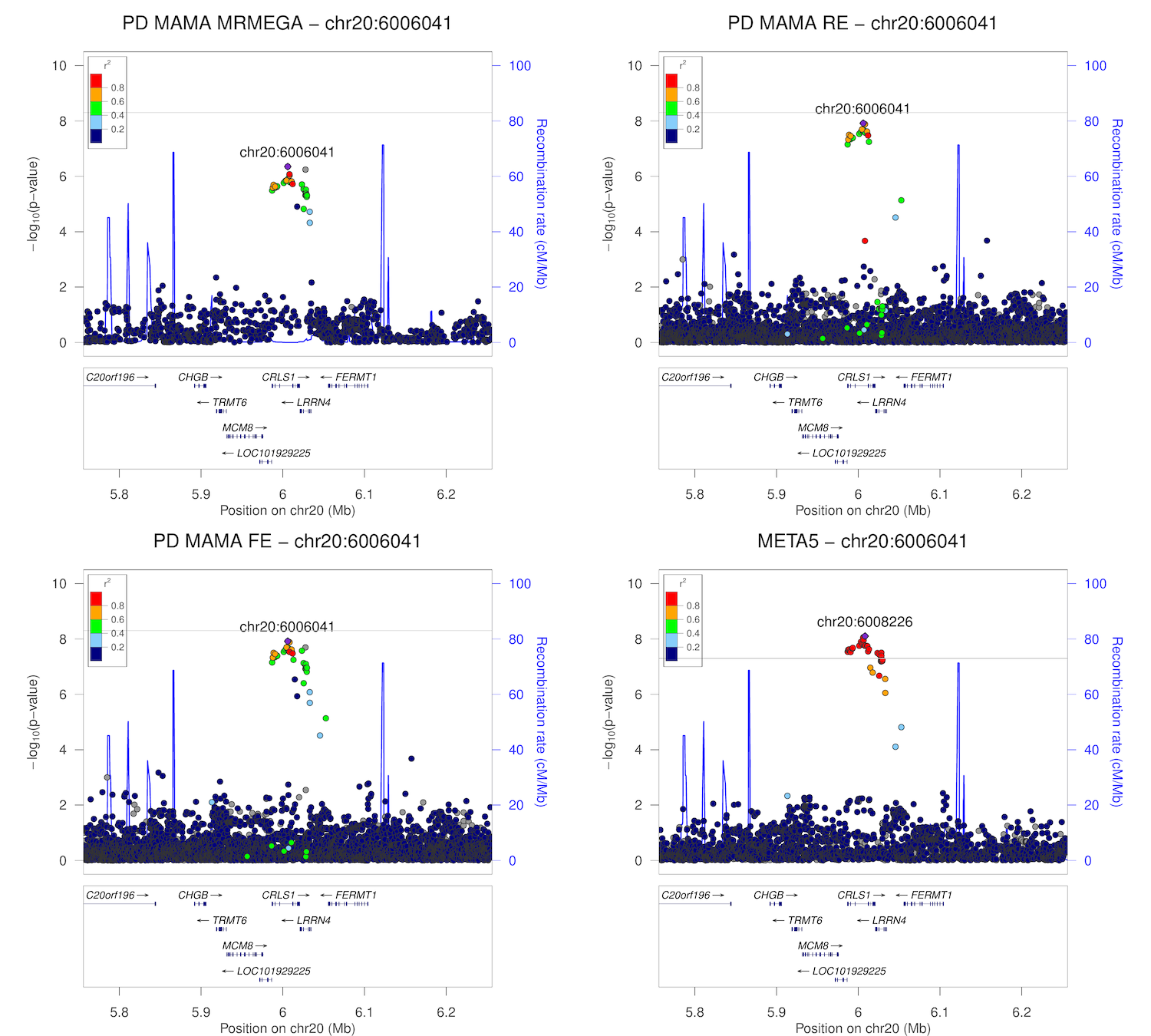

### chr20_30170924-30670924.png

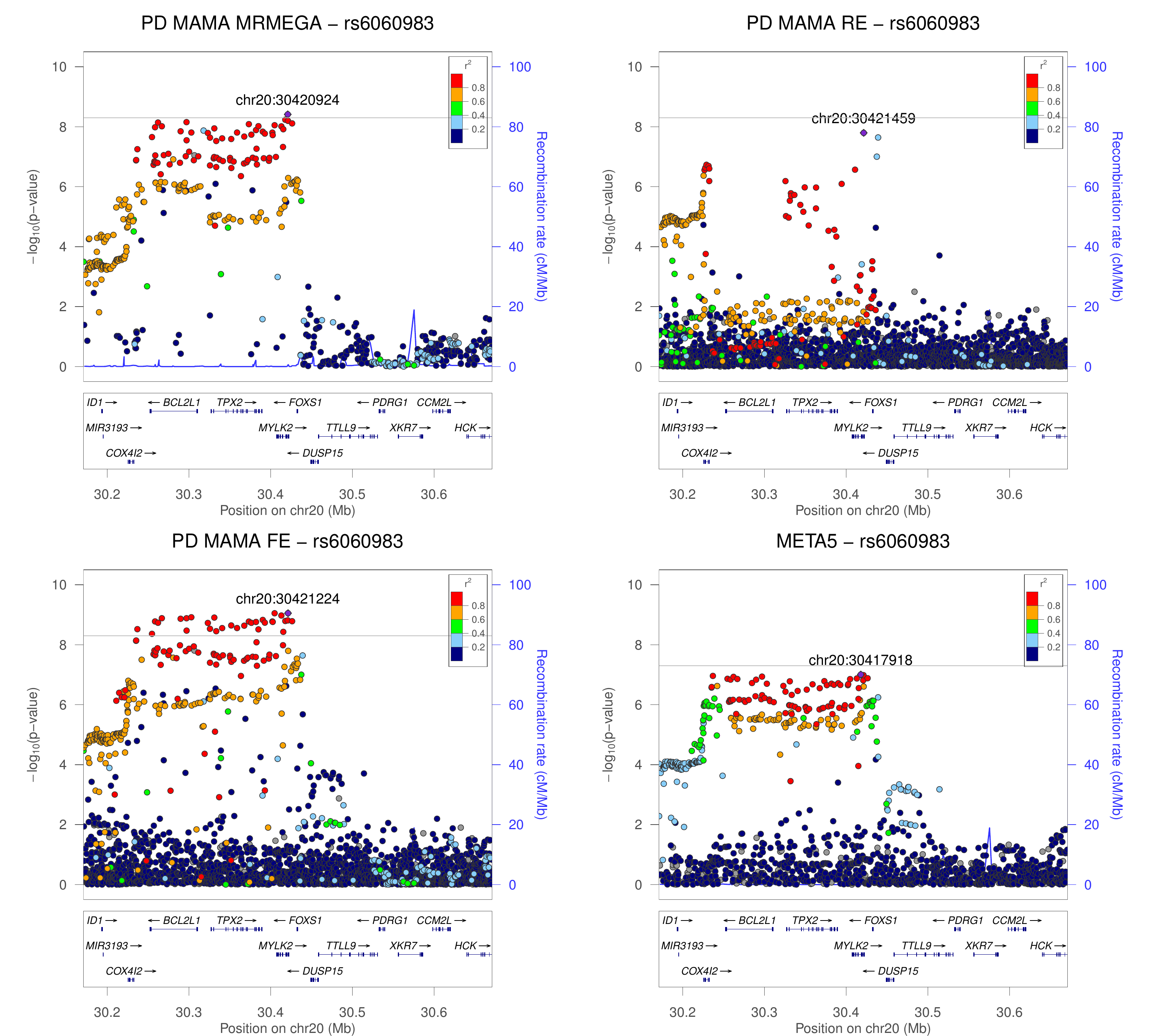

### chr21_16562552-17062552.png

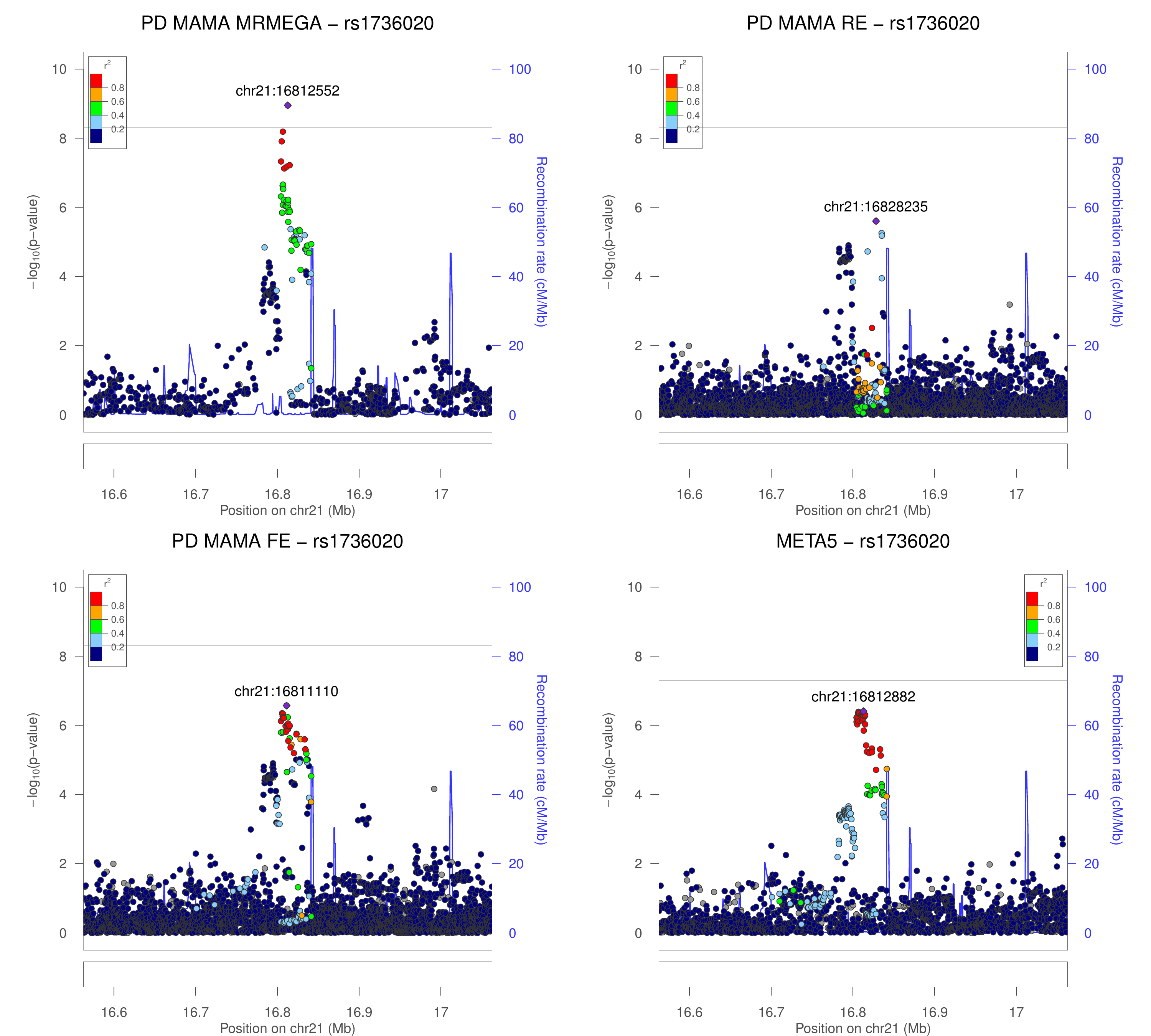

### chr21_38602361-39102361.png

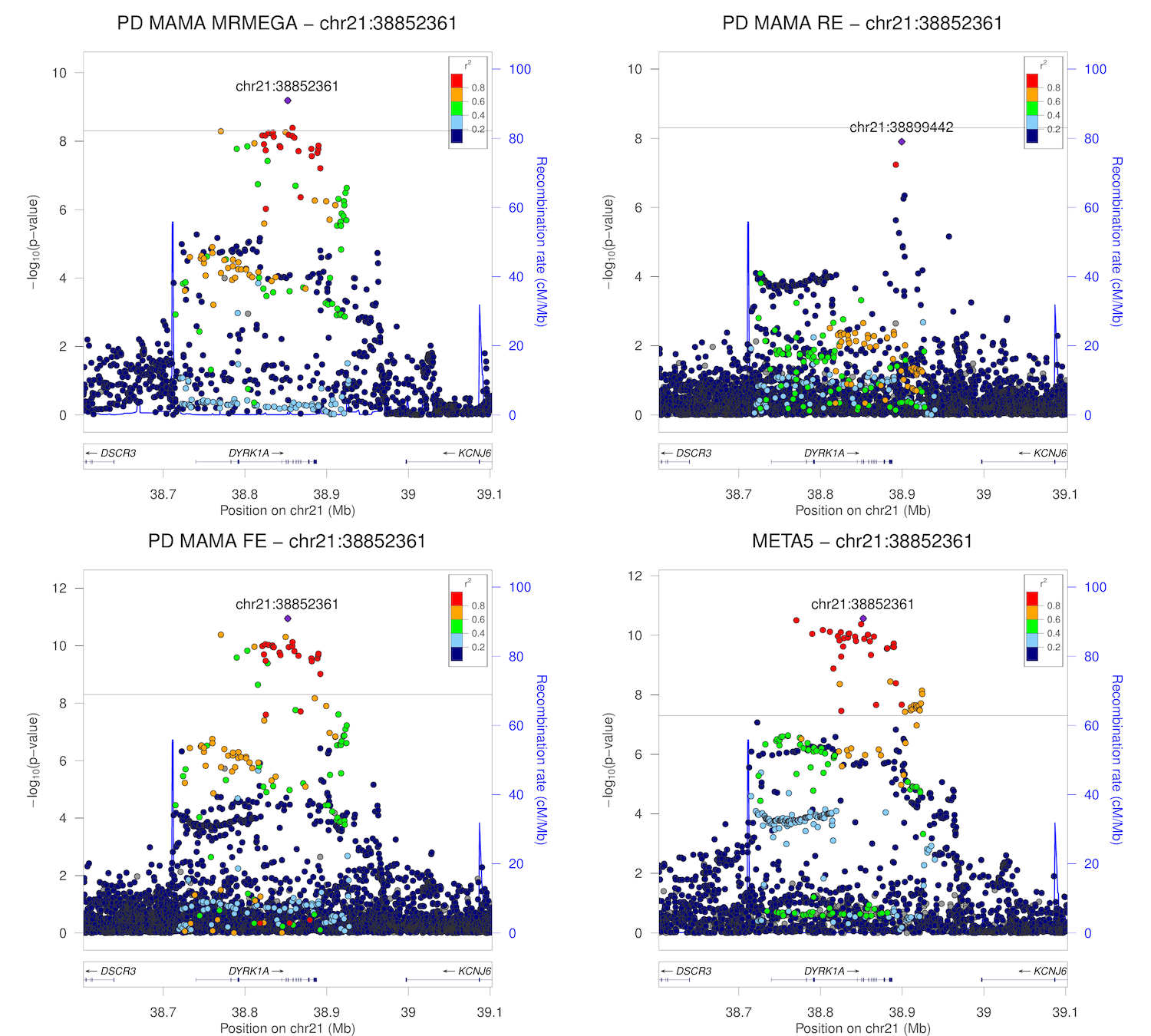

### chr22_41184158-41684158.png

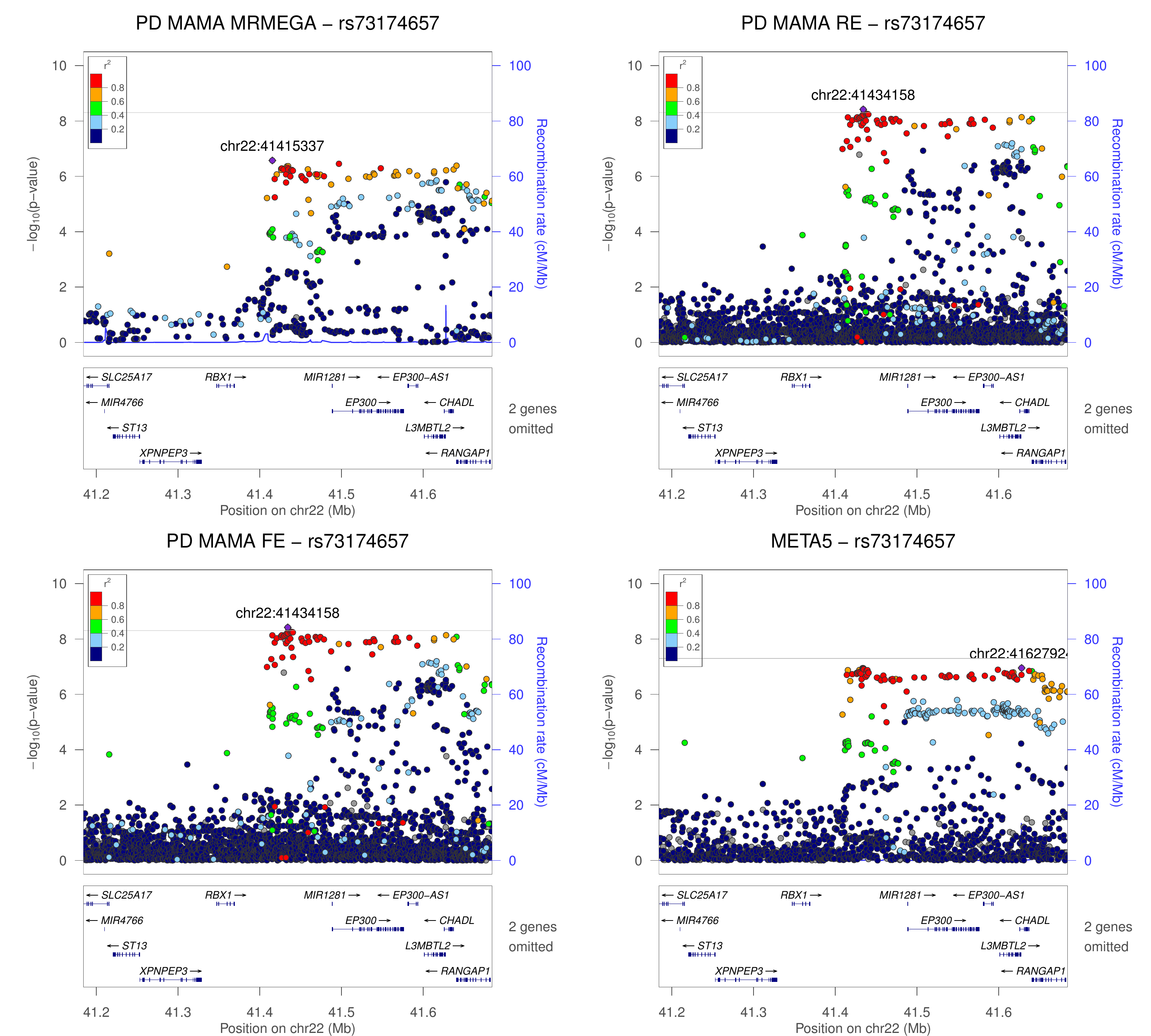

### chr22_50558017-51058017.png

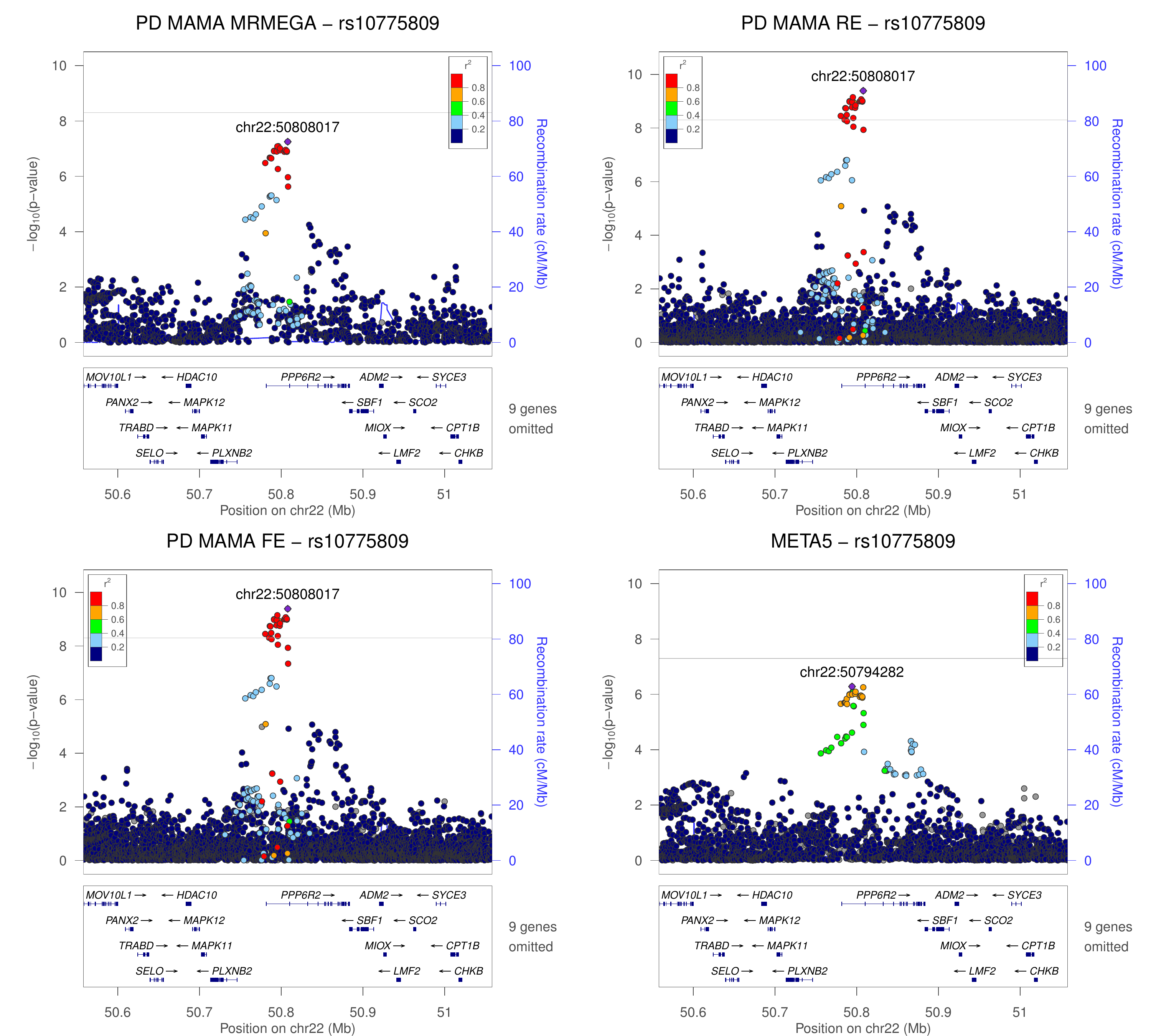

### DropViz_all_level1.png

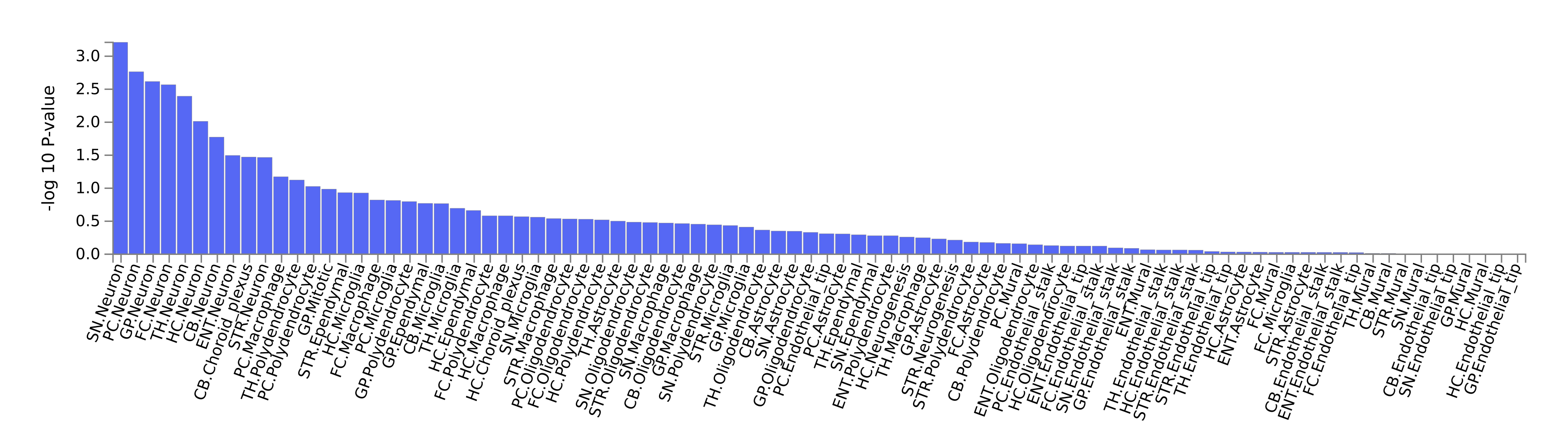

### DropViz_Substantia_Nigra_level2.png

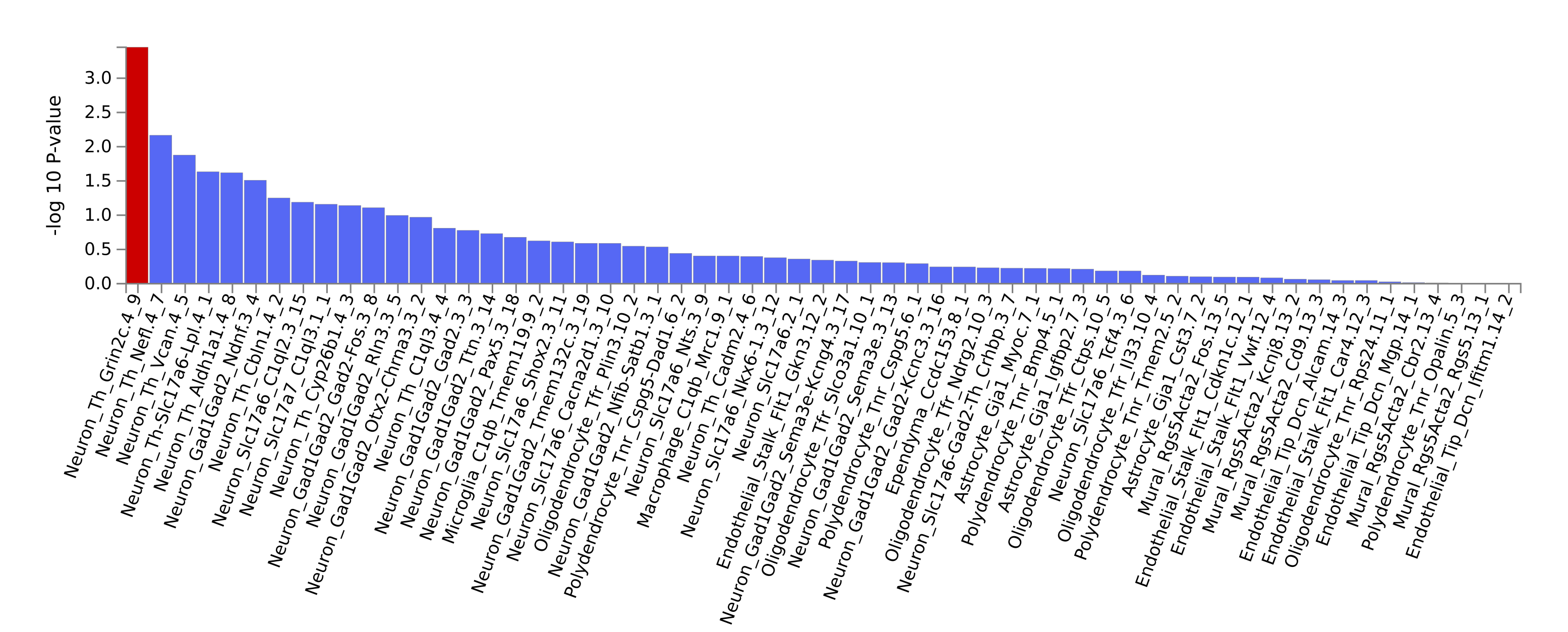

### Linnarsson_GSE76381_Human_Midbrain.png

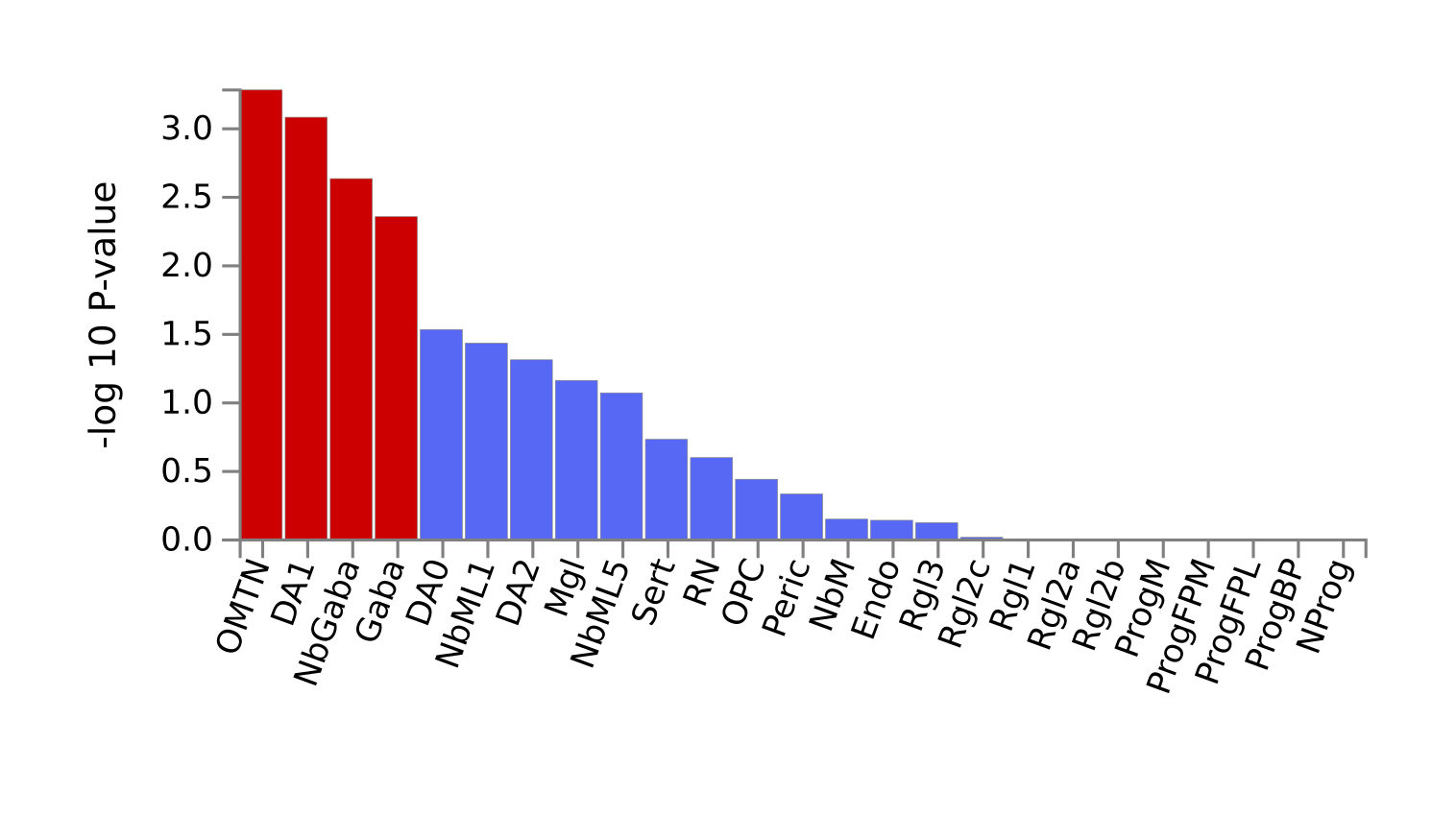

### Supplementary Figure S10A.pdf

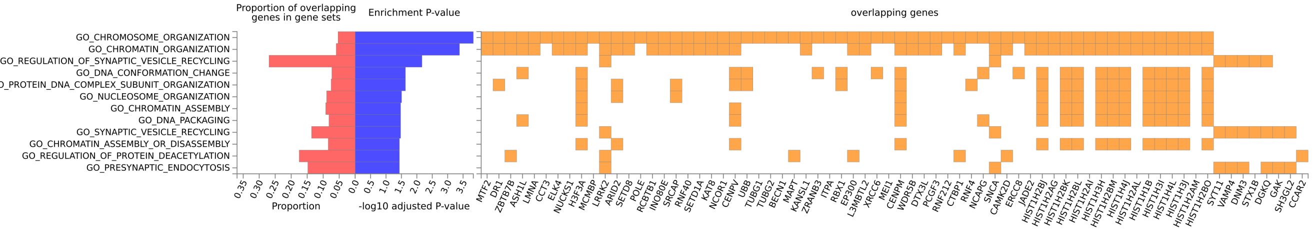

### Supplementary Figure S10B.pdf

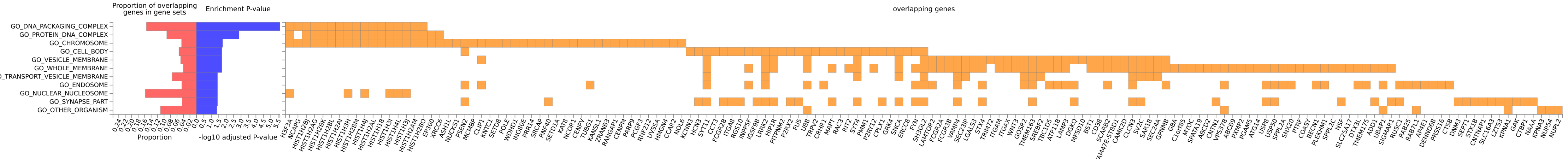

### Supplementary Figure S10C.pdf

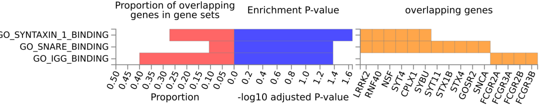

### Supplementary Figure S10D.pdf

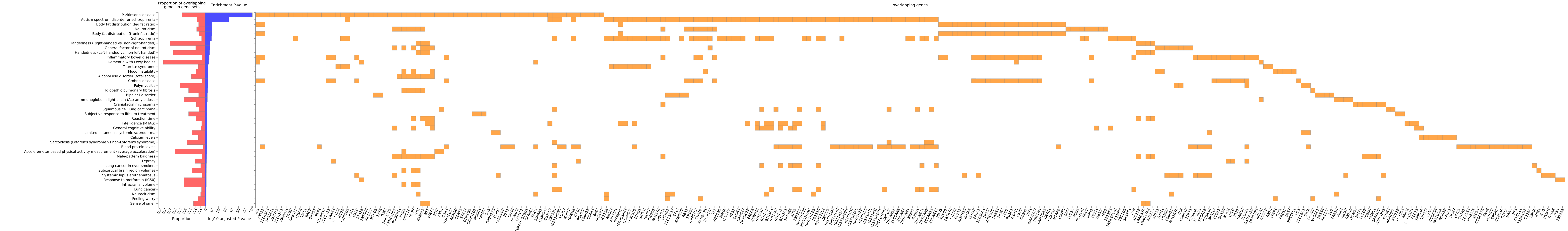

### Supplementary Figure S13 - Miami plot of RE (top) and EUR (bottom).png

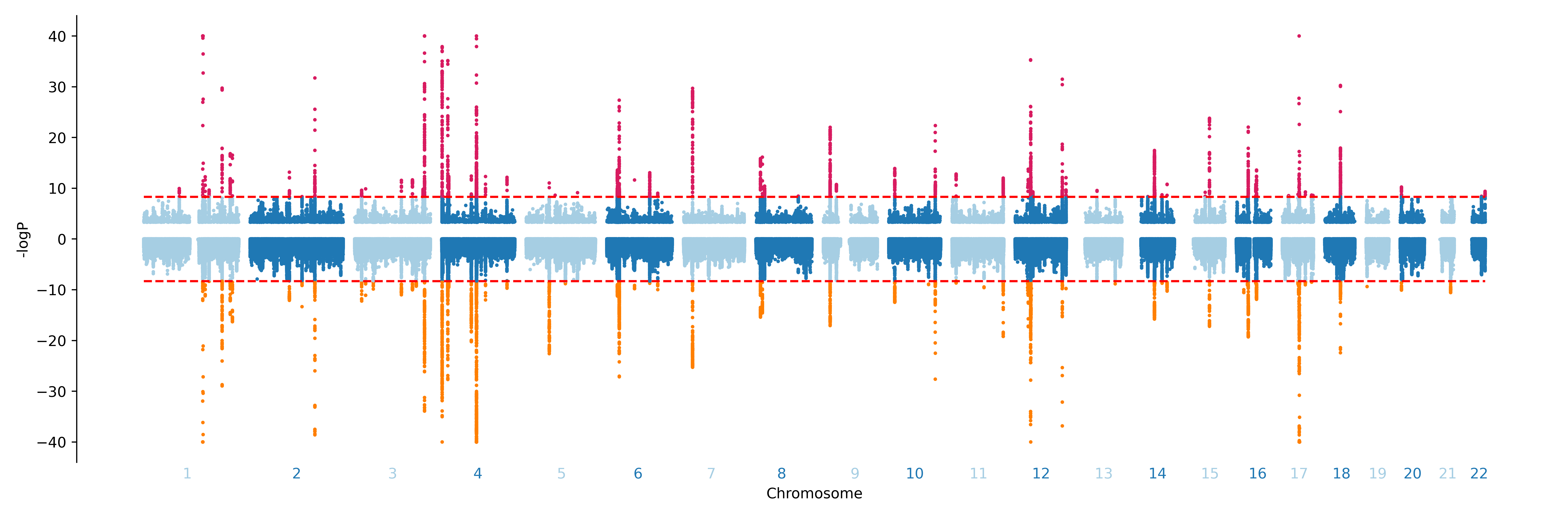
